## Supplementary Tables for "White blood cells and severe COVID-19: a Mendelian randomization study": Supplementary Information. Tables S2-21.docx

* **Address for Correspondence:**

Dr. Kaixiong Ye

**Table S2. 401 SNPs significantly associated with white blood cell count used IVs in forward MR analyses derived from Vuckovic D et al.** Chr: Chromosome; EA: Effect allele; NEA: Non-effect allele; EAF: Effect allele frequency.

| SNP | Chr | Pos | EA | NEA | EAF | Beta | SE | P-value |
| --- | --- | --- | --- | --- | --- | --- | --- | --- |
| rs4908508 | 1 | 8800780 | C | T | 0.206 | 0.028 | 0.003 | 5.3e-27 |
| rs111877086 | 1 | 9702591 | C | T | 0.192 | -0.019 | 0.003 | 2.5e-12 |
| rs284317 | 1 | 10731625 | G | A | 0.495 | 0.019 | 0.002 | 2e-18 |
| rs10917107 | 1 | 22329414 | A | G | 0.690 | -0.015 | 0.002 | 1.7e-10 |
| rs60918921 | 1 | 23850590 | A | C | 0.618 | -0.020 | 0.002 | 5.6e-20 |
| rs71638504 | 1 | 27327822 | A | G | 0.056 | -0.030 | 0.005 | 3.4e-10 |
| rs3762297 | 1 | 31231680 | T | C | 0.183 | 0.026 | 0.003 | 6e-22 |
| rs3917932 | 1 | 36943916 | G | C | 0.577 | -0.044 | 0.002 | 1.6e-95 |
| rs11800162 | 1 | 36970512 | T | C | 0.020 | 0.053 | 0.008 | 4.6e-12 |
| rs3754224 | 1 | 43423622 | C | T | 0.269 | -0.016 | 0.002 | 4.2e-11 |
| rs9429088 | 1 | 46497500 | A | T | 0.435 | -0.022 | 0.002 | 3.4e-25 |
| rs12121236 | 1 | 56617911 | A | C | 0.352 | 0.015 | 0.002 | 7.6e-12 |
| rs11208712 | 1 | 66148652 | C | T | 0.356 | -0.044 | 0.002 | 4.6e-89 |
| rs61780038 | 1 | 67487376 | A | C | 0.559 | -0.018 | 0.002 | 1.2e-14 |
| rs41313381 | 1 | 79411968 | A | C | 0.031 | 0.050 | 0.006 | 6.8e-17 |
| rs150649461 | 1 | 92925654 | C | G | 0.015 | 0.065 | 0.009 | 4.9e-13 |
| rs56674564 | 1 | 101097718 | C | T | 0.354 | -0.016 | 0.002 | 1.8e-12 |
| rs6696259 | 1 | 101221482 | C | G | 0.410 | 0.020 | 0.002 | 7.7e-21 |
| rs9429767 | 1 | 110496087 | A | G | 0.198 | 0.018 | 0.003 | 6.5e-11 |
| rs2476601 | 1 | 114377568 | G | A | 0.899 | 0.052 | 0.004 | 8.1e-50 |
| rs4970996 | 1 | 150506589 | C | G | 0.751 | 0.015 | 0.002 | 2.7e-09 |
| rs568035 | 1 | 156110167 | T | C | 0.064 | -0.032 | 0.004 | 1.8e-13 |
| rs34599082 | 1 | 159175494 | T | C | 0.014 | -0.165 | 0.009 | 2.6e-73 |
| rs11587213 | 1 | 161184875 | G | A | 0.183 | -0.019 | 0.003 | 1.5e-12 |
| rs192123799 | 1 | 161608301 | C | T | 0.125 | -0.034 | 0.003 | 1.4e-24 |
| rs4657690 | 1 | 167601061 | A | G | 0.576 | 0.013 | 0.002 | 5e-10 |
| rs3795503 | 1 | 180905694 | T | C | 0.315 | -0.016 | 0.002 | 2.1e-12 |
| rs1779807 | 1 | 182164250 | T | C | 0.333 | 0.014 | 0.002 | 1.9e-10 |
| rs41272536 | 1 | 183440531 | G | A | 0.046 | -0.033 | 0.005 | 1.2e-10 |
| rs10494783 | 1 | 198663661 | A | G | 0.051 | -0.047 | 0.005 | 3.6e-22 |
| rs1060286 | 1 | 207250300 | A | G | 0.611 | 0.013 | 0.002 | 6.9e-10 |
| rs4844390 | 1 | 207934849 | G | A | 0.222 | -0.025 | 0.003 | 1.2e-23 |
| rs10916617 | 1 | 224637070 | T | C | 0.212 | 0.026 | 0.003 | 3.5e-23 |
| rs2615061 | 1 | 225895806 | A | G | 0.116 | -0.029 | 0.003 | 5.8e-18 |
| rs7552783 | 1 | 227174145 | C | T | 0.512 | -0.017 | 0.002 | 9.2e-14 |
| rs533483 | 1 | 234765256 | A | G | 0.246 | -0.017 | 0.002 | 3.3e-11 |
| rs9970896 | 1 | 236104981 | T | A | 0.888 | -0.060 | 0.003 | 8.2e-71 |
| rs34298354 | 1 | 247588053 | T | C | 0.122 | -0.025 | 0.003 | 6.8e-15 |
| rs56188865 | 1 | 247606276 | C | T | 0.372 | -0.026 | 0.002 | 1.1e-32 |
| rs1469217 | 2 | 12950124 | C | T | 0.493 | 0.019 | 0.002 | 3.1e-19 |
| rs4632345 | 2 | 16702654 | A | G | 0.678 | -0.015 | 0.002 | 1.7e-10 |
| rs377110161 | 2 | 23963462 | G | A | 0.128 | 0.036 | 0.003 | 2.9e-29 |
| rs1260326 | 2 | 27730940 | C | T | 0.606 | -0.033 | 0.002 | 3.1e-53 |
| rs1509396 | 2 | 28612213 | C | G | 0.507 | 0.013 | 0.002 | 7.8e-10 |
| rs935655 | 2 | 46067445 | G | T | 0.762 | 0.016 | 0.002 | 1.7e-10 |
| rs75475627 | 2 | 54787592 | G | C | 0.076 | 0.035 | 0.004 | 6.5e-18 |
| rs2421200 | 2 | 61711815 | T | G | 0.487 | -0.022 | 0.002 | 3e-26 |
| rs7569257 | 2 | 65656976 | A | G | 0.409 | 0.026 | 0.002 | 3e-33 |
| rs4599108 | 2 | 85543222 | T | C | 0.486 | -0.018 | 0.002 | 1.3e-16 |
| rs7575217 | 2 | 101776932 | G | A | 0.673 | 0.023 | 0.002 | 9.2e-24 |
| rs7370196 | 2 | 102297387 | C | T | 0.685 | 0.016 | 0.002 | 4.9e-12 |
| rs13011687 | 2 | 102535941 | C | A | 0.257 | 0.014 | 0.002 | 2.3e-09 |
| rs35898589 | 2 | 112915809 | G | A | 0.048 | 0.031 | 0.005 | 4.1e-10 |
| rs55709272 | 2 | 113867288 | C | T | 0.437 | 0.035 | 0.002 | 6.7e-61 |
| rs77524493 | 2 | 128618594 | T | A | 0.029 | 0.041 | 0.007 | 1.7e-10 |
| rs79716587 | 2 | 143886819 | A | G | 0.126 | -0.036 | 0.003 | 1.7e-29 |
| rs3856364 | 2 | 145477217 | G | C | 0.681 | 0.015 | 0.002 | 1.3e-10 |
| rs62173240 | 2 | 160352482 | A | G | 0.023 | -0.054 | 0.007 | 7.4e-14 |
| rs2729707 | 2 | 160687231 | G | A | 0.832 | -0.032 | 0.003 | 1.5e-29 |
| rs2068330 | 2 | 163237390 | G | C | 0.358 | 0.019 | 0.002 | 3.3e-17 |
| rs2632372 | 2 | 169717541 | C | T | 0.504 | -0.021 | 0.002 | 8e-24 |
| rs17270868 | 2 | 182214669 | C | T | 0.241 | 0.033 | 0.002 | 1.7e-40 |
| rs7573465 | 2 | 182315885 | T | G | 0.557 | 0.036 | 0.002 | 2.4e-62 |
| rs12987470 | 2 | 188323311 | T | A | 0.295 | -0.017 | 0.002 | 1.5e-13 |
| rs13392977 | 2 | 192514856 | A | G | 0.054 | 0.033 | 0.005 | 1.3e-12 |
| rs6760993 | 2 | 202110917 | A | G | 0.530 | 0.016 | 0.002 | 1.1e-14 |
| rs1047891 | 2 | 211540507 | A | C | 0.316 | -0.020 | 0.002 | 1.6e-18 |
| rs114050631 | 2 | 219020958 | T | C | 0.011 | -0.139 | 0.011 | 6.4e-38 |
| rs72951772 | 2 | 219990997 | A | C | 0.127 | -0.020 | 0.003 | 7.4e-10 |
| rs6755895 | 2 | 232579795 | C | T | 0.227 | -0.024 | 0.003 | 3.9e-21 |
| rs10164769 | 2 | 237779229 | T | C | 0.740 | 0.027 | 0.002 | 7.2e-28 |
| rs1822534 | 3 | 12266804 | G | A | 0.392 | -0.029 | 0.002 | 1.7e-39 |
| rs479404 | 3 | 27363064 | C | T | 0.269 | -0.015 | 0.002 | 5e-10 |
| rs2371108 | 3 | 27757018 | T | G | 0.387 | 0.021 | 0.002 | 3e-22 |
| rs3732378 | 3 | 39307162 | A | G | 0.173 | 0.023 | 0.003 | 7.3e-16 |
| rs9872570 | 3 | 42896515 | A | T | 0.385 | -0.026 | 0.002 | 1.6e-32 |
| rs4682844 | 3 | 46982737 | C | T | 0.419 | -0.027 | 0.002 | 4.1e-33 |
| rs6779340 | 3 | 58033701 | G | C | 0.336 | -0.020 | 0.002 | 7.6e-19 |
| rs832187 | 3 | 63833050 | T | C | 0.634 | -0.014 | 0.002 | 6.1e-11 |
| rs6445424 | 3 | 64717575 | A | C | 0.238 | -0.016 | 0.002 | 2.5e-10 |
| rs17737336 | 3 | 70764744 | C | T | 0.237 | -0.017 | 0.003 | 3.2e-11 |
| rs7639292 | 3 | 107295665 | T | C | 0.170 | -0.026 | 0.003 | 7.1e-20 |
| rs34198449 | 3 | 128381270 | A | G | 0.270 | -0.029 | 0.002 | 2.9e-33 |
| rs10935013 | 3 | 132227935 | A | G | 0.113 | 0.022 | 0.003 | 6.5e-11 |
| rs11712552 | 3 | 140957762 | G | A | 0.127 | 0.030 | 0.003 | 1.3e-20 |
| rs9819371 | 3 | 141206800 | T | C | 0.065 | -0.048 | 0.004 | 6.4e-29 |
| rs6440050 | 3 | 141622749 | C | G | 0.501 | -0.013 | 0.002 | 2.4e-10 |
| rs6440732 | 3 | 150990510 | A | C | 0.825 | 0.023 | 0.003 | 1.8e-16 |
| rs55730982 | 3 | 156797208 | G | T | 0.401 | 0.014 | 0.002 | 7.6e-11 |
| rs12494724 | 3 | 169320998 | T | C | 0.634 | -0.014 | 0.002 | 5.2e-11 |
| rs4074672 | 3 | 183730295 | T | C | 0.369 | 0.019 | 0.002 | 1.6e-18 |
| rs9867398 | 3 | 185912816 | T | C | 0.094 | 0.022 | 0.004 | 2.3e-09 |
| rs4490348 | 3 | 186204424 | A | G | 0.410 | 0.013 | 0.002 | 1.9e-09 |
| rs2089979 | 3 | 196501413 | G | A | 0.416 | -0.035 | 0.002 | 2.5e-59 |
| rs56011263 | 4 | 702972 | C | T | 0.388 | 0.022 | 0.002 | 3.1e-24 |
| rs73191188 | 4 | 3105200 | A | G | 0.353 | -0.016 | 0.002 | 1e-12 |
| rs6831368 | 4 | 6969919 | G | A | 0.362 | -0.020 | 0.002 | 4.1e-20 |
| rs28530750 | 4 | 36312542 | A | G | 0.043 | 0.058 | 0.005 | 6.8e-28 |
| rs987121 | 4 | 38329783 | T | A | 0.603 | 0.015 | 0.002 | 1.4e-11 |
| rs13132853 | 4 | 38680015 | G | A | 0.361 | 0.024 | 0.002 | 3.5e-27 |
| rs218264 | 4 | 55408875 | T | A | 0.249 | 0.029 | 0.002 | 8.8e-32 |
| rs723585 | 4 | 55503194 | G | A | 0.486 | -0.019 | 0.002 | 8.5e-20 |
| rs2412771 | 4 | 57761417 | C | T | 0.417 | -0.020 | 0.002 | 9.2e-20 |
| rs11723621 | 4 | 72615362 | G | A | 0.292 | -0.031 | 0.002 | 3.8e-39 |
| rs16850073 | 4 | 74703999 | T | C | 0.375 | 0.043 | 0.002 | 2.6e-83 |
| rs11725704 | 4 | 74959996 | G | A | 0.376 | 0.065 | 0.002 | 7.5e-194 |
| rs116746683 | 4 | 83471084 | T | C | 0.062 | -0.032 | 0.004 | 4.1e-13 |
| rs28378092 | 4 | 90228807 | T | C | 0.458 | 0.013 | 0.002 | 3.4e-09 |
| rs113473633 | 4 | 103449131 | G | A | 0.026 | -0.050 | 0.007 | 1e-12 |
| rs144317085 | 4 | 105806108 | T | A | 0.034 | 0.049 | 0.006 | 2.6e-16 |
| rs74693727 | 4 | 120330155 | T | C | 0.328 | -0.017 | 0.002 | 1.5e-14 |
| rs11735662 | 4 | 145026126 | T | C | 0.034 | 0.053 | 0.006 | 5.7e-20 |
| rs2290846 | 4 | 151199080 | A | G | 0.287 | 0.018 | 0.002 | 7.6e-14 |
| rs6817881 | 4 | 152223904 | T | C | 0.500 | 0.014 | 0.002 | 1.3e-10 |
| rs4535497 | 5 | 1107428 | A | C | 0.570 | -0.013 | 0.002 | 4.3e-10 |
| rs7705526 | 5 | 1285974 | A | C | 0.326 | 0.030 | 0.002 | 1.2e-38 |
| rs464609 | 5 | 34654477 | A | G | 0.544 | 0.014 | 0.002 | 1.7e-10 |
| rs3822733 | 5 | 35875134 | C | T | 0.259 | 0.022 | 0.002 | 3e-20 |
| rs13155659 | 5 | 54987585 | C | T | 0.699 | -0.014 | 0.002 | 7.8e-10 |
| rs11744663 | 5 | 57315635 | A | G | 0.192 | -0.020 | 0.003 | 8.2e-14 |
| rs2964199 | 5 | 57532775 | C | T | 0.682 | -0.016 | 0.002 | 7.8e-13 |
| rs11741775 | 5 | 68590395 | T | C | 0.443 | -0.022 | 0.002 | 1.8e-24 |
| rs4703890 | 5 | 71748624 | A | G | 0.882 | -0.039 | 0.003 | 7.2e-32 |
| rs6876611 | 5 | 96243598 | T | A | 0.486 | -0.013 | 0.002 | 2.8e-09 |
| rs141543178 | 5 | 108133865 | T | A | 0.026 | 0.044 | 0.007 | 6.7e-11 |
| rs257063 | 5 | 114806819 | T | C | 0.743 | -0.015 | 0.002 | 7e-10 |
| rs79272926 | 5 | 118726637 | A | C | 0.220 | -0.025 | 0.003 | 4.4e-22 |
| rs4836224 | 5 | 122092875 | C | A | 0.426 | 0.014 | 0.002 | 1.9e-11 |
| rs2631367 | 5 | 131705458 | G | C | 0.522 | 0.034 | 0.002 | 3.4e-58 |
| rs79237520 | 5 | 134712566 | T | C | 0.023 | 0.043 | 0.007 | 1.5e-09 |
| rs6580229 | 5 | 141510754 | G | A | 0.617 | 0.025 | 0.002 | 8.6e-31 |
| rs2082382 | 5 | 148200553 | A | G | 0.548 | -0.021 | 0.002 | 1.9e-22 |
| rs4704826 | 5 | 156392082 | A | C | 0.635 | -0.016 | 0.002 | 4e-13 |
| rs10053217 | 5 | 173102585 | G | A | 0.355 | 0.014 | 0.002 | 8.1e-10 |
| rs2594836 | 5 | 173205318 | A | G | 0.722 | -0.036 | 0.002 | 1.4e-50 |
| rs13180726 | 5 | 179126457 | A | G | 0.827 | -0.022 | 0.003 | 1.5e-14 |
| rs12203592 | 6 | 396321 | T | C | 0.220 | 0.024 | 0.003 | 4.2e-20 |
| rs1761678 | 6 | 7090640 | T | G | 0.873 | -0.021 | 0.003 | 9.3e-11 |
| rs1285886 | 6 | 7140831 | A | G | 0.197 | 0.029 | 0.003 | 1.2e-27 |
| rs1334576 | 6 | 7211818 | A | G | 0.425 | -0.013 | 0.002 | 1.3e-09 |
| rs1087924 | 6 | 10523826 | C | G | 0.529 | -0.013 | 0.002 | 4.3e-10 |
| rs1144700 | 6 | 16744687 | T | C | 0.178 | -0.031 | 0.003 | 3.3e-29 |
| rs9350354 | 6 | 21393645 | G | A | 0.362 | 0.016 | 0.002 | 1.5e-13 |
| rs62392759 | 6 | 25804685 | A | C | 0.125 | 0.021 | 0.003 | 4.9e-11 |
| rs73740434 | 6 | 27144980 | A | C | 0.082 | -0.066 | 0.005 | 2.8e-37 |
| rs6902400 | 6 | 30425276 | T | C | 0.179 | 0.034 | 0.003 | 4.5e-34 |
| rs2905757 | 6 | 31022828 | T | C | 0.324 | 0.069 | 0.002 | 1e-200 |
| rs9265546 | 6 | 31298202 | A | G | 0.223 | -0.067 | 0.003 | 2.3e-112 |
| rs200801362 | 6 | 31555480 | C | T | 0.161 | -0.090 | 0.004 | 1e-116 |
| rs74973088 | 6 | 32457874 | C | T | 0.465 | 0.077 | 0.002 | 1e-200 |
| rs9274305 | 6 | 32632000 | T | C | 0.205 | -0.052 | 0.003 | 2.7e-75 |
| rs149066177 | 6 | 34219560 | A | T | 0.112 | -0.021 | 0.003 | 3.4e-10 |
| rs6902672 | 6 | 41165840 | C | T | 0.207 | -0.019 | 0.003 | 4.4e-13 |
| rs11965885 | 6 | 43693094 | T | G | 0.582 | -0.016 | 0.002 | 5.5e-14 |
| rs6937438 | 6 | 43815364 | A | G | 0.707 | 0.016 | 0.002 | 7.8e-12 |
| rs900403 | 6 | 44591850 | A | G | 0.425 | -0.013 | 0.002 | 3.6e-10 |
| rs2012011 | 6 | 46893355 | A | G | 0.135 | -0.027 | 0.003 | 2.5e-18 |
| rs71548708 | 6 | 87837599 | T | C | 0.497 | -0.018 | 0.002 | 3.7e-17 |
| rs4707609 | 6 | 90946479 | C | T | 0.362 | -0.022 | 0.002 | 4.1e-24 |
| rs364663 | 6 | 105443189 | A | T | 0.557 | -0.014 | 0.002 | 1.3e-10 |
| rs6927569 | 6 | 109621494 | C | T | 0.523 | 0.023 | 0.002 | 2.3e-28 |
| rs1741820 | 6 | 122721510 | G | A | 0.545 | 0.021 | 0.002 | 4.6e-22 |
| rs35786788 | 6 | 135419042 | A | G | 0.253 | -0.050 | 0.002 | 2.4e-90 |
| rs9402702 | 6 | 135634006 | C | T | 0.492 | 0.020 | 0.002 | 4.9e-21 |
| rs6924387 | 6 | 137082948 | G | A | 0.410 | 0.017 | 0.002 | 1.2e-14 |
| rs72978754 | 6 | 138053203 | C | T | 0.066 | -0.029 | 0.004 | 1.2e-11 |
| rs9390461 | 6 | 147701217 | G | A | 0.540 | 0.016 | 0.002 | 8.5e-14 |
| rs1738074 | 6 | 159465977 | C | T | 0.565 | -0.020 | 0.002 | 3.7e-21 |
| rs71536939 | 6 | 170486950 | G | A | 0.785 | 0.017 | 0.003 | 1.2e-09 |
| rs798540 | 7 | 2764130 | A | G | 0.297 | 0.019 | 0.002 | 8.3e-16 |
| rs2462661 | 7 | 6702311 | G | T | 0.576 | 0.017 | 0.002 | 1.1e-15 |
| rs56195338 | 7 | 8107922 | A | G | 0.058 | -0.032 | 0.005 | 1.1e-12 |
| rs7776857 | 7 | 22754768 | T | G | 0.655 | -0.019 | 0.002 | 1.4e-17 |
| rs2158799 | 7 | 28277107 | G | C | 0.610 | 0.043 | 0.002 | 5e-88 |
| rs56388170 | 7 | 28724374 | T | G | 0.292 | 0.060 | 0.002 | 7.6e-144 |
| rs2710804 | 7 | 36084529 | C | T | 0.377 | 0.023 | 0.002 | 7.1e-26 |
| rs3735485 | 7 | 45009341 | G | A | 0.845 | 0.040 | 0.003 | 1.2e-42 |
| rs865031 | 7 | 47448640 | C | A | 0.431 | -0.013 | 0.002 | 3.9e-10 |
| rs6583435 | 7 | 50293405 | A | C | 0.676 | 0.019 | 0.002 | 1.5e-16 |
| rs4689 | 7 | 56067016 | T | G | 0.760 | -0.018 | 0.002 | 1.5e-12 |
| rs4276561 | 7 | 66120952 | C | T | 0.046 | -0.037 | 0.005 | 2e-13 |
| rs33951980 | 7 | 73029437 | T | C | 0.130 | -0.025 | 0.003 | 7.2e-16 |
| rs79864016 | 7 | 75655838 | G | A | 0.166 | -0.018 | 0.003 | 7.6e-10 |
| rs8179 | 7 | 92236164 | C | T | 0.791 | -0.038 | 0.003 | 4.4e-48 |
| rs445 | 7 | 92408370 | T | C | 0.096 | -0.100 | 0.004 | 8.6e-169 |
| rs2107717 | 7 | 98005398 | C | T | 0.679 | -0.014 | 0.002 | 1.3e-09 |
| rs342293 | 7 | 106372219 | G | C | 0.459 | -0.017 | 0.002 | 2.4e-16 |
| rs11768743 | 7 | 129632081 | T | A | 0.214 | 0.016 | 0.003 | 3.9e-09 |
| rs9656395 | 7 | 130575884 | G | A | 0.095 | -0.030 | 0.004 | 1.5e-16 |
| rs7803075 | 7 | 130742066 | G | A | 0.739 | -0.026 | 0.002 | 5e-27 |
| rs13231262 | 7 | 148886058 | T | C | 0.656 | -0.016 | 0.002 | 5.9e-13 |
| rs7464263 | 8 | 11434176 | T | A | 0.529 | 0.025 | 0.002 | 9.5e-31 |
| rs12550612 | 8 | 22966769 | A | G | 0.824 | -0.034 | 0.003 | 9.2e-34 |
| rs2322586 | 8 | 27323182 | G | C | 0.857 | -0.019 | 0.003 | 3.2e-10 |
| rs2979489 | 8 | 30280833 | A | G | 0.742 | 0.018 | 0.002 | 2.5e-14 |
| rs28571765 | 8 | 55449281 | C | T | 0.209 | -0.022 | 0.003 | 3.9e-17 |
| rs12676105 | 8 | 56795439 | T | C | 0.505 | -0.017 | 0.002 | 1.9e-16 |
| rs7822169 | 8 | 61417284 | G | A | 0.371 | 0.021 | 0.002 | 5.4e-22 |
| rs7846314 | 8 | 61650831 | T | A | 0.187 | 0.049 | 0.003 | 4.4e-71 |
| rs4623401 | 8 | 68797804 | G | A | 0.467 | -0.019 | 0.002 | 1.8e-19 |
| rs1947897 | 8 | 78946874 | C | G | 0.332 | -0.016 | 0.002 | 3.4e-13 |
| rs62510269 | 8 | 79012343 | G | A | 0.147 | -0.037 | 0.003 | 1.4e-34 |
| rs10808538 | 8 | 87169063 | A | G | 0.297 | 0.014 | 0.002 | 9.1e-10 |
| rs10104995 | 8 | 103901234 | T | C | 0.231 | -0.022 | 0.003 | 2e-18 |
| rs2957421 | 8 | 106275376 | A | G | 0.243 | 0.016 | 0.002 | 1.8e-10 |
| rs4734879 | 8 | 106583124 | G | A | 0.275 | -0.022 | 0.002 | 5.3e-20 |
| rs71510325 | 8 | 126298640 | T | C | 0.598 | 0.014 | 0.002 | 7.1e-11 |
| rs4733823 | 8 | 129014697 | T | C | 0.456 | 0.021 | 0.002 | 4.9e-23 |
| rs59697075 | 8 | 130618150 | T | C | 0.588 | -0.038 | 0.002 | 3.2e-69 |
| rs145209947 | 8 | 130694185 | A | C | 0.015 | -0.075 | 0.010 | 1.4e-14 |
| rs7005996 | 8 | 142241681 | T | C | 0.906 | 0.026 | 0.004 | 5.1e-13 |
| rs34634548 | 8 | 142329460 | A | C | 0.358 | -0.028 | 0.002 | 3.9e-37 |
| rs2992836 | 9 | 276053 | T | G | 0.188 | 0.024 | 0.003 | 1e-18 |
| rs2219143 | 9 | 2622278 | A | G | 0.393 | 0.020 | 0.002 | 4.8e-19 |
| rs385893 | 9 | 4763176 | C | T | 0.524 | 0.020 | 0.002 | 7.1e-22 |
| rs10758669 | 9 | 4981602 | A | C | 0.659 | -0.014 | 0.002 | 2.8e-10 |
| rs7036656 | 9 | 21990457 | T | C | 0.722 | 0.029 | 0.002 | 4.2e-35 |
| rs10757287 | 9 | 22143570 | T | A | 0.162 | -0.035 | 0.003 | 1.1e-33 |
| rs3793537 | 9 | 35687556 | C | G | 0.293 | 0.021 | 0.002 | 1.2e-18 |
| rs12343705 | 9 | 38197187 | T | A | 0.490 | -0.014 | 0.002 | 2e-10 |
| rs611461 | 9 | 79325615 | G | A | 0.759 | -0.018 | 0.002 | 1.5e-13 |
| rs696825 | 9 | 86583076 | T | C | 0.252 | -0.025 | 0.002 | 4.1e-25 |
| rs1288649 | 9 | 91426717 | G | T | 0.613 | -0.021 | 0.002 | 3.7e-21 |
| rs10992433 | 9 | 95483503 | G | A | 0.231 | 0.017 | 0.003 | 4.6e-11 |
| rs1930249 | 9 | 112735377 | G | A | 0.172 | 0.022 | 0.003 | 3e-15 |
| rs1411424 | 9 | 113892963 | A | G | 0.523 | -0.028 | 0.002 | 1.7e-38 |
| rs7851073 | 9 | 114654460 | T | C | 0.309 | 0.017 | 0.002 | 4.1e-13 |
| rs113702392 | 9 | 116039908 | T | C | 0.226 | -0.017 | 0.003 | 7.5e-11 |
| rs10760127 | 9 | 123692940 | C | G | 0.569 | 0.014 | 0.002 | 1.2e-10 |
| rs10986338 | 9 | 127191232 | A | G | 0.651 | -0.018 | 0.002 | 2.7e-16 |
| rs2519093 | 9 | 136141870 | T | C | 0.184 | -0.044 | 0.003 | 6.7e-57 |
| rs2905064 | 9 | 136949558 | T | A | 0.484 | 0.015 | 0.002 | 1.2e-12 |
| rs12555241 | 9 | 139387676 | A | G | 0.278 | 0.032 | 0.002 | 1.8e-41 |
| rs9329341 | 10 | 13532447 | A | G | 0.580 | 0.017 | 0.002 | 1.3e-15 |
| rs10828725 | 10 | 25218243 | T | G | 0.369 | -0.050 | 0.002 | 1.8e-113 |
| rs61838426 | 10 | 26735804 | T | G | 0.323 | -0.016 | 0.002 | 5.2e-13 |
| rs2807742 | 10 | 28781367 | A | G | 0.770 | 0.034 | 0.003 | 1.2e-41 |
| rs2091084 | 10 | 30492267 | C | T | 0.653 | 0.017 | 0.002 | 1.3e-13 |
| rs72790861 | 10 | 44880207 | G | C | 0.308 | -0.026 | 0.002 | 1.1e-28 |
| rs61850684 | 10 | 50252936 | A | G | 0.235 | -0.024 | 0.003 | 2.3e-21 |
| rs4746522 | 10 | 64437684 | A | G | 0.277 | 0.015 | 0.002 | 1.2e-10 |
| rs7082470 | 10 | 65277026 | A | G | 0.471 | -0.029 | 0.002 | 3.4e-41 |
| rs10997821 | 10 | 69570691 | G | A | 0.104 | -0.023 | 0.003 | 2.3e-11 |
| rs3747869 | 10 | 73520632 | C | A | 0.901 | 0.038 | 0.004 | 2e-27 |
| rs7098414 | 10 | 82214586 | C | A | 0.732 | 0.015 | 0.002 | 5.8e-10 |
| rs1412445 | 10 | 91002804 | T | C | 0.338 | 0.019 | 0.002 | 9.1e-17 |
| rs11187838 | 10 | 96038686 | A | G | 0.434 | -0.019 | 0.002 | 9.8e-20 |
| rs61863767 | 10 | 99084426 | T | C | 0.597 | 0.046 | 0.002 | 4e-100 |
| rs11593526 | 10 | 104228017 | A | C | 0.539 | 0.016 | 0.002 | 2.3e-14 |
| rs2803606 | 10 | 113902827 | T | C | 0.246 | -0.015 | 0.002 | 5e-10 |
| rs180943 | 10 | 115719472 | G | C | 0.587 | -0.019 | 0.002 | 4.3e-18 |
| rs12784071 | 10 | 115854445 | T | C | 0.230 | 0.017 | 0.003 | 4.6e-12 |
| rs3781454 | 10 | 126348565 | A | G | 0.681 | 0.025 | 0.002 | 2.3e-28 |
| rs14408 | 11 | 308314 | C | T | 0.368 | 0.044 | 0.002 | 9.7e-89 |
| rs1468102 | 11 | 3004526 | G | C | 0.306 | 0.017 | 0.002 | 1.2e-13 |
| rs7949917 | 11 | 10475370 | A | G | 0.596 | 0.014 | 0.002 | 1.6e-10 |
| rs11022177 | 11 | 12133478 | G | C | 0.310 | -0.017 | 0.002 | 1.5e-13 |
| rs2049045 | 11 | 27694241 | C | G | 0.186 | 0.017 | 0.003 | 5.9e-10 |
| rs1227996 | 11 | 47965189 | G | C | 0.661 | 0.027 | 0.002 | 1.9e-32 |
| rs174549 | 11 | 61571382 | A | G | 0.309 | -0.027 | 0.002 | 1.7e-31 |
| rs10896064 | 11 | 65641033 | C | G | 0.532 | 0.019 | 0.002 | 2.3e-18 |
| rs4980661 | 11 | 69306579 | A | G | 0.527 | 0.017 | 0.002 | 3.5e-15 |
| rs12363256 | 11 | 100520680 | A | T | 0.327 | 0.018 | 0.002 | 5.6e-15 |
| rs228606 | 11 | 108087847 | T | G | 0.411 | 0.018 | 0.002 | 2.3e-16 |
| rs238914 | 11 | 113984109 | A | C | 0.396 | 0.029 | 0.002 | 5.3e-41 |
| rs148713124 | 11 | 118225200 | T | G | 0.009 | 0.107 | 0.011 | 9.9e-21 |
| rs71468750 | 11 | 120205560 | A | G | 0.413 | 0.014 | 0.002 | 2e-10 |
| rs8705 | 11 | 128328913 | A | G | 0.318 | -0.028 | 0.002 | 7.3e-35 |
| rs632887 | 12 | 3392351 | G | A | 0.410 | 0.014 | 0.002 | 3.9e-11 |
| rs7955734 | 12 | 4333159 | G | C | 0.210 | -0.028 | 0.003 | 2.1e-27 |
| rs2286599 | 12 | 6499533 | A | G | 0.142 | 0.037 | 0.003 | 3e-33 |
| rs2024385 | 12 | 12888438 | A | T | 0.428 | 0.017 | 0.002 | 1.1e-14 |
| rs7134738 | 12 | 26358170 | C | T | 0.351 | -0.014 | 0.002 | 1e-10 |
| rs739842 | 12 | 48202046 | C | T | 0.635 | -0.016 | 0.002 | 3.3e-12 |
| rs7978263 | 12 | 49118291 | T | C | 0.314 | -0.016 | 0.002 | 9.3e-12 |
| rs11169302 | 12 | 50578705 | G | T | 0.394 | 0.021 | 0.002 | 1.2e-22 |
| rs2452793 | 12 | 51539508 | A | G | 0.880 | -0.020 | 0.003 | 4.8e-10 |
| rs706809 | 12 | 52294257 | C | T | 0.775 | 0.022 | 0.003 | 4.3e-18 |
| rs1245035 | 12 | 64976049 | A | C | 0.629 | 0.015 | 0.002 | 3.4e-12 |
| rs971546 | 12 | 68591919 | G | T | 0.704 | 0.016 | 0.002 | 2e-11 |
| rs4761234 | 12 | 69732105 | C | T | 0.485 | 0.015 | 0.002 | 8.8e-13 |
| rs12424773 | 12 | 80033612 | T | C | 0.691 | -0.015 | 0.002 | 1.1e-10 |
| rs7487314 | 12 | 88836215 | T | G | 0.701 | -0.025 | 0.002 | 1.1e-27 |
| rs17041439 | 12 | 101873240 | C | A | 0.056 | 0.041 | 0.005 | 1.2e-18 |
| rs11835066 | 12 | 109844678 | C | A | 0.050 | 0.029 | 0.005 | 2.9e-09 |
| rs10774624 | 12 | 111833788 | A | G | 0.515 | -0.067 | 0.002 | 1e-200 |
| rs75468472 | 12 | 112363239 | C | T | 0.015 | 0.071 | 0.011 | 4.8e-10 |
| rs11064881 | 12 | 120146925 | A | G | 0.073 | -0.034 | 0.004 | 1.8e-16 |
| rs34944476 | 12 | 124006269 | T | C | 0.219 | 0.017 | 0.003 | 2.2e-11 |
| rs9863 | 12 | 124421453 | C | T | 0.327 | -0.020 | 0.002 | 9e-18 |
| rs76428106 | 13 | 28604007 | C | T | 0.013 | 0.167 | 0.010 | 2.2e-65 |
| rs138028125 | 13 | 28712689 | G | C | 0.035 | 0.056 | 0.006 | 5.6e-21 |
| rs9508005 | 13 | 28789794 | G | T | 0.095 | -0.023 | 0.004 | 1.6e-09 |
| rs7326825 | 13 | 50113450 | A | G | 0.707 | 0.022 | 0.002 | 8.7e-21 |
| rs706606 | 13 | 50833707 | C | T | 0.531 | 0.017 | 0.002 | 4.2e-15 |
| rs3812849 | 13 | 74701736 | C | A | 0.264 | 0.021 | 0.002 | 1.1e-17 |
| rs17785077 | 13 | 99635611 | A | G | 0.103 | 0.023 | 0.003 | 7.5e-11 |
| rs12874404 | 13 | 108993494 | G | A | 0.053 | -0.037 | 0.005 | 2.8e-15 |
| rs2260302 | 13 | 114150850 | A | G | 0.258 | 0.023 | 0.002 | 1.3e-21 |
| rs9590390 | 13 | 114920867 | A | G | 0.294 | 0.016 | 0.002 | 8.1e-12 |
| rs7159281 | 14 | 23756604 | G | A | 0.669 | -0.014 | 0.002 | 5.7e-10 |
| rs2038700 | 14 | 25461989 | C | T | 0.394 | 0.035 | 0.002 | 1.7e-56 |
| rs17386684 | 14 | 35602087 | G | T | 0.178 | 0.024 | 0.003 | 2.4e-17 |
| rs696 | 14 | 35871093 | T | C | 0.363 | 0.017 | 0.002 | 4.8e-14 |
| rs10138752 | 14 | 69179971 | T | C | 0.078 | -0.040 | 0.004 | 2.8e-24 |
| rs175714 | 14 | 75981856 | C | T | 0.577 | 0.013 | 0.002 | 1.1e-09 |
| rs2205190 | 14 | 77886723 | G | T | 0.521 | -0.014 | 0.002 | 2.6e-11 |
| rs9806027 | 14 | 81668707 | C | G | 0.554 | -0.018 | 0.002 | 6e-17 |
| rs117068593 | 14 | 93118229 | T | C | 0.189 | -0.029 | 0.003 | 1.6e-26 |
| rs36084521 | 14 | 93516398 | G | T | 0.111 | 0.023 | 0.003 | 9.2e-12 |
| rs10146962 | 14 | 101170540 | C | T | 0.335 | -0.016 | 0.002 | 1.1e-12 |
| rs56227024 | 14 | 103861706 | G | A | 0.283 | -0.019 | 0.002 | 2.3e-16 |
| rs4924450 | 15 | 40597229 | A | G | 0.704 | 0.016 | 0.002 | 1.5e-11 |
| rs72726027 | 15 | 42248826 | C | T | 0.111 | -0.045 | 0.003 | 4e-41 |
| rs60695341 | 15 | 51010271 | T | C | 0.197 | -0.030 | 0.003 | 5.6e-29 |
| rs398474 | 15 | 62799035 | A | G | 0.273 | 0.016 | 0.002 | 1.9e-11 |
| rs7180079 | 15 | 64629873 | G | A | 0.879 | 0.036 | 0.003 | 9.2e-28 |
| rs743580 | 15 | 74328116 | G | A | 0.490 | 0.014 | 0.002 | 1.3e-10 |
| rs67538199 | 15 | 75361650 | C | T | 0.161 | -0.020 | 0.003 | 1.2e-11 |
| rs8024737 | 15 | 77249703 | C | T | 0.542 | 0.013 | 0.002 | 3.9e-09 |
| rs2562751 | 15 | 80251294 | A | G | 0.760 | 0.015 | 0.003 | 1.8e-09 |
| rs7177338 | 15 | 91428636 | A | G | 0.525 | -0.017 | 0.002 | 4.2e-15 |
| rs28576226 | 15 | 101713668 | A | G | 0.124 | 0.034 | 0.003 | 2.8e-26 |
| rs75893715 | 16 | 1360091 | T | C | 0.271 | 0.018 | 0.002 | 1.2e-13 |
| rs35929659 | 16 | 2165630 | C | T | 0.180 | -0.016 | 0.003 | 3.7e-09 |
| rs6500550 | 16 | 3746241 | T | C | 0.302 | -0.022 | 0.002 | 4.1e-22 |
| rs17199009 | 16 | 3867081 | G | A | 0.070 | 0.027 | 0.004 | 9.4e-11 |
| rs61739285 | 16 | 27480797 | T | C | 0.034 | -0.035 | 0.006 | 1.3e-09 |
| rs11574938 | 16 | 30485393 | C | G | 0.517 | 0.040 | 0.002 | 1.5e-79 |
| rs28853644 | 16 | 30801027 | T | C | 0.270 | -0.019 | 0.002 | 7.6e-16 |
| rs2080506 | 16 | 50164672 | T | C | 0.156 | -0.018 | 0.003 | 5.9e-10 |
| rs11644125 | 16 | 57058974 | T | C | 0.600 | -0.018 | 0.002 | 8.4e-17 |
| rs12918121 | 16 | 67187795 | T | C | 0.018 | -0.051 | 0.008 | 3.2e-10 |
| rs117784871 | 16 | 68138091 | T | G | 0.057 | 0.029 | 0.005 | 1.7e-09 |
| rs11642655 | 16 | 81610480 | A | G | 0.565 | 0.015 | 0.002 | 8.2e-13 |
| rs247826 | 16 | 84582965 | T | C | 0.222 | 0.035 | 0.003 | 2.9e-41 |
| rs9939427 | 16 | 86016091 | A | G | 0.231 | 0.022 | 0.003 | 7.7e-18 |
| rs59404182 | 16 | 88852866 | G | A | 0.199 | -0.020 | 0.003 | 8.7e-14 |
| rs2229075 | 17 | 1359363 | G | T | 0.722 | 0.016 | 0.002 | 2e-11 |
| rs7225843 | 17 | 2001825 | C | T | 0.203 | -0.033 | 0.003 | 7.1e-37 |
| rs62089694 | 17 | 2723868 | G | C | 0.161 | 0.021 | 0.003 | 5.2e-13 |
| rs8071086 | 17 | 4612866 | G | A | 0.294 | 0.015 | 0.002 | 5.9e-11 |
| rs9896414 | 17 | 5015895 | T | C | 0.509 | -0.018 | 0.002 | 7.1e-17 |
| rs1029809 | 17 | 16156235 | G | A | 0.475 | -0.026 | 0.002 | 1.6e-33 |
| rs55767800 | 17 | 17674967 | C | T | 0.635 | -0.020 | 0.002 | 2.2e-19 |
| rs8070966 | 17 | 27196748 | C | T | 0.844 | -0.021 | 0.003 | 2.3e-13 |
| rs2259855 | 17 | 28097860 | C | T | 0.526 | -0.026 | 0.002 | 1.4e-33 |
| rs79730542 | 17 | 37298789 | C | T | 0.031 | -0.037 | 0.006 | 4.4e-09 |
| rs11651596 | 17 | 38056116 | C | T | 0.468 | -0.065 | 0.002 | 1e-200 |
| rs576566496 | 17 | 38140269 | A | G | 0.027 | 0.085 | 0.007 | 1.1e-34 |
| rs74725931 | 17 | 38196327 | C | T | 0.038 | 0.058 | 0.006 | 3.6e-24 |
| rs563136944 | 17 | 38470429 | G | T | 0.356 | -0.023 | 0.003 | 1e-19 |
| rs4796657 | 17 | 40589978 | A | C | 0.573 | -0.014 | 0.002 | 4.3e-11 |
| rs138284624 | 17 | 56385573 | T | C | 0.013 | 0.110 | 0.009 | 3.4e-31 |
| rs8081395 | 17 | 57801761 | A | G | 0.545 | 0.038 | 0.002 | 3e-70 |
| rs2084312 | 17 | 72695211 | T | C | 0.800 | 0.029 | 0.003 | 6.8e-27 |
| rs12939873 | 17 | 74460148 | C | T | 0.657 | -0.013 | 0.002 | 3.9e-09 |
| rs8079218 | 17 | 76247744 | G | A | 0.498 | -0.015 | 0.002 | 2.3e-12 |
| rs9747839 | 17 | 81084522 | G | C | 0.478 | 0.020 | 0.002 | 2.8e-17 |
| rs4468717 | 18 | 3457606 | T | C | 0.077 | -0.024 | 0.004 | 1.7e-09 |
| rs303753 | 18 | 21074922 | A | G | 0.346 | -0.021 | 0.002 | 4.7e-20 |
| rs16978075 | 18 | 42113151 | C | T | 0.110 | 0.027 | 0.003 | 4.4e-15 |
| rs55874505 | 18 | 43816804 | G | T | 0.584 | -0.019 | 0.002 | 3.4e-18 |
| rs9947760 | 18 | 48148741 | C | T | 0.372 | 0.021 | 0.002 | 1.6e-21 |
| rs527544 | 18 | 51762866 | T | G | 0.424 | 0.016 | 0.002 | 4.4e-13 |
| rs72933640 | 18 | 60054077 | C | G | 0.107 | -0.021 | 0.003 | 1.8e-09 |
| rs17758695 | 18 | 60920854 | T | C | 0.029 | -0.062 | 0.006 | 6.6e-23 |
| rs763361 | 18 | 67531642 | C | T | 0.527 | -0.013 | 0.002 | 3.2e-10 |
| rs72973711 | 18 | 74072245 | T | A | 0.067 | -0.033 | 0.004 | 1.7e-14 |
| rs4807440 | 19 | 1026477 | T | G | 0.638 | 0.026 | 0.002 | 3.4e-30 |
| rs873289 | 19 | 1175396 | G | A | 0.474 | -0.013 | 0.002 | 2.7e-10 |
| rs2358581 | 19 | 10391611 | G | T | 0.734 | -0.029 | 0.002 | 1.2e-33 |
| rs895819 | 19 | 13947292 | C | T | 0.337 | 0.020 | 0.002 | 6.7e-18 |
| rs59326120 | 19 | 16504352 | C | T | 0.156 | 0.054 | 0.003 | 1.5e-75 |
| rs57070010 | 19 | 17650608 | C | G | 0.112 | 0.022 | 0.003 | 3.2e-11 |
| rs4805881 | 19 | 33896432 | C | A | 0.665 | -0.016 | 0.002 | 4.3e-13 |
| rs2194067 | 19 | 35464727 | T | C | 0.417 | 0.014 | 0.002 | 9.8e-11 |
| rs4760 | 19 | 44153100 | G | A | 0.156 | -0.070 | 0.003 | 5.8e-127 |
| rs11673093 | 19 | 45742094 | A | G | 0.260 | -0.054 | 0.002 | 1.4e-108 |
| rs177918 | 19 | 47634313 | G | T | 0.894 | -0.032 | 0.003 | 2e-20 |
| rs35112940 | 19 | 51738917 | A | G | 0.218 | -0.024 | 0.003 | 1.8e-20 |
| rs6045615 | 20 | 1931582 | C | A | 0.328 | -0.018 | 0.002 | 3.4e-15 |
| rs2294256 | 20 | 8174058 | T | A | 0.482 | -0.015 | 0.002 | 4.3e-13 |
| rs6086540 | 20 | 8603950 | C | G | 0.507 | -0.023 | 0.002 | 1e-27 |
| rs34952318 | 20 | 11177055 | A | G | 0.049 | -0.034 | 0.005 | 5e-12 |
| rs2185558 | 20 | 36905894 | C | T | 0.140 | 0.019 | 0.003 | 1.5e-09 |
| rs6029234 | 20 | 39259278 | C | G | 0.626 | 0.025 | 0.002 | 2.5e-29 |
| rs1800961 | 20 | 43042364 | T | C | 0.031 | -0.042 | 0.006 | 5.1e-12 |
| rs143003731 | 20 | 47936696 | T | C | 0.008 | -0.074 | 0.013 | 4e-09 |
| rs1060402 | 20 | 49205095 | A | G | 0.518 | -0.017 | 0.002 | 5.1e-16 |
| rs1997577 | 21 | 16371102 | T | A | 0.155 | -0.019 | 0.003 | 2.5e-10 |
| rs9977672 | 21 | 40463283 | A | G | 0.258 | -0.021 | 0.002 | 2.5e-18 |
| rs73377344 | 21 | 47804358 | C | T | 0.554 | -0.017 | 0.002 | 1.8e-14 |
| rs740422 | 22 | 17619292 | T | C | 0.218 | 0.016 | 0.003 | 1.3e-09 |
| rs5746451 | 22 | 18126020 | C | T | 0.504 | 0.020 | 0.002 | 7.3e-22 |
| rs2240717 | 22 | 19969106 | G | A | 0.350 | 0.022 | 0.002 | 3.6e-22 |
| rs13055027 | 22 | 24641953 | T | G | 0.276 | -0.019 | 0.002 | 3.1e-15 |
| rs62227042 | 22 | 29632882 | T | A | 0.416 | -0.019 | 0.002 | 1.1e-17 |
| rs9680589 | 22 | 31853484 | A | G | 0.051 | -0.029 | 0.005 | 2.2e-09 |
| rs139402 | 22 | 39546145 | C | T | 0.438 | -0.022 | 0.002 | 1.3e-24 |
| rs47340 | 22 | 43562829 | T | G | 0.396 | 0.015 | 0.002 | 1.5e-12 |

**Table S3. 367 SNPs significantly associated with eosinophil count used IVs in forward MR analyses derived from Vuckovic D et al.** Chr: Chromosome; EA: Effect allele; NEA: Non-effect allele; EAF: Effect allele frequency.

| SNP | Chr | Pos | EA | NEA | EAF | Beta | SE | P-value |
| --- | --- | --- | --- | --- | --- | --- | --- | --- |
| rs12097268 | 1 | 2477358 | A | T | 0.482 | -0.013 | 0.002 | 3.2e-10 |
| rs301802 | 1 | 8497307 | A | T | 0.584 | 0.019 | 0.002 | 2.5e-18 |
| rs1294028 | 1 | 9364634 | G | A | 0.610 | 0.015 | 0.002 | 8.2e-12 |
| rs9430574 | 1 | 9709072 | A | G | 0.325 | -0.021 | 0.002 | 4.1e-20 |
| rs35249183 | 1 | 12099345 | G | A | 0.100 | 0.040 | 0.004 | 5.1e-29 |
| rs2502994 | 1 | 24199363 | C | T | 0.569 | 0.021 | 0.002 | 1.9e-22 |
| rs10794666 | 1 | 25250830 | T | C | 0.573 | 0.017 | 0.002 | 3e-16 |
| rs6699190 | 1 | 26531789 | T | G | 0.663 | -0.014 | 0.002 | 3.1e-10 |
| rs79516637 | 1 | 28608721 | A | G | 0.067 | -0.032 | 0.004 | 4.1e-14 |
| rs1004870 | 1 | 42370787 | T | C | 0.589 | 0.026 | 0.002 | 4e-34 |
| rs567910619 | 1 | 65210790 | T | C | 0.011 | -0.088 | 0.011 | 4.9e-16 |
| rs12408934 | 1 | 65423447 | A | G | 0.103 | -0.042 | 0.003 | 4.8e-33 |
| rs41313381 | 1 | 79411968 | A | C | 0.031 | 0.049 | 0.006 | 3e-16 |
| rs6672038 | 1 | 87751670 | T | C | 0.119 | 0.037 | 0.003 | 3.2e-29 |
| rs1088318 | 1 | 93733057 | T | C | 0.379 | -0.023 | 0.002 | 7e-26 |
| rs74431709 | 1 | 101653125 | G | T | 0.111 | -0.042 | 0.003 | 7.4e-35 |
| rs1494316 | 1 | 111319655 | C | G | 0.235 | -0.016 | 0.002 | 1.1e-10 |
| rs2793925 | 1 | 120652158 | A | T | 0.684 | 0.017 | 0.002 | 3.7e-13 |
| rs11204682 | 1 | 150595537 | T | G | 0.223 | -0.042 | 0.003 | 6.4e-60 |
| rs111548294 | 1 | 150911337 | T | C | 0.013 | 0.088 | 0.010 | 2e-20 |
| rs34448954 | 1 | 161168189 | T | C | 0.106 | -0.028 | 0.003 | 6.5e-16 |
| rs6672031 | 1 | 172796505 | G | C | 0.487 | 0.019 | 0.002 | 4.3e-19 |
| rs12137747 | 1 | 173213790 | A | C | 0.469 | 0.018 | 0.002 | 8.3e-17 |
| rs6658698 | 1 | 181060791 | A | G | 0.618 | -0.015 | 0.002 | 1.6e-11 |
| rs17849502 | 1 | 183532580 | T | G | 0.052 | -0.029 | 0.005 | 1e-09 |
| rs2296618 | 1 | 198666232 | G | A | 0.136 | -0.039 | 0.003 | 9.3e-37 |
| rs1036332 | 1 | 199012478 | C | A | 0.738 | -0.032 | 0.002 | 8e-41 |
| rs10793767 | 1 | 204549564 | C | A | 0.675 | 0.014 | 0.002 | 6.6e-10 |
| rs41298997 | 1 | 206655331 | T | C | 0.235 | 0.016 | 0.002 | 4.8e-11 |
| rs59150958 | 1 | 221106304 | T | C | 0.153 | -0.027 | 0.003 | 1.2e-19 |
| rs12142635 | 1 | 226976332 | A | C | 0.057 | -0.030 | 0.005 | 2e-10 |
| rs11810108 | 1 | 229668671 | C | T | 0.229 | -0.017 | 0.003 | 1.8e-11 |
| rs72755295 | 1 | 242034263 | G | A | 0.033 | -0.037 | 0.006 | 3.2e-10 |
| rs10165200 | 2 | 8439629 | A | G | 0.371 | -0.025 | 0.002 | 5.2e-30 |
| rs62105489 | 2 | 8757326 | T | C | 0.053 | -0.032 | 0.005 | 1.3e-11 |
| rs11127154 | 2 | 28685967 | T | A | 0.492 | -0.024 | 0.002 | 9.8e-28 |
| rs11890306 | 2 | 28691488 | T | C | 0.423 | 0.013 | 0.002 | 2.3e-09 |
| rs17682575 | 2 | 43152304 | T | C | 0.202 | -0.019 | 0.003 | 2.2e-12 |
| rs113542380 | 2 | 43464818 | A | G | 0.075 | -0.028 | 0.004 | 6.2e-12 |
| rs778756 | 2 | 61781994 | G | A | 0.580 | -0.016 | 0.002 | 8.4e-14 |
| rs7569084 | 2 | 65656969 | T | C | 0.585 | 0.019 | 0.002 | 1.7e-18 |
| rs12622600 | 2 | 71279940 | T | C | 0.270 | -0.016 | 0.002 | 4.6e-11 |
| rs11126424 | 2 | 74262353 | C | T | 0.306 | -0.015 | 0.002 | 3.4e-11 |
| rs2366640 | 2 | 85810734 | C | A | 0.319 | -0.016 | 0.002 | 1.6e-12 |
| rs2579505 | 2 | 97167962 | C | T | 0.638 | 0.044 | 0.002 | 1.5e-90 |
| rs13418548 | 2 | 102917239 | T | C | 0.138 | -0.093 | 0.003 | 1e-200 |
| rs1465641 | 2 | 106438936 | T | C | 0.385 | 0.015 | 0.002 | 1.3e-11 |
| rs72836346 | 2 | 111876613 | C | G | 0.078 | 0.071 | 0.004 | 1.2e-68 |
| rs4848139 | 2 | 112418002 | T | C | 0.525 | 0.018 | 0.002 | 3.5e-18 |
| rs11678022 | 2 | 136809033 | T | A | 0.731 | -0.019 | 0.002 | 1.4e-14 |
| rs79716587 | 2 | 143886819 | A | G | 0.126 | -0.025 | 0.003 | 5.5e-15 |
| rs1427499 | 2 | 145400317 | G | A | 0.710 | 0.019 | 0.002 | 1.1e-15 |
| rs10195713 | 2 | 158520905 | T | C | 0.864 | 0.028 | 0.003 | 1.5e-18 |
| rs10930337 | 2 | 169695031 | T | C | 0.285 | 0.014 | 0.002 | 4e-09 |
| rs6731125 | 2 | 182308836 | C | T | 0.565 | 0.018 | 0.002 | 1.9e-16 |
| rs10174238 | 2 | 191973034 | A | G | 0.766 | 0.015 | 0.003 | 3.5e-09 |
| rs1519602 | 2 | 197027792 | G | T | 0.649 | 0.015 | 0.002 | 5.8e-11 |
| rs13021247 | 2 | 207926818 | A | G | 0.315 | -0.014 | 0.002 | 1.4e-09 |
| rs715 | 2 | 211543055 | C | T | 0.311 | 0.018 | 0.002 | 4.3e-15 |
| rs12619285 | 2 | 213824045 | G | A | 0.265 | -0.064 | 0.002 | 1.2e-156 |
| rs35409523 | 2 | 213908457 | A | G | 0.076 | 0.052 | 0.004 | 7.4e-38 |
| rs7423615 | 2 | 231116874 | T | C | 0.187 | 0.023 | 0.003 | 5.5e-18 |
| rs1057258 | 2 | 234115629 | T | C | 0.178 | -0.032 | 0.003 | 5.4e-31 |
| rs34290285 | 2 | 242698640 | A | G | 0.256 | -0.046 | 0.002 | 8.8e-80 |
| rs76474320 | 2 | 242806022 | C | T | 0.078 | 0.031 | 0.004 | 3.5e-15 |
| rs1695315 | 3 | 3153339 | A | G | 0.605 | -0.036 | 0.002 | 9.3e-60 |
| rs4684037 | 3 | 10228040 | G | C | 0.143 | -0.020 | 0.003 | 6.3e-11 |
| rs310747 | 3 | 12259545 | G | T | 0.625 | -0.030 | 0.002 | 9.9e-42 |
| rs76643909 | 3 | 12441367 | G | T | 0.150 | 0.027 | 0.003 | 5.4e-20 |
| rs13073683 | 3 | 25393837 | C | T | 0.399 | 0.016 | 0.002 | 3.6e-13 |
| rs1353286 | 3 | 27772014 | G | T | 0.453 | 0.020 | 0.002 | 2.1e-21 |
| rs7646283 | 3 | 33046480 | T | C | 0.367 | 0.026 | 0.002 | 8.2e-32 |
| rs2228467 | 3 | 42906116 | C | T | 0.061 | 0.062 | 0.004 | 1.7e-45 |
| rs4072859 | 3 | 49032205 | C | G | 0.659 | 0.027 | 0.002 | 9.3e-30 |
| rs4677079 | 3 | 72125863 | T | G | 0.498 | 0.014 | 0.002 | 1.7e-11 |
| rs7649812 | 3 | 98449191 | G | C | 0.242 | 0.017 | 0.002 | 3.1e-12 |
| rs4618204 | 3 | 101281534 | C | T | 0.444 | 0.018 | 0.002 | 4.3e-17 |
| rs528288236 | 3 | 111807413 | A | G | 0.019 | -0.047 | 0.008 | 2.3e-09 |
| rs9868582 | 3 | 112052927 | T | G | 0.452 | -0.024 | 0.002 | 2e-29 |
| rs2399441 | 3 | 112587012 | C | T | 0.350 | -0.019 | 0.002 | 2.8e-18 |
| rs6806253 | 3 | 128282697 | G | A | 0.164 | -0.088 | 0.003 | 1e-200 |
| rs142062847 | 3 | 128395208 | C | T | 0.437 | 0.018 | 0.002 | 6.3e-14 |
| rs2712399 | 3 | 128432953 | G | C | 0.409 | -0.025 | 0.002 | 6e-30 |
| rs73238201 | 3 | 142204728 | T | C | 0.179 | -0.017 | 0.003 | 3.9e-10 |
| rs1516527 | 3 | 148609704 | C | T | 0.951 | -0.033 | 0.005 | 2.6e-11 |
| rs4680250 | 3 | 150941492 | G | C | 0.694 | -0.016 | 0.002 | 1.9e-11 |
| rs76830965 | 3 | 159637678 | A | C | 0.118 | -0.023 | 0.003 | 7.3e-12 |
| rs191674933 | 3 | 169488691 | T | A | 0.240 | 0.025 | 0.002 | 1.3e-23 |
| rs4464488 | 3 | 171498211 | T | C | 0.414 | -0.014 | 0.002 | 8.9e-11 |
| rs4074672 | 3 | 183730295 | T | C | 0.369 | 0.015 | 0.002 | 2.8e-12 |
| rs9815874 | 3 | 188441161 | T | C | 0.299 | 0.029 | 0.002 | 3.7e-35 |
| rs9818987 | 3 | 194402430 | T | C | 0.306 | 0.014 | 0.002 | 2.9e-09 |
| rs12152276 | 3 | 196368501 | G | A | 0.081 | -0.035 | 0.004 | 4.8e-20 |
| rs4916555 | 3 | 196538648 | T | C | 0.475 | 0.018 | 0.002 | 9.3e-18 |
| rs1828803 | 4 | 2689449 | A | C | 0.393 | 0.013 | 0.002 | 1.2e-09 |
| rs6448432 | 4 | 26098810 | A | G | 0.306 | 0.014 | 0.002 | 1.8e-09 |
| rs73232881 | 4 | 38664131 | C | T | 0.214 | 0.069 | 0.003 | 1.3e-156 |
| rs6855379 | 4 | 39037026 | G | C | 0.584 | -0.016 | 0.002 | 6e-13 |
| rs2412771 | 4 | 57761417 | C | T | 0.417 | -0.013 | 0.002 | 1.3e-09 |
| rs17005891 | 4 | 83547862 | A | G | 0.184 | -0.055 | 0.003 | 2.9e-90 |
| rs13105682 | 4 | 102702364 | G | T | 0.059 | -0.029 | 0.005 | 3.4e-10 |
| rs113473633 | 4 | 103449131 | G | A | 0.026 | -0.063 | 0.007 | 1.5e-19 |
| rs62324212 | 4 | 123560939 | A | C | 0.437 | 0.021 | 0.002 | 5.4e-22 |
| rs6836610 | 4 | 123601697 | A | G | 0.303 | 0.019 | 0.002 | 6.3e-17 |
| rs13120371 | 4 | 139092719 | G | A | 0.327 | 0.016 | 0.002 | 3.4e-13 |
| rs11931711 | 4 | 153291513 | T | C | 0.282 | 0.021 | 0.002 | 1.6e-18 |
| rs10062687 | 5 | 10624866 | G | T | 0.233 | 0.025 | 0.003 | 2e-22 |
| rs16903574 | 5 | 14610309 | G | C | 0.076 | 0.030 | 0.004 | 1.7e-13 |
| rs4594881 | 5 | 35846815 | T | G | 0.341 | -0.036 | 0.002 | 8.1e-58 |
| rs4703589 | 5 | 72097351 | C | T | 0.531 | 0.013 | 0.002 | 2.1e-10 |
| rs4703730 | 5 | 76549688 | T | C | 0.517 | -0.017 | 0.002 | 4.6e-16 |
| rs34495 | 5 | 98265807 | T | G | 0.304 | -0.019 | 0.002 | 6e-17 |
| rs10455025 | 5 | 110404999 | C | A | 0.355 | 0.041 | 0.002 | 8.6e-75 |
| rs35936927 | 5 | 110768362 | G | A | 0.522 | -0.018 | 0.002 | 2.4e-14 |
| rs17516457 | 5 | 131590387 | C | T | 0.417 | -0.065 | 0.002 | 1e-200 |
| rs375509059 | 5 | 131960939 | T | C | 0.046 | 0.045 | 0.005 | 2.2e-18 |
| rs256869 | 5 | 132135372 | C | T | 0.085 | 0.023 | 0.004 | 1.8e-09 |
| rs17653687 | 5 | 133468000 | G | A | 0.179 | -0.019 | 0.003 | 1.3e-11 |
| rs10062349 | 5 | 141509597 | A | G | 0.617 | 0.040 | 0.002 | 1.9e-76 |
| rs55977204 | 5 | 142501095 | C | T | 0.113 | 0.021 | 0.003 | 3.5e-10 |
| rs56330463 | 5 | 148200011 | C | T | 0.552 | 0.041 | 0.002 | 3.3e-81 |
| rs6869605 | 5 | 150452866 | C | A | 0.123 | -0.024 | 0.003 | 3.6e-14 |
| rs2431097 | 5 | 159890885 | T | C | 0.485 | 0.014 | 0.002 | 5.9e-11 |
| rs62385501 | 5 | 171950231 | A | T | 0.308 | -0.015 | 0.002 | 1.5e-10 |
| rs6556313 | 5 | 176792491 | G | A | 0.332 | 0.020 | 0.002 | 3.8e-18 |
| rs2666969 | 6 | 336410 | G | A | 0.254 | -0.042 | 0.003 | 2.5e-38 |
| rs9504361 | 6 | 577820 | G | A | 0.446 | -0.032 | 0.002 | 9.4e-49 |
| rs186984319 | 6 | 16702251 | C | T | 0.317 | -0.014 | 0.002 | 4.2e-09 |
| rs3132682 | 6 | 30044388 | C | G | 0.588 | -0.055 | 0.002 | 5.1e-144 |
| rs9266321 | 6 | 31330360 | C | G | 0.184 | 0.048 | 0.003 | 9.6e-69 |
| rs111936749 | 6 | 31523012 | T | C | 0.252 | 0.041 | 0.003 | 1.7e-34 |
| rs62395789 | 6 | 31572801 | A | G | 0.044 | 0.042 | 0.005 | 8.2e-16 |
| rs529454848 | 6 | 32525948 | A | T | 0.146 | -0.036 | 0.004 | 4.7e-22 |
| rs28383314 | 6 | 32587213 | C | T | 0.625 | 0.063 | 0.002 | 5e-184 |
| rs9462094 | 6 | 35513603 | T | C | 0.431 | -0.023 | 0.002 | 3.4e-20 |
| rs11751949 | 6 | 42219296 | A | G | 0.071 | -0.031 | 0.004 | 1e-13 |
| rs76236447 | 6 | 42225993 | C | G | 0.072 | 0.039 | 0.004 | 5.1e-21 |
| rs9395112 | 6 | 45682806 | G | A | 0.156 | 0.020 | 0.003 | 7.9e-12 |
| rs1095671 | 6 | 88016300 | A | G | 0.571 | -0.015 | 0.002 | 6.1e-13 |
| rs62408224 | 6 | 90955995 | G | A | 0.351 | -0.041 | 0.002 | 1.3e-77 |
| rs62420764 | 6 | 106799970 | C | T | 0.140 | 0.019 | 0.003 | 8.2e-10 |
| rs12208103 | 6 | 107442431 | T | C | 0.376 | -0.031 | 0.002 | 1.8e-44 |
| rs1915834 | 6 | 109597624 | A | C | 0.480 | 0.014 | 0.002 | 3.6e-11 |
| rs9389268 | 6 | 135419631 | G | A | 0.260 | -0.047 | 0.002 | 1.1e-82 |
| rs6924387 | 6 | 137082948 | G | A | 0.410 | 0.017 | 0.002 | 1.8e-15 |
| rs113496608 | 6 | 138161666 | A | G | 0.030 | -0.043 | 0.006 | 6.7e-12 |
| rs13207791 | 6 | 143249749 | G | A | 0.089 | 0.024 | 0.004 | 2.6e-10 |
| rs149110519 | 6 | 144385777 | T | C | 0.036 | 0.042 | 0.006 | 4.7e-13 |
| rs577734383 | 7 | 2881732 | A | G | 0.236 | 0.016 | 0.003 | 4.3e-10 |
| rs6971710 | 7 | 3140173 | A | G | 0.201 | 0.030 | 0.003 | 3.1e-29 |
| rs13245462 | 7 | 8074608 | T | C | 0.102 | -0.021 | 0.004 | 3e-09 |
| rs10950642 | 7 | 17016646 | A | G | 0.363 | -0.019 | 0.002 | 3.5e-18 |
| rs57834782 | 7 | 20502828 | A | T | 0.245 | -0.059 | 0.002 | 2.6e-125 |
| rs12537614 | 7 | 22789551 | C | G | 0.592 | -0.028 | 0.002 | 1.4e-37 |
| rs60600003 | 7 | 37382465 | G | T | 0.101 | 0.041 | 0.004 | 1.3e-31 |
| rs150640087 | 7 | 50444152 | T | G | 0.016 | 0.099 | 0.009 | 5e-31 |
| rs17450294 | 7 | 50763459 | C | T | 0.086 | -0.031 | 0.004 | 5e-16 |
| rs13226583 | 7 | 75454152 | T | A | 0.116 | -0.062 | 0.003 | 5.8e-78 |
| rs55879743 | 7 | 75470364 | T | C | 0.066 | 0.074 | 0.004 | 3.6e-65 |
| rs62473720 | 7 | 77254547 | G | A | 0.318 | 0.014 | 0.002 | 6.3e-10 |
| rs4272 | 7 | 92236829 | G | A | 0.211 | 0.028 | 0.003 | 8.7e-28 |
| rs445 | 7 | 92408370 | T | C | 0.096 | -0.053 | 0.004 | 7.5e-49 |
| rs56072276 | 7 | 98778222 | A | G | 0.688 | -0.016 | 0.002 | 5.6e-12 |
| rs12538988 | 7 | 101810359 | A | C | 0.291 | 0.018 | 0.002 | 4.2e-14 |
| rs12705849 | 7 | 112782556 | A | G | 0.407 | -0.021 | 0.002 | 4.7e-23 |
| rs3807307 | 7 | 128579202 | C | T | 0.464 | -0.016 | 0.002 | 1e-14 |
| rs56179563 | 7 | 129685597 | A | G | 0.389 | 0.018 | 0.002 | 6.7e-16 |
| rs3110791 | 7 | 135367828 | C | T | 0.624 | 0.015 | 0.002 | 7.6e-12 |
| rs12530946 | 7 | 148887942 | G | A | 0.614 | 0.043 | 0.002 | 6.1e-85 |
| rs4871849 | 8 | 22964719 | A | G | 0.708 | -0.033 | 0.002 | 1.7e-45 |
| rs17377462 | 8 | 27281912 | A | G | 0.248 | -0.017 | 0.002 | 1.5e-11 |
| rs2133480 | 8 | 47923945 | A | C | 0.094 | 0.029 | 0.004 | 3.8e-15 |
| rs45577137 | 8 | 48651633 | G | A | 0.045 | -0.061 | 0.005 | 5.2e-29 |
| rs7846314 | 8 | 61650831 | T | A | 0.187 | -0.033 | 0.003 | 1.4e-34 |
| rs6989099 | 8 | 66898262 | C | T | 0.317 | -0.018 | 0.002 | 5.6e-15 |
| rs4739140 | 8 | 79753010 | T | C | 0.610 | -0.016 | 0.002 | 2.6e-13 |
| rs10957978 | 8 | 81285139 | T | G | 0.647 | -0.016 | 0.002 | 1e-12 |
| rs2011566 | 8 | 95971921 | A | G | 0.512 | 0.013 | 0.002 | 1.1e-09 |
| rs12681644 | 8 | 119972974 | T | C | 0.211 | -0.021 | 0.003 | 3.3e-15 |
| rs4870977 | 8 | 127526842 | C | G | 0.871 | -0.019 | 0.003 | 8.9e-10 |
| rs11786536 | 8 | 129000416 | A | G | 0.164 | -0.028 | 0.003 | 1.9e-22 |
| rs7840212 | 8 | 130599247 | T | C | 0.336 | -0.040 | 0.002 | 1.8e-70 |
| rs4236746 | 8 | 130699861 | G | A | 0.975 | 0.056 | 0.007 | 2.3e-16 |
| rs117961539 | 8 | 144989466 | A | G | 0.039 | -0.033 | 0.006 | 3.9e-09 |
| rs34173062 | 8 | 145158607 | A | G | 0.072 | 0.054 | 0.004 | 5.8e-35 |
| rs460631 | 9 | 4851440 | G | A | 0.885 | 0.027 | 0.003 | 6.6e-16 |
| rs62541534 | 9 | 5028921 | G | C | 0.259 | 0.032 | 0.002 | 1.5e-40 |
| rs4142528 | 9 | 6172296 | A | T | 0.672 | -0.076 | 0.002 | 1e-200 |
| rs1547258 | 9 | 6523056 | C | T | 0.708 | -0.020 | 0.002 | 1.3e-17 |
| rs10962679 | 9 | 16905441 | C | T | 0.755 | -0.015 | 0.002 | 4.1e-10 |
| rs7036656 | 9 | 21990457 | T | C | 0.722 | 0.018 | 0.002 | 9.3e-15 |
| rs9410887 | 9 | 86465395 | C | T | 0.253 | 0.022 | 0.002 | 3e-19 |
| rs1331691 | 9 | 94129137 | C | T | 0.184 | -0.030 | 0.003 | 2.7e-27 |
| rs496475 | 9 | 113638236 | G | T | 0.387 | 0.035 | 0.002 | 1.5e-58 |
| rs7040707 | 9 | 117680732 | G | A | 0.048 | -0.034 | 0.005 | 5.6e-10 |
| rs911603 | 9 | 117697584 | A | C | 0.403 | -0.025 | 0.002 | 2.9e-29 |
| rs2773818 | 9 | 135859519 | A | C | 0.447 | 0.014 | 0.002 | 5.2e-11 |
| rs74612091 | 9 | 135877278 | A | T | 0.063 | 0.060 | 0.004 | 4.9e-43 |
| rs115478735 | 9 | 136149711 | T | A | 0.184 | -0.028 | 0.003 | 1.5e-24 |
| rs72766638 | 9 | 136931778 | A | C | 0.164 | -0.025 | 0.003 | 2.8e-18 |
| rs72775768 | 9 | 139324574 | T | C | 0.289 | 0.022 | 0.002 | 2.5e-20 |
| rs11253517 | 10 | 970524 | A | G | 0.238 | 0.038 | 0.002 | 1.2e-51 |
| rs1323650 | 10 | 6033049 | T | G | 0.681 | -0.014 | 0.002 | 1.8e-09 |
| rs10905281 | 10 | 8109583 | A | C | 0.176 | 0.021 | 0.003 | 2.9e-14 |
| rs2646438 | 10 | 8565675 | A | G | 0.566 | -0.021 | 0.002 | 3.6e-22 |
| rs1444782 | 10 | 9058671 | A | G | 0.423 | -0.040 | 0.002 | 2.1e-79 |
| rs2992335 | 10 | 26727934 | G | C | 0.598 | -0.035 | 0.002 | 1.1e-57 |
| rs2807742 | 10 | 28781367 | A | G | 0.770 | 0.038 | 0.003 | 8.7e-51 |
| rs17482472 | 10 | 44859618 | A | G | 0.099 | -0.034 | 0.004 | 3.5e-21 |
| rs16917546 | 10 | 64397538 | C | T | 0.369 | -0.045 | 0.002 | 5.5e-93 |
| rs72834751 | 10 | 64559465 | T | C | 0.013 | -0.069 | 0.010 | 6.2e-12 |
| rs10998535 | 10 | 70805373 | T | C | 0.215 | 0.016 | 0.003 | 1.2e-09 |
| rs564443 | 10 | 89807917 | A | G | 0.163 | 0.024 | 0.003 | 1.2e-16 |
| rs7897422 | 10 | 90849704 | C | T | 0.191 | 0.021 | 0.003 | 7.1e-15 |
| rs7918084 | 10 | 94429467 | T | C | 0.550 | 0.030 | 0.002 | 4.7e-45 |
| rs11591571 | 10 | 104342804 | A | G | 0.339 | 0.016 | 0.002 | 4.1e-13 |
| rs17126931 | 10 | 111754633 | C | T | 0.129 | -0.022 | 0.003 | 1e-11 |
| rs7080536 | 10 | 115348046 | A | G | 0.043 | -0.046 | 0.005 | 4.4e-18 |
| rs1059091 | 11 | 309127 | G | A | 0.321 | 0.034 | 0.002 | 9.8e-51 |
| rs9666598 | 11 | 325386 | G | C | 0.843 | -0.021 | 0.003 | 4.2e-13 |
| rs58833930 | 11 | 2325997 | T | C | 0.113 | -0.024 | 0.003 | 2.5e-12 |
| rs34439695 | 11 | 33901483 | T | C | 0.035 | -0.041 | 0.006 | 3.4e-12 |
| rs3824867 | 11 | 47468569 | G | A | 0.712 | 0.018 | 0.002 | 1.2e-14 |
| rs3016175 | 11 | 60830367 | G | A | 0.881 | 0.021 | 0.003 | 4.7e-10 |
| rs174548 | 11 | 61571348 | G | C | 0.314 | -0.023 | 0.002 | 9e-24 |
| rs689274 | 11 | 65665988 | G | A | 0.543 | 0.033 | 0.002 | 4e-54 |
| rs7936323 | 11 | 76293758 | A | G | 0.478 | 0.041 | 0.002 | 3.2e-83 |
| rs11236813 | 11 | 76343427 | C | G | 0.101 | -0.031 | 0.004 | 5.6e-18 |
| rs4409785 | 11 | 95311422 | C | T | 0.172 | 0.018 | 0.003 | 6.2e-11 |
| rs637064 | 11 | 108140909 | T | C | 0.555 | 0.025 | 0.002 | 4.4e-32 |
| rs964184 | 11 | 116648917 | C | G | 0.867 | 0.028 | 0.003 | 7.5e-20 |
| rs7123726 | 11 | 118694547 | C | T | 0.210 | 0.019 | 0.003 | 2.2e-13 |
| rs10893844 | 11 | 128185850 | C | G | 0.502 | 0.024 | 0.002 | 4.5e-30 |
| rs605093 | 11 | 128604232 | T | G | 0.473 | -0.017 | 0.002 | 5.9e-16 |
| rs12820863 | 12 | 4318723 | T | C | 0.351 | 0.022 | 0.002 | 1.2e-22 |
| rs1800692 | 12 | 6442346 | G | A | 0.587 | -0.019 | 0.002 | 8e-19 |
| rs3759332 | 12 | 6491078 | C | T | 0.391 | -0.013 | 0.002 | 3.8e-09 |
| rs7302975 | 12 | 32138924 | C | T | 0.779 | -0.023 | 0.003 | 7.3e-19 |
| rs12581511 | 12 | 46573788 | G | C | 0.172 | -0.018 | 0.003 | 1.6e-10 |
| rs10876550 | 12 | 54712308 | A | G | 0.559 | 0.014 | 0.002 | 1.4e-11 |
| rs1689510 | 12 | 56396768 | C | G | 0.338 | 0.027 | 0.002 | 6.7e-34 |
| rs3024971 | 12 | 57493727 | G | T | 0.107 | -0.039 | 0.003 | 6.7e-30 |
| rs10859277 | 12 | 92521198 | T | C | 0.536 | -0.024 | 0.002 | 1.4e-28 |
| rs12313790 | 12 | 94632933 | A | C | 0.085 | -0.025 | 0.004 | 1.1e-10 |
| rs1530161 | 12 | 96804649 | G | A | 0.433 | 0.013 | 0.002 | 1.1e-09 |
| rs73191842 | 12 | 110986507 | G | T | 0.022 | 0.057 | 0.007 | 2.3e-15 |
| rs1265564 | 12 | 111708458 | C | A | 0.441 | 0.082 | 0.002 | 1e-200 |
| rs182686482 | 12 | 112350744 | C | T | 0.015 | 0.104 | 0.011 | 1.9e-21 |
| rs373582220 | 12 | 112847812 | A | G | 0.372 | -0.041 | 0.003 | 7e-51 |
| rs1557865 | 12 | 113342598 | G | T | 0.817 | -0.017 | 0.003 | 1.8e-09 |
| rs1169286 | 12 | 121419056 | C | T | 0.426 | -0.013 | 0.002 | 7.2e-10 |
| rs28866344 | 12 | 123885484 | G | A | 0.907 | -0.028 | 0.004 | 1.2e-14 |
| rs12861824 | 13 | 31079184 | C | T | 0.273 | -0.020 | 0.003 | 3.2e-15 |
| rs7986796 | 13 | 40351064 | T | G | 0.627 | 0.025 | 0.002 | 5e-30 |
| rs34373380 | 13 | 41244217 | A | T | 0.305 | 0.037 | 0.002 | 1.3e-58 |
| rs9533117 | 13 | 43046812 | T | A | 0.239 | -0.019 | 0.002 | 1.8e-14 |
| rs201798 | 13 | 50954721 | A | G | 0.615 | 0.021 | 0.002 | 6.6e-22 |
| rs7327960 | 13 | 74682892 | C | T | 0.826 | 0.017 | 0.003 | 6.7e-10 |
| rs9557201 | 13 | 99989538 | G | A | 0.272 | 0.019 | 0.002 | 2.6e-15 |
| rs912131 | 13 | 100032346 | G | A | 0.702 | 0.027 | 0.002 | 4.3e-32 |
| rs9590475 | 13 | 114961483 | A | G | 0.413 | -0.016 | 0.002 | 1.4e-13 |
| rs10137343 | 14 | 23382640 | T | A | 0.604 | 0.013 | 0.002 | 3.4e-09 |
| rs2239633 | 14 | 23589057 | A | G | 0.484 | 0.035 | 0.002 | 7.9e-61 |
| rs2004925 | 14 | 25445362 | A | G | 0.393 | 0.016 | 0.002 | 2.1e-13 |
| rs8020739 | 14 | 35882492 | T | G | 0.643 | 0.025 | 0.002 | 2.6e-28 |
| rs12432697 | 14 | 38053334 | T | G | 0.254 | -0.016 | 0.002 | 1.3e-10 |
| rs6573020 | 14 | 55871452 | T | C | 0.434 | 0.021 | 0.002 | 1.3e-22 |
| rs7152277 | 14 | 65535391 | C | G | 0.399 | 0.016 | 0.002 | 3.8e-13 |
| rs1296535 | 14 | 69297986 | T | C | 0.779 | -0.018 | 0.003 | 1.7e-12 |
| rs1861160 | 14 | 73365712 | A | G | 0.317 | 0.015 | 0.002 | 2e-11 |
| rs175705 | 14 | 75975648 | G | C | 0.719 | 0.039 | 0.002 | 8.7e-61 |
| rs67856193 | 14 | 93024616 | G | C | 0.308 | 0.024 | 0.002 | 9.7e-26 |
| rs117068593 | 14 | 93118229 | T | C | 0.189 | -0.025 | 0.003 | 3e-20 |
| rs11555542 | 14 | 94417531 | C | T | 0.063 | 0.066 | 0.004 | 4.5e-51 |
| rs11628569 | 14 | 103190362 | G | A | 0.290 | -0.018 | 0.002 | 1.9e-14 |
| rs4906317 | 14 | 103814500 | T | G | 0.504 | -0.014 | 0.002 | 6.4e-10 |
| rs62006172 | 15 | 38904527 | A | G | 0.042 | -0.038 | 0.005 | 3.9e-13 |
| rs7173571 | 15 | 41712807 | C | T | 0.526 | 0.014 | 0.002 | 1.5e-10 |
| rs34212866 | 15 | 43702964 | G | A | 0.221 | 0.022 | 0.003 | 1e-17 |
| rs11638256 | 15 | 58772807 | C | T | 0.120 | 0.020 | 0.003 | 2.6e-09 |
| rs11637887 | 15 | 60765216 | T | C | 0.193 | -0.016 | 0.003 | 4.7e-09 |
| rs16943117 | 15 | 61078836 | T | C | 0.192 | -0.018 | 0.003 | 5.9e-11 |
| rs3101486 | 15 | 67346220 | T | G | 0.765 | 0.018 | 0.003 | 2.8e-13 |
| rs17293632 | 15 | 67442596 | T | C | 0.236 | 0.030 | 0.003 | 3e-33 |
| rs7498094 | 15 | 68486097 | T | C | 0.519 | -0.013 | 0.002 | 1.6e-09 |
| rs6494871 | 15 | 70767978 | T | C | 0.595 | 0.015 | 0.002 | 3.1e-12 |
| rs2115535 | 15 | 80195199 | A | G | 0.568 | 0.034 | 0.002 | 4.3e-56 |
| rs1138358 | 15 | 80263345 | C | A | 0.264 | -0.030 | 0.002 | 1.7e-36 |
| rs13511 | 15 | 86284817 | C | T | 0.577 | -0.017 | 0.002 | 1.7e-14 |
| rs5011652 | 15 | 91166608 | C | G | 0.759 | 0.020 | 0.002 | 2.9e-15 |
| rs4785903 | 16 | 2838274 | C | T | 0.755 | -0.034 | 0.002 | 7.1e-43 |
| rs9934736 | 16 | 3519252 | G | A | 0.047 | 0.031 | 0.005 | 6.8e-10 |
| rs7186106 | 16 | 11210831 | A | G | 0.354 | -0.047 | 0.002 | 5.2e-97 |
| rs2072130 | 16 | 27356398 | T | C | 0.365 | 0.030 | 0.002 | 3.7e-41 |
| rs7201518 | 16 | 30082508 | T | G | 0.480 | -0.030 | 0.002 | 5.3e-45 |
| rs17881535 | 16 | 31108709 | G | C | 0.240 | -0.017 | 0.002 | 2.5e-12 |
| rs9924483 | 16 | 48646258 | T | C | 0.310 | -0.023 | 0.002 | 7.3e-24 |
| rs1684578 | 16 | 57069650 | T | G | 0.416 | 0.017 | 0.002 | 1.5e-14 |
| rs2161647 | 16 | 57503330 | A | C | 0.037 | -0.041 | 0.006 | 2.1e-13 |
| rs12918121 | 16 | 67187795 | T | C | 0.018 | -0.047 | 0.008 | 4.4e-09 |
| rs570765844 | 16 | 67618243 | C | T | 0.080 | 0.026 | 0.004 | 8.4e-11 |
| rs1170439 | 16 | 68608511 | C | T | 0.779 | 0.025 | 0.003 | 4.4e-23 |
| rs13339407 | 16 | 78156295 | G | A | 0.306 | -0.014 | 0.002 | 2.1e-09 |
| rs301161 | 16 | 85810349 | A | G | 0.850 | 0.027 | 0.003 | 7e-20 |
| rs12932970 | 16 | 86004110 | C | T | 0.352 | 0.016 | 0.002 | 2.6e-13 |
| rs17175830 | 16 | 88558164 | A | G | 0.237 | 0.033 | 0.003 | 2.8e-39 |
| rs112036266 | 17 | 2834143 | T | C | 0.176 | 0.017 | 0.003 | 4.4e-10 |
| rs34210653 | 17 | 4535314 | A | G | 0.021 | -0.176 | 0.007 | 1.6e-124 |
| rs74480102 | 17 | 7742601 | A | G | 0.043 | -0.084 | 0.005 | 1.1e-57 |
| rs12941068 | 17 | 17737857 | A | G | 0.285 | -0.015 | 0.002 | 2e-10 |
| rs58745116 | 17 | 33803592 | A | G | 0.392 | -0.015 | 0.002 | 2.1e-11 |
| rs79989390 | 17 | 37962845 | A | G | 0.034 | -0.072 | 0.006 | 4.4e-35 |
| rs72836561 | 17 | 41926126 | T | C | 0.031 | -0.054 | 0.006 | 4.2e-19 |
| rs1971374 | 17 | 43659738 | C | T | 0.213 | -0.033 | 0.003 | 6.4e-38 |
| rs12952581 | 17 | 47448346 | A | G | 0.362 | 0.029 | 0.002 | 8.1e-39 |
| rs4968392 | 17 | 57799204 | G | A | 0.841 | 0.020 | 0.003 | 9.4e-12 |
| rs7215391 | 17 | 64232022 | T | C | 0.257 | -0.014 | 0.002 | 4.9e-09 |
| rs62086903 | 17 | 66016006 | C | T | 0.231 | 0.030 | 0.003 | 1.7e-32 |
| rs180506 | 17 | 68274205 | A | G | 0.776 | -0.023 | 0.003 | 4e-20 |
| rs78057960 | 17 | 73779075 | T | C | 0.296 | -0.018 | 0.002 | 8.7e-15 |
| rs12946146 | 17 | 81079703 | T | C | 0.486 | 0.016 | 0.002 | 1.2e-12 |
| rs2847266 | 18 | 12773338 | T | C | 0.717 | -0.015 | 0.002 | 5.9e-10 |
| rs12968697 | 18 | 20625431 | T | C | 0.627 | 0.017 | 0.002 | 2.4e-15 |
| rs9955853 | 18 | 43829655 | T | C | 0.559 | -0.013 | 0.002 | 4.9e-09 |
| rs57633475 | 18 | 46469650 | G | A | 0.122 | -0.026 | 0.003 | 1.1e-15 |
| rs637716 | 18 | 48481308 | A | G | 0.589 | -0.013 | 0.002 | 8.9e-10 |
| rs73963711 | 18 | 60874413 | T | C | 0.210 | 0.023 | 0.003 | 9.4e-18 |
| rs954954 | 18 | 60902328 | C | A | 0.105 | -0.035 | 0.003 | 1.7e-23 |
| rs17758695 | 18 | 60920854 | T | C | 0.029 | -0.121 | 0.006 | 1.7e-82 |
| rs1395269 | 18 | 61377644 | G | T | 0.301 | -0.024 | 0.002 | 1.1e-25 |
| rs3848640 | 19 | 1037986 | G | A | 0.329 | -0.016 | 0.002 | 1.3e-11 |
| rs36084354 | 19 | 1079959 | A | G | 0.092 | -0.044 | 0.004 | 1.1e-33 |
| rs3746165 | 19 | 1102211 | G | A | 0.469 | -0.014 | 0.002 | 5.4e-11 |
| rs61731111 | 19 | 3179517 | T | C | 0.011 | -0.131 | 0.010 | 1.5e-37 |
| rs34466956 | 19 | 3353622 | T | C | 0.568 | -0.013 | 0.002 | 2.2e-09 |
| rs410867 | 19 | 16427111 | G | A | 0.216 | -0.061 | 0.003 | 2.2e-122 |
| rs3786586 | 19 | 16495287 | G | A | 0.155 | 0.032 | 0.003 | 6.9e-27 |
| rs8108623 | 19 | 18408519 | A | C | 0.626 | 0.018 | 0.002 | 4.9e-14 |
| rs2043293 | 19 | 19751456 | A | C | 0.826 | -0.017 | 0.003 | 2.3e-09 |
| rs117710327 | 19 | 33726578 | A | C | 0.067 | -0.036 | 0.004 | 9.1e-17 |
| rs412884 | 19 | 40219449 | C | T | 0.672 | 0.057 | 0.002 | 1.1e-140 |
| rs62117160 | 19 | 45232161 | A | G | 0.045 | -0.048 | 0.005 | 1.1e-20 |
| rs73036520 | 19 | 45749484 | C | G | 0.254 | -0.031 | 0.002 | 7.9e-36 |
| rs113577760 | 19 | 46365177 | C | T | 0.091 | -0.040 | 0.004 | 3.3e-27 |
| rs6077868 | 20 | 10649761 | T | C | 0.206 | -0.016 | 0.003 | 5.9e-10 |
| rs3790315 | 20 | 17630045 | T | C | 0.580 | -0.018 | 0.002 | 9e-17 |
| rs80054178 | 20 | 30294682 | C | T | 0.023 | -0.084 | 0.007 | 1e-31 |
| rs6141755 | 20 | 31163565 | T | G | 0.236 | -0.016 | 0.003 | 8e-11 |
| rs6017247 | 20 | 42657841 | C | T | 0.729 | -0.025 | 0.002 | 1.5e-24 |
| rs6013210 | 20 | 50165634 | G | C | 0.118 | -0.023 | 0.003 | 1.9e-12 |
| rs7264802 | 20 | 62692440 | G | A | 0.748 | -0.016 | 0.002 | 2.9e-10 |
| rs2223043 | 21 | 16438793 | G | A | 0.305 | 0.020 | 0.002 | 9.1e-17 |
| rs11701443 | 21 | 16931215 | C | T | 0.202 | 0.017 | 0.003 | 3.7e-10 |
| rs1000005 | 21 | 34433051 | C | G | 0.591 | -0.020 | 0.002 | 3.1e-20 |
| rs34872427 | 21 | 36294044 | C | T | 0.415 | -0.020 | 0.002 | 7.7e-21 |
| rs2242886 | 21 | 36387806 | T | C | 0.071 | -0.102 | 0.004 | 1.4e-132 |
| rs9979383 | 21 | 36715761 | T | C | 0.630 | 0.030 | 0.002 | 1.1e-42 |
| rs150229150 | 21 | 45641853 | T | C | 0.125 | -0.021 | 0.003 | 5.1e-11 |
| rs11703539 | 22 | 17594755 | A | G | 0.102 | -0.022 | 0.003 | 4.3e-10 |
| rs5747308 | 22 | 18133500 | C | A | 0.505 | 0.015 | 0.002 | 1.2e-12 |
| rs7288670 | 22 | 24621826 | G | A | 0.306 | -0.025 | 0.002 | 2.6e-28 |
| rs140074 | 22 | 31725634 | G | C | 0.486 | 0.023 | 0.002 | 2.4e-26 |
| rs41280005 | 22 | 37316133 | A | G | 0.167 | -0.029 | 0.003 | 1e-23 |
| rs228951 | 22 | 37529798 | G | A | 0.363 | -0.022 | 0.002 | 3.7e-22 |
| rs2413631 | 22 | 41359786 | A | G | 0.250 | 0.015 | 0.002 | 1.6e-09 |
| rs34780507 | 22 | 41405753 | G | A | 0.066 | -0.059 | 0.004 | 1.4e-43 |
| rs78023664 | 22 | 46647844 | A | G | 0.092 | -0.027 | 0.004 | 5.3e-13 |

**Table S4. 388 SNPs significantly associated with eosinophil count used IVs in forward MR analyses derived from Chen MH et al.** Chr: Chromosome; EA: Effect allele; NEA: Non-effect allele; EAF: Effect allele frequency.

| SNP | Chr | Pos | EA | NEA | EAF | Beta | SE | P-value |
| --- | --- | --- | --- | --- | --- | --- | --- | --- |
| rs2419313 | 10 | 111750851 | A | G | 0.143 | -0.021 | 0.003 | 5.21e-13 |
| rs7080536 | 10 | 115348046 | A | G | 0.043 | -0.044 | 0.005 | 1.17e-18 |
| rs2992333 | 10 | 26727454 | A | G | 0.599 | -0.034 | 0.002 | 1.35e-63 |
| rs2807740 | 10 | 28784483 | T | C | 0.770 | 0.035 | 0.002 | 1.05e-50 |
| rs17482472 | 10 | 44859618 | A | G | 0.099 | -0.031 | 0.003 | 5.34e-21 |
| rs1323650 | 10 | 6033049 | T | G | 0.681 | -0.014 | 0.002 | 2.53e-10 |
| rs10995240 | 10 | 64388631 | C | G | 0.368 | -0.044 | 0.002 | 1.55e-101 |
| rs72834751 | 10 | 64559465 | T | C | 0.013 | -0.072 | 0.009 | 1.5e-14 |
| rs3747869 | 10 | 73520632 | C | A | 0.901 | 0.020 | 0.003 | 2.31e-09 |
| rs4746153 | 10 | 75598282 | C | G | 0.185 | 0.016 | 0.003 | 8.58e-10 |
| rs11255507 | 10 | 8109615 | G | T | 0.178 | 0.020 | 0.003 | 1.05e-14 |
| rs2646438 | 10 | 8565675 | A | G | 0.566 | -0.021 | 0.002 | 1.2e-24 |
| rs495149 | 10 | 89795523 | T | C | 0.164 | 0.024 | 0.003 | 1.15e-19 |
| rs962993 | 10 | 9053132 | T | C | 0.422 | -0.040 | 0.002 | 4.92e-90 |
| rs7897422 | 10 | 90849704 | C | T | 0.192 | 0.021 | 0.003 | 1.85e-16 |
| rs2497318 | 10 | 94432000 | T | C | 0.443 | -0.030 | 0.002 | 1.96e-49 |
| rs1539174 | 10 | 974870 | G | C | 0.237 | 0.038 | 0.002 | 1.12e-59 |
| rs637064 | 11 | 108140909 | T | C | 0.556 | 0.024 | 0.002 | 5.43e-34 |
| rs964184 | 11 | 116648917 | C | G | 0.867 | 0.029 | 0.003 | 4.39e-23 |
| rs10893844 | 11 | 128185850 | C | G | 0.502 | 0.025 | 0.002 | 3.49e-35 |
| rs668248 | 11 | 128586262 | C | G | 0.616 | -0.017 | 0.002 | 5.49e-16 |
| rs214080 | 11 | 17299762 | G | A | 0.580 | 0.013 | 0.002 | 1.37e-10 |
| rs58833930 | 11 | 2325997 | T | C | 0.113 | -0.023 | 0.003 | 1.03e-13 |
| rs1059091 | 11 | 309127 | G | A | 0.321 | 0.034 | 0.002 | 3.08e-56 |
| rs9666598 | 11 | 325386 | G | C | 0.843 | -0.020 | 0.003 | 1.36e-12 |
| rs34439695 | 11 | 33901483 | T | C | 0.035 | -0.038 | 0.005 | 3.67e-12 |
| rs80116434 | 11 | 57110622 | A | G | 0.142 | -0.020 | 0.003 | 9.09e-12 |
| rs174548 | 11 | 61571348 | G | C | 0.314 | -0.023 | 0.002 | 4.08e-27 |
| rs11231727 | 11 | 64011854 | T | C | 0.431 | -0.012 | 0.002 | 1.34e-09 |
| rs634534 | 11 | 65665256 | G | A | 0.542 | 0.033 | 0.002 | 5.83e-61 |
| rs7936434 | 11 | 76293805 | C | G | 0.478 | 0.043 | 0.002 | 4.08e-104 |
| rs11236814 | 11 | 76343428 | T | A | 0.102 | -0.030 | 0.003 | 3.46e-20 |
| rs4409785 | 11 | 95311422 | C | T | 0.172 | 0.016 | 0.003 | 4.25e-10 |
| rs11571404 | 12 | 1041450 | T | C | 0.203 | 0.015 | 0.002 | 5.57e-10 |
| rs73202462 | 12 | 110266430 | A | G | 0.020 | 0.057 | 0.008 | 2.14e-13 |
| rs11065822 | 12 | 111600134 | T | G | 0.354 | 0.065 | 0.002 | 1e-200 |
| rs115647629 | 12 | 111811903 | A | G | 0.026 | -0.067 | 0.007 | 6.69e-25 |
| rs6490291 | 12 | 112177775 | A | T | 0.964 | -0.066 | 0.006 | 1.11e-29 |
| rs146730870 | 12 | 112332996 | A | G | 0.010 | -0.087 | 0.010 | 4.95e-17 |
| rs148219449 | 12 | 113339873 | A | G | 0.014 | 0.064 | 0.010 | 6.02e-11 |
| rs28532037 | 12 | 123883406 | A | G | 0.907 | -0.030 | 0.003 | 4.19e-18 |
| rs4931002 | 12 | 32143169 | A | C | 0.779 | -0.022 | 0.002 | 1.73e-19 |
| rs12820863 | 12 | 4318723 | T | C | 0.352 | 0.022 | 0.002 | 1.07e-25 |
| rs12581511 | 12 | 46573788 | G | C | 0.173 | -0.018 | 0.003 | 2.07e-11 |
| rs1689510 | 12 | 56396768 | C | G | 0.338 | 0.027 | 0.002 | 1.06e-36 |
| rs3024971 | 12 | 57493727 | G | T | 0.107 | -0.040 | 0.003 | 5.04e-36 |
| rs1800692 | 12 | 6442346 | G | A | 0.588 | -0.018 | 0.002 | 8.92e-19 |
| rs10777378 | 12 | 92518680 | A | G | 0.536 | -0.022 | 0.002 | 1.28e-29 |
| rs10745763 | 12 | 96811386 | T | G | 0.423 | 0.013 | 0.002 | 2.41e-11 |
| rs12100034 | 13 | 114956773 | A | G | 0.359 | -0.017 | 0.002 | 2.6e-16 |
| rs12861824 | 13 | 31079184 | C | T | 0.275 | -0.021 | 0.002 | 2.59e-18 |
| rs7986796 | 13 | 40351064 | T | G | 0.628 | 0.024 | 0.002 | 4.91e-32 |
| rs6563842 | 13 | 41244297 | T | G | 0.306 | 0.036 | 0.002 | 3.39e-61 |
| rs71429414 | 13 | 42945821 | A | G | 0.196 | -0.019 | 0.002 | 1.03e-13 |
| rs201798 | 13 | 50954721 | A | G | 0.615 | 0.020 | 0.002 | 5.71e-23 |
| rs7327960 | 13 | 74682892 | C | T | 0.827 | 0.018 | 0.003 | 1.91e-11 |
| rs2182885 | 13 | 99855124 | A | G | 0.605 | 0.026 | 0.002 | 2.05e-38 |
| rs11628569 | 14 | 103190362 | G | A | 0.290 | -0.017 | 0.002 | 4.7e-14 |
| rs2239633 | 14 | 23589057 | A | G | 0.484 | 0.035 | 0.002 | 1.5e-68 |
| rs7141943 | 14 | 25438250 | G | A | 0.395 | 0.016 | 0.002 | 7e-15 |
| rs8020739 | 14 | 35882492 | T | G | 0.644 | 0.025 | 0.002 | 2.57e-32 |
| rs8012643 | 14 | 37692864 | T | C | 0.286 | 0.015 | 0.002 | 1.91e-12 |
| rs6573020 | 14 | 55871452 | T | C | 0.433 | 0.021 | 0.002 | 2.31e-24 |
| rs8011585 | 14 | 65522223 | A | T | 0.397 | 0.016 | 0.002 | 1.5e-14 |
| rs1296535 | 14 | 69297986 | T | C | 0.777 | -0.019 | 0.002 | 3.35e-15 |
| rs12878610 | 14 | 73389048 | C | T | 0.476 | -0.013 | 0.002 | 3.95e-11 |
| rs175705 | 14 | 75975648 | G | C | 0.719 | 0.038 | 0.002 | 8.8e-67 |
| rs67856193 | 14 | 93024616 | G | C | 0.308 | 0.024 | 0.002 | 1.63e-27 |
| rs117068593 | 14 | 93118229 | T | C | 0.190 | -0.026 | 0.003 | 1.17e-24 |
| rs11555542 | 14 | 94417531 | C | T | 0.063 | 0.064 | 0.004 | 2.12e-54 |
| rs62006172 | 15 | 38904527 | A | G | 0.042 | -0.038 | 0.005 | 4.62e-14 |
| rs34212866 | 15 | 43702964 | G | A | 0.221 | 0.020 | 0.002 | 5.74e-17 |
| rs11071559 | 15 | 61069988 | T | C | 0.131 | -0.022 | 0.003 | 4.94e-14 |
| rs62011287 | 15 | 63791228 | G | A | 0.344 | -0.013 | 0.002 | 1.32e-09 |
| rs17293632 | 15 | 67442596 | T | C | 0.236 | 0.030 | 0.002 | 8.92e-37 |
| rs7257 | 15 | 80191343 | A | G | 0.567 | 0.034 | 0.002 | 3.6e-63 |
| rs8026803 | 15 | 80260014 | C | T | 0.261 | -0.031 | 0.002 | 1.67e-43 |
| rs13511 | 15 | 86284817 | C | T | 0.577 | -0.016 | 0.002 | 4.13e-16 |
| rs725613 | 16 | 11169683 | G | T | 0.356 | -0.047 | 0.002 | 4.78e-116 |
| rs121564 | 16 | 20717360 | T | G | 0.270 | -0.017 | 0.002 | 1.26e-13 |
| rs3785356 | 16 | 27349168 | T | C | 0.298 | 0.030 | 0.002 | 1.7e-43 |
| rs4280242 | 16 | 2830482 | T | C | 0.754 | -0.033 | 0.002 | 1.03e-45 |
| rs9939774 | 16 | 30068354 | T | C | 0.405 | -0.029 | 0.002 | 1.98e-47 |
| rs1039341 | 16 | 48574869 | T | C | 0.311 | -0.024 | 0.002 | 8.79e-29 |
| rs1684578 | 16 | 57069650 | T | G | 0.416 | 0.016 | 0.002 | 5.15e-16 |
| rs16956811 | 16 | 57444002 | G | T | 0.081 | -0.029 | 0.004 | 1.75e-15 |
| rs8050508 | 16 | 67303358 | T | C | 0.031 | 0.035 | 0.006 | 1.01e-09 |
| rs1170439 | 16 | 68608511 | C | T | 0.779 | 0.023 | 0.002 | 7.86e-21 |
| rs8044920 | 16 | 69838676 | T | C | 0.376 | 0.012 | 0.002 | 3.59e-09 |
| rs3850107 | 16 | 78150341 | G | C | 0.294 | -0.014 | 0.002 | 3.94e-10 |
| rs17689159 | 16 | 79742390 | C | T | 0.315 | -0.014 | 0.002 | 1.4e-11 |
| rs301162 | 16 | 85810647 | G | A | 0.853 | 0.026 | 0.003 | 1.16e-20 |
| rs17175830 | 16 | 88558164 | A | G | 0.238 | 0.032 | 0.002 | 4.13e-42 |
| rs61426394 | 16 | 9053519 | C | G | 0.067 | -0.025 | 0.004 | 6.99e-10 |
| rs12941068 | 17 | 17737857 | A | G | 0.286 | -0.014 | 0.002 | 9.59e-10 |
| rs112036266 | 17 | 2834143 | T | C | 0.176 | 0.017 | 0.003 | 1.48e-10 |
| rs58745116 | 17 | 33803592 | A | G | 0.391 | -0.015 | 0.002 | 8.21e-14 |
| rs79989390 | 17 | 37962845 | A | G | 0.035 | -0.069 | 0.005 | 3.28e-37 |
| rs145947882 | 17 | 41809207 | C | A | 0.026 | -0.060 | 0.006 | 2.98e-21 |
| rs34210653 | 17 | 4535314 | A | G | 0.021 | -0.176 | 0.007 | 8.11e-140 |
| rs9889262 | 17 | 47398070 | A | T | 0.364 | 0.028 | 0.002 | 4.18e-43 |
| rs72831272 | 17 | 57448441 | A | G | 0.029 | 0.035 | 0.006 | 3.16e-09 |
| rs11869228 | 17 | 57800358 | T | C | 0.841 | 0.021 | 0.003 | 3.45e-14 |
| rs62086903 | 17 | 66016006 | C | T | 0.232 | 0.030 | 0.002 | 2.27e-37 |
| rs180506 | 17 | 68274205 | A | G | 0.776 | -0.023 | 0.002 | 5.82e-22 |
| rs55868524 | 17 | 7170665 | A | G | 0.606 | 0.013 | 0.002 | 9.91e-11 |
| rs8076052 | 17 | 73779198 | C | A | 0.296 | -0.018 | 0.002 | 4.33e-17 |
| rs74480102 | 17 | 7742601 | A | G | 0.043 | -0.083 | 0.005 | 2.52e-63 |
| rs9709216 | 17 | 81081455 | G | T | 0.487 | 0.018 | 0.002 | 1.21e-16 |
| rs397187 | 17 | 8784759 | C | T | 0.430 | -0.012 | 0.002 | 1.28e-09 |
| rs2847266 | 18 | 12773338 | T | C | 0.717 | -0.014 | 0.002 | 1.22e-10 |
| rs9675999 | 18 | 20627691 | A | G | 0.627 | 0.016 | 0.002 | 1.59e-15 |
| rs57633475 | 18 | 46469650 | G | A | 0.122 | -0.025 | 0.003 | 6.36e-16 |
| rs2850542 | 18 | 48403560 | T | G | 0.556 | -0.013 | 0.002 | 6.5e-11 |
| rs73963711 | 18 | 60874413 | T | C | 0.211 | 0.022 | 0.002 | 3.28e-19 |
| rs954954 | 18 | 60902328 | C | A | 0.105 | -0.035 | 0.003 | 3.57e-27 |
| rs17758695 | 18 | 60920854 | T | C | 0.030 | -0.122 | 0.006 | 1.32e-93 |
| rs7241187 | 18 | 61375974 | C | T | 0.301 | -0.024 | 0.002 | 9.38e-28 |
| rs36084354 | 19 | 1079959 | A | G | 0.092 | -0.045 | 0.003 | 1.17e-39 |
| rs4588110 | 19 | 1102139 | G | A | 0.469 | -0.015 | 0.002 | 3.37e-13 |
| rs410867 | 19 | 16427111 | G | A | 0.217 | -0.059 | 0.002 | 4.12e-134 |
| rs3786586 | 19 | 16495287 | G | A | 0.155 | 0.032 | 0.003 | 4.42e-31 |
| rs8108623 | 19 | 18408519 | A | C | 0.625 | 0.018 | 0.002 | 1.91e-16 |
| rs1529745 | 19 | 19517325 | G | C | 0.175 | 0.016 | 0.003 | 2.9e-09 |
| rs61731111 | 19 | 3179517 | T | C | 0.011 | -0.132 | 0.010 | 7.57e-43 |
| rs34466956 | 19 | 3353622 | T | C | 0.568 | -0.014 | 0.002 | 2.6e-11 |
| rs118013485 | 19 | 33726577 | A | G | 0.067 | -0.034 | 0.004 | 1.54e-16 |
| rs412884 | 19 | 40219449 | C | T | 0.672 | 0.058 | 0.002 | 2.22e-164 |
| rs62117160 | 19 | 45232161 | A | G | 0.045 | -0.048 | 0.005 | 2.15e-23 |
| rs117955557 | 19 | 45755368 | T | G | 0.256 | -0.030 | 0.002 | 1.44e-40 |
| rs76793172 | 19 | 46359794 | T | C | 0.092 | -0.042 | 0.003 | 8.07e-34 |
| rs2967592 | 19 | 8598280 | C | T | 0.122 | -0.018 | 0.003 | 3.29e-09 |
| rs111759324 | 1 | 101652522 | T | C | 0.111 | -0.043 | 0.003 | 3.56e-42 |
| rs1160413 | 1 | 111315694 | G | A | 0.240 | -0.015 | 0.002 | 2.97e-10 |
| rs35249183 | 1 | 12099345 | G | A | 0.100 | 0.040 | 0.003 | 1.46e-32 |
| rs11204682 | 1 | 150595537 | T | G | 0.223 | -0.041 | 0.002 | 4.89e-67 |
| rs71628184 | 1 | 160596935 | T | C | 0.094 | 0.021 | 0.003 | 8.32e-10 |
| rs34448954 | 1 | 161168189 | T | C | 0.106 | -0.025 | 0.003 | 3.87e-15 |
| rs6672031 | 1 | 172796505 | G | C | 0.486 | 0.020 | 0.002 | 1.9e-23 |
| rs1099448 | 1 | 173231898 | T | C | 0.466 | 0.018 | 0.002 | 8.32e-19 |
| rs4652560 | 1 | 181057188 | T | A | 0.618 | -0.015 | 0.002 | 5.23e-14 |
| rs17849502 | 1 | 183532580 | T | G | 0.052 | -0.027 | 0.004 | 1.29e-09 |
| rs17668272 | 1 | 198623842 | T | G | 0.118 | -0.042 | 0.003 | 7.48e-42 |
| rs1036332 | 1 | 199012478 | C | A | 0.737 | -0.033 | 0.002 | 1.61e-47 |
| rs1414517 | 1 | 221092838 | G | C | 0.187 | -0.025 | 0.003 | 1.36e-22 |
| rs708776 | 1 | 226923505 | T | G | 0.941 | 0.027 | 0.004 | 2.91e-10 |
| rs4148757 | 1 | 229693846 | C | A | 0.210 | -0.017 | 0.002 | 2.24e-12 |
| rs3218148 | 1 | 23851787 | A | G | 0.540 | -0.018 | 0.002 | 7.08e-19 |
| rs2502995 | 1 | 24199290 | C | T | 0.570 | 0.022 | 0.002 | 1.06e-27 |
| rs4149909 | 1 | 242023898 | G | A | 0.033 | -0.033 | 0.006 | 2.07e-09 |
| rs12097268 | 1 | 2477358 | A | T | 0.482 | -0.013 | 0.002 | 3.23e-11 |
| rs113105190 | 1 | 28614481 | C | T | 0.068 | -0.033 | 0.004 | 5.19e-17 |
| rs4310436 | 1 | 39489221 | A | G | 0.120 | 0.019 | 0.003 | 9.15e-10 |
| rs1004870 | 1 | 42370787 | T | C | 0.589 | 0.026 | 0.002 | 1.27e-38 |
| rs77625297 | 1 | 65362766 | C | G | 0.035 | -0.046 | 0.006 | 2.79e-16 |
| rs12408934 | 1 | 65423447 | A | G | 0.103 | -0.041 | 0.003 | 2.36e-36 |
| rs41313381 | 1 | 79411968 | A | C | 0.031 | 0.050 | 0.006 | 7.05e-19 |
| rs159963 | 1 | 8504421 | A | C | 0.582 | 0.019 | 0.002 | 9.88e-22 |
| rs6684992 | 1 | 87752000 | T | A | 0.119 | 0.038 | 0.003 | 6.06e-35 |
| rs4908835 | 1 | 9357460 | C | T | 0.161 | 0.019 | 0.003 | 1.8e-12 |
| rs661126 | 1 | 93678968 | G | T | 0.624 | 0.023 | 0.002 | 9.02e-29 |
| rs4529711 | 1 | 9709735 | C | T | 0.320 | -0.020 | 0.002 | 3.7e-20 |
| rs3790163 | 20 | 10647951 | G | A | 0.792 | -0.015 | 0.003 | 1.35e-09 |
| rs6080761 | 20 | 17629162 | A | G | 0.419 | 0.018 | 0.002 | 4.75e-20 |
| rs80054178 | 20 | 30294682 | C | T | 0.022 | -0.082 | 0.007 | 8.57e-34 |
| rs6141755 | 20 | 31163565 | T | G | 0.236 | -0.017 | 0.002 | 8.3e-13 |
| rs6103572 | 20 | 42657862 | C | T | 0.729 | -0.024 | 0.002 | 5.96e-27 |
| rs3746420 | 20 | 50140627 | C | G | 0.061 | -0.032 | 0.004 | 2.26e-14 |
| rs2223043 | 21 | 16438793 | G | A | 0.307 | 0.019 | 0.002 | 3.23e-17 |
| rs11701475 | 21 | 16931347 | C | T | 0.223 | 0.015 | 0.002 | 1.06e-09 |
| rs11088236 | 21 | 34416187 | T | C | 0.453 | 0.018 | 0.002 | 5.17e-20 |
| rs28421324 | 21 | 36286938 | T | A | 0.102 | 0.036 | 0.003 | 1.57e-27 |
| rs56117721 | 21 | 36398586 | A | T | 0.072 | -0.104 | 0.004 | 6.64e-160 |
| rs114152720 | 21 | 36434894 | A | G | 0.030 | -0.036 | 0.006 | 8.08e-10 |
| rs9979383 | 21 | 36715761 | T | C | 0.630 | 0.028 | 0.002 | 8e-43 |
| rs2838317 | 21 | 44991791 | C | A | 0.550 | 0.013 | 0.002 | 1.2e-10 |
| rs7354779 | 21 | 45670770 | C | T | 0.271 | 0.018 | 0.002 | 2.71e-15 |
| rs11703539 | 22 | 17594755 | A | G | 0.102 | -0.022 | 0.003 | 2.82e-11 |
| rs5747308 | 22 | 18133500 | C | A | 0.505 | 0.016 | 0.002 | 4.82e-15 |
| rs7288670 | 22 | 24621826 | G | A | 0.306 | -0.026 | 0.002 | 4.51e-34 |
| rs739427 | 22 | 31659101 | C | G | 0.512 | -0.022 | 0.002 | 1.06e-29 |
| rs60175411 | 22 | 37311858 | A | G | 0.167 | -0.030 | 0.003 | 1.25e-28 |
| rs743002 | 22 | 41404939 | C | T | 0.066 | -0.059 | 0.004 | 2.9e-48 |
| rs73176685 | 22 | 41781094 | G | C | 0.243 | 0.024 | 0.002 | 1.07e-24 |
| rs8142080 | 22 | 46647429 | T | G | 0.102 | -0.024 | 0.003 | 5.72e-13 |
| rs72998585 | 2 | 102858490 | T | A | 0.134 | -0.091 | 0.003 | 1e-200 |
| rs6736701 | 2 | 106390440 | T | G | 0.409 | 0.013 | 0.002 | 1.98e-10 |
| rs72836346 | 2 | 111876613 | C | G | 0.079 | 0.070 | 0.004 | 4.38e-76 |
| rs4848139 | 2 | 112418002 | T | C | 0.525 | 0.019 | 0.002 | 4.82e-21 |
| rs925966 | 2 | 136806959 | G | C | 0.759 | -0.018 | 0.002 | 1.32e-14 |
| rs79716587 | 2 | 143886819 | A | G | 0.127 | -0.024 | 0.003 | 3.99e-16 |
| rs1427499 | 2 | 145400317 | G | A | 0.710 | 0.019 | 0.002 | 1.3e-17 |
| rs1406449 | 2 | 148495285 | C | T | 0.462 | -0.012 | 0.002 | 8.69e-10 |
| rs10195713 | 2 | 158520905 | T | C | 0.864 | 0.025 | 0.003 | 2.74e-17 |
| rs10930337 | 2 | 169695031 | T | C | 0.285 | 0.015 | 0.002 | 2.03e-11 |
| rs6731125 | 2 | 182308836 | C | T | 0.565 | 0.017 | 0.002 | 1.04e-17 |
| rs1519602 | 2 | 197027792 | G | T | 0.648 | 0.013 | 0.002 | 7.43e-10 |
| rs62183994 | 2 | 204614872 | T | C | 0.051 | 0.027 | 0.005 | 2.65e-09 |
| rs715 | 2 | 211543055 | C | T | 0.311 | 0.018 | 0.002 | 3.1e-16 |
| rs6750754 | 2 | 213830187 | G | T | 0.264 | -0.064 | 0.002 | 8.35e-179 |
| rs35409523 | 2 | 213908457 | A | G | 0.076 | 0.050 | 0.004 | 1.43e-40 |
| rs2713548 | 2 | 227156662 | T | C | 0.633 | 0.013 | 0.002 | 1.02e-09 |
| rs7423615 | 2 | 231116874 | T | C | 0.187 | 0.023 | 0.003 | 5.1e-20 |
| rs1057258 | 2 | 234115629 | T | C | 0.178 | -0.032 | 0.003 | 2.52e-35 |
| rs34290285 | 2 | 242698640 | A | G | 0.257 | -0.046 | 0.002 | 4.73e-91 |
| rs76474320 | 2 | 242806022 | C | T | 0.077 | 0.028 | 0.004 | 5.76e-14 |
| rs80066203 | 2 | 26220710 | T | C | 0.068 | -0.024 | 0.004 | 8.53e-10 |
| rs11695281 | 2 | 28686568 | T | C | 0.514 | -0.023 | 0.002 | 4.09e-31 |
| rs6753015 | 2 | 28875673 | T | A | 0.315 | -0.013 | 0.002 | 2.42e-09 |
| rs10165678 | 2 | 30446568 | A | G | 0.758 | -0.015 | 0.002 | 1.9e-10 |
| rs17682575 | 2 | 43152304 | T | C | 0.201 | -0.018 | 0.002 | 1.33e-12 |
| rs149290349 | 2 | 43451957 | A | G | 0.075 | -0.027 | 0.004 | 1.18e-12 |
| rs778756 | 2 | 61781994 | G | A | 0.580 | -0.016 | 0.002 | 1.57e-15 |
| rs7569084 | 2 | 65656969 | T | C | 0.584 | 0.019 | 0.002 | 1.19e-20 |
| rs346835 | 2 | 8438693 | T | C | 0.328 | -0.027 | 0.002 | 7.55e-37 |
| rs62105489 | 2 | 8757326 | T | C | 0.053 | -0.030 | 0.004 | 1.13e-11 |
| rs56268488 | 2 | 95902245 | A | G | 0.086 | -0.021 | 0.004 | 4.14e-09 |
| rs2579505 | 2 | 97167962 | C | T | 0.638 | 0.042 | 0.002 | 5.79e-94 |
| rs4618204 | 3 | 101281534 | C | T | 0.444 | 0.018 | 0.002 | 1.09e-19 |
| rs4684037 | 3 | 10228040 | G | C | 0.144 | -0.021 | 0.003 | 4.24e-13 |
| rs111413886 | 3 | 111821047 | A | G | 0.031 | -0.040 | 0.006 | 3.16e-12 |
| rs1131199 | 3 | 112059768 | G | C | 0.530 | -0.025 | 0.002 | 9.55e-36 |
| rs2399441 | 3 | 112587012 | C | T | 0.351 | -0.019 | 0.002 | 8.08e-20 |
| rs10222469 | 3 | 121977125 | C | A | 0.819 | 0.017 | 0.003 | 4.95e-11 |
| rs310747 | 3 | 12259545 | G | T | 0.625 | -0.029 | 0.002 | 1.65e-44 |
| rs6802327 | 3 | 12410892 | T | G | 0.146 | 0.027 | 0.003 | 2.43e-21 |
| rs62270951 | 3 | 128225277 | G | A | 0.037 | -0.043 | 0.005 | 1.92e-16 |
| rs73238201 | 3 | 142204728 | T | C | 0.179 | -0.016 | 0.003 | 1.52e-10 |
| rs1516527 | 3 | 148609704 | C | T | 0.951 | -0.032 | 0.005 | 2.25e-12 |
| rs4680250 | 3 | 150941492 | G | C | 0.693 | -0.016 | 0.002 | 4.85e-13 |
| rs76830965 | 3 | 159637678 | A | C | 0.118 | -0.023 | 0.003 | 3.27e-14 |
| rs4464488 | 3 | 171498211 | T | C | 0.415 | -0.014 | 0.002 | 1.99e-12 |
| rs4074672 | 3 | 183730295 | T | C | 0.369 | 0.015 | 0.002 | 1.48e-12 |
| rs9815874 | 3 | 188441161 | T | C | 0.300 | 0.028 | 0.002 | 1.93e-39 |
| rs9818987 | 3 | 194402430 | T | C | 0.306 | 0.014 | 0.002 | 2.6e-10 |
| rs7636495 | 3 | 196367936 | A | G | 0.181 | -0.024 | 0.003 | 1.84e-20 |
| rs2089979 | 3 | 196501413 | G | A | 0.416 | -0.015 | 0.002 | 1.25e-13 |
| rs13073683 | 3 | 25393837 | C | T | 0.399 | 0.016 | 0.002 | 1.32e-14 |
| rs1353286 | 3 | 27772014 | G | T | 0.453 | 0.020 | 0.002 | 3.09e-24 |
| rs1695315 | 3 | 3153339 | A | G | 0.605 | -0.034 | 0.002 | 1.56e-61 |
| rs7646283 | 3 | 33046480 | T | C | 0.368 | 0.026 | 0.002 | 2.33e-36 |
| rs2228467 | 3 | 42906116 | C | T | 0.062 | 0.062 | 0.004 | 8.83e-52 |
| rs13316620 | 3 | 49045355 | C | T | 0.659 | 0.027 | 0.002 | 1.16e-33 |
| rs12487980 | 3 | 71760041 | A | C | 0.634 | 0.013 | 0.002 | 1.5e-09 |
| rs7646695 | 3 | 72172181 | T | C | 0.180 | 0.018 | 0.003 | 1.82e-11 |
| rs7649812 | 3 | 98449191 | G | C | 0.243 | 0.016 | 0.002 | 3.74e-12 |
| rs13105682 | 4 | 102702364 | G | T | 0.060 | -0.027 | 0.004 | 8.22e-10 |
| rs113473633 | 4 | 103449131 | G | A | 0.026 | -0.063 | 0.007 | 3.47e-22 |
| rs6827756 | 4 | 123184411 | C | T | 0.638 | -0.020 | 0.002 | 5.39e-23 |
| rs2390353 | 4 | 123602097 | C | T | 0.232 | 0.023 | 0.002 | 3.04e-22 |
| rs13120371 | 4 | 139092719 | G | A | 0.327 | 0.015 | 0.002 | 3e-13 |
| rs11931711 | 4 | 153291513 | T | C | 0.282 | 0.019 | 0.002 | 3.48e-18 |
| rs7441808 | 4 | 26090375 | G | A | 0.301 | 0.014 | 0.002 | 1.22e-10 |
| rs1828803 | 4 | 2689449 | A | C | 0.392 | 0.013 | 0.002 | 1.28e-10 |
| rs73232881 | 4 | 38664131 | C | T | 0.213 | 0.067 | 0.002 | 8.22e-171 |
| rs2566133 | 4 | 39022515 | C | T | 0.584 | -0.014 | 0.002 | 1.06e-12 |
| rs62308111 | 4 | 57738491 | T | G | 0.221 | 0.015 | 0.002 | 1.5e-10 |
| rs7687708 | 4 | 6906076 | G | T | 0.220 | -0.015 | 0.002 | 1.56e-09 |
| rs13138355 | 4 | 83545976 | T | C | 0.186 | -0.055 | 0.003 | 2.35e-102 |
| rs10062687 | 5 | 10624866 | G | T | 0.233 | 0.024 | 0.002 | 9.47e-25 |
| rs139640694 | 5 | 109938754 | G | A | 0.093 | 0.025 | 0.003 | 3.54e-13 |
| rs79881201 | 5 | 110427795 | T | C | 0.360 | 0.040 | 0.002 | 4.72e-85 |
| rs10059018 | 5 | 110501604 | T | G | 0.201 | -0.023 | 0.002 | 3.38e-21 |
| rs17516457 | 5 | 131590387 | C | T | 0.417 | -0.064 | 0.002 | 1e-200 |
| rs137906075 | 5 | 131972916 | G | A | 0.046 | 0.043 | 0.005 | 1.39e-18 |
| rs17653687 | 5 | 133468000 | G | A | 0.179 | -0.018 | 0.003 | 4.06e-12 |
| rs7700687 | 5 | 141491985 | T | C | 0.617 | 0.040 | 0.002 | 4.82e-86 |
| rs55977204 | 5 | 142501095 | C | T | 0.113 | 0.020 | 0.003 | 3.6e-10 |
| rs16903574 | 5 | 14610309 | G | C | 0.078 | 0.029 | 0.004 | 1.17e-13 |
| rs56330463 | 5 | 148200011 | C | T | 0.553 | 0.039 | 0.002 | 5.38e-86 |
| rs73272842 | 5 | 150453888 | A | G | 0.123 | -0.025 | 0.003 | 4.84e-17 |
| rs2431097 | 5 | 159890885 | T | C | 0.486 | 0.015 | 0.002 | 5.95e-14 |
| rs62385501 | 5 | 171950231 | A | T | 0.308 | -0.014 | 0.002 | 1.68e-11 |
| rs6556313 | 5 | 176792491 | G | A | 0.332 | 0.019 | 0.002 | 2.16e-20 |
| rs1961220 | 5 | 35844125 | A | G | 0.340 | -0.035 | 0.002 | 2.64e-63 |
| rs4703589 | 5 | 72097351 | C | T | 0.533 | 0.014 | 0.002 | 3.26e-13 |
| rs4703730 | 5 | 76549688 | T | C | 0.517 | -0.016 | 0.002 | 6.91e-16 |
| rs34495 | 5 | 98265807 | T | G | 0.304 | -0.020 | 0.002 | 3.67e-20 |
| rs62420764 | 6 | 106799970 | C | T | 0.140 | 0.019 | 0.003 | 4.21e-11 |
| rs12208103 | 6 | 107442431 | T | C | 0.378 | -0.031 | 0.002 | 6.83e-50 |
| rs783646 | 6 | 117286220 | C | G | 0.817 | -0.019 | 0.003 | 2.41e-13 |
| rs9389268 | 6 | 135419631 | G | A | 0.256 | -0.045 | 0.002 | 3.69e-87 |
| rs6924387 | 6 | 137082948 | G | A | 0.410 | 0.017 | 0.002 | 2.19e-16 |
| rs113496608 | 6 | 138161666 | A | G | 0.030 | -0.045 | 0.006 | 1.13e-14 |
| rs13207791 | 6 | 143249749 | G | A | 0.089 | 0.026 | 0.004 | 4.3e-13 |
| rs149110519 | 6 | 144385777 | T | C | 0.036 | 0.038 | 0.005 | 1.39e-12 |
| rs3093023 | 6 | 167534290 | A | G | 0.434 | 0.013 | 0.002 | 2.27e-10 |
| rs41546314 | 6 | 29910378 | T | C | 0.290 | 0.060 | 0.002 | 1.29e-166 |
| rs6929022 | 6 | 33562447 | G | A | 0.282 | -0.029 | 0.002 | 1.39e-40 |
| rs10456428 | 6 | 35012302 | T | C | 0.235 | -0.023 | 0.002 | 1.17e-22 |
| rs56987668 | 6 | 35319253 | C | T | 0.037 | -0.045 | 0.005 | 1.21e-17 |
| rs3793084 | 6 | 42142244 | C | T | 0.183 | 0.015 | 0.003 | 2.89e-09 |
| rs9349322 | 6 | 45634092 | C | A | 0.147 | 0.022 | 0.003 | 4.04e-15 |
| rs9504361 | 6 | 577820 | G | A | 0.447 | -0.032 | 0.002 | 7.98e-56 |
| rs138305038 | 6 | 88014416 | G | C | 0.474 | -0.016 | 0.002 | 2.07e-15 |
| rs62408224 | 6 | 90955995 | G | A | 0.350 | -0.042 | 0.002 | 5.69e-92 |
| rs73187850 | 7 | 101867824 | T | A | 0.278 | 0.018 | 0.002 | 8.69e-17 |
| rs12705849 | 7 | 112782556 | A | G | 0.407 | -0.021 | 0.002 | 2.55e-24 |
| rs7797428 | 7 | 124886883 | T | C | 0.469 | 0.012 | 0.002 | 5.85e-10 |
| rs3823536 | 7 | 128579666 | A | G | 0.467 | -0.018 | 0.002 | 3.16e-19 |
| rs56179563 | 7 | 129685597 | A | G | 0.389 | 0.018 | 0.002 | 3.87e-18 |
| rs3110791 | 7 | 135367828 | C | T | 0.623 | 0.015 | 0.002 | 7.49e-14 |
| rs12530946 | 7 | 148887942 | G | A | 0.613 | 0.043 | 0.002 | 3.99e-97 |
| rs12540285 | 7 | 150312474 | G | A | 0.221 | 0.015 | 0.002 | 1.01e-09 |
| rs6979947 | 7 | 157005863 | G | A | 0.266 | 0.013 | 0.002 | 3.75e-09 |
| rs10950642 | 7 | 17016646 | A | G | 0.363 | -0.019 | 0.002 | 4.15e-20 |
| rs61210140 | 7 | 20503007 | A | G | 0.245 | -0.058 | 0.002 | 7.12e-141 |
| rs4722171 | 7 | 22785717 | G | A | 0.592 | -0.027 | 0.002 | 1.83e-41 |
| rs6971710 | 7 | 3140173 | A | G | 0.201 | 0.028 | 0.002 | 2.75e-30 |
| rs60600003 | 7 | 37382465 | G | T | 0.100 | 0.043 | 0.003 | 3.05e-38 |
| rs1037674 | 7 | 50217850 | T | G | 0.292 | 0.013 | 0.002 | 1.63e-09 |
| rs13226583 | 7 | 75454152 | T | A | 0.116 | -0.062 | 0.003 | 1.59e-88 |
| rs55879743 | 7 | 75470364 | T | C | 0.066 | 0.078 | 0.004 | 7.07e-82 |
| rs62473720 | 7 | 77254547 | G | A | 0.317 | 0.014 | 0.002 | 2.64e-11 |
| rs56195338 | 7 | 8107922 | A | G | 0.058 | -0.030 | 0.004 | 1.88e-12 |
| rs4272 | 7 | 92236829 | G | A | 0.212 | 0.028 | 0.002 | 2.68e-30 |
| rs445 | 7 | 92408370 | T | C | 0.097 | -0.053 | 0.003 | 2.84e-56 |
| rs12681644 | 8 | 119972974 | T | C | 0.212 | -0.019 | 0.002 | 1.23e-15 |
| rs13251643 | 8 | 126240768 | T | A | 0.044 | 0.030 | 0.005 | 3.54e-10 |
| rs4870977 | 8 | 127526842 | C | G | 0.871 | -0.019 | 0.003 | 2.4e-10 |
| rs11786536 | 8 | 129000416 | A | G | 0.165 | -0.027 | 0.003 | 5.11e-23 |
| rs7840212 | 8 | 130599247 | T | C | 0.337 | -0.040 | 0.002 | 7.11e-81 |
| rs4236746 | 8 | 130699861 | G | A | 0.975 | 0.057 | 0.006 | 6.78e-19 |
| rs117961539 | 8 | 144989466 | A | G | 0.040 | -0.031 | 0.005 | 2.63e-09 |
| rs34173062 | 8 | 145158607 | A | G | 0.073 | 0.055 | 0.004 | 1.55e-40 |
| rs3208305 | 8 | 19823648 | T | A | 0.287 | 0.013 | 0.002 | 8.64e-10 |
| rs4871849 | 8 | 22964719 | A | G | 0.707 | -0.033 | 0.002 | 1.06e-51 |
| rs10088702 | 8 | 27263107 | T | C | 0.264 | -0.017 | 0.002 | 2.1e-13 |
| rs45577137 | 8 | 48651633 | G | A | 0.045 | -0.060 | 0.005 | 5.1e-31 |
| rs62539154 | 8 | 48677930 | A | G | 0.097 | 0.027 | 0.003 | 7.2e-16 |
| rs7846314 | 8 | 61650831 | T | A | 0.187 | -0.033 | 0.003 | 3.92e-38 |
| rs6989099 | 8 | 66898262 | C | T | 0.317 | -0.018 | 0.002 | 8.58e-17 |
| rs2887502 | 8 | 79756591 | T | C | 0.590 | -0.014 | 0.002 | 1.74e-11 |
| rs6986109 | 8 | 81286298 | T | G | 0.704 | -0.016 | 0.002 | 4.24e-14 |
| rs4240624 | 8 | 9184231 | A | G | 0.909 | -0.020 | 0.003 | 3.62e-09 |
| rs574183 | 8 | 95973816 | G | A | 0.389 | -0.014 | 0.002 | 1.2e-11 |
| rs7026022 | 9 | 102562216 | C | A | 0.378 | 0.012 | 0.002 | 3.38e-09 |
| rs496475 | 9 | 113638236 | G | T | 0.387 | 0.035 | 0.002 | 1.83e-66 |
| rs911603 | 9 | 117697584 | A | C | 0.404 | -0.025 | 0.002 | 1.42e-33 |
| rs74612091 | 9 | 135877278 | A | T | 0.063 | 0.061 | 0.004 | 1.23e-49 |
| rs460631 | 9 | 4851440 | G | A | 0.885 | 0.026 | 0.003 | 1.88e-16 |
| rs7868130 | 9 | 4998401 | T | C | 0.259 | 0.032 | 0.002 | 3.35e-46 |
| rs4142528 | 9 | 6172296 | A | T | 0.672 | -0.075 | 0.002 | 1e-200 |
| rs1547258 | 9 | 6523056 | C | T | 0.708 | -0.019 | 0.002 | 1.49e-18 |
| rs295273 | 9 | 86466968 | A | G | 0.256 | 0.021 | 0.002 | 1.5e-19 |
| rs6479336 | 9 | 94110288 | A | T | 0.185 | -0.029 | 0.003 | 3.44e-30 |
| rs78691875 | 12 | 113070264 | C | A | 0.979 | 0.051 | 0.007 | 1.69e-13 |
| rs12928503 | 16 | 67377216 | C | T | 0.980 | 0.049 | 0.007 | 1.92e-11 |
| rs151036504 | 6 | 31409578 | C | T | 0.985 | -0.062 | 0.008 | 2.5e-14 |
| rs150640087 | 7 | 50444152 | G | T | 0.984 | -0.100 | 0.008 | 1.8e-35 |
| rs10892296 | 11 | 118718729 | T | C | 0.195 | 0.018 | 0.003 | 2.63e-12 |
| rs12577643 | 11 | 47467172 | T | A | 0.326 | 0.017 | 0.002 | 4.31e-15 |
| rs11559982 | 12 | 54711574 | G | A | 0.556 | 0.014 | 0.002 | 1.92e-12 |
| rs4906317 | 14 | 103814500 | T | G | 0.505 | -0.015 | 0.002 | 9.44e-12 |
| rs8010804 | 14 | 23396480 | G | T | 0.614 | 0.013 | 0.002 | 6.51e-10 |
| rs4924544 | 15 | 41693455 | T | G | 0.525 | 0.015 | 0.002 | 4.26e-13 |
| rs76758258 | 15 | 91190935 | A | T | 0.240 | -0.020 | 0.002 | 2.4e-16 |
| rs7184899 | 16 | 4472087 | A | T | 0.704 | -0.014 | 0.002 | 1.16e-09 |
| rs9906974 | 17 | 43822772 | C | T | 0.220 | -0.034 | 0.002 | 8.37e-44 |
| rs4789797 | 17 | 80527895 | A | G | 0.587 | 0.012 | 0.002 | 2.67e-09 |
| rs3848640 | 19 | 1037986 | G | A | 0.329 | -0.016 | 0.002 | 9.78e-13 |
| rs2793925 | 1 | 120652158 | A | T | 0.684 | 0.017 | 0.002 | 7.93e-14 |
| rs111548294 | 1 | 150911337 | T | C | 0.013 | 0.087 | 0.009 | 2.61e-21 |
| rs12116645 | 1 | 204524474 | T | G | 0.675 | 0.016 | 0.002 | 6.31e-13 |
| rs192397873 | 1 | 28002091 | C | T | 0.053 | 0.030 | 0.005 | 1.56e-10 |
| rs2422631 | 20 | 1556758 | C | T | 0.739 | -0.014 | 0.002 | 2.04e-09 |
| rs7264802 | 20 | 62692440 | G | A | 0.749 | -0.015 | 0.002 | 1.07e-10 |
| rs12622600 | 2 | 71279940 | T | C | 0.270 | -0.016 | 0.002 | 9.11e-12 |
| rs11126424 | 2 | 74262353 | C | T | 0.306 | -0.016 | 0.002 | 1.84e-12 |
| rs2366640 | 2 | 85810734 | C | A | 0.319 | -0.017 | 0.002 | 5.08e-14 |
| rs112924476 | 3 | 128265131 | A | G | 0.119 | -0.098 | 0.003 | 1e-200 |
| rs6765218 | 3 | 128380332 | G | A | 0.254 | 0.045 | 0.002 | 2.65e-80 |
| rs191674933 | 3 | 169488691 | T | A | 0.240 | 0.025 | 0.002 | 4.26e-25 |
| rs186984319 | 6 | 16702251 | C | T | 0.318 | -0.013 | 0.002 | 4.37e-09 |
| rs9270464 | 6 | 32558914 | G | A | 0.345 | -0.035 | 0.002 | 5.57e-52 |
| rs9272400 | 6 | 32604898 | A | G | 0.655 | 0.055 | 0.002 | 5.11e-130 |
| rs2666969 | 6 | 336410 | G | A | 0.256 | -0.040 | 0.003 | 7.32e-38 |
| rs77396612 | 7 | 138737519 | C | A | 0.025 | 0.040 | 0.007 | 4.33e-09 |
| rs572698961 | 7 | 50802158 | A | C | 0.086 | -0.031 | 0.004 | 4.86e-17 |
| rs56072276 | 7 | 98778222 | A | G | 0.688 | -0.016 | 0.002 | 6.88e-13 |
| rs7040707 | 9 | 117680732 | G | A | 0.048 | -0.031 | 0.005 | 1.69e-09 |
| rs115478735 | 9 | 136149711 | T | A | 0.183 | -0.028 | 0.003 | 1.1e-25 |
| rs146671954 | 9 | 136934203 | A | G | 0.164 | -0.025 | 0.003 | 2.1e-20 |
| rs68142670 | 9 | 139324573 | C | T | 0.278 | 0.022 | 0.002 | 3.07e-21 |
| rs10962679 | 9 | 16905441 | C | T | 0.754 | -0.014 | 0.002 | 1.26e-09 |
| rs7874405 | 9 | 21980944 | T | C | 0.712 | 0.018 | 0.002 | 9.15e-16 |
| rs12339911 | 9 | 34845984 | A | T | 0.613 | -0.013 | 0.002 | 6.18e-10 |
| rs193101979 | 5 | 129836370 | I | D | 0.160 | 0.017 | 0.003 | 3e-09 |

**Table S5. 146 SNPs significantly associated with basophil count used IVs in forward MR analyses derived from Vuckovic D et al.** Chr: Chromosome; EA: Effect allele; NEA: Non-effect allele; EAF: Effect allele frequency.

| SNP | Chr | Pos | EA | NEA | EAF | Beta | SE | P-value |
| --- | --- | --- | --- | --- | --- | --- | --- | --- |
| rs35076930 | 1 | 17677636 | T | C | 0.293 | 0.021 | 0.003 | 1.3e-13 |
| rs12097415 | 1 | 21506276 | A | T | 0.547 | -0.021 | 0.002 | 9.3e-21 |
| rs3917932 | 1 | 36943916 | G | C | 0.577 | -0.015 | 0.002 | 1.3e-11 |
| rs17613339 | 1 | 118139540 | T | C | 0.128 | -0.022 | 0.003 | 6.7e-11 |
| rs12143614 | 1 | 150932696 | A | T | 0.117 | -0.023 | 0.003 | 5e-11 |
| rs12075 | 1 | 159175354 | A | G | 0.579 | 0.031 | 0.002 | 2.5e-43 |
| rs2926468 | 1 | 161612578 | C | T | 0.468 | -0.019 | 0.002 | 5e-17 |
| rs10798014 | 1 | 185394834 | G | A | 0.321 | -0.014 | 0.002 | 4.7e-09 |
| rs12123922 | 1 | 205140436 | A | G | 0.441 | -0.031 | 0.002 | 1.5e-44 |
| rs1086893 | 1 | 212563114 | C | T | 0.344 | 0.032 | 0.002 | 3.2e-43 |
| rs10927074 | 1 | 236103965 | C | T | 0.892 | 0.088 | 0.004 | 4.7e-135 |
| rs62105478 | 2 | 8735869 | A | G | 0.052 | -0.031 | 0.005 | 8.9e-10 |
| rs12470883 | 2 | 65651851 | A | G | 0.409 | 0.022 | 0.002 | 8.1e-23 |
| rs4851592 | 2 | 103087641 | T | C | 0.303 | 0.017 | 0.002 | 1.5e-12 |
| rs1427499 | 2 | 145400317 | G | A | 0.710 | 0.017 | 0.002 | 6.3e-12 |
| rs78862952 | 2 | 146506667 | C | T | 0.092 | 0.026 | 0.004 | 2.4e-11 |
| rs1424951 | 2 | 148533708 | C | T | 0.334 | 0.015 | 0.002 | 2.4e-10 |
| rs4602187 | 2 | 181834018 | T | C | 0.683 | 0.015 | 0.002 | 2.2e-10 |
| rs4667282 | 2 | 182312809 | A | T | 0.561 | 0.016 | 0.002 | 3e-13 |
| rs73987603 | 2 | 213844822 | A | G | 0.166 | 0.024 | 0.003 | 1.2e-15 |
| rs10716631 | 2 | 219138170 | G | T | 0.536 | 0.017 | 0.002 | 6e-14 |
| rs4324460 | 3 | 3120542 | G | T | 0.219 | 0.017 | 0.003 | 8.3e-11 |
| rs1669340 | 3 | 3198380 | T | G | 0.839 | -0.025 | 0.003 | 1.1e-16 |
| rs6766037 | 3 | 16908186 | G | A | 0.218 | -0.041 | 0.003 | 1.1e-52 |
| rs12497690 | 3 | 27795397 | C | A | 0.371 | 0.016 | 0.002 | 6.7e-12 |
| rs3181077 | 3 | 46250652 | T | C | 0.718 | -0.043 | 0.002 | 1.2e-69 |
| rs13063578 | 3 | 47087837 | A | T | 0.401 | 0.017 | 0.002 | 2.4e-13 |
| rs6782812 | 3 | 128317997 | A | G | 0.893 | 0.069 | 0.004 | 9.2e-82 |
| rs9819371 | 3 | 141206800 | T | C | 0.065 | -0.033 | 0.004 | 3.5e-13 |
| rs1973791 | 3 | 187416634 | A | C | 0.617 | -0.018 | 0.002 | 7.4e-15 |
| rs13434265 | 3 | 196489422 | T | C | 0.415 | -0.013 | 0.002 | 4.9e-09 |
| rs11930388 | 4 | 9985884 | T | C | 0.211 | -0.016 | 0.003 | 4.9e-09 |
| rs10023310 | 4 | 79629863 | C | T | 0.601 | 0.017 | 0.002 | 3.3e-13 |
| rs906149 | 4 | 83537777 | G | C | 0.221 | -0.017 | 0.003 | 1.4e-10 |
| rs55913164 | 4 | 87898930 | C | T | 0.190 | -0.031 | 0.003 | 8.9e-29 |
| rs374187209 | 4 | 103410817 | A | T | 0.453 | -0.016 | 0.002 | 1e-11 |
| rs10214237 | 5 | 35883734 | C | T | 0.275 | -0.019 | 0.002 | 4.1e-14 |
| rs34500 | 5 | 98294885 | A | G | 0.084 | -0.028 | 0.004 | 1.7e-12 |
| rs2271352 | 5 | 126091428 | C | G | 0.226 | 0.046 | 0.003 | 2.8e-66 |
| rs6579875 | 5 | 150782574 | T | C | 0.389 | -0.016 | 0.002 | 4.8e-12 |
| rs2594836 | 5 | 173205318 | A | G | 0.722 | -0.017 | 0.002 | 6.4e-12 |
| rs62393575 | 6 | 7058516 | C | G | 0.053 | -0.030 | 0.005 | 3.1e-09 |
| rs1100575 | 6 | 22373710 | A | G | 0.545 | 0.022 | 0.002 | 9.3e-23 |
| rs2524079 | 6 | 31242174 | A | G | 0.417 | 0.031 | 0.002 | 1.3e-44 |
| rs9267893 | 6 | 32202221 | T | A | 0.210 | -0.026 | 0.003 | 3.9e-21 |
| rs141378803 | 6 | 32490016 | A | G | 0.233 | 0.027 | 0.003 | 6.4e-18 |
| rs915125 | 6 | 82463376 | T | C | 0.279 | -0.034 | 0.002 | 3.3e-43 |
| rs72895231 | 6 | 82679396 | A | C | 0.596 | -0.017 | 0.002 | 9e-14 |
| rs6927569 | 6 | 109621494 | C | T | 0.523 | 0.019 | 0.002 | 1.3e-18 |
| rs9376098 | 6 | 135499460 | A | T | 0.349 | 0.026 | 0.002 | 2e-28 |
| rs73049252 | 7 | 8016602 | A | G | 0.059 | -0.044 | 0.005 | 1.2e-20 |
| rs2158799 | 7 | 28277107 | G | C | 0.610 | 0.016 | 0.002 | 2.2e-12 |
| rs56388170 | 7 | 28724374 | T | G | 0.292 | 0.034 | 0.002 | 5.5e-44 |
| rs149007767 | 7 | 50370254 | T | C | 0.162 | 0.031 | 0.003 | 4.4e-24 |
| rs1186222 | 7 | 75247329 | T | C | 0.462 | -0.027 | 0.002 | 1.9e-33 |
| rs11974227 | 7 | 80222408 | T | C | 0.064 | 0.028 | 0.005 | 5.4e-10 |
| rs8179 | 7 | 92236164 | C | T | 0.792 | -0.034 | 0.003 | 3.5e-36 |
| rs445 | 7 | 92408370 | T | C | 0.096 | -0.087 | 0.004 | 2.5e-119 |
| rs11772895 | 7 | 143081942 | C | G | 0.277 | 0.040 | 0.002 | 3.3e-58 |
| rs9314614 | 8 | 6697731 | G | C | 0.533 | 0.033 | 0.002 | 7.1e-49 |
| rs2738104 | 8 | 6792257 | A | G | 0.673 | 0.021 | 0.002 | 1.2e-18 |
| rs10086568 | 8 | 6900336 | A | G | 0.321 | -0.041 | 0.002 | 6.8e-67 |
| rs877116 | 8 | 10712945 | T | G | 0.589 | 0.017 | 0.002 | 3.2e-14 |
| rs9644063 | 8 | 22974450 | C | T | 0.823 | -0.027 | 0.003 | 1.6e-20 |
| rs45577137 | 8 | 48651633 | G | A | 0.045 | -0.052 | 0.006 | 1.3e-19 |
| rs16923637 | 8 | 59567885 | T | G | 0.286 | 0.014 | 0.002 | 3.9e-09 |
| rs7818415 | 8 | 70727663 | A | T | 0.485 | -0.014 | 0.002 | 1e-09 |
| rs4541868 | 8 | 106590705 | A | C | 0.275 | -0.018 | 0.002 | 3.9e-13 |
| rs10956401 | 8 | 129002419 | A | G | 0.343 | -0.017 | 0.002 | 2.7e-13 |
| rs3731211 | 9 | 21986847 | A | T | 0.722 | 0.017 | 0.002 | 4.2e-12 |
| rs12376511 | 9 | 22142756 | C | T | 0.162 | -0.019 | 0.003 | 1.2e-10 |
| rs2150052 | 9 | 113945067 | T | A | 0.505 | -0.014 | 0.002 | 5.7e-10 |
| rs11253511 | 10 | 964832 | T | C | 0.238 | 0.052 | 0.003 | 1.4e-87 |
| rs11595895 | 10 | 26734518 | A | C | 0.403 | 0.024 | 0.002 | 2.7e-26 |
| rs2998286 | 10 | 28780373 | C | T | 0.769 | 0.025 | 0.003 | 4.4e-21 |
| rs1897191 | 10 | 77143467 | G | A | 0.617 | -0.019 | 0.002 | 2.3e-16 |
| rs56014906 | 10 | 102103508 | G | A | 0.368 | -0.018 | 0.002 | 3.9e-15 |
| rs11195959 | 10 | 114199287 | A | G | 0.096 | 0.023 | 0.004 | 1.8e-09 |
| rs3781452 | 10 | 126355129 | T | C | 0.633 | 0.022 | 0.002 | 7.1e-21 |
| rs1058900 | 11 | 308290 | C | T | 0.568 | 0.018 | 0.002 | 8.2e-15 |
| rs12278324 | 11 | 3862058 | A | C | 0.619 | 0.016 | 0.002 | 2.3e-12 |
| rs742631 | 11 | 32077472 | T | C | 0.282 | -0.016 | 0.002 | 6.6e-11 |
| rs531612 | 11 | 65705432 | T | C | 0.509 | 0.013 | 0.002 | 4.4e-09 |
| rs74472890 | 11 | 72946279 | C | T | 0.049 | 0.119 | 0.005 | 4.7e-120 |
| rs4753251 | 11 | 89654536 | A | T | 0.730 | -0.016 | 0.003 | 9.2e-11 |
| rs2606724 | 11 | 113957880 | A | G | 0.455 | 0.017 | 0.002 | 3.2e-13 |
| rs10893845 | 11 | 128186882 | G | T | 0.502 | -0.014 | 0.002 | 8.5e-11 |
| rs2286599 | 12 | 6499533 | A | G | 0.142 | 0.020 | 0.003 | 3.4e-10 |
| rs146970669 | 12 | 27103449 | A | G | 0.093 | 0.026 | 0.004 | 9.9e-12 |
| rs117053853 | 12 | 51720047 | A | G | 0.011 | 0.124 | 0.011 | 2.8e-31 |
| rs2118140 | 12 | 66698461 | G | A | 0.528 | 0.015 | 0.002 | 2.6e-11 |
| rs75084335 | 12 | 76981753 | G | C | 0.069 | 0.035 | 0.004 | 3e-15 |
| rs3184504 | 12 | 111884608 | C | T | 0.519 | -0.029 | 0.002 | 3.1e-39 |
| rs11064881 | 12 | 120146925 | A | G | 0.073 | -0.033 | 0.004 | 5.5e-15 |
| rs4475963 | 12 | 129302255 | G | T | 0.370 | -0.017 | 0.002 | 2.4e-13 |
| rs76428106 | 13 | 28604007 | C | T | 0.013 | 0.082 | 0.010 | 8.8e-16 |
| rs149391212 | 14 | 23544999 | T | C | 0.016 | 0.069 | 0.009 | 1.8e-14 |
| rs2239630 | 14 | 23589349 | G | A | 0.558 | 0.028 | 0.002 | 8e-37 |
| rs114917114 | 14 | 35366971 | G | A | 0.353 | 0.014 | 0.002 | 2.7e-09 |
| rs11158159 | 14 | 57857162 | C | G | 0.773 | 0.031 | 0.003 | 9.6e-30 |
| rs72721631 | 14 | 75807762 | A | C | 0.238 | -0.021 | 0.003 | 4.3e-16 |
| rs2289511 | 14 | 88454910 | A | G | 0.582 | -0.017 | 0.002 | 6.9e-15 |
| rs79429038 | 15 | 42677739 | A | G | 0.133 | 0.023 | 0.003 | 3.3e-12 |
| rs2070596 | 15 | 50545159 | A | T | 0.211 | 0.020 | 0.003 | 6.5e-13 |
| rs3848148 | 15 | 64637536 | C | T | 0.943 | 0.035 | 0.005 | 4.1e-13 |
| rs1847271 | 15 | 69632367 | A | G | 0.624 | 0.016 | 0.002 | 1.2e-12 |
| rs7166645 | 15 | 83740277 | A | G | 0.387 | 0.015 | 0.002 | 1.1e-10 |
| rs2074585 | 15 | 91009484 | A | G | 0.514 | -0.036 | 0.002 | 2.8e-60 |
| rs8026614 | 15 | 100051136 | T | C | 0.472 | -0.014 | 0.002 | 9.5e-10 |
| rs67175901 | 15 | 101748227 | T | C | 0.108 | 0.029 | 0.004 | 1.4e-16 |
| rs875740 | 16 | 16123048 | A | C | 0.666 | -0.019 | 0.002 | 6.7e-16 |
| rs7196129 | 16 | 30471109 | C | T | 0.528 | 0.019 | 0.002 | 1.2e-16 |
| rs12927351 | 16 | 74596618 | A | C | 0.198 | 0.017 | 0.003 | 8.8e-10 |
| rs247830 | 16 | 84581925 | C | T | 0.304 | 0.015 | 0.002 | 1.6e-10 |
| rs4782302 | 16 | 88524389 | C | A | 0.436 | 0.018 | 0.002 | 4.4e-16 |
| rs7503461 | 17 | 2883320 | T | C | 0.319 | -0.015 | 0.002 | 7.3e-10 |
| rs12453682 | 17 | 37770005 | T | C | 0.695 | -0.019 | 0.002 | 1.4e-14 |
| rs12941811 | 17 | 38159335 | C | T | 0.578 | -0.048 | 0.002 | 6.9e-102 |
| rs8178414 | 17 | 56345363 | T | C | 0.013 | 0.088 | 0.010 | 3.9e-19 |
| rs34097845 | 17 | 56358429 | T | C | 0.057 | 0.049 | 0.005 | 1.9e-23 |
| rs1295927 | 17 | 57929535 | G | A | 0.447 | -0.027 | 0.002 | 1.1e-33 |
| rs2384952 | 17 | 72744512 | C | T | 0.539 | -0.017 | 0.002 | 3.6e-13 |
| rs479032 | 18 | 9850581 | A | G | 0.656 | -0.016 | 0.002 | 1.1e-11 |
| rs17758695 | 18 | 60920854 | T | C | 0.029 | -0.104 | 0.007 | 1.6e-57 |
| rs62132278 | 19 | 836654 | T | C | 0.256 | -0.031 | 0.003 | 1.2e-32 |
| rs2336068 | 19 | 5813519 | C | A | 0.276 | 0.015 | 0.003 | 7.4e-10 |
| rs2967595 | 19 | 8566299 | T | C | 0.168 | -0.030 | 0.003 | 3.3e-23 |
| rs873636 | 19 | 16514771 | G | A | 0.308 | 0.024 | 0.002 | 2.6e-23 |
| rs55990904 | 19 | 18302239 | C | T | 0.274 | -0.016 | 0.002 | 1.4e-10 |
| rs8113682 | 19 | 19743730 | G | T | 0.748 | -0.023 | 0.003 | 1.4e-19 |
| rs78744187 | 19 | 33754548 | T | C | 0.081 | -0.088 | 0.004 | 8.3e-105 |
| rs34158728 | 19 | 38903032 | A | G | 0.029 | 0.105 | 0.007 | 1.8e-56 |
| rs431329 | 19 | 40221107 | G | C | 0.663 | -0.017 | 0.002 | 6.9e-14 |
| rs12459419 | 19 | 51728477 | T | C | 0.324 | -0.016 | 0.002 | 3.1e-12 |
| rs7250849 | 19 | 52158316 | T | G | 0.107 | 0.021 | 0.004 | 3.5e-09 |
| rs6045612 | 20 | 1931001 | T | C | 0.263 | -0.016 | 0.003 | 1.1e-10 |
| rs4911102 | 20 | 31179500 | T | C | 0.234 | -0.017 | 0.003 | 2.9e-10 |
| rs6141333 | 20 | 31263324 | A | C | 0.147 | 0.025 | 0.003 | 1.6e-15 |
| rs4931 | 20 | 43530234 | C | A | 0.271 | -0.024 | 0.002 | 1.6e-21 |
| rs6091176 | 20 | 49150510 | T | C | 0.261 | 0.015 | 0.003 | 2.1e-09 |
| rs310631 | 20 | 62196253 | A | C | 0.411 | 0.014 | 0.002 | 1.1e-09 |
| rs11702209 | 21 | 38808428 | G | C | 0.118 | -0.031 | 0.003 | 6.3e-20 |
| rs2836154 | 21 | 39485842 | T | C | 0.336 | 0.017 | 0.002 | 8.8e-14 |
| rs5756822 | 22 | 38196474 | T | C | 0.556 | -0.014 | 0.002 | 4.6e-10 |
| rs761702 | 22 | 41404970 | A | G | 0.065 | 0.038 | 0.004 | 2.5e-17 |
| rs5766582 | 22 | 45632528 | T | C | 0.475 | -0.018 | 0.002 | 1.4e-16 |

**Table S6. 166 SNPs significantly associated with basophil count used IVs in forward MR analyses derived from Chen MH et al.** Chr: Chromosome; EA: Effect allele; NEA: Non-effect allele; EAF: Effect allele frequency.

| SNP | Chr | Pos | EA | NEA | EAF | Beta | SE | P-value |
| --- | --- | --- | --- | --- | --- | --- | --- | --- |
| rs61872688 | 10 | 101273546 | A | G | 0.381 | -0.013 | 0.002 | 1.04e-09 |
| rs10883782 | 10 | 104583932 | G | A | 0.169 | 0.018 | 0.003 | 1.28e-10 |
| rs3781452 | 10 | 126355129 | T | C | 0.632 | 0.020 | 0.002 | 1.34e-20 |
| rs11595895 | 10 | 26734518 | A | C | 0.403 | 0.024 | 0.002 | 1.23e-29 |
| rs2998286 | 10 | 28780373 | C | T | 0.770 | 0.021 | 0.002 | 1.31e-17 |
| rs7078507 | 10 | 77140864 | G | A | 0.609 | -0.016 | 0.002 | 2.54e-14 |
| rs2606724 | 11 | 113957880 | A | G | 0.453 | 0.016 | 0.002 | 1.33e-13 |
| rs10893844 | 11 | 128185850 | C | G | 0.501 | -0.015 | 0.002 | 4.22e-14 |
| rs695113 | 11 | 128562098 | T | C | 0.696 | 0.017 | 0.002 | 3.93e-14 |
| rs734095 | 11 | 2323198 | G | C | 0.113 | -0.028 | 0.003 | 2.04e-17 |
| rs6421984 | 11 | 305619 | C | T | 0.514 | 0.018 | 0.002 | 6.97e-19 |
| rs742631 | 11 | 32077472 | T | C | 0.284 | -0.017 | 0.002 | 4.83e-13 |
| rs10835333 | 11 | 3957766 | G | A | 0.349 | 0.015 | 0.002 | 1.68e-11 |
| rs10896064 | 11 | 65641033 | C | G | 0.532 | 0.013 | 0.002 | 8.01e-11 |
| rs74472890 | 11 | 72946279 | C | T | 0.049 | 0.103 | 0.005 | 1.02e-103 |
| rs4753251 | 11 | 89654536 | A | T | 0.728 | -0.022 | 0.002 | 3.42e-20 |
| rs3184504 | 12 | 111884608 | C | T | 0.519 | -0.028 | 0.002 | 5.08e-43 |
| rs11064881 | 12 | 120146925 | A | G | 0.074 | -0.032 | 0.004 | 2.05e-16 |
| rs4475963 | 12 | 129302255 | G | T | 0.370 | -0.014 | 0.002 | 2.11e-10 |
| rs146970669 | 12 | 27103449 | A | G | 0.094 | 0.022 | 0.004 | 2.43e-10 |
| rs17860282 | 12 | 51740350 | T | C | 0.011 | 0.105 | 0.010 | 2.62e-26 |
| rs2286599 | 12 | 6499533 | A | G | 0.142 | 0.023 | 0.003 | 1.15e-14 |
| rs2118140 | 12 | 66698461 | G | A | 0.528 | 0.018 | 0.002 | 4.03e-19 |
| rs75084335 | 12 | 76981753 | G | C | 0.069 | 0.028 | 0.004 | 3.75e-12 |
| rs76428106 | 13 | 28604007 | C | T | 0.013 | 0.082 | 0.009 | 2.78e-18 |
| rs2701863 | 13 | 41170140 | T | C | 0.306 | 0.052 | 0.007 | 1.74e-13 |
| rs2239635 | 14 | 23588731 | C | G | 0.704 | 0.034 | 0.002 | 1.89e-52 |
| rs11158159 | 14 | 57857162 | C | G | 0.770 | 0.028 | 0.003 | 4.59e-28 |
| rs72721631 | 14 | 75807762 | A | C | 0.237 | -0.019 | 0.002 | 1.6e-15 |
| rs2289511 | 14 | 88454910 | A | G | 0.582 | -0.016 | 0.002 | 3.3e-15 |
| rs72697295 | 14 | 93069980 | C | G | 0.181 | -0.018 | 0.003 | 8e-11 |
| rs67175901 | 15 | 101748227 | T | C | 0.108 | 0.026 | 0.003 | 8.24e-15 |
| rs79429038 | 15 | 42677739 | A | G | 0.132 | 0.021 | 0.003 | 1.33e-11 |
| rs2070596 | 15 | 50545159 | A | T | 0.211 | 0.023 | 0.003 | 3.3e-19 |
| rs62021606 | 15 | 64548734 | G | T | 0.051 | -0.034 | 0.005 | 2e-13 |
| rs7496362 | 15 | 65758874 | G | C | 0.366 | -0.013 | 0.002 | 2.71e-09 |
| rs12443468 | 15 | 81888088 | G | A | 0.226 | 0.015 | 0.002 | 3.04e-09 |
| rs2074585 | 15 | 91009484 | A | G | 0.515 | -0.031 | 0.002 | 2.71e-51 |
| rs875740 | 16 | 16123048 | A | C | 0.665 | -0.018 | 0.002 | 1.52e-16 |
| rs7196129 | 16 | 30471109 | C | T | 0.529 | 0.017 | 0.002 | 7.19e-16 |
| rs9928015 | 16 | 57570561 | T | G | 0.302 | -0.015 | 0.002 | 2.01e-11 |
| rs247833 | 16 | 84581684 | A | G | 0.249 | 0.016 | 0.002 | 1.42e-11 |
| rs12447180 | 16 | 88517722 | C | T | 0.324 | 0.023 | 0.002 | 2.06e-26 |
| rs7503461 | 17 | 2883320 | T | C | 0.319 | -0.013 | 0.002 | 9.6e-10 |
| rs12453682 | 17 | 37770005 | T | C | 0.695 | -0.018 | 0.002 | 3.6e-16 |
| rs12941811 | 17 | 38159335 | C | T | 0.578 | -0.046 | 0.002 | 7.71e-111 |
| rs8178414 | 17 | 56345363 | T | C | 0.013 | 0.074 | 0.009 | 4.48e-16 |
| rs34097845 | 17 | 56358429 | T | C | 0.057 | 0.043 | 0.005 | 4.58e-21 |
| rs1295927 | 17 | 57929535 | G | A | 0.448 | -0.025 | 0.002 | 5.83e-34 |
| rs2959356 | 18 | 23590203 | G | A | 0.298 | 0.014 | 0.002 | 4.14e-10 |
| rs17758695 | 18 | 60920854 | T | C | 0.030 | -0.125 | 0.006 | 1.47e-91 |
| rs561102 | 18 | 9851388 | T | C | 0.655 | -0.014 | 0.002 | 8.12e-11 |
| rs2607278 | 19 | 16568197 | T | C | 0.306 | 0.023 | 0.002 | 1.79e-24 |
| rs8113682 | 19 | 19743730 | G | T | 0.747 | -0.022 | 0.002 | 6.41e-20 |
| rs78744187 | 19 | 33754548 | T | C | 0.082 | -0.130 | 0.004 | 1e-200 |
| rs34158728 | 19 | 38903032 | A | G | 0.028 | 0.095 | 0.006 | 3.24e-53 |
| rs12459419 | 19 | 51728477 | T | C | 0.323 | -0.017 | 0.002 | 2.44e-14 |
| rs7250849 | 19 | 52158316 | T | G | 0.107 | 0.022 | 0.003 | 1.01e-10 |
| rs76427287 | 19 | 837190 | C | T | 0.254 | -0.029 | 0.002 | 2.07e-31 |
| rs12143614 | 1 | 150932696 | A | T | 0.117 | -0.023 | 0.003 | 4.72e-13 |
| rs12075 | 1 | 159175354 | A | G | 0.580 | 0.028 | 0.002 | 6.02e-41 |
| rs17625587 | 1 | 198990494 | A | G | 0.264 | 0.015 | 0.002 | 1.25e-10 |
| rs1086893 | 1 | 212563114 | C | T | 0.343 | 0.030 | 0.002 | 2.41e-43 |
| rs4060971 | 1 | 21506930 | T | C | 0.551 | -0.021 | 0.002 | 1.85e-23 |
| rs10927074 | 1 | 236103965 | C | T | 0.892 | 0.079 | 0.003 | 3e-127 |
| rs56043070 | 1 | 247719769 | A | G | 0.071 | -0.026 | 0.004 | 6.04e-11 |
| rs7515985 | 1 | 28997021 | G | A | 0.416 | 0.014 | 0.002 | 6.05e-11 |
| rs1537061 | 1 | 87739219 | C | T | 0.099 | -0.025 | 0.003 | 2.55e-13 |
| rs6045612 | 20 | 1931001 | T | C | 0.264 | -0.017 | 0.002 | 7.18e-13 |
| rs4911102 | 20 | 31179500 | T | C | 0.234 | -0.022 | 0.002 | 6.46e-19 |
| rs6141781 | 20 | 31263342 | T | C | 0.147 | 0.033 | 0.003 | 4.12e-29 |
| rs6029234 | 20 | 39259278 | C | G | 0.626 | 0.013 | 0.002 | 3.46e-09 |
| rs16989483 | 20 | 43537200 | C | T | 0.274 | -0.021 | 0.002 | 3.15e-20 |
| rs6091176 | 20 | 49150510 | T | C | 0.261 | 0.014 | 0.002 | 4.8e-09 |
| rs310631 | 20 | 62196253 | A | C | 0.412 | 0.012 | 0.002 | 4.42e-09 |
| rs2834670 | 21 | 36280376 | G | A | 0.172 | 0.028 | 0.003 | 1.69e-18 |
| rs138595256 | 21 | 36789420 | G | C | 0.026 | 0.064 | 0.007 | 1.82e-20 |
| rs7285377 | 22 | 19987202 | T | G | 0.284 | 0.014 | 0.002 | 1.86e-10 |
| rs34288539 | 22 | 24642009 | T | C | 0.274 | -0.015 | 0.002 | 6.34e-11 |
| rs5756822 | 22 | 38196474 | T | C | 0.556 | -0.012 | 0.002 | 3.79e-09 |
| rs34780507 | 22 | 41405753 | G | A | 0.066 | 0.035 | 0.004 | 2.96e-17 |
| rs5766582 | 22 | 45632528 | T | C | 0.475 | -0.017 | 0.002 | 1.77e-16 |
| rs6543144 | 2 | 103092575 | G | A | 0.313 | 0.014 | 0.002 | 7.43e-10 |
| rs62160676 | 2 | 112167931 | C | T | 0.296 | -0.017 | 0.002 | 3.57e-13 |
| rs1427499 | 2 | 145400317 | G | A | 0.710 | 0.021 | 0.002 | 1.57e-20 |
| rs77785849 | 2 | 146580195 | A | C | 0.033 | 0.036 | 0.006 | 4.94e-10 |
| rs1598207 | 2 | 148757870 | G | A | 0.303 | 0.020 | 0.002 | 1.41e-19 |
| rs4602187 | 2 | 181834018 | T | C | 0.683 | 0.014 | 0.002 | 2.53e-10 |
| rs1449263 | 2 | 182319301 | T | C | 0.556 | 0.016 | 0.002 | 5.15e-14 |
| rs73987603 | 2 | 213844822 | A | G | 0.166 | 0.024 | 0.003 | 2.64e-18 |
| rs13419763 | 2 | 219134950 | T | C | 0.562 | 0.016 | 0.002 | 4.43e-15 |
| rs79140637 | 2 | 65084123 | A | G | 0.054 | -0.027 | 0.005 | 2.09e-09 |
| rs2028900 | 2 | 85767735 | T | C | 0.449 | -0.014 | 0.002 | 1.25e-11 |
| rs62105478 | 2 | 8735869 | A | G | 0.052 | -0.032 | 0.005 | 1.09e-11 |
| rs74535412 | 3 | 128297689 | A | G | 0.035 | 0.038 | 0.006 | 2.31e-11 |
| rs2335235 | 3 | 128315725 | C | G | 0.108 | -0.088 | 0.003 | 4.63e-157 |
| rs9819371 | 3 | 141206800 | T | C | 0.065 | -0.030 | 0.004 | 7.05e-13 |
| rs7613595 | 3 | 16908518 | C | A | 0.218 | -0.036 | 0.002 | 1.55e-48 |
| rs6780544 | 3 | 187424334 | A | G | 0.623 | -0.016 | 0.002 | 9.32e-15 |
| rs2089979 | 3 | 196501413 | G | A | 0.416 | -0.014 | 0.002 | 2.59e-11 |
| rs12497690 | 3 | 27795397 | C | A | 0.371 | 0.015 | 0.002 | 6.48e-12 |
| rs4324460 | 3 | 3120542 | G | T | 0.219 | 0.024 | 0.003 | 6.15e-22 |
| rs1669340 | 3 | 3198380 | T | G | 0.839 | -0.034 | 0.003 | 3.09e-34 |
| rs3181077 | 3 | 46250652 | T | C | 0.718 | -0.037 | 0.002 | 2.23e-59 |
| rs11097787 | 4 | 103407342 | T | C | 0.403 | -0.015 | 0.002 | 3.83e-12 |
| rs906149 | 4 | 83537777 | G | C | 0.221 | -0.019 | 0.002 | 5.16e-14 |
| rs56406125 | 4 | 87940205 | T | G | 0.190 | -0.035 | 0.003 | 6.04e-41 |
| rs2271352 | 5 | 126091428 | C | G | 0.226 | 0.042 | 0.002 | 1.44e-65 |
| rs449454 | 5 | 141533062 | G | A | 0.617 | 0.013 | 0.002 | 1.56e-09 |
| rs2594836 | 5 | 173205318 | A | G | 0.722 | -0.018 | 0.002 | 1.87e-14 |
| rs13188960 | 5 | 35853319 | T | G | 0.279 | -0.019 | 0.002 | 5.08e-17 |
| rs9376098 | 6 | 135499460 | A | T | 0.349 | 0.023 | 0.002 | 4.15e-27 |
| rs1205896 | 6 | 22341469 | A | G | 0.504 | 0.024 | 0.002 | 4.46e-32 |
| rs1029239 | 6 | 30138162 | G | C | 0.468 | -0.014 | 0.002 | 2.17e-12 |
| rs2308557 | 6 | 31239417 | T | C | 0.351 | 0.031 | 0.002 | 1.34e-46 |
| rs1883638 | 6 | 34830754 | T | C | 0.162 | -0.017 | 0.003 | 1.43e-09 |
| rs62393575 | 6 | 7058516 | C | G | 0.053 | -0.031 | 0.005 | 1.33e-11 |
| rs915125 | 6 | 82463376 | T | C | 0.281 | -0.029 | 0.002 | 2.18e-37 |
| rs905670 | 6 | 90958502 | A | G | 0.350 | -0.014 | 0.002 | 2.49e-11 |
| rs56179563 | 7 | 129685597 | A | G | 0.389 | 0.015 | 0.002 | 6.1e-12 |
| rs11772895 | 7 | 143081942 | C | G | 0.278 | 0.034 | 0.002 | 9.23e-50 |
| rs2158799 | 7 | 28277107 | G | C | 0.610 | 0.014 | 0.002 | 2.57e-11 |
| rs56388170 | 7 | 28724374 | T | G | 0.293 | 0.031 | 0.002 | 3.34e-43 |
| rs149007767 | 7 | 50370254 | T | C | 0.162 | 0.027 | 0.003 | 3.48e-20 |
| rs11768817 | 7 | 80223839 | G | A | 0.064 | 0.025 | 0.004 | 1.22e-09 |
| rs8179 | 7 | 92236164 | C | T | 0.791 | -0.028 | 0.003 | 2.75e-29 |
| rs445 | 7 | 92408370 | T | C | 0.097 | -0.076 | 0.003 | 2.69e-105 |
| rs4541868 | 8 | 106590705 | A | C | 0.276 | -0.019 | 0.002 | 2.61e-16 |
| rs7819602 | 8 | 10726842 | G | C | 0.613 | 0.016 | 0.002 | 1.98e-14 |
| rs4876400 | 8 | 119114110 | G | A | 0.626 | -0.017 | 0.002 | 6e-16 |
| rs7832357 | 8 | 126516197 | G | A | 0.342 | -0.015 | 0.002 | 8.9e-13 |
| rs9644063 | 8 | 22974450 | C | T | 0.822 | -0.024 | 0.003 | 1.49e-18 |
| rs45577137 | 8 | 48651633 | G | A | 0.045 | -0.045 | 0.005 | 2.08e-17 |
| rs16923637 | 8 | 59567885 | T | G | 0.286 | 0.014 | 0.002 | 4.79e-10 |
| rs2977799 | 8 | 6696927 | G | A | 0.509 | 0.027 | 0.002 | 1.17e-40 |
| rs2738104 | 8 | 6792257 | A | G | 0.672 | 0.018 | 0.002 | 4.01e-17 |
| rs55690609 | 8 | 6902043 | A | G | 0.323 | -0.035 | 0.002 | 8.93e-57 |
| rs2150052 | 9 | 113945067 | T | A | 0.506 | -0.013 | 0.002 | 8.76e-11 |
| rs1633768 | 9 | 135879138 | T | C | 0.275 | -0.017 | 0.002 | 7.02e-13 |
| rs4503179 | 9 | 21950879 | A | G | 0.724 | 0.017 | 0.002 | 3.77e-13 |
| rs12376511 | 9 | 22142756 | C | T | 0.163 | -0.021 | 0.003 | 1.03e-13 |
| rs7044519 | 9 | 82233842 | G | T | 0.161 | 0.017 | 0.003 | 2.37e-09 |
| rs2548992 | 5 | 131808668 | A | G | 0.267 | 0.014 | 0.002 | 4.65e-09 |
| rs2148204 | 10 | 102096354 | A | C | 0.364 | -0.017 | 0.002 | 2.86e-14 |
| rs12762973 | 10 | 960761 | G | C | 0.237 | 0.048 | 0.002 | 3.85e-83 |
| rs33931987 | 16 | 87903382 | G | A | 0.208 | 0.018 | 0.003 | 4.73e-11 |
| rs2384952 | 17 | 72744512 | C | T | 0.539 | -0.015 | 0.002 | 5.14e-12 |
| rs7249415 | 19 | 40223579 | A | C | 0.667 | -0.015 | 0.002 | 2.02e-11 |
| rs2967602 | 19 | 8569064 | C | T | 0.166 | -0.028 | 0.003 | 9.21e-23 |
| rs2926468 | 1 | 161612578 | C | T | 0.462 | -0.018 | 0.002 | 2.54e-16 |
| rs35076930 | 1 | 17677636 | T | C | 0.296 | 0.017 | 0.003 | 6.51e-11 |
| rs6692170 | 1 | 205074019 | A | G | 0.477 | -0.028 | 0.002 | 1.45e-41 |
| rs2009069 | 21 | 39596365 | T | C | 0.415 | 0.015 | 0.002 | 1.94e-12 |
| rs7576126 | 2 | 65658369 | A | G | 0.407 | 0.020 | 0.002 | 3.01e-21 |
| rs10212483 | 3 | 3160214 | A | G | 0.043 | -0.033 | 0.005 | 3.63e-10 |
| rs13063578 | 3 | 47087837 | A | T | 0.401 | 0.017 | 0.002 | 1.32e-14 |
| rs10023310 | 4 | 79629863 | C | T | 0.601 | 0.017 | 0.002 | 4.19e-15 |
| rs4481234 | 4 | 9956821 | C | T | 0.210 | -0.017 | 0.003 | 1.56e-10 |
| rs181548 | 5 | 150847783 | A | G | 0.386 | -0.014 | 0.002 | 3.99e-11 |
| rs111536497 | 5 | 98187510 | A | G | 0.091 | -0.027 | 0.004 | 6.8e-13 |
| rs9487032 | 6 | 109605323 | T | G | 0.484 | 0.022 | 0.002 | 2.61e-26 |
| rs111596698 | 6 | 32487123 | G | T | 0.262 | -0.025 | 0.003 | 2.35e-22 |
| rs72895231 | 6 | 82679396 | A | C | 0.596 | -0.016 | 0.002 | 2.99e-13 |
| rs2705789 | 7 | 75270073 | A | T | 0.459 | -0.024 | 0.002 | 2.9e-28 |
| rs140289677 | 7 | 8013293 | G | A | 0.058 | -0.039 | 0.005 | 9.58e-18 |
| rs74883958 | 10 | 1104030 | I | D | 0.644 | 0.014 | 0.002 | 5.1e-10 |

**Table S7. 350 SNPs significantly associated with neutrophil count used IVs in forward MR analyses derived from Vuckovic D et al.** Chr: Chromosome; EA: Effect allele; NEA: Non-effect allele; EAF: Effect allele frequency.

| SNP | Chr | Pos | EA | NEA | EAF | Beta | SE | P-value |
| --- | --- | --- | --- | --- | --- | --- | --- | --- |
| rs301819 | 1 | 8501786 | G | A | 0.588 | -0.014 | 0.002 | 3.5e-10 |
| rs6577536 | 1 | 8910110 | G | A | 0.509 | 0.025 | 0.002 | 1e-30 |
| rs284317 | 1 | 10731625 | G | A | 0.495 | 0.014 | 0.002 | 6.6e-11 |
| rs10917107 | 1 | 22329414 | A | G | 0.690 | -0.019 | 0.002 | 1.3e-16 |
| rs60918921 | 1 | 23850590 | A | C | 0.618 | -0.017 | 0.002 | 1.1e-14 |
| rs79567479 | 1 | 27011533 | C | T | 0.080 | -0.034 | 0.004 | 2.3e-18 |
| rs3762297 | 1 | 31231680 | T | C | 0.183 | 0.026 | 0.003 | 4.8e-21 |
| rs3917932 | 1 | 36943916 | G | C | 0.577 | -0.051 | 0.002 | 7e-121 |
| rs12139676 | 1 | 42201847 | A | T | 0.371 | -0.016 | 0.002 | 6.7e-13 |
| rs3754224 | 1 | 43423622 | C | T | 0.269 | -0.017 | 0.002 | 2.7e-12 |
| rs9429088 | 1 | 46497500 | A | T | 0.435 | -0.022 | 0.002 | 9.1e-24 |
| rs12121236 | 1 | 56617911 | A | C | 0.352 | 0.016 | 0.002 | 1.7e-12 |
| rs74076327 | 1 | 56979431 | C | T | 0.045 | -0.036 | 0.005 | 3.1e-12 |
| rs34293785 | 1 | 66137192 | C | T | 0.364 | -0.046 | 0.002 | 5.4e-93 |
| rs41313381 | 1 | 79411968 | A | C | 0.031 | 0.061 | 0.006 | 4.3e-24 |
| rs115490375 | 1 | 93427987 | G | A | 0.025 | 0.042 | 0.007 | 6.4e-10 |
| rs694180 | 1 | 111726213 | G | A | 0.658 | 0.018 | 0.002 | 1.5e-15 |
| rs6679677 | 1 | 114303808 | A | C | 0.101 | -0.037 | 0.004 | 1.1e-25 |
| rs140441527 | 1 | 150026632 | G | A | 0.093 | -0.025 | 0.004 | 2.3e-11 |
| rs501791 | 1 | 156089873 | T | C | 0.044 | -0.037 | 0.005 | 3.2e-12 |
| rs34599082 | 1 | 159175494 | T | C | 0.014 | -0.181 | 0.009 | 2.3e-86 |
| rs150041157 | 1 | 161566312 | G | C | 0.526 | 0.017 | 0.002 | 1.1e-13 |
| rs55970405 | 1 | 161617820 | C | G | 0.268 | -0.022 | 0.002 | 1.5e-18 |
| rs10918721 | 1 | 167568952 | G | T | 0.510 | 0.013 | 0.002 | 6.6e-10 |
| rs60124939 | 1 | 174075302 | T | C | 0.174 | -0.018 | 0.003 | 1.1e-10 |
| rs41272536 | 1 | 183440531 | G | A | 0.046 | -0.035 | 0.005 | 1.5e-11 |
| rs16843346 | 1 | 198543027 | T | C | 0.029 | -0.038 | 0.006 | 3.4e-09 |
| rs200639793 | 1 | 200304877 | A | G | 0.712 | 0.014 | 0.002 | 2.3e-09 |
| rs896319 | 1 | 205205651 | T | G | 0.912 | 0.027 | 0.004 | 1.4e-12 |
| rs12141901 | 1 | 208031234 | A | G | 0.242 | -0.023 | 0.002 | 1.5e-19 |
| rs1855829 | 1 | 221018832 | T | G | 0.316 | 0.018 | 0.002 | 2.7e-14 |
| rs10916617 | 1 | 224637070 | T | C | 0.212 | 0.026 | 0.003 | 2.4e-23 |
| rs7552783 | 1 | 227174145 | C | T | 0.512 | -0.024 | 0.002 | 1.6e-25 |
| rs486650 | 1 | 234819885 | T | C | 0.471 | -0.013 | 0.002 | 3.3e-10 |
| rs2065919 | 1 | 234928045 | G | C | 0.562 | 0.021 | 0.002 | 2.6e-20 |
| rs9970896 | 1 | 236104981 | T | A | 0.888 | -0.058 | 0.003 | 1.6e-65 |
| rs61838753 | 1 | 247569300 | C | A | 0.672 | -0.019 | 0.002 | 6.3e-17 |
| rs56188865 | 1 | 247606276 | C | T | 0.372 | -0.027 | 0.002 | 3.7e-34 |
| rs2113818 | 2 | 12890860 | C | T | 0.499 | 0.013 | 0.002 | 1.6e-09 |
| rs36101491 | 2 | 24387532 | T | C | 0.297 | 0.022 | 0.002 | 2.1e-20 |
| rs1866560 | 2 | 27328553 | G | T | 0.556 | -0.014 | 0.002 | 3.6e-10 |
| rs1260326 | 2 | 27730940 | C | T | 0.606 | -0.033 | 0.002 | 2.7e-52 |
| rs116447416 | 2 | 37501547 | G | A | 0.035 | -0.035 | 0.006 | 1.7e-09 |
| rs17030394 | 2 | 43354280 | G | A | 0.385 | -0.015 | 0.002 | 1.6e-11 |
| rs77552263 | 2 | 43786818 | A | G | 0.078 | 0.047 | 0.004 | 5.6e-32 |
| rs75475627 | 2 | 54787592 | G | C | 0.076 | 0.034 | 0.004 | 1.2e-16 |
| rs2421200 | 2 | 61711815 | T | G | 0.487 | -0.020 | 0.002 | 2.7e-20 |
| rs2028150 | 2 | 65655012 | G | C | 0.409 | 0.026 | 0.002 | 7.9e-32 |
| rs4599108 | 2 | 85543222 | T | C | 0.486 | -0.020 | 0.002 | 1.2e-20 |
| rs2309996 | 2 | 101735838 | G | T | 0.627 | 0.023 | 0.002 | 3.1e-26 |
| rs35789178 | 2 | 102603925 | G | T | 0.187 | 0.018 | 0.003 | 2.7e-11 |
| rs10181102 | 2 | 111835489 | C | T | 0.274 | 0.016 | 0.002 | 4.3e-11 |
| rs35898589 | 2 | 112915809 | G | A | 0.048 | 0.031 | 0.005 | 1.1e-09 |
| rs55709272 | 2 | 113867288 | C | T | 0.437 | 0.039 | 0.002 | 2.5e-73 |
| rs7604081 | 2 | 127866535 | C | T | 0.056 | 0.031 | 0.005 | 6e-11 |
| rs354703 | 2 | 143883952 | C | T | 0.593 | 0.015 | 0.002 | 2.3e-11 |
| rs3856364 | 2 | 145477217 | G | C | 0.681 | 0.016 | 0.002 | 2.6e-12 |
| rs188653407 | 2 | 160671563 | G | C | 0.023 | -0.056 | 0.007 | 4.5e-15 |
| rs12692567 | 2 | 160678144 | A | G | 0.833 | -0.037 | 0.003 | 8.4e-39 |
| rs2632372 | 2 | 169717541 | C | T | 0.504 | -0.021 | 0.002 | 9.1e-23 |
| rs7573465 | 2 | 182315885 | T | G | 0.557 | 0.022 | 0.002 | 1.5e-24 |
| rs13013264 | 2 | 192521279 | T | C | 0.072 | 0.027 | 0.004 | 1.5e-10 |
| rs79710932 | 2 | 218947121 | T | G | 0.077 | -0.032 | 0.004 | 1e-15 |
| rs114050631 | 2 | 219020958 | T | C | 0.011 | -0.159 | 0.011 | 2.2e-48 |
| rs1521134 | 2 | 219995186 | T | G | 0.643 | 0.013 | 0.002 | 3.4e-09 |
| rs79047930 | 2 | 220165527 | T | C | 0.015 | 0.053 | 0.009 | 2.6e-09 |
| rs6755895 | 2 | 232579795 | C | T | 0.227 | -0.018 | 0.003 | 8.7e-13 |
| rs10164769 | 2 | 237779229 | T | C | 0.740 | 0.027 | 0.002 | 7.1e-29 |
| rs56217149 | 3 | 12962914 | A | G | 0.151 | 0.018 | 0.003 | 2e-09 |
| rs4682867 | 3 | 42905465 | T | C | 0.383 | -0.022 | 0.002 | 1.5e-23 |
| rs11920354 | 3 | 47262246 | A | C | 0.388 | -0.016 | 0.002 | 4.5e-12 |
| rs6779340 | 3 | 58033701 | G | C | 0.336 | -0.017 | 0.002 | 3.5e-13 |
| rs9842724 | 3 | 63804761 | C | T | 0.803 | -0.016 | 0.003 | 3.9e-09 |
| rs12487658 | 3 | 71528943 | T | C | 0.802 | 0.019 | 0.003 | 1.2e-12 |
| rs4504118 | 3 | 98422588 | C | G | 0.556 | 0.013 | 0.002 | 1.3e-09 |
| rs191832862 | 3 | 107303068 | G | A | 0.169 | -0.025 | 0.003 | 1.1e-17 |
| rs3732360 | 3 | 119536581 | T | C | 0.747 | 0.017 | 0.002 | 4.1e-12 |
| rs6782812 | 3 | 128317997 | A | G | 0.893 | -0.065 | 0.003 | 2.7e-77 |
| rs6764912 | 3 | 128372487 | A | G | 0.067 | -0.039 | 0.004 | 1.7e-18 |
| rs113799399 | 3 | 140926791 | T | C | 0.126 | 0.034 | 0.003 | 8.1e-25 |
| rs9819371 | 3 | 141206800 | T | C | 0.065 | -0.046 | 0.004 | 1.8e-26 |
| rs55730982 | 3 | 156797208 | G | T | 0.401 | 0.014 | 0.002 | 1.2e-10 |
| rs10936588 | 3 | 169319801 | A | G | 0.634 | -0.013 | 0.002 | 1.6e-09 |
| rs4074672 | 3 | 183730295 | T | C | 0.369 | 0.018 | 0.002 | 7.5e-16 |
| rs789858 | 3 | 194405966 | T | C | 0.403 | 0.015 | 0.002 | 3.5e-11 |
| rs7626444 | 3 | 196504902 | C | G | 0.423 | -0.022 | 0.002 | 1.9e-23 |
| rs35734242 | 4 | 706700 | C | T | 0.428 | 0.024 | 0.002 | 6.6e-29 |
| rs73191188 | 4 | 3105200 | A | G | 0.353 | -0.016 | 0.002 | 1.5e-12 |
| rs6831368 | 4 | 6969919 | G | A | 0.362 | -0.023 | 0.002 | 4.6e-24 |
| rs28530750 | 4 | 36312542 | A | G | 0.043 | 0.061 | 0.005 | 7.3e-30 |
| rs218264 | 4 | 55408875 | T | A | 0.249 | 0.034 | 0.002 | 8.5e-43 |
| rs723585 | 4 | 55503194 | G | A | 0.486 | -0.022 | 0.002 | 2e-25 |
| rs2412771 | 4 | 57761417 | C | T | 0.417 | -0.015 | 0.002 | 3.8e-12 |
| rs2201124 | 4 | 72597009 | T | C | 0.332 | -0.022 | 0.002 | 1.4e-21 |
| rs16850073 | 4 | 74703999 | T | C | 0.375 | 0.053 | 0.002 | 8.9e-124 |
| rs146149115 | 4 | 74768492 | G | A | 0.010 | -0.090 | 0.011 | 1.3e-15 |
| rs16850408 | 4 | 74932807 | A | C | 0.369 | 0.069 | 0.002 | 1e-200 |
| rs17049930 | 4 | 105579471 | C | T | 0.028 | 0.044 | 0.007 | 1.5e-11 |
| rs7679673 | 4 | 106061534 | A | C | 0.376 | -0.025 | 0.002 | 1.3e-28 |
| rs4145952 | 4 | 120155806 | A | C | 0.401 | -0.014 | 0.002 | 1.2e-10 |
| rs11735662 | 4 | 145026126 | T | C | 0.034 | 0.049 | 0.006 | 1.2e-16 |
| rs2290846 | 4 | 151199080 | A | G | 0.287 | 0.018 | 0.002 | 8.2e-15 |
| rs6817881 | 4 | 152223904 | T | C | 0.500 | 0.013 | 0.002 | 1.5e-09 |
| rs4535497 | 5 | 1107428 | A | C | 0.570 | -0.015 | 0.002 | 4.7e-12 |
| rs7705526 | 5 | 1285974 | A | C | 0.326 | 0.035 | 0.002 | 2.4e-50 |
| rs11744663 | 5 | 57315635 | A | G | 0.192 | -0.017 | 0.003 | 3.7e-10 |
| rs11741775 | 5 | 68590395 | T | C | 0.443 | -0.026 | 0.002 | 1.8e-33 |
| rs71576926 | 5 | 68828803 | A | T | 0.784 | 0.016 | 0.003 | 3.3e-09 |
| rs4703890 | 5 | 71748624 | A | G | 0.882 | -0.044 | 0.003 | 1.2e-40 |
| rs716848 | 5 | 96265000 | A | C | 0.436 | 0.014 | 0.002 | 2.2e-11 |
| rs141543178 | 5 | 108133865 | T | A | 0.026 | 0.041 | 0.007 | 3.6e-09 |
| rs257063 | 5 | 114806819 | T | C | 0.743 | -0.015 | 0.002 | 2.5e-09 |
| rs4836224 | 5 | 122092875 | C | A | 0.426 | 0.014 | 0.002 | 2.3e-10 |
| rs2631367 | 5 | 131705458 | G | C | 0.522 | 0.024 | 0.002 | 4.7e-30 |
| rs329117 | 5 | 133860101 | C | T | 0.582 | 0.014 | 0.002 | 8.9e-11 |
| rs6580229 | 5 | 141510754 | G | A | 0.617 | 0.026 | 0.002 | 9.9e-32 |
| rs2082382 | 5 | 148200553 | A | G | 0.548 | -0.031 | 0.002 | 1.4e-47 |
| rs284440 | 5 | 156170215 | G | T | 0.837 | -0.021 | 0.003 | 4.1e-13 |
| rs2561758 | 5 | 173205282 | G | A | 0.723 | -0.033 | 0.002 | 8.9e-43 |
| rs13190036 | 5 | 176737720 | G | A | 0.892 | -0.022 | 0.003 | 1.6e-10 |
| rs1134924 | 5 | 179126090 | T | C | 0.506 | -0.017 | 0.002 | 4.3e-15 |
| rs155784 | 5 | 179354966 | G | A | 0.353 | 0.014 | 0.002 | 2.8e-10 |
| rs61025394 | 6 | 7085817 | A | G | 0.211 | -0.022 | 0.003 | 7.9e-17 |
| rs144721194 | 6 | 7223858 | G | T | 0.023 | -0.043 | 0.007 | 2.6e-09 |
| rs3777755 | 6 | 12159699 | T | C | 0.310 | 0.015 | 0.002 | 1.8e-10 |
| rs1144700 | 6 | 16744687 | T | C | 0.179 | -0.033 | 0.003 | 7.1e-32 |
| rs4712614 | 6 | 21382765 | G | T | 0.622 | -0.022 | 0.002 | 3.2e-23 |
| rs141928748 | 6 | 28296315 | G | C | 0.045 | -0.032 | 0.005 | 3.7e-10 |
| rs9261145 | 6 | 29984865 | T | C | 0.098 | 0.028 | 0.004 | 3.8e-15 |
| rs144626001 | 6 | 31374671 | T | C | 0.019 | -0.049 | 0.008 | 7.6e-10 |
| rs4713470 | 6 | 31472821 | C | T | 0.312 | 0.046 | 0.002 | 2e-83 |
| rs200801362 | 6 | 31555480 | C | T | 0.161 | -0.060 | 0.004 | 6.3e-51 |
| rs138408209 | 6 | 32537509 | T | A | 0.090 | -0.039 | 0.005 | 6.5e-16 |
| rs9271578 | 6 | 32590635 | T | A | 0.502 | 0.063 | 0.002 | 5.6e-190 |
| rs9274305 | 6 | 32632000 | T | C | 0.205 | -0.038 | 0.003 | 6.7e-40 |
| rs16895923 | 6 | 42633368 | G | A | 0.167 | 0.026 | 0.003 | 3.2e-19 |
| rs729761 | 6 | 43804571 | G | T | 0.711 | 0.017 | 0.002 | 2.5e-12 |
| rs915125 | 6 | 82463376 | T | C | 0.279 | -0.017 | 0.002 | 6.3e-13 |
| rs9362415 | 6 | 87968565 | G | A | 0.490 | -0.027 | 0.002 | 6.1e-36 |
| rs207253 | 6 | 90808423 | A | G | 0.408 | -0.014 | 0.002 | 2.1e-09 |
| rs364663 | 6 | 105443189 | A | T | 0.557 | -0.013 | 0.002 | 3.3e-09 |
| rs6927569 | 6 | 109621494 | C | T | 0.523 | 0.027 | 0.002 | 2.7e-37 |
| rs1490384 | 6 | 126851160 | T | C | 0.500 | -0.015 | 0.002 | 1.6e-12 |
| rs35786788 | 6 | 135419042 | A | G | 0.253 | -0.036 | 0.003 | 6e-47 |
| rs62431534 | 6 | 135832523 | G | C | 0.544 | 0.020 | 0.002 | 1.1e-19 |
| rs6924387 | 6 | 137082948 | G | A | 0.410 | 0.017 | 0.002 | 1.2e-14 |
| rs72978754 | 6 | 138053203 | C | T | 0.066 | -0.027 | 0.004 | 6.2e-10 |
| rs9390461 | 6 | 147701217 | G | A | 0.540 | 0.017 | 0.002 | 5.1e-16 |
| rs212409 | 6 | 159470058 | A | G | 0.553 | -0.022 | 0.002 | 1.6e-23 |
| rs4724795 | 7 | 6377836 | A | G | 0.291 | -0.015 | 0.002 | 5.2e-10 |
| rs6954012 | 7 | 6755724 | C | A | 0.112 | 0.026 | 0.003 | 5.2e-14 |
| rs56195338 | 7 | 8107922 | A | G | 0.058 | -0.039 | 0.005 | 3e-17 |
| rs7776857 | 7 | 22754768 | T | G | 0.655 | -0.015 | 0.002 | 1.4e-11 |
| rs2158799 | 7 | 28277107 | G | C | 0.610 | 0.052 | 0.002 | 8.7e-125 |
| rs56388170 | 7 | 28724374 | T | G | 0.292 | 0.071 | 0.002 | 5.3e-195 |
| rs2710804 | 7 | 36084529 | C | T | 0.377 | 0.021 | 0.002 | 4.2e-22 |
| rs1557934 | 7 | 45022257 | C | T | 0.843 | 0.026 | 0.003 | 8.9e-19 |
| rs10252457 | 7 | 47337530 | G | A | 0.425 | -0.013 | 0.002 | 3.9e-09 |
| rs1456896 | 7 | 50304461 | T | C | 0.667 | 0.024 | 0.002 | 2.1e-26 |
| rs149007767 | 7 | 50370254 | T | C | 0.162 | 0.031 | 0.003 | 7.4e-25 |
| rs1540652 | 7 | 65650277 | C | T | 0.890 | -0.027 | 0.004 | 3.6e-14 |
| rs4718353 | 7 | 65941448 | A | G | 0.045 | -0.034 | 0.005 | 3e-11 |
| rs62466318 | 7 | 73042085 | T | C | 0.203 | -0.024 | 0.003 | 5.3e-19 |
| rs42033 | 7 | 92237533 | T | A | 0.208 | 0.041 | 0.003 | 1.3e-54 |
| rs445 | 7 | 92408370 | T | C | 0.096 | -0.097 | 0.004 | 2e-156 |
| rs6465668 | 7 | 97961469 | C | T | 0.482 | -0.015 | 0.002 | 1e-12 |
| rs342242 | 7 | 106338989 | C | T | 0.454 | -0.016 | 0.002 | 3.2e-14 |
| rs11556924 | 7 | 129663496 | T | C | 0.389 | 0.016 | 0.002 | 3e-13 |
| rs2015210 | 7 | 130737982 | G | T | 0.751 | -0.021 | 0.002 | 1.8e-17 |
| rs13231262 | 7 | 148886058 | T | C | 0.656 | -0.020 | 0.002 | 2.1e-19 |
| rs2977806 | 8 | 6692968 | T | C | 0.531 | 0.013 | 0.002 | 5.1e-10 |
| rs55690609 | 8 | 6902043 | A | G | 0.322 | -0.014 | 0.002 | 3.4e-10 |
| rs2126260 | 8 | 9185081 | C | T | 0.798 | 0.017 | 0.003 | 1.1e-10 |
| rs11781592 | 8 | 10511121 | A | G | 0.572 | 0.025 | 0.002 | 4.6e-30 |
| rs12550612 | 8 | 22966769 | A | G | 0.824 | -0.032 | 0.003 | 1.3e-29 |
| rs2979489 | 8 | 30280833 | A | G | 0.742 | 0.021 | 0.002 | 9.1e-18 |
| rs45577137 | 8 | 48651633 | G | A | 0.045 | 0.037 | 0.006 | 1.3e-11 |
| rs28571765 | 8 | 55449281 | C | T | 0.209 | -0.022 | 0.003 | 1.4e-16 |
| rs7816785 | 8 | 56797418 | T | C | 0.606 | -0.021 | 0.002 | 1.3e-20 |
| rs609264 | 8 | 61548546 | G | T | 0.528 | 0.013 | 0.002 | 1.7e-09 |
| rs7846314 | 8 | 61650831 | T | A | 0.187 | 0.067 | 0.003 | 6.5e-130 |
| rs34685450 | 8 | 61916962 | A | T | 0.253 | -0.015 | 0.003 | 4.1e-09 |
| rs57039640 | 8 | 68864171 | T | C | 0.334 | -0.019 | 0.002 | 2.8e-16 |
| rs62510269 | 8 | 79012343 | G | A | 0.147 | -0.029 | 0.003 | 6.9e-21 |
| rs10808538 | 8 | 87169063 | A | G | 0.297 | 0.014 | 0.002 | 2.7e-09 |
| rs2679741 | 8 | 103881011 | G | C | 0.366 | -0.020 | 0.002 | 1.3e-19 |
| rs4734879 | 8 | 106583124 | G | A | 0.275 | -0.021 | 0.002 | 7.2e-18 |
| rs2980888 | 8 | 126507308 | C | T | 0.699 | 0.023 | 0.002 | 1.7e-23 |
| rs59697075 | 8 | 130618150 | T | C | 0.588 | -0.038 | 0.002 | 1.6e-66 |
| rs145209947 | 8 | 130694185 | A | C | 0.015 | -0.069 | 0.010 | 2e-12 |
| rs7005996 | 8 | 142241681 | T | C | 0.906 | 0.024 | 0.004 | 6.3e-11 |
| rs34634548 | 8 | 142329460 | A | C | 0.358 | -0.028 | 0.002 | 1.1e-35 |
| rs2992836 | 9 | 276053 | T | G | 0.188 | 0.027 | 0.003 | 3.8e-22 |
| rs2219143 | 9 | 2622278 | A | G | 0.393 | 0.020 | 0.002 | 3.3e-19 |
| rs385893 | 9 | 4763176 | C | T | 0.524 | 0.027 | 0.002 | 5.3e-36 |
| rs10758669 | 9 | 4981602 | A | C | 0.659 | -0.015 | 0.002 | 2.9e-11 |
| rs680775 | 9 | 79322674 | G | A | 0.759 | -0.020 | 0.003 | 1.7e-15 |
| rs696825 | 9 | 86583076 | T | C | 0.253 | -0.022 | 0.002 | 4.5e-19 |
| rs116852611 | 9 | 112745176 | T | A | 0.135 | 0.020 | 0.003 | 2e-10 |
| rs7866863 | 9 | 114657707 | A | G | 0.343 | 0.014 | 0.002 | 3e-10 |
| rs78610102 | 9 | 116039668 | T | G | 0.225 | -0.020 | 0.003 | 3.6e-15 |
| rs10986338 | 9 | 127191232 | A | G | 0.651 | -0.015 | 0.002 | 2.6e-11 |
| rs2519093 | 9 | 136141870 | T | C | 0.184 | -0.041 | 0.003 | 1.4e-49 |
| rs2810489 | 9 | 136922700 | A | G | 0.258 | 0.021 | 0.002 | 8.7e-18 |
| rs11145986 | 9 | 139319847 | G | A | 0.290 | 0.033 | 0.002 | 1.8e-45 |
| rs9329341 | 10 | 13532447 | A | G | 0.580 | 0.019 | 0.002 | 7.9e-18 |
| rs56278466 | 10 | 17875857 | G | T | 0.662 | -0.014 | 0.002 | 2e-09 |
| rs692594 | 10 | 18265893 | C | G | 0.491 | 0.015 | 0.002 | 8.1e-12 |
| rs10828722 | 10 | 25199951 | G | A | 0.370 | -0.040 | 0.002 | 4.3e-73 |
| rs2807742 | 10 | 28781367 | A | G | 0.770 | 0.034 | 0.003 | 8e-41 |
| rs72790861 | 10 | 44880207 | G | C | 0.308 | -0.020 | 0.002 | 6.8e-18 |
| rs2393969 | 10 | 65140440 | C | A | 0.471 | -0.029 | 0.002 | 1.1e-41 |
| rs10997821 | 10 | 69570691 | G | A | 0.104 | -0.024 | 0.004 | 4.4e-12 |
| rs3747869 | 10 | 73520632 | C | A | 0.901 | 0.032 | 0.004 | 1.4e-19 |
| rs2785072 | 10 | 89765388 | T | A | 0.168 | -0.022 | 0.003 | 1.2e-12 |
| rs1412445 | 10 | 91002804 | T | C | 0.338 | 0.021 | 0.002 | 1.1e-20 |
| rs9633675 | 10 | 96210400 | C | G | 0.478 | -0.019 | 0.002 | 3e-18 |
| rs10882895 | 10 | 99075300 | G | A | 0.597 | 0.044 | 0.002 | 3.1e-90 |
| rs4556473 | 10 | 104454700 | C | T | 0.384 | -0.014 | 0.002 | 9.2e-10 |
| rs76863268 | 10 | 111995452 | A | T | 0.344 | -0.016 | 0.002 | 3.9e-12 |
| rs151595 | 10 | 115719765 | C | T | 0.628 | -0.014 | 0.002 | 3.9e-10 |
| rs3781454 | 10 | 126348565 | A | G | 0.681 | 0.024 | 0.002 | 2.1e-25 |
| rs14408 | 11 | 308314 | C | T | 0.368 | 0.050 | 0.002 | 1.9e-110 |
| rs1468102 | 11 | 3004526 | G | C | 0.306 | 0.018 | 0.002 | 2.5e-14 |
| rs899013 | 11 | 10476689 | G | A | 0.596 | 0.013 | 0.002 | 4.2e-09 |
| rs1159649 | 11 | 12135604 | G | C | 0.316 | -0.014 | 0.002 | 1.9e-09 |
| rs10765999 | 11 | 12859917 | T | C | 0.677 | -0.014 | 0.002 | 3.6e-09 |
| rs55876153 | 11 | 47416636 | A | G | 0.323 | 0.020 | 0.002 | 2.2e-18 |
| rs7396753 | 11 | 60021508 | C | T | 0.592 | 0.015 | 0.002 | 2.2e-12 |
| rs174544 | 11 | 61567753 | A | C | 0.308 | -0.026 | 0.002 | 9.3e-30 |
| rs545500 | 11 | 65629934 | C | G | 0.667 | 0.017 | 0.002 | 1.9e-13 |
| rs55904328 | 11 | 76267477 | A | G | 0.314 | -0.014 | 0.002 | 2.4e-09 |
| rs7129527 | 11 | 108044995 | G | A | 0.411 | 0.017 | 0.002 | 4.1e-15 |
| rs1791807 | 11 | 113954234 | T | C | 0.415 | -0.018 | 0.002 | 3e-16 |
| rs238914 | 11 | 113984109 | A | C | 0.396 | 0.029 | 0.002 | 3.9e-39 |
| rs1715429 | 11 | 118083664 | G | A | 0.753 | 0.022 | 0.002 | 4.6e-18 |
| rs1783921 | 11 | 128094803 | C | T | 0.205 | 0.018 | 0.003 | 1.7e-11 |
| rs8705 | 11 | 128328913 | A | G | 0.318 | -0.020 | 0.002 | 7.9e-18 |
| rs632887 | 12 | 3392351 | G | A | 0.409 | 0.016 | 0.002 | 1.4e-12 |
| rs11611647 | 12 | 4333919 | C | T | 0.210 | -0.024 | 0.003 | 8.7e-20 |
| rs2280503 | 12 | 51138687 | C | A | 0.344 | 0.019 | 0.002 | 8e-18 |
| rs1700159 | 12 | 52305786 | T | C | 0.772 | 0.021 | 0.003 | 5.5e-16 |
| rs1245035 | 12 | 64976049 | A | C | 0.629 | 0.017 | 0.002 | 6.7e-14 |
| rs7968902 | 12 | 66363070 | G | T | 0.570 | 0.014 | 0.002 | 9e-10 |
| rs4761234 | 12 | 69732105 | C | T | 0.485 | 0.023 | 0.002 | 6.4e-26 |
| rs7487314 | 12 | 88836215 | T | G | 0.701 | -0.025 | 0.002 | 6.3e-27 |
| rs17041439 | 12 | 101873240 | C | A | 0.056 | 0.027 | 0.005 | 4.3e-09 |
| rs3184504 | 12 | 111884608 | C | T | 0.519 | -0.031 | 0.002 | 3.1e-47 |
| rs11064881 | 12 | 120146925 | A | G | 0.073 | -0.035 | 0.004 | 8.3e-18 |
| rs610578 | 12 | 121194565 | G | A | 0.662 | -0.014 | 0.002 | 2.3e-09 |
| rs61955089 | 12 | 123851372 | C | T | 0.026 | 0.045 | 0.007 | 1.2e-11 |
| rs117808697 | 13 | 28513538 | T | A | 0.073 | 0.034 | 0.005 | 1.2e-13 |
| rs2504235 | 13 | 28612886 | G | A | 0.638 | 0.026 | 0.002 | 1.6e-30 |
| rs17521426 | 13 | 42868208 | A | C | 0.238 | 0.022 | 0.003 | 3.5e-18 |
| rs7326825 | 13 | 50113450 | A | G | 0.707 | 0.021 | 0.002 | 2e-19 |
| rs11841945 | 13 | 73630059 | C | G | 0.579 | 0.013 | 0.002 | 1.1e-09 |
| rs150861794 | 13 | 109003805 | T | C | 0.018 | -0.053 | 0.009 | 7e-10 |
| rs2260766 | 13 | 114186800 | G | A | 0.281 | 0.024 | 0.002 | 3.1e-24 |
| rs2038700 | 14 | 25461989 | C | T | 0.394 | 0.034 | 0.002 | 6.5e-54 |
| rs72664840 | 14 | 35596323 | T | C | 0.178 | 0.024 | 0.003 | 2.2e-17 |
| rs72731564 | 14 | 69280158 | T | C | 0.186 | 0.020 | 0.003 | 1.4e-12 |
| rs12884741 | 14 | 69870944 | G | A | 0.065 | 0.044 | 0.004 | 9.6e-24 |
| rs2205190 | 14 | 77886723 | G | T | 0.521 | -0.016 | 0.002 | 5.4e-13 |
| rs9806027 | 14 | 81668707 | C | G | 0.554 | -0.018 | 0.002 | 4.3e-17 |
| rs10498635 | 14 | 93103309 | T | C | 0.184 | -0.029 | 0.003 | 1.9e-25 |
| rs12588718 | 14 | 101149605 | C | G | 0.672 | 0.015 | 0.002 | 4e-11 |
| rs2494748 | 14 | 105258892 | T | C | 0.616 | 0.013 | 0.002 | 1.8e-09 |
| rs4983590 | 14 | 105646179 | A | G | 0.609 | -0.017 | 0.002 | 3.7e-14 |
| rs4924450 | 15 | 40597229 | A | G | 0.704 | 0.015 | 0.002 | 6.6e-10 |
| rs7183879 | 15 | 42260456 | G | A | 0.111 | -0.045 | 0.003 | 8e-39 |
| rs2663553 | 15 | 51207818 | C | G | 0.540 | 0.020 | 0.002 | 3.4e-21 |
| rs7180079 | 15 | 64629873 | G | A | 0.879 | 0.032 | 0.003 | 2e-22 |
| rs7496362 | 15 | 65758874 | G | C | 0.366 | 0.017 | 0.002 | 1.7e-14 |
| rs67538199 | 15 | 75361650 | C | T | 0.161 | -0.022 | 0.003 | 1.1e-13 |
| rs8024737 | 15 | 77249703 | C | T | 0.542 | 0.013 | 0.002 | 5e-10 |
| rs72756503 | 15 | 86062567 | T | C | 0.147 | -0.018 | 0.003 | 3.7e-09 |
| rs7183988 | 15 | 91428589 | G | T | 0.526 | -0.017 | 0.002 | 9.2e-16 |
| rs67175901 | 15 | 101748227 | T | C | 0.108 | 0.045 | 0.003 | 4.1e-39 |
| rs11647740 | 16 | 1347862 | C | T | 0.593 | -0.018 | 0.002 | 8.3e-16 |
| rs35929659 | 16 | 2165630 | C | T | 0.180 | -0.019 | 0.003 | 2.3e-11 |
| rs6500550 | 16 | 3746241 | T | C | 0.302 | -0.024 | 0.002 | 9e-24 |
| rs16958642 | 16 | 11864032 | A | G | 0.083 | 0.025 | 0.004 | 1.8e-10 |
| rs61739285 | 16 | 27480797 | T | C | 0.034 | -0.042 | 0.006 | 1.1e-12 |
| rs12716977 | 16 | 30482803 | C | T | 0.517 | 0.020 | 0.002 | 3.7e-21 |
| rs1362623 | 16 | 49885785 | T | C | 0.210 | 0.019 | 0.003 | 2.5e-12 |
| rs74250734 | 16 | 50212589 | G | T | 0.095 | -0.025 | 0.004 | 2.6e-11 |
| rs11648664 | 16 | 53122486 | A | G | 0.404 | -0.014 | 0.002 | 8.6e-11 |
| rs36026517 | 16 | 74597758 | C | G | 0.198 | 0.023 | 0.003 | 2.8e-17 |
| rs8045100 | 16 | 81603400 | T | C | 0.541 | 0.020 | 0.002 | 7.7e-21 |
| rs305082 | 16 | 85936978 | C | T | 0.171 | 0.037 | 0.003 | 3.1e-38 |
| rs72803323 | 16 | 89373324 | T | C | 0.040 | 0.034 | 0.005 | 3.2e-10 |
| rs9905106 | 17 | 1373518 | C | T | 0.735 | 0.022 | 0.002 | 1e-19 |
| rs7225843 | 17 | 2001825 | C | T | 0.203 | -0.023 | 0.003 | 1.4e-17 |
| rs3213878 | 17 | 5039138 | T | C | 0.106 | 0.022 | 0.003 | 4e-10 |
| rs12936529 | 17 | 16168784 | T | C | 0.476 | -0.037 | 0.002 | 4.3e-66 |
| rs11653826 | 17 | 27653016 | T | C | 0.099 | 0.026 | 0.004 | 9e-13 |
| rs2617884 | 17 | 28120430 | C | G | 0.497 | -0.020 | 0.002 | 3.1e-20 |
| rs11651596 | 17 | 38056116 | C | T | 0.468 | -0.066 | 0.002 | 1e-200 |
| rs576566496 | 17 | 38140269 | A | G | 0.027 | 0.091 | 0.007 | 6.1e-39 |
| rs74725931 | 17 | 38196327 | C | T | 0.038 | 0.063 | 0.006 | 1e-27 |
| rs563136944 | 17 | 38470429 | G | T | 0.356 | -0.028 | 0.003 | 6.6e-27 |
| rs184482331 | 17 | 40702760 | T | C | 0.079 | 0.024 | 0.004 | 3.5e-09 |
| rs11656151 | 17 | 44068492 | G | A | 0.239 | -0.016 | 0.003 | 1.4e-10 |
| rs138284624 | 17 | 56385573 | T | C | 0.013 | 0.130 | 0.010 | 4.7e-42 |
| rs529080407 | 17 | 57252243 | C | A | 0.043 | 0.045 | 0.007 | 1.8e-11 |
| rs11650106 | 17 | 57850251 | T | C | 0.545 | 0.031 | 0.002 | 4.7e-48 |
| rs9916458 | 17 | 72688028 | A | G | 0.782 | 0.018 | 0.003 | 2.3e-12 |
| rs2384952 | 17 | 72744512 | C | T | 0.539 | -0.017 | 0.002 | 1.2e-14 |
| rs2240775 | 17 | 74710128 | C | T | 0.471 | -0.015 | 0.002 | 2.3e-12 |
| rs72901753 | 17 | 76244283 | C | G | 0.326 | -0.015 | 0.002 | 1.5e-10 |
| rs4468717 | 18 | 3457606 | T | C | 0.077 | -0.028 | 0.004 | 3.2e-12 |
| rs7235882 | 18 | 19684119 | T | C | 0.092 | 0.024 | 0.004 | 5.8e-11 |
| rs303753 | 18 | 21074922 | A | G | 0.346 | -0.023 | 0.002 | 1.1e-23 |
| rs78285907 | 18 | 21622003 | T | A | 0.089 | 0.023 | 0.004 | 4.1e-09 |
| rs9965539 | 18 | 41973779 | A | G | 0.133 | 0.033 | 0.003 | 1.4e-25 |
| rs8093990 | 18 | 42122473 | C | A | 0.432 | 0.014 | 0.002 | 1.8e-10 |
| rs55874505 | 18 | 43816804 | G | T | 0.584 | -0.019 | 0.002 | 1.5e-16 |
| rs8098724 | 18 | 48125899 | A | G | 0.212 | -0.018 | 0.003 | 9.3e-12 |
| rs66723169 | 18 | 57808978 | A | C | 0.230 | -0.015 | 0.003 | 2.2e-09 |
| rs1108167 | 18 | 60163025 | G | A | 0.286 | -0.018 | 0.002 | 7.7e-14 |
| rs72973714 | 18 | 74076020 | T | C | 0.067 | -0.039 | 0.004 | 1.4e-19 |
| rs930232 | 19 | 1036018 | A | G | 0.460 | 0.022 | 0.002 | 3.1e-24 |
| rs539088769 | 19 | 3228641 | G | T | 0.065 | -0.039 | 0.005 | 1.4e-13 |
| rs263067 | 19 | 5028094 | C | A | 0.664 | 0.017 | 0.002 | 2.8e-13 |
| rs571497 | 19 | 7827830 | A | G | 0.153 | -0.039 | 0.003 | 8.5e-39 |
| rs2116942 | 19 | 10334663 | G | T | 0.604 | -0.022 | 0.002 | 3.4e-23 |
| rs11085767 | 19 | 11370547 | T | C | 0.164 | 0.018 | 0.003 | 1.4e-09 |
| rs1985157 | 19 | 18513594 | C | T | 0.411 | 0.026 | 0.002 | 1.5e-33 |
| rs58434384 | 19 | 19786099 | G | A | 0.085 | 0.032 | 0.004 | 6.8e-17 |
| rs4805881 | 19 | 33896432 | C | A | 0.665 | -0.014 | 0.002 | 8.9e-10 |
| rs4760 | 19 | 44153100 | G | A | 0.156 | -0.075 | 0.003 | 3.2e-142 |
| rs11673093 | 19 | 45742094 | A | G | 0.260 | -0.047 | 0.002 | 7.3e-83 |
| rs386243 | 19 | 49127490 | T | C | 0.240 | -0.026 | 0.003 | 8.1e-26 |
| rs2243602 | 20 | 1546742 | A | T | 0.563 | -0.014 | 0.002 | 7.3e-11 |
| rs633284 | 20 | 2904143 | T | A | 0.516 | 0.016 | 0.002 | 4.9e-13 |
| rs579111 | 20 | 3763872 | C | T | 0.636 | -0.013 | 0.002 | 4.2e-09 |
| rs2294256 | 20 | 8174058 | T | A | 0.482 | -0.016 | 0.002 | 2.5e-13 |
| rs6029234 | 20 | 39259278 | C | G | 0.626 | 0.017 | 0.002 | 1.3e-14 |
| rs1800961 | 20 | 43042364 | T | C | 0.031 | -0.056 | 0.006 | 2e-19 |
| rs143003731 | 20 | 47936696 | T | C | 0.008 | -0.086 | 0.013 | 9.3e-12 |
| rs6011066 | 20 | 62364180 | A | G | 0.674 | 0.015 | 0.002 | 1.5e-11 |
| rs62239920 | 21 | 18944889 | A | G | 0.203 | -0.017 | 0.003 | 3.1e-10 |
| rs9977672 | 21 | 40463283 | A | G | 0.258 | -0.024 | 0.002 | 4.2e-22 |
| rs35990176 | 21 | 44472118 | C | A | 0.449 | 0.013 | 0.002 | 3.9e-09 |
| rs73377344 | 21 | 47804358 | C | T | 0.554 | -0.014 | 0.002 | 1.7e-10 |
| rs5994158 | 22 | 17567898 | G | A | 0.807 | -0.017 | 0.003 | 8.6e-10 |
| rs5746451 | 22 | 18126020 | C | T | 0.504 | 0.022 | 0.002 | 2.2e-25 |
| rs5746638 | 22 | 18907997 | T | C | 0.285 | -0.014 | 0.002 | 2.9e-09 |
| rs35955747 | 22 | 31813058 | T | A | 0.539 | 0.019 | 0.002 | 8.7e-18 |
| rs139402 | 22 | 39546145 | C | T | 0.438 | -0.014 | 0.002 | 3.3e-10 |

**Table S8. 383 SNPs significantly associated with neutrophil count used IVs in forward MR analyses derived from Chen MH et al.** Chr: Chromosome; EA: Effect allele; NEA: Non-effect allele; EAF: Effect allele frequency.

| SNP | Chr | Pos | EA | NEA | EAF | Beta | SE | P-value |
| --- | --- | --- | --- | --- | --- | --- | --- | --- |
| rs7917772 | 10 | 104487443 | A | G | 0.631 | 0.016 | 0.002 | 5.41e-16 |
| rs10509912 | 10 | 112029405 | A | T | 0.135 | -0.021 | 0.003 | 7.02e-14 |
| rs72836628 | 10 | 113902454 | T | C | 0.248 | -0.015 | 0.002 | 4.07e-12 |
| rs180941 | 10 | 115720674 | A | G | 0.625 | -0.013 | 0.002 | 8.33e-11 |
| rs3781454 | 10 | 126348565 | A | G | 0.678 | 0.024 | 0.002 | 6.26e-31 |
| rs9419387 | 10 | 133758358 | A | G | 0.405 | -0.012 | 0.002 | 2.66e-10 |
| rs10906393 | 10 | 13536512 | T | A | 0.578 | 0.017 | 0.002 | 9.66e-19 |
| rs56278466 | 10 | 17875857 | G | T | 0.637 | -0.014 | 0.002 | 1.53e-10 |
| rs692594 | 10 | 18265893 | C | G | 0.490 | 0.012 | 0.002 | 8.23e-11 |
| rs9804265 | 10 | 25214050 | C | A | 0.367 | -0.040 | 0.002 | 2.79e-89 |
| rs2807742 | 10 | 28781367 | A | G | 0.771 | 0.033 | 0.002 | 1.16e-45 |
| rs1571956 | 10 | 30504708 | A | G | 0.646 | 0.014 | 0.002 | 1.43e-11 |
| rs72790862 | 10 | 44880260 | C | T | 0.307 | -0.021 | 0.002 | 6.71e-24 |
| rs10995477 | 10 | 65010672 | C | T | 0.474 | -0.030 | 0.002 | 2.86e-56 |
| rs3747869 | 10 | 73520632 | C | A | 0.901 | 0.033 | 0.003 | 1.62e-22 |
| rs2802372 | 10 | 81047575 | C | A | 0.435 | -0.014 | 0.002 | 7.44e-12 |
| rs1412445 | 10 | 91002804 | T | C | 0.338 | 0.021 | 0.002 | 2.64e-25 |
| rs1977289 | 10 | 96301907 | C | T | 0.479 | -0.018 | 0.002 | 2.14e-21 |
| rs10786325 | 10 | 99068738 | G | C | 0.596 | 0.042 | 0.002 | 6.06e-104 |
| rs4909932 | 11 | 10475967 | G | A | 0.598 | 0.013 | 0.002 | 8.36e-12 |
| rs7934719 | 11 | 108341864 | T | C | 0.413 | 0.018 | 0.002 | 2.82e-20 |
| rs61904448 | 11 | 113958121 | C | T | 0.284 | -0.015 | 0.002 | 2.37e-11 |
| rs73000965 | 11 | 113982321 | A | T | 0.316 | 0.029 | 0.002 | 6.63e-45 |
| rs8705 | 11 | 128328913 | A | G | 0.317 | -0.018 | 0.002 | 1.97e-18 |
| rs4757955 | 11 | 12876492 | A | C | 0.680 | -0.015 | 0.002 | 5.28e-13 |
| rs1840595 | 11 | 18162482 | T | C | 0.230 | 0.015 | 0.002 | 1.29e-10 |
| rs10833024 | 11 | 3010390 | T | C | 0.310 | 0.016 | 0.002 | 4.51e-15 |
| rs7929589 | 11 | 59975078 | T | C | 0.407 | -0.015 | 0.002 | 1.79e-14 |
| rs7947929 | 11 | 65625191 | A | G | 0.666 | 0.016 | 0.002 | 1.38e-14 |
| rs2282611 | 11 | 76154846 | G | T | 0.320 | -0.013 | 0.002 | 1.03e-09 |
| rs7131178 | 11 | 93462607 | T | A | 0.812 | 0.014 | 0.002 | 4.79e-09 |
| rs28588142 | 12 | 10097609 | T | C | 0.188 | 0.015 | 0.002 | 6.6e-10 |
| rs3184504 | 12 | 111884608 | C | T | 0.517 | -0.029 | 0.002 | 1.9e-52 |
| rs11064881 | 12 | 120146925 | A | G | 0.073 | -0.034 | 0.004 | 2.37e-20 |
| rs610578 | 12 | 121194565 | G | A | 0.663 | -0.015 | 0.002 | 6.27e-13 |
| rs1168669 | 12 | 122228694 | C | T | 0.814 | -0.015 | 0.002 | 3.87e-09 |
| rs632887 | 12 | 3392351 | G | A | 0.411 | 0.014 | 0.002 | 1.06e-12 |
| rs10849020 | 12 | 4332009 | G | C | 0.210 | -0.024 | 0.002 | 2.34e-24 |
| rs12426444 | 12 | 50686512 | A | G | 0.358 | 0.018 | 0.002 | 6.66e-19 |
| rs1700159 | 12 | 52305786 | T | C | 0.774 | 0.022 | 0.002 | 1.92e-20 |
| rs1245035 | 12 | 64976049 | A | C | 0.628 | 0.016 | 0.002 | 3.39e-16 |
| rs1042725 | 12 | 66358347 | T | C | 0.494 | 0.013 | 0.002 | 3.33e-11 |
| rs4761234 | 12 | 69732105 | C | T | 0.484 | 0.022 | 0.002 | 4.25e-29 |
| rs11104881 | 12 | 88843474 | C | T | 0.702 | -0.026 | 0.002 | 7.84e-35 |
| rs2260766 | 13 | 114186800 | G | A | 0.282 | 0.023 | 0.002 | 5.22e-27 |
| rs76603681 | 13 | 28496468 | A | G | 0.032 | 0.041 | 0.006 | 2.65e-13 |
| rs76428106 | 13 | 28604007 | C | T | 0.013 | 0.104 | 0.009 | 6.25e-32 |
| rs78738581 | 13 | 42843630 | A | G | 0.232 | 0.021 | 0.002 | 1.13e-20 |
| rs7325851 | 13 | 50113215 | G | A | 0.741 | 0.016 | 0.002 | 6.33e-13 |
| rs2296028 | 13 | 52345637 | C | G | 0.175 | -0.015 | 0.003 | 2.16e-09 |
| rs9543219 | 13 | 73638478 | T | C | 0.574 | 0.013 | 0.002 | 5.06e-11 |
| rs12588718 | 14 | 101149605 | C | G | 0.666 | 0.015 | 0.002 | 4.29e-12 |
| rs2494748 | 14 | 105258892 | T | C | 0.616 | 0.013 | 0.002 | 1.5e-10 |
| rs11625865 | 14 | 105644421 | A | G | 0.610 | -0.015 | 0.002 | 1.26e-13 |
| rs2038700 | 14 | 25461989 | C | T | 0.394 | 0.033 | 0.002 | 1.25e-64 |
| rs72664840 | 14 | 35596323 | T | C | 0.179 | 0.021 | 0.003 | 9.21e-17 |
| rs72731564 | 14 | 69280158 | T | C | 0.187 | 0.020 | 0.002 | 2.95e-16 |
| rs34765661 | 14 | 69852940 | C | T | 0.065 | 0.042 | 0.004 | 8.58e-27 |
| rs4903580 | 14 | 77850978 | T | C | 0.456 | 0.017 | 0.002 | 1.16e-17 |
| rs10498635 | 14 | 93103309 | T | C | 0.184 | -0.030 | 0.002 | 8.73e-34 |
| rs67175901 | 15 | 101748227 | T | C | 0.107 | 0.044 | 0.003 | 1.99e-43 |
| rs4924450 | 15 | 40597229 | A | G | 0.705 | 0.014 | 0.002 | 3.4e-10 |
| rs72726027 | 15 | 42248826 | C | T | 0.112 | -0.043 | 0.003 | 1.71e-45 |
| rs2663553 | 15 | 51207818 | C | G | 0.541 | 0.021 | 0.002 | 4.92e-27 |
| rs780142 | 15 | 62797964 | G | T | 0.274 | 0.013 | 0.002 | 3.84e-09 |
| rs2062250 | 15 | 64672002 | A | G | 0.939 | 0.047 | 0.004 | 9.62e-31 |
| rs7496362 | 15 | 65758874 | G | C | 0.366 | 0.018 | 0.002 | 4.26e-18 |
| rs11072516 | 15 | 75224810 | A | T | 0.448 | 0.015 | 0.002 | 6.36e-15 |
| rs4886830 | 15 | 77094418 | T | A | 0.490 | 0.012 | 0.002 | 1.61e-09 |
| rs4843073 | 15 | 86124419 | T | C | 0.645 | 0.014 | 0.002 | 1.01e-11 |
| rs7183988 | 15 | 91428589 | G | T | 0.527 | -0.017 | 0.002 | 6.26e-18 |
| rs16958642 | 16 | 11864032 | A | G | 0.083 | 0.022 | 0.004 | 2.77e-10 |
| rs4984803 | 16 | 1349929 | A | G | 0.596 | -0.018 | 0.002 | 1.34e-17 |
| rs35929659 | 16 | 2165630 | C | T | 0.180 | -0.020 | 0.003 | 1.21e-13 |
| rs61739285 | 16 | 27480797 | T | C | 0.033 | -0.040 | 0.005 | 1.15e-13 |
| rs11574938 | 16 | 30485393 | C | G | 0.520 | 0.020 | 0.002 | 1.57e-25 |
| rs6500550 | 16 | 3746241 | T | C | 0.304 | -0.022 | 0.002 | 5.53e-26 |
| rs1362623 | 16 | 49885785 | T | C | 0.208 | 0.018 | 0.002 | 1.49e-14 |
| rs11648664 | 16 | 53122486 | A | G | 0.402 | -0.014 | 0.002 | 5.43e-13 |
| rs75974417 | 16 | 57063494 | T | C | 0.211 | -0.017 | 0.002 | 2.32e-12 |
| rs12927351 | 16 | 74596618 | A | C | 0.199 | 0.022 | 0.002 | 2.83e-19 |
| rs12930850 | 16 | 81602212 | G | A | 0.537 | 0.019 | 0.002 | 1.29e-22 |
| rs72832055 | 16 | 81869387 | A | G | 0.313 | -0.012 | 0.002 | 3.09e-09 |
| rs305082 | 16 | 85936978 | C | T | 0.172 | 0.037 | 0.003 | 3.04e-45 |
| rs72803323 | 16 | 89373324 | T | C | 0.040 | 0.034 | 0.005 | 3.56e-12 |
| rs9905106 | 17 | 1373518 | C | T | 0.734 | 0.023 | 0.002 | 1.62e-24 |
| rs7210990 | 17 | 16170764 | A | C | 0.478 | -0.037 | 0.002 | 9.18e-83 |
| rs16961474 | 17 | 17146619 | A | G | 0.137 | -0.017 | 0.003 | 1.11e-09 |
| rs7225843 | 17 | 2001825 | C | T | 0.203 | -0.022 | 0.002 | 2.69e-20 |
| rs4795474 | 17 | 27228198 | A | G | 0.124 | 0.019 | 0.003 | 3.23e-11 |
| rs7214252 | 17 | 27486673 | A | G | 0.211 | 0.015 | 0.002 | 2.11e-10 |
| rs3115086 | 17 | 28025949 | T | C | 0.503 | 0.020 | 0.002 | 1.73e-24 |
| rs79730542 | 17 | 37298789 | C | T | 0.030 | -0.043 | 0.006 | 3.32e-13 |
| rs12946510 | 17 | 37912377 | T | C | 0.473 | -0.059 | 0.002 | 1e-200 |
| rs74725931 | 17 | 38196327 | C | T | 0.037 | 0.067 | 0.005 | 9.03e-37 |
| rs111975659 | 17 | 38570168 | T | C | 0.027 | -0.041 | 0.006 | 1.96e-11 |
| rs11656151 | 17 | 44068492 | G | A | 0.238 | -0.016 | 0.002 | 1.08e-11 |
| rs78813154 | 17 | 5267002 | T | G | 0.144 | 0.019 | 0.003 | 7.13e-12 |
| rs56378716 | 17 | 56356502 | G | A | 0.013 | 0.129 | 0.009 | 3.72e-50 |
| rs2665405 | 17 | 57875292 | A | G | 0.548 | 0.030 | 0.002 | 6.74e-53 |
| rs749780 | 17 | 72699384 | A | C | 0.732 | 0.019 | 0.002 | 4.94e-18 |
| rs2001613 | 17 | 75383679 | T | C | 0.472 | 0.012 | 0.002 | 2.12e-09 |
| rs72901753 | 17 | 76244283 | C | G | 0.327 | -0.015 | 0.002 | 1.23e-11 |
| rs7235882 | 18 | 19684119 | T | C | 0.093 | 0.023 | 0.003 | 2.4e-12 |
| rs303753 | 18 | 21074922 | A | G | 0.345 | -0.023 | 0.002 | 9.57e-29 |
| rs78285907 | 18 | 21622003 | T | A | 0.089 | 0.022 | 0.003 | 3.11e-10 |
| rs4468717 | 18 | 3457606 | T | C | 0.078 | -0.025 | 0.004 | 4.59e-12 |
| rs9965539 | 18 | 41973779 | A | G | 0.134 | 0.031 | 0.003 | 1.35e-27 |
| rs8093990 | 18 | 42122473 | C | A | 0.430 | 0.012 | 0.002 | 1.63e-10 |
| rs17742008 | 18 | 48124763 | A | C | 0.212 | -0.018 | 0.002 | 4.1e-14 |
| rs656306 | 18 | 51705700 | A | C | 0.359 | 0.015 | 0.002 | 8.72e-14 |
| rs763362 | 18 | 67531797 | G | A | 0.399 | 0.016 | 0.002 | 2.23e-15 |
| rs72973711 | 18 | 74072245 | T | A | 0.067 | -0.036 | 0.004 | 5.53e-21 |
| rs10409243 | 19 | 10332988 | T | C | 0.601 | -0.022 | 0.002 | 5.55e-26 |
| rs930232 | 19 | 1036018 | A | G | 0.460 | 0.021 | 0.002 | 2.91e-25 |
| rs4808683 | 19 | 17862925 | G | C | 0.458 | -0.012 | 0.002 | 3.67e-10 |
| rs1985157 | 19 | 18513594 | C | T | 0.410 | 0.026 | 0.002 | 1.29e-38 |
| rs56408111 | 19 | 19793545 | C | T | 0.084 | 0.031 | 0.004 | 1.11e-18 |
| rs4805881 | 19 | 33896432 | C | A | 0.665 | -0.014 | 0.002 | 3.05e-12 |
| rs4760 | 19 | 44153100 | G | A | 0.153 | -0.075 | 0.003 | 1.42e-171 |
| rs11673093 | 19 | 45742094 | A | G | 0.260 | -0.049 | 0.002 | 1.45e-109 |
| rs386243 | 19 | 49127490 | T | C | 0.250 | -0.026 | 0.002 | 1.03e-30 |
| rs28540102 | 19 | 4975763 | C | T | 0.659 | 0.017 | 0.002 | 8.51e-17 |
| rs571497 | 19 | 7827830 | A | G | 0.155 | -0.038 | 0.003 | 9.76e-45 |
| rs284317 | 1 | 10731625 | G | A | 0.496 | 0.014 | 0.002 | 1.6e-11 |
| rs694180 | 1 | 111726213 | G | A | 0.656 | 0.019 | 0.002 | 1.24e-20 |
| rs2476601 | 1 | 114377568 | G | A | 0.900 | 0.038 | 0.003 | 5.82e-33 |
| rs7555995 | 1 | 150212707 | G | C | 0.285 | -0.015 | 0.002 | 2.86e-12 |
| rs114427170 | 1 | 159240076 | A | G | 0.037 | 0.032 | 0.005 | 2.81e-09 |
| rs2312675 | 1 | 166922533 | C | T | 0.212 | -0.020 | 0.002 | 2.61e-17 |
| rs60124939 | 1 | 174075302 | T | C | 0.176 | -0.018 | 0.003 | 1.52e-12 |
| rs41272536 | 1 | 183440531 | G | A | 0.046 | -0.036 | 0.005 | 2.33e-13 |
| rs16843350 | 1 | 198543095 | T | A | 0.030 | -0.034 | 0.006 | 2.85e-09 |
| rs2808519 | 1 | 200361914 | T | G | 0.738 | 0.016 | 0.002 | 1.5e-13 |
| rs4844622 | 1 | 208034329 | T | C | 0.240 | -0.023 | 0.002 | 1.44e-25 |
| rs35915186 | 1 | 224654623 | C | T | 0.215 | 0.024 | 0.002 | 3.67e-25 |
| rs11580229 | 1 | 227210820 | A | G | 0.462 | -0.023 | 0.002 | 2.76e-30 |
| rs533483 | 1 | 234765256 | A | G | 0.244 | -0.017 | 0.002 | 6.13e-14 |
| rs4626924 | 1 | 234909298 | T | C | 0.551 | 0.019 | 0.002 | 4.99e-23 |
| rs1886654 | 1 | 236105910 | C | T | 0.892 | -0.058 | 0.003 | 3.08e-80 |
| rs35020138 | 1 | 247570221 | C | T | 0.675 | -0.018 | 0.002 | 1.42e-17 |
| rs56188865 | 1 | 247606276 | C | T | 0.374 | -0.027 | 0.002 | 1.93e-43 |
| rs3762297 | 1 | 31231680 | T | C | 0.183 | 0.024 | 0.003 | 2.83e-21 |
| rs3917932 | 1 | 36943916 | G | C | 0.577 | -0.052 | 0.002 | 3e-154 |
| rs3754224 | 1 | 43423622 | C | T | 0.270 | -0.017 | 0.002 | 2.7e-14 |
| rs74076327 | 1 | 56979431 | C | T | 0.045 | -0.039 | 0.005 | 7.85e-17 |
| rs6678033 | 1 | 66077624 | A | G | 0.368 | -0.043 | 0.002 | 1.46e-102 |
| rs41313381 | 1 | 79411968 | A | C | 0.031 | 0.059 | 0.005 | 1.16e-26 |
| rs301819 | 1 | 8501786 | G | A | 0.584 | -0.014 | 0.002 | 6.96e-13 |
| rs10864368 | 1 | 8918313 | C | T | 0.503 | 0.024 | 0.002 | 4.01e-35 |
| rs150649461 | 1 | 92925654 | C | G | 0.015 | 0.053 | 0.008 | 1.9e-10 |
| rs116631966 | 1 | 93229909 | A | G | 0.025 | 0.040 | 0.006 | 1.02e-10 |
| rs2254458 | 20 | 1551485 | T | C | 0.645 | -0.013 | 0.002 | 1.76e-10 |
| rs156338 | 20 | 1854678 | T | C | 0.436 | -0.012 | 0.002 | 3.32e-09 |
| rs633284 | 20 | 2904143 | T | A | 0.516 | 0.015 | 0.002 | 1.74e-13 |
| rs4812447 | 20 | 39272620 | G | A | 0.440 | 0.017 | 0.002 | 1.49e-18 |
| rs1800961 | 20 | 43042364 | T | C | 0.031 | -0.052 | 0.006 | 8.99e-21 |
| rs1555275 | 20 | 55969270 | G | A | 0.650 | -0.014 | 0.002 | 2.3e-11 |
| rs2315008 | 20 | 62343956 | G | T | 0.667 | 0.016 | 0.002 | 1.64e-15 |
| rs12481262 | 20 | 8126920 | C | T | 0.460 | -0.015 | 0.002 | 4.79e-13 |
| rs9977672 | 21 | 40463283 | A | G | 0.259 | -0.022 | 0.002 | 7.34e-24 |
| rs1788493 | 21 | 44469063 | T | C | 0.500 | 0.013 | 0.002 | 1.88e-11 |
| rs2839166 | 21 | 47656712 | C | T | 0.398 | -0.013 | 0.002 | 1.91e-11 |
| rs41433144 | 22 | 17569993 | A | T | 0.043 | -0.035 | 0.005 | 1.45e-12 |
| rs5747308 | 22 | 18133500 | C | A | 0.504 | 0.022 | 0.002 | 2.3e-30 |
| rs5746638 | 22 | 18907997 | T | C | 0.286 | -0.014 | 0.002 | 9.41e-10 |
| rs47341 | 22 | 43560763 | T | C | 0.396 | 0.013 | 0.002 | 2.26e-10 |
| rs738408 | 22 | 44324730 | T | C | 0.217 | -0.016 | 0.002 | 7.74e-12 |
| rs35789178 | 2 | 102603925 | G | T | 0.186 | 0.018 | 0.002 | 3.41e-13 |
| rs11691193 | 2 | 111854524 | T | C | 0.218 | 0.015 | 0.002 | 2.52e-10 |
| rs75726191 | 2 | 112914684 | T | C | 0.135 | 0.023 | 0.003 | 1.29e-15 |
| rs6734238 | 2 | 113841030 | G | A | 0.403 | 0.039 | 0.002 | 1.58e-90 |
| rs11683933 | 2 | 127886955 | A | C | 0.055 | 0.031 | 0.004 | 5.72e-13 |
| rs6432335 | 2 | 12928971 | G | T | 0.452 | 0.012 | 0.002 | 2.42e-10 |
| rs354703 | 2 | 143883952 | C | T | 0.593 | 0.015 | 0.002 | 4.08e-14 |
| rs3856364 | 2 | 145477217 | G | C | 0.680 | 0.014 | 0.002 | 5.28e-12 |
| rs188653407 | 2 | 160671563 | G | C | 0.023 | -0.051 | 0.006 | 2.58e-15 |
| rs2729707 | 2 | 160687231 | G | A | 0.831 | -0.036 | 0.003 | 2.68e-44 |
| rs11689257 | 2 | 162958353 | A | G | 0.070 | -0.025 | 0.004 | 2.51e-10 |
| rs4632345 | 2 | 16702654 | A | G | 0.678 | -0.012 | 0.002 | 3.24e-09 |
| rs2632372 | 2 | 169717541 | C | T | 0.504 | -0.021 | 0.002 | 6.68e-28 |
| rs62189859 | 2 | 182254087 | C | T | 0.244 | 0.014 | 0.002 | 2.96e-10 |
| rs6740847 | 2 | 182308352 | G | A | 0.562 | 0.022 | 0.002 | 3.95e-29 |
| rs13392977 | 2 | 192514856 | A | G | 0.054 | 0.028 | 0.004 | 3.73e-11 |
| rs1047891 | 2 | 211540507 | A | C | 0.315 | -0.019 | 0.002 | 8.79e-21 |
| rs114050631 | 2 | 219020958 | T | C | 0.011 | -0.163 | 0.010 | 8.74e-59 |
| rs11695689 | 2 | 220018091 | C | A | 0.640 | 0.012 | 0.002 | 2.88e-09 |
| rs79047930 | 2 | 220165527 | T | C | 0.015 | 0.053 | 0.008 | 1.64e-10 |
| rs55729107 | 2 | 232582085 | G | A | 0.226 | -0.018 | 0.002 | 7.87e-14 |
| rs1057258 | 2 | 234115629 | T | C | 0.178 | 0.017 | 0.003 | 2.83e-10 |
| rs4074882 | 2 | 237780727 | C | T | 0.742 | 0.028 | 0.002 | 5.61e-37 |
| rs36101491 | 2 | 24387532 | T | C | 0.294 | 0.021 | 0.002 | 1.41e-23 |
| rs1260326 | 2 | 27730940 | C | T | 0.602 | -0.033 | 0.002 | 6.91e-62 |
| rs116447416 | 2 | 37501547 | G | A | 0.035 | -0.037 | 0.005 | 1.47e-12 |
| rs17030394 | 2 | 43354280 | G | A | 0.386 | -0.016 | 0.002 | 6.29e-16 |
| rs77552263 | 2 | 43786818 | A | G | 0.078 | 0.049 | 0.004 | 7.19e-42 |
| rs75475627 | 2 | 54787592 | G | C | 0.077 | 0.033 | 0.004 | 3.76e-19 |
| rs2421200 | 2 | 61711815 | T | G | 0.487 | -0.021 | 0.002 | 1.03e-26 |
| rs6731993 | 2 | 65642097 | T | A | 0.408 | 0.023 | 0.002 | 2.45e-31 |
| rs7639292 | 3 | 107295665 | T | C | 0.169 | -0.022 | 0.003 | 4.83e-18 |
| rs58605236 | 3 | 119697032 | A | T | 0.373 | -0.014 | 0.002 | 2.53e-12 |
| rs2734031 | 3 | 128301390 | C | T | 0.908 | -0.068 | 0.003 | 4.2e-91 |
| rs6764912 | 3 | 128372487 | A | G | 0.065 | -0.037 | 0.004 | 3.14e-20 |
| rs56217149 | 3 | 12962914 | A | G | 0.151 | 0.017 | 0.003 | 4.45e-10 |
| rs11712552 | 3 | 140957762 | G | A | 0.126 | 0.030 | 0.003 | 6.45e-26 |
| rs9819371 | 3 | 141206800 | T | C | 0.065 | -0.043 | 0.004 | 3.64e-28 |
| rs2046934 | 3 | 151057642 | A | G | 0.823 | 0.015 | 0.002 | 1.66e-09 |
| rs56082403 | 3 | 156797225 | C | T | 0.404 | 0.015 | 0.002 | 5.08e-15 |
| rs11716015 | 3 | 169320058 | T | C | 0.636 | -0.013 | 0.002 | 1.16e-10 |
| rs3749440 | 3 | 183702089 | G | A | 0.372 | 0.017 | 0.002 | 2.2e-18 |
| rs56174170 | 3 | 184091881 | C | T | 0.139 | 0.017 | 0.003 | 2.04e-09 |
| rs34834004 | 3 | 185991986 | G | T | 0.506 | -0.013 | 0.002 | 6.77e-11 |
| rs789858 | 3 | 194405966 | T | C | 0.404 | 0.014 | 0.002 | 1.07e-12 |
| rs7626444 | 3 | 196504902 | C | G | 0.422 | -0.021 | 0.002 | 1.27e-26 |
| rs1366045 | 3 | 42909050 | C | T | 0.385 | -0.022 | 0.002 | 1.2e-28 |
| rs6442061 | 3 | 47203783 | T | G | 0.596 | 0.017 | 0.002 | 3.68e-17 |
| rs6779340 | 3 | 58033701 | G | C | 0.337 | -0.015 | 0.002 | 8.69e-14 |
| rs12487658 | 3 | 71528943 | T | C | 0.802 | 0.017 | 0.002 | 2.06e-12 |
| rs11721064 | 3 | 98408826 | T | G | 0.609 | 0.014 | 0.002 | 3.22e-13 |
| rs17213043 | 4 | 105546979 | G | C | 0.028 | 0.046 | 0.006 | 1.97e-15 |
| rs7679673 | 4 | 106061534 | A | C | 0.380 | -0.024 | 0.002 | 2.15e-33 |
| rs4145952 | 4 | 120155806 | A | C | 0.399 | -0.013 | 0.002 | 1.06e-11 |
| rs11735662 | 4 | 145026126 | T | C | 0.034 | 0.048 | 0.005 | 5.59e-19 |
| rs2290846 | 4 | 151199080 | A | G | 0.284 | 0.019 | 0.002 | 9.92e-19 |
| rs6817881 | 4 | 152223904 | T | C | 0.499 | 0.014 | 0.002 | 1.35e-13 |
| rs6855981 | 4 | 3148276 | A | G | 0.373 | -0.016 | 0.002 | 1.85e-16 |
| rs28530750 | 4 | 36312542 | A | G | 0.043 | 0.056 | 0.005 | 2.36e-31 |
| rs218264 | 4 | 55408875 | T | A | 0.251 | 0.032 | 0.002 | 5.59e-46 |
| rs723585 | 4 | 55503194 | G | A | 0.484 | -0.022 | 0.002 | 2.97e-31 |
| rs7684253 | 4 | 57727311 | T | C | 0.550 | 0.014 | 0.002 | 3.29e-13 |
| rs6831368 | 4 | 6969919 | G | A | 0.362 | -0.022 | 0.002 | 3.65e-27 |
| rs35734242 | 4 | 706700 | C | T | 0.428 | 0.023 | 0.002 | 5.42e-31 |
| rs11723621 | 4 | 72615362 | G | A | 0.291 | -0.027 | 0.002 | 2.67e-37 |
| rs16850073 | 4 | 74703999 | T | C | 0.374 | 0.052 | 0.002 | 2.12e-149 |
| rs370655 | 4 | 74903134 | C | T | 0.394 | 0.062 | 0.002 | 1e-200 |
| rs1445171 | 5 | 100046075 | C | T | 0.517 | 0.014 | 0.002 | 1.15e-13 |
| rs4535497 | 5 | 1107428 | A | C | 0.570 | -0.014 | 0.002 | 1.75e-12 |
| rs257063 | 5 | 114806819 | T | C | 0.741 | -0.015 | 0.002 | 1.54e-11 |
| rs1966479 | 5 | 118627271 | G | A | 0.660 | 0.014 | 0.002 | 1.21e-12 |
| rs6878780 | 5 | 122093740 | C | T | 0.426 | 0.013 | 0.002 | 6.01e-12 |
| rs7705526 | 5 | 1285974 | A | C | 0.327 | 0.034 | 0.002 | 9.19e-54 |
| rs2522051 | 5 | 131797578 | C | T | 0.454 | 0.024 | 0.002 | 1.88e-35 |
| rs6860961 | 5 | 133849687 | C | T | 0.583 | 0.012 | 0.002 | 2.3e-10 |
| rs6881942 | 5 | 141518653 | T | C | 0.556 | 0.021 | 0.002 | 2.76e-28 |
| rs114363252 | 5 | 142539162 | A | G | 0.074 | 0.023 | 0.004 | 9.53e-10 |
| rs284440 | 5 | 156170215 | G | T | 0.838 | -0.020 | 0.003 | 6.16e-15 |
| rs2561758 | 5 | 173205282 | G | A | 0.723 | -0.034 | 0.002 | 3.49e-55 |
| rs13190036 | 5 | 176737720 | G | A | 0.891 | -0.022 | 0.003 | 3.63e-12 |
| rs10447304 | 5 | 179172098 | A | G | 0.506 | -0.017 | 0.002 | 5.58e-18 |
| rs152232 | 5 | 179353282 | C | T | 0.384 | 0.014 | 0.002 | 4.45e-12 |
| rs62360185 | 5 | 57274612 | G | A | 0.191 | -0.016 | 0.002 | 8.05e-11 |
| rs12658947 | 5 | 57552464 | G | A | 0.257 | 0.013 | 0.002 | 3.01e-09 |
| rs6859727 | 5 | 71742622 | C | T | 0.878 | -0.040 | 0.003 | 4.29e-43 |
| rs2432142 | 5 | 96275201 | A | G | 0.435 | 0.015 | 0.002 | 1.89e-14 |
| rs6927569 | 6 | 109621494 | C | T | 0.523 | 0.026 | 0.002 | 2.4e-41 |
| rs3777755 | 6 | 12159699 | T | C | 0.311 | 0.014 | 0.002 | 4.78e-11 |
| rs62429983 | 6 | 121862273 | A | G | 0.145 | 0.016 | 0.003 | 1.77e-09 |
| rs1490384 | 6 | 126851160 | T | C | 0.501 | -0.015 | 0.002 | 2.19e-14 |
| rs9402685 | 6 | 135419688 | C | T | 0.256 | -0.034 | 0.002 | 1.05e-52 |
| rs12214269 | 6 | 135846518 | A | G | 0.545 | 0.019 | 0.002 | 6.6e-24 |
| rs6924387 | 6 | 137082948 | G | A | 0.412 | 0.017 | 0.002 | 1.73e-18 |
| rs72978754 | 6 | 138053203 | C | T | 0.066 | -0.026 | 0.004 | 5.38e-11 |
| rs9390461 | 6 | 147701217 | G | A | 0.539 | 0.018 | 0.002 | 3.49e-20 |
| rs10945542 | 6 | 158752931 | T | C | 0.521 | 0.012 | 0.002 | 4.82e-10 |
| rs212409 | 6 | 159470058 | A | G | 0.554 | -0.021 | 0.002 | 4.71e-28 |
| rs6915310 | 6 | 16758002 | T | C | 0.174 | -0.032 | 0.003 | 1.71e-36 |
| rs4712614 | 6 | 21382765 | G | T | 0.620 | -0.020 | 0.002 | 1.04e-23 |
| rs4712850 | 6 | 24805574 | T | G | 0.508 | 0.014 | 0.002 | 3.35e-13 |
| rs72843538 | 6 | 25808742 | A | T | 0.126 | 0.019 | 0.003 | 1.41e-10 |
| rs3828914 | 6 | 31465819 | T | C | 0.305 | 0.043 | 0.002 | 1.04e-94 |
| rs12194148 | 6 | 32444198 | T | G | 0.469 | 0.059 | 0.002 | 1e-200 |
| rs2395618 | 6 | 35299507 | T | C | 0.686 | 0.016 | 0.002 | 8.98e-15 |
| rs16895923 | 6 | 42633368 | G | A | 0.166 | 0.025 | 0.003 | 1.62e-21 |
| rs6905891 | 6 | 43759189 | A | G | 0.081 | -0.028 | 0.004 | 3.26e-15 |
| rs1089946 | 6 | 52310094 | A | T | 0.614 | 0.013 | 0.002 | 8.29e-11 |
| rs6936191 | 6 | 7201660 | C | T | 0.608 | 0.018 | 0.002 | 9.87e-20 |
| rs915125 | 6 | 82463376 | T | C | 0.282 | -0.018 | 0.002 | 1.92e-16 |
| rs6454596 | 6 | 87980735 | C | G | 0.490 | -0.025 | 0.002 | 1.38e-39 |
| rs7753008 | 6 | 90809639 | C | T | 0.387 | -0.013 | 0.002 | 1.08e-10 |
| rs342242 | 7 | 106338989 | C | T | 0.453 | -0.016 | 0.002 | 3.62e-16 |
| rs35759345 | 7 | 116388021 | T | C | 0.450 | 0.013 | 0.002 | 6.19e-11 |
| rs9656395 | 7 | 130575884 | G | A | 0.095 | -0.023 | 0.003 | 4.08e-12 |
| rs7803075 | 7 | 130742066 | G | A | 0.735 | -0.019 | 0.002 | 2.19e-18 |
| rs13231262 | 7 | 148886058 | T | C | 0.656 | -0.020 | 0.002 | 1.34e-23 |
| rs12530608 | 7 | 18023617 | G | A | 0.146 | -0.016 | 0.003 | 3.63e-09 |
| rs7776857 | 7 | 22754768 | T | G | 0.657 | -0.015 | 0.002 | 3.16e-13 |
| rs798547 | 7 | 2761177 | C | T | 0.296 | 0.014 | 0.002 | 1.46e-10 |
| rs2158799 | 7 | 28277107 | G | C | 0.611 | 0.052 | 0.002 | 6.53e-152 |
| rs56388170 | 7 | 28724374 | T | G | 0.294 | 0.070 | 0.002 | 1e-200 |
| rs2710804 | 7 | 36084529 | C | T | 0.376 | 0.020 | 0.002 | 3.76e-23 |
| rs3735485 | 7 | 45009341 | G | A | 0.845 | 0.029 | 0.003 | 4.2e-28 |
| rs10252457 | 7 | 47337530 | G | A | 0.426 | -0.014 | 0.002 | 1.82e-12 |
| rs7341532 | 7 | 48135306 | T | G | 0.522 | 0.011 | 0.002 | 3.46e-09 |
| rs876036 | 7 | 50307710 | C | T | 0.321 | 0.021 | 0.002 | 6.88e-25 |
| rs778732 | 7 | 65822360 | T | C | 0.107 | 0.027 | 0.003 | 6.53e-18 |
| rs62465399 | 7 | 65941570 | T | C | 0.045 | -0.033 | 0.005 | 1.66e-12 |
| rs1474419 | 7 | 6692605 | C | T | 0.579 | 0.018 | 0.002 | 2.55e-19 |
| rs33951980 | 7 | 73029437 | T | C | 0.129 | -0.028 | 0.003 | 6.63e-22 |
| rs73049276 | 7 | 8022016 | A | G | 0.059 | -0.040 | 0.004 | 5.13e-22 |
| rs42033 | 7 | 92237533 | T | A | 0.209 | 0.038 | 0.002 | 5.51e-59 |
| rs445 | 7 | 92408370 | T | C | 0.098 | -0.091 | 0.003 | 1.36e-175 |
| rs7790229 | 7 | 97919338 | A | G | 0.493 | -0.014 | 0.002 | 1.45e-12 |
| rs4734879 | 8 | 106583124 | G | A | 0.276 | -0.021 | 0.002 | 2.63e-23 |
| rs1991651 | 8 | 10706411 | G | C | 0.618 | 0.023 | 0.002 | 7.4e-31 |
| rs2954038 | 8 | 126507389 | A | C | 0.697 | 0.021 | 0.002 | 2.64e-23 |
| rs145209947 | 8 | 130694185 | A | C | 0.014 | -0.069 | 0.009 | 1.96e-14 |
| rs7005996 | 8 | 142241681 | T | C | 0.906 | 0.023 | 0.003 | 3.33e-11 |
| rs6985508 | 8 | 142337734 | A | G | 0.358 | -0.027 | 0.002 | 3.36e-39 |
| rs34215892 | 8 | 21767240 | A | G | 0.028 | 0.038 | 0.006 | 2.3e-10 |
| rs12550612 | 8 | 22966769 | A | G | 0.822 | -0.031 | 0.003 | 1.09e-33 |
| rs6468341 | 8 | 30279355 | C | T | 0.743 | 0.019 | 0.002 | 7.1e-18 |
| rs45577137 | 8 | 48651633 | G | A | 0.045 | 0.036 | 0.005 | 2.63e-12 |
| rs28571765 | 8 | 55449281 | C | T | 0.206 | -0.022 | 0.002 | 3.8e-21 |
| rs7816785 | 8 | 56797418 | T | C | 0.606 | -0.021 | 0.002 | 4.31e-27 |
| rs609264 | 8 | 61548546 | G | T | 0.528 | 0.013 | 0.002 | 7.93e-11 |
| rs7846314 | 8 | 61650831 | T | A | 0.187 | 0.064 | 0.002 | 4.7e-149 |
| rs4276676 | 8 | 68795618 | C | T | 0.470 | -0.018 | 0.002 | 1.16e-20 |
| rs12716647 | 8 | 6901304 | C | G | 0.637 | 0.014 | 0.002 | 1.03e-12 |
| rs16939607 | 8 | 79013333 | A | G | 0.146 | -0.028 | 0.003 | 1.99e-25 |
| rs7815046 | 8 | 87054885 | C | A | 0.303 | -0.013 | 0.002 | 4.65e-10 |
| rs6998846 | 8 | 9193341 | A | G | 0.783 | 0.015 | 0.002 | 4.54e-11 |
| rs117468663 | 9 | 112745175 | T | A | 0.134 | 0.019 | 0.003 | 8.74e-12 |
| rs1411424 | 9 | 113892963 | A | G | 0.523 | -0.011 | 0.002 | 2.51e-09 |
| rs7866863 | 9 | 114657707 | A | G | 0.341 | 0.013 | 0.002 | 1e-10 |
| rs17831500 | 9 | 116047472 | C | A | 0.224 | -0.020 | 0.002 | 5.64e-18 |
| rs12378064 | 9 | 127191540 | T | C | 0.650 | -0.014 | 0.002 | 1.99e-11 |
| rs2519093 | 9 | 136141870 | T | C | 0.185 | -0.040 | 0.002 | 9.83e-59 |
| rs2157770 | 9 | 136921464 | G | A | 0.287 | 0.021 | 0.002 | 2.67e-21 |
| rs4413892 | 9 | 139330158 | A | G | 0.278 | 0.034 | 0.002 | 1.65e-51 |
| rs9411293 | 9 | 139929015 | G | C | 0.337 | -0.014 | 0.002 | 4.19e-11 |
| rs2219143 | 9 | 2622278 | A | G | 0.394 | 0.019 | 0.002 | 1.3e-20 |
| rs2992836 | 9 | 276053 | T | G | 0.186 | 0.028 | 0.002 | 6.65e-29 |
| rs3739873 | 9 | 34978431 | A | G | 0.220 | -0.014 | 0.002 | 3.45e-09 |
| rs3793537 | 9 | 35687556 | C | G | 0.291 | 0.013 | 0.002 | 2.55e-09 |
| rs385893 | 9 | 4763176 | C | T | 0.521 | 0.023 | 0.002 | 7.7e-34 |
| rs10758669 | 9 | 4981602 | A | C | 0.658 | -0.014 | 0.002 | 1.85e-12 |
| rs626416 | 9 | 79326680 | C | G | 0.762 | -0.020 | 0.002 | 5.77e-18 |
| rs796007 | 9 | 86577541 | A | G | 0.253 | -0.021 | 0.002 | 9.41e-21 |
| rs10992394 | 9 | 95433830 | A | G | 0.231 | 0.014 | 0.002 | 1.14e-09 |
| rs150861794 | 13 | 109003805 | C | T | 0.980 | 0.053 | 0.008 | 2.47e-12 |
| rs34599082 | 1 | 159175494 | C | T | 0.987 | 0.173 | 0.008 | 7.27e-94 |
| rs138645114 | 10 | 69586392 | G | T | 0.056 | -0.035 | 0.005 | 2.04e-14 |
| rs74149356 | 10 | 93785201 | T | A | 0.169 | 0.016 | 0.003 | 2.06e-09 |
| rs1715460 | 11 | 118122052 | G | C | 0.741 | 0.021 | 0.002 | 4.32e-19 |
| rs14408 | 11 | 308314 | C | T | 0.368 | 0.049 | 0.002 | 1.21e-115 |
| rs11823949 | 11 | 47723512 | A | G | 0.347 | 0.019 | 0.002 | 1.56e-20 |
| rs174557 | 11 | 61581368 | G | A | 0.307 | -0.025 | 0.002 | 3.19e-31 |
| rs35375443 | 11 | 9729432 | T | A | 0.285 | 0.015 | 0.002 | 9.14e-12 |
| rs370591186 | 12 | 123885930 | T | C | 0.026 | 0.048 | 0.006 | 1.89e-13 |
| rs549435091 | 12 | 27393975 | G | T | 0.067 | -0.028 | 0.004 | 6.2e-11 |
| rs149859245 | 14 | 81710883 | C | T | 0.782 | -0.019 | 0.002 | 1.41e-14 |
| rs576566496 | 17 | 38140269 | A | G | 0.026 | 0.092 | 0.006 | 9.87e-46 |
| rs79135050 | 17 | 38215203 | C | A | 0.030 | 0.080 | 0.006 | 2.29e-37 |
| rs113686433 | 17 | 56803943 | G | A | 0.135 | 0.022 | 0.003 | 8.3e-13 |
| rs55874505 | 18 | 43816804 | G | T | 0.584 | -0.018 | 0.002 | 4.06e-19 |
| rs12232734 | 18 | 74253960 | A | T | 0.186 | 0.016 | 0.003 | 9.43e-10 |
| rs150665764 | 19 | 14499614 | C | G | 0.017 | 0.047 | 0.008 | 3.75e-09 |
| rs61801809 | 1 | 161557512 | T | C | 0.355 | -0.013 | 0.002 | 1.79e-09 |
| rs140884047 | 1 | 161610372 | C | A | 0.848 | 0.027 | 0.003 | 1.28e-21 |
| rs6593925 | 1 | 205100663 | C | G | 0.908 | 0.024 | 0.003 | 9.24e-13 |
| rs12037562 | 1 | 248047350 | C | T | 0.660 | -0.013 | 0.002 | 2.56e-10 |
| rs11800162 | 1 | 36970512 | T | C | 0.020 | 0.064 | 0.007 | 5.07e-19 |
| rs140958972 | 1 | 46428213 | G | A | 0.435 | -0.020 | 0.002 | 1.45e-23 |
| rs12136896 | 1 | 56617956 | A | G | 0.358 | 0.015 | 0.002 | 8.71e-14 |
| rs62239920 | 21 | 18944889 | A | G | 0.203 | -0.016 | 0.003 | 1.56e-10 |
| rs143188161 | 22 | 24403541 | T | C | 0.522 | 0.013 | 0.002 | 3.59e-10 |
| rs35955747 | 22 | 31813058 | T | A | 0.539 | 0.019 | 0.002 | 1.34e-21 |
| rs2309996 | 2 | 101735838 | G | T | 0.625 | 0.023 | 0.002 | 4.27e-30 |
| rs4954387 | 2 | 136784344 | G | A | 0.248 | 0.014 | 0.002 | 3.67e-09 |
| rs2891550 | 2 | 27334624 | T | C | 0.554 | -0.013 | 0.002 | 1.91e-10 |
| rs4599108 | 2 | 85543222 | T | C | 0.488 | -0.019 | 0.002 | 6.58e-22 |
| rs3733120 | 3 | 63850299 | C | G | 0.788 | -0.015 | 0.002 | 1.63e-09 |
| rs547380175 | 4 | 26290470 | T | A | 0.043 | -0.030 | 0.005 | 2.55e-09 |
| rs10479004 | 5 | 131842977 | T | G | 0.512 | 0.014 | 0.002 | 7.07e-13 |
| rs11960649 | 5 | 148203144 | C | A | 0.553 | -0.031 | 0.002 | 1.96e-53 |
| rs66514959 | 5 | 68611583 | A | G | 0.444 | -0.026 | 0.002 | 2.48e-38 |
| rs9260150 | 6 | 29911029 | T | G | 0.092 | 0.032 | 0.004 | 1.73e-19 |
| rs9274305 | 6 | 32632000 | T | C | 0.192 | -0.038 | 0.003 | 2.66e-43 |
| rs62454712 | 7 | 6498057 | C | T | 0.239 | -0.018 | 0.002 | 2.64e-14 |
| rs10104995 | 8 | 103901234 | T | C | 0.231 | -0.023 | 0.002 | 3.36e-22 |
| rs59697075 | 8 | 130618150 | T | C | 0.587 | -0.038 | 0.002 | 6.4e-80 |
| rs6502497 | 17 | 16102530 | D | I | 0.464 | 0.033 | 0.002 | 2.67e-54 |
| rs534234053 | 6 | 32490048 | G | A | 0.074 | 0.053 | 0.006 | 5.15e-21 |

**Table S9. 421 SNPs significantly associated with monocyte count used IVs in forward MR analyses derived from Vuckovic D et al.** Chr: Chromosome; EA: Effect allele; NEA: Non-effect allele; EAF: Effect allele frequency.

| SNP | Chr | Pos | EA | NEA | EAF | Beta | SE | P-value |
| --- | --- | --- | --- | --- | --- | --- | --- | --- |
| rs4240895 | 1 | 9713386 | T | C | 0.388 | -0.023 | 0.002 | 2.8e-27 |
| rs620405 | 1 | 10554794 | C | T | 0.684 | 0.015 | 0.002 | 1.6e-11 |
| rs284324 | 1 | 10724236 | A | G | 0.494 | 0.022 | 0.002 | 8e-25 |
| rs11247913 | 1 | 26648551 | C | G | 0.762 | 0.028 | 0.002 | 5.8e-31 |
| rs141459612 | 1 | 28215835 | T | A | 0.011 | 0.077 | 0.010 | 4.1e-14 |
| rs35351292 | 1 | 31208042 | T | C | 0.267 | 0.015 | 0.002 | 6.7e-11 |
| rs77046911 | 1 | 41330726 | T | C | 0.112 | 0.029 | 0.003 | 1.5e-18 |
| rs80200046 | 1 | 43423932 | A | G | 0.172 | -0.027 | 0.003 | 1.2e-22 |
| rs11211176 | 1 | 46223086 | A | G | 0.532 | -0.016 | 0.002 | 1.8e-14 |
| rs10788942 | 1 | 53210521 | T | A | 0.470 | -0.017 | 0.002 | 5.2e-17 |
| rs12117663 | 1 | 54892079 | T | C | 0.432 | -0.017 | 0.002 | 5.5e-16 |
| rs72675573 | 1 | 56636881 | T | C | 0.367 | 0.017 | 0.002 | 2e-14 |
| rs1933295 | 1 | 62107021 | G | A | 0.776 | -0.024 | 0.003 | 1.1e-21 |
| rs72683129 | 1 | 66069781 | G | A | 0.148 | 0.017 | 0.003 | 4.7e-09 |
| rs12562761 | 1 | 67396739 | T | C | 0.650 | -0.018 | 0.002 | 1.2e-16 |
| rs41313381 | 1 | 79411968 | A | C | 0.031 | 0.042 | 0.006 | 1.6e-12 |
| rs528433 | 1 | 89987940 | G | A | 0.524 | -0.015 | 0.002 | 2.6e-13 |
| rs191524 | 1 | 91584916 | G | A | 0.658 | -0.014 | 0.002 | 5.6e-11 |
| rs11466597 | 1 | 92185351 | C | T | 0.012 | 0.291 | 0.010 | 1e-200 |
| rs564161800 | 1 | 92423062 | T | A | 0.035 | 0.084 | 0.007 | 2.2e-33 |
| rs115340020 | 1 | 92766438 | A | G | 0.028 | -0.097 | 0.006 | 7.7e-54 |
| rs12138486 | 1 | 93865391 | T | C | 0.781 | 0.018 | 0.003 | 4.4e-13 |
| rs28549287 | 1 | 110230138 | A | G | 0.774 | -0.016 | 0.003 | 1.2e-09 |
| rs333947 | 1 | 110470764 | A | G | 0.150 | 0.037 | 0.003 | 1.8e-36 |
| rs11102144 | 1 | 111195493 | G | A | 0.169 | -0.017 | 0.003 | 5.9e-10 |
| rs67224956 | 1 | 118154575 | C | T | 0.169 | 0.026 | 0.003 | 1.9e-21 |
| rs11204682 | 1 | 150595537 | T | G | 0.223 | -0.075 | 0.003 | 4.4e-195 |
| rs3806232 | 1 | 153364130 | C | T | 0.118 | 0.029 | 0.003 | 8.3e-20 |
| rs142676169 | 1 | 156243941 | T | C | 0.020 | -0.048 | 0.008 | 1.3e-09 |
| rs3026940 | 1 | 159130696 | T | A | 0.008 | -0.099 | 0.013 | 4.2e-14 |
| rs3027012 | 1 | 159174123 | T | C | 0.182 | 0.028 | 0.003 | 1e-24 |
| rs10908820 | 1 | 160999479 | G | A | 0.796 | 0.017 | 0.003 | 1.5e-10 |
| rs35902866 | 1 | 169544387 | T | C | 0.399 | -0.018 | 0.002 | 4e-18 |
| rs10489182 | 1 | 169710669 | G | A | 0.189 | 0.026 | 0.003 | 4e-22 |
| rs3795503 | 1 | 180905694 | T | C | 0.315 | -0.019 | 0.002 | 4.2e-18 |
| rs7364556 | 1 | 185401229 | C | T | 0.679 | 0.026 | 0.002 | 1.5e-30 |
| rs9787298 | 1 | 204276040 | C | A | 0.211 | -0.016 | 0.003 | 4.5e-10 |
| rs2018104 | 1 | 205309382 | G | A | 0.299 | 0.013 | 0.002 | 4.2e-09 |
| rs9438393 | 1 | 205782718 | G | A | 0.414 | 0.019 | 0.002 | 2.4e-19 |
| rs7522307 | 1 | 207998783 | C | G | 0.125 | -0.022 | 0.003 | 1.7e-12 |
| rs6702347 | 1 | 212612911 | A | C | 0.298 | 0.023 | 0.002 | 1.3e-24 |
| rs1414519 | 1 | 221134350 | A | G | 0.572 | 0.017 | 0.002 | 1.8e-15 |
| rs113292043 | 1 | 223231657 | T | C | 0.067 | -0.027 | 0.004 | 1.1e-10 |
| rs6696074 | 1 | 225962690 | T | C | 0.565 | 0.018 | 0.002 | 2.4e-16 |
| rs4626924 | 1 | 234909298 | T | C | 0.551 | -0.021 | 0.002 | 9.5e-23 |
| rs4269828 | 1 | 235257838 | G | A | 0.559 | 0.013 | 0.002 | 1.1e-09 |
| rs6429432 | 1 | 236107241 | C | A | 0.892 | -0.092 | 0.003 | 3.8e-167 |
| rs6429439 | 1 | 243993530 | T | A | 0.868 | 0.021 | 0.003 | 2.6e-11 |
| rs61165644 | 1 | 244487940 | G | A | 0.141 | -0.027 | 0.003 | 9.2e-19 |
| rs4335411 | 1 | 249191706 | A | G | 0.762 | -0.019 | 0.003 | 2.1e-13 |
| rs3111414 | 2 | 8443859 | G | C | 0.796 | 0.018 | 0.003 | 4.5e-12 |
| rs7572278 | 2 | 8563029 | A | T | 0.203 | 0.022 | 0.003 | 1.1e-17 |
| rs2059427 | 2 | 12893977 | T | C | 0.372 | 0.021 | 0.002 | 6.4e-22 |
| rs6753534 | 2 | 27752871 | T | C | 0.565 | 0.014 | 0.002 | 1e-10 |
| rs649729 | 2 | 31464385 | A | T | 0.697 | 0.019 | 0.002 | 1e-16 |
| rs28498283 | 2 | 43360065 | T | A | 0.256 | 0.022 | 0.002 | 3.5e-19 |
| rs6706095 | 2 | 46075677 | G | T | 0.760 | 0.020 | 0.002 | 4.9e-16 |
| rs75475627 | 2 | 54787592 | G | C | 0.076 | 0.024 | 0.004 | 2.5e-09 |
| rs778752 | 2 | 61776902 | G | C | 0.613 | -0.015 | 0.002 | 1.3e-12 |
| rs13026184 | 2 | 64930786 | G | C | 0.243 | 0.017 | 0.002 | 1.4e-12 |
| rs79709502 | 2 | 65087232 | G | C | 0.222 | 0.015 | 0.003 | 2.1e-09 |
| rs13417564 | 2 | 65665273 | T | G | 0.582 | 0.029 | 0.002 | 6e-44 |
| rs58448550 | 2 | 96880147 | T | G | 0.327 | 0.016 | 0.002 | 3.2e-12 |
| rs6543084 | 2 | 102294664 | C | T | 0.579 | 0.013 | 0.002 | 2.4e-09 |
| rs150449635 | 2 | 111752151 | C | T | 0.022 | 0.149 | 0.007 | 1.1e-90 |
| rs72836307 | 2 | 111776154 | T | C | 0.157 | -0.041 | 0.003 | 4.3e-47 |
| rs72836346 | 2 | 111876613 | C | G | 0.078 | 0.034 | 0.004 | 1.1e-17 |
| rs6735269 | 2 | 136885734 | A | G | 0.735 | -0.041 | 0.002 | 6.8e-67 |
| rs34005843 | 2 | 148548683 | G | C | 0.294 | 0.020 | 0.002 | 8.9e-18 |
| rs1549387 | 2 | 160417540 | G | A | 0.512 | 0.018 | 0.002 | 8e-19 |
| rs3931 | 2 | 169721381 | G | A | 0.321 | 0.022 | 0.002 | 4.7e-22 |
| rs4374330 | 2 | 182058432 | T | C | 0.747 | -0.032 | 0.002 | 1.1e-41 |
| rs6740847 | 2 | 182308352 | G | A | 0.564 | 0.129 | 0.002 | 1e-200 |
| rs34743120 | 2 | 182425330 | A | G | 0.019 | -0.046 | 0.008 | 1.2e-09 |
| rs145447411 | 2 | 182846767 | C | T | 0.024 | 0.043 | 0.007 | 6.7e-10 |
| rs4416233 | 2 | 196992495 | T | C | 0.720 | 0.014 | 0.002 | 3.3e-09 |
| rs10931936 | 2 | 202143928 | C | T | 0.732 | 0.014 | 0.002 | 3.2e-09 |
| rs6736362 | 2 | 219115108 | T | C | 0.562 | 0.021 | 0.002 | 1.3e-23 |
| rs6436124 | 2 | 220041433 | A | C | 0.609 | 0.015 | 0.002 | 7.6e-12 |
| rs2304336 | 2 | 225750861 | T | C | 0.157 | -0.042 | 0.003 | 4e-49 |
| rs4470337 | 2 | 228293381 | G | A | 0.598 | -0.013 | 0.002 | 1.9e-09 |
| rs13002735 | 2 | 232268884 | C | A | 0.238 | -0.015 | 0.002 | 2.5e-09 |
| rs34236350 | 2 | 241568326 | T | C | 0.183 | -0.033 | 0.003 | 2.9e-35 |
| rs1822534 | 3 | 12266804 | G | A | 0.392 | -0.032 | 0.002 | 5.5e-52 |
| rs869785 | 3 | 24347800 | C | T | 0.671 | -0.018 | 0.002 | 2.8e-16 |
| rs73058593 | 3 | 39281374 | C | G | 0.217 | 0.037 | 0.003 | 2.2e-49 |
| rs200623415 | 3 | 42831059 | A | C | 0.088 | 0.030 | 0.004 | 1.4e-15 |
| rs2228467 | 3 | 42906116 | C | T | 0.061 | 0.163 | 0.004 | 1e-200 |
| rs875890 | 3 | 45945287 | A | T | 0.260 | -0.017 | 0.002 | 3.7e-13 |
| rs2213290 | 3 | 46406367 | T | C | 0.409 | 0.037 | 0.002 | 7.2e-66 |
| rs374231748 | 3 | 46813660 | A | G | 0.054 | -0.033 | 0.005 | 1.3e-11 |
| rs13094390 | 3 | 71817778 | G | A | 0.116 | -0.021 | 0.003 | 6.9e-11 |
| rs9809116 | 3 | 72397279 | G | A | 0.408 | 0.019 | 0.002 | 3.6e-18 |
| rs62276455 | 3 | 98417481 | A | T | 0.443 | 0.024 | 0.002 | 2.8e-30 |
| rs167924 | 3 | 107379837 | G | A | 0.622 | 0.014 | 0.002 | 2.4e-10 |
| rs62263808 | 3 | 123086587 | A | G | 0.217 | -0.023 | 0.003 | 2.3e-19 |
| rs2291079 | 3 | 124483089 | G | C | 0.220 | -0.015 | 0.003 | 3.8e-09 |
| rs11713343 | 3 | 128185399 | A | G | 0.196 | -0.025 | 0.003 | 3.8e-22 |
| rs2245626 | 3 | 128287687 | G | A | 0.674 | 0.075 | 0.002 | 1e-200 |
| rs2669833 | 3 | 129604169 | A | G | 0.157 | -0.018 | 0.003 | 2.2e-10 |
| rs62410436 | 3 | 136597549 | A | C | 0.681 | 0.016 | 0.002 | 1.1e-12 |
| rs4683611 | 3 | 141290903 | C | T | 0.201 | -0.017 | 0.003 | 3.4e-09 |
| rs6440013 | 3 | 141304571 | G | A | 0.401 | -0.024 | 0.002 | 5.4e-30 |
| rs10804681 | 3 | 141660675 | T | A | 0.842 | -0.020 | 0.003 | 2.2e-12 |
| rs16831133 | 3 | 159923576 | C | T | 0.398 | -0.014 | 0.002 | 4.3e-11 |
| rs7633965 | 3 | 168864643 | C | A | 0.930 | 0.027 | 0.004 | 3.6e-11 |
| rs231988 | 3 | 172276979 | C | T | 0.868 | 0.027 | 0.003 | 5.8e-18 |
| rs9815073 | 3 | 188115682 | A | C | 0.343 | -0.023 | 0.002 | 1.4e-23 |
| rs7626444 | 3 | 196504902 | C | G | 0.423 | -0.029 | 0.002 | 2e-43 |
| rs2269486 | 4 | 2267566 | T | A | 0.161 | 0.021 | 0.003 | 1.5e-13 |
| rs6815294 | 4 | 7042349 | A | G | 0.572 | 0.039 | 0.002 | 1e-73 |
| rs73809166 | 4 | 36305937 | C | T | 0.023 | 0.045 | 0.007 | 2e-10 |
| rs2711981 | 4 | 39039258 | T | C | 0.663 | -0.023 | 0.002 | 1.4e-25 |
| rs723585 | 4 | 55503194 | G | A | 0.486 | -0.016 | 0.002 | 5.9e-15 |
| rs9637714 | 4 | 57731446 | C | T | 0.289 | -0.016 | 0.002 | 3.6e-12 |
| rs2282679 | 4 | 72608383 | G | T | 0.291 | -0.014 | 0.002 | 3.7e-09 |
| rs113172748 | 4 | 79639615 | G | A | 0.670 | 0.017 | 0.002 | 9.2e-14 |
| rs6832609 | 4 | 83556571 | G | A | 0.185 | -0.049 | 0.003 | 2.3e-73 |
| rs4566648 | 4 | 84159219 | G | T | 0.342 | -0.025 | 0.002 | 8.4e-31 |
| rs141936164 | 4 | 103401723 | G | A | 0.354 | -0.019 | 0.002 | 2.1e-18 |
| rs144317085 | 4 | 105806108 | T | A | 0.034 | 0.058 | 0.006 | 3.7e-23 |
| rs6533483 | 4 | 110887989 | A | G | 0.403 | -0.019 | 0.002 | 9.4e-20 |
| rs9307475 | 4 | 120368556 | A | G | 0.327 | -0.014 | 0.002 | 3.6e-10 |
| rs4565031 | 4 | 145044759 | A | G | 0.550 | -0.023 | 0.002 | 1.6e-27 |
| rs72720206 | 4 | 146105777 | A | G | 0.325 | -0.017 | 0.002 | 4.4e-14 |
| rs7687559 | 4 | 154385925 | C | G | 0.399 | 0.015 | 0.002 | 3.6e-12 |
| rs113862443 | 4 | 157725371 | G | C | 0.083 | -0.025 | 0.004 | 4.5e-11 |
| rs964751 | 5 | 17399882 | T | C | 0.531 | -0.013 | 0.002 | 1e-09 |
| rs116619972 | 5 | 32214314 | A | G | 0.027 | -0.046 | 0.007 | 2.5e-12 |
| rs4865956 | 5 | 54882505 | A | T | 0.696 | -0.019 | 0.002 | 2e-17 |
| rs28722705 | 5 | 55453942 | T | A | 0.148 | 0.036 | 0.003 | 1.2e-34 |
| rs74735005 | 5 | 57202727 | C | T | 0.207 | -0.016 | 0.003 | 4.9e-10 |
| rs6452473 | 5 | 71741094 | G | T | 0.877 | -0.032 | 0.003 | 3.5e-23 |
| rs31243 | 5 | 75594360 | G | A | 0.095 | -0.022 | 0.004 | 1.7e-09 |
| rs165944 | 5 | 88110363 | T | C | 0.765 | -0.016 | 0.002 | 1.4e-10 |
| rs1902801 | 5 | 110982340 | A | G | 0.280 | -0.017 | 0.002 | 7.4e-14 |
| rs2631367 | 5 | 131705458 | G | C | 0.522 | 0.022 | 0.002 | 3.6e-26 |
| rs12332674 | 5 | 132322852 | G | T | 0.152 | -0.017 | 0.003 | 2.2e-09 |
| rs329125 | 5 | 133871101 | A | C | 0.209 | -0.017 | 0.003 | 1.2e-10 |
| rs4362932 | 5 | 149483906 | T | C | 0.267 | 0.034 | 0.002 | 3.1e-47 |
| rs17656204 | 5 | 149501803 | T | C | 0.323 | -0.056 | 0.002 | 3.8e-135 |
| rs360017 | 5 | 173207353 | G | A | 0.775 | -0.023 | 0.003 | 7.4e-20 |
| rs6883116 | 5 | 179230321 | C | T | 0.417 | -0.017 | 0.002 | 1.3e-15 |
| rs55693514 | 6 | 6708847 | T | A | 0.703 | 0.015 | 0.002 | 7.2e-11 |
| rs9505057 | 6 | 7171407 | G | T | 0.203 | 0.030 | 0.003 | 4.6e-31 |
| rs707794 | 6 | 10519890 | A | C | 0.530 | -0.031 | 0.002 | 5.4e-48 |
| rs707846 | 6 | 16711874 | G | A | 0.509 | -0.017 | 0.002 | 1.4e-15 |
| rs28505913 | 6 | 21826915 | T | C | 0.141 | 0.019 | 0.003 | 5.6e-10 |
| rs261945 | 6 | 30272417 | G | C | 0.612 | -0.027 | 0.002 | 3.3e-37 |
| rs2523562 | 6 | 31330618 | T | C | 0.248 | -0.069 | 0.002 | 3e-176 |
| rs200801362 | 6 | 31555480 | C | T | 0.160 | -0.074 | 0.004 | 1e-81 |
| rs28361033 | 6 | 31973863 | T | C | 0.440 | 0.023 | 0.002 | 5.9e-26 |
| rs41552812 | 6 | 32632689 | T | C | 0.070 | -0.076 | 0.005 | 3.2e-46 |
| rs56007794 | 6 | 41990827 | T | A | 0.243 | 0.042 | 0.002 | 4.8e-67 |
| rs79894332 | 6 | 44591721 | A | G | 0.101 | -0.049 | 0.003 | 1.3e-44 |
| rs115202835 | 6 | 45533239 | A | G | 0.068 | 0.028 | 0.004 | 1e-11 |
| rs17470494 | 6 | 46883834 | G | A | 0.102 | -0.033 | 0.003 | 7.9e-22 |
| rs632497 | 6 | 53443926 | G | A | 0.198 | -0.020 | 0.003 | 1.7e-14 |
| rs915125 | 6 | 82463376 | T | C | 0.279 | 0.042 | 0.002 | 3.2e-74 |
| rs9480737 | 6 | 107442277 | G | A | 0.322 | -0.022 | 0.002 | 1.2e-23 |
| rs77791617 | 6 | 114083374 | T | C | 0.060 | 0.040 | 0.004 | 9e-20 |
| rs509596 | 6 | 122799541 | A | G | 0.554 | 0.018 | 0.002 | 7.6e-18 |
| rs34164109 | 6 | 135421176 | T | C | 0.261 | -0.038 | 0.002 | 4.6e-58 |
| rs9389287 | 6 | 135684975 | T | C | 0.278 | 0.017 | 0.002 | 8.2e-12 |
| rs9494571 | 6 | 137111121 | T | C | 0.593 | -0.016 | 0.002 | 2.3e-13 |
| rs2797670 | 6 | 137571613 | A | G | 0.648 | -0.013 | 0.002 | 4.2e-09 |
| rs6920282 | 6 | 140013130 | A | G | 0.326 | -0.014 | 0.002 | 1.1e-09 |
| rs149110519 | 6 | 144385777 | T | C | 0.036 | 0.082 | 0.006 | 7.5e-48 |
| rs9390460 | 6 | 147694334 | C | T | 0.540 | 0.013 | 0.002 | 1.6e-09 |
| rs2817441 | 6 | 156938699 | T | C | 0.280 | -0.021 | 0.002 | 6.7e-19 |
| rs3012415 | 6 | 170489464 | T | C | 0.822 | 0.021 | 0.003 | 4e-15 |
| rs2273215 | 6 | 170586082 | A | G | 0.459 | -0.018 | 0.002 | 2.4e-18 |
| rs2895 | 7 | 2568770 | C | T | 0.099 | 0.023 | 0.004 | 1.2e-10 |
| rs798531 | 7 | 2770067 | C | G | 0.428 | 0.016 | 0.002 | 1.1e-14 |
| rs6796 | 7 | 6502367 | C | T | 0.278 | 0.047 | 0.002 | 2.1e-90 |
| rs75814517 | 7 | 17145526 | A | G | 0.056 | -0.033 | 0.005 | 8.4e-13 |
| rs17437411 | 7 | 27198086 | T | C | 0.069 | -0.036 | 0.004 | 2.7e-18 |
| rs17156536 | 7 | 28279488 | A | C | 0.158 | 0.025 | 0.003 | 6.4e-18 |
| rs10238435 | 7 | 28724491 | T | C | 0.239 | -0.024 | 0.003 | 8.8e-22 |
| rs55959091 | 7 | 44771671 | A | G | 0.131 | 0.022 | 0.003 | 1.9e-12 |
| rs4385425 | 7 | 50307334 | G | A | 0.323 | -0.062 | 0.002 | 6.9e-168 |
| rs149007767 | 7 | 50370254 | T | C | 0.162 | -0.075 | 0.003 | 4.1e-142 |
| rs11766800 | 7 | 50435617 | T | A | 0.322 | 0.020 | 0.002 | 1.1e-19 |
| rs41430449 | 7 | 50798525 | G | C | 0.067 | -0.032 | 0.004 | 3.4e-14 |
| rs3981841 | 7 | 65184339 | C | G | 0.567 | -0.014 | 0.002 | 1.9e-11 |
| rs178405 | 7 | 73513045 | T | G | 0.786 | -0.016 | 0.003 | 2.8e-10 |
| rs445 | 7 | 92408370 | T | C | 0.096 | -0.093 | 0.004 | 2.3e-152 |
| rs6465668 | 7 | 97961469 | C | T | 0.482 | -0.017 | 0.002 | 1.4e-15 |
| rs142473518 | 7 | 99894971 | G | A | 0.190 | 0.018 | 0.003 | 6.4e-12 |
| rs4727673 | 7 | 107021301 | A | C | 0.526 | 0.014 | 0.002 | 2.2e-11 |
| rs71243006 | 7 | 114633041 | G | T | 0.104 | -0.021 | 0.003 | 6.7e-10 |
| rs1973325 | 7 | 137879428 | G | A | 0.692 | -0.028 | 0.002 | 1.3e-34 |
| rs28744543 | 7 | 138705803 | A | G | 0.509 | -0.013 | 0.002 | 1.9e-09 |
| rs71552657 | 7 | 143082534 | A | G | 0.211 | 0.021 | 0.003 | 2.5e-16 |
| rs4840638 | 8 | 6698248 | G | T | 0.536 | -0.024 | 0.002 | 9.6e-30 |
| rs17078436 | 8 | 6827165 | C | T | 0.016 | 0.076 | 0.008 | 2.5e-19 |
| rs7826487 | 8 | 6880925 | G | A | 0.116 | -0.050 | 0.003 | 8.5e-52 |
| rs6988319 | 8 | 6915673 | G | C | 0.608 | 0.015 | 0.002 | 1e-11 |
| rs7460395 | 8 | 11835375 | A | G | 0.512 | -0.016 | 0.002 | 1.1e-14 |
| rs62501136 | 8 | 23089429 | A | G | 0.222 | 0.021 | 0.003 | 8e-17 |
| rs3735759 | 8 | 27203039 | T | C | 0.867 | -0.019 | 0.003 | 5.5e-10 |
| rs62505144 | 8 | 30355038 | T | G | 0.230 | -0.019 | 0.002 | 8.7e-15 |
| rs4737009 | 8 | 41630405 | A | G | 0.236 | 0.017 | 0.002 | 1.6e-11 |
| rs45577137 | 8 | 48651633 | G | A | 0.045 | -0.068 | 0.005 | 3e-36 |
| rs333613 | 8 | 56785685 | T | C | 0.611 | -0.018 | 0.002 | 2e-17 |
| rs7824937 | 8 | 61392724 | G | A | 0.367 | 0.018 | 0.002 | 1.4e-16 |
| rs144345073 | 8 | 68811043 | A | T | 0.399 | -0.024 | 0.002 | 1.3e-28 |
| rs62510269 | 8 | 79012343 | G | A | 0.147 | -0.025 | 0.003 | 2e-17 |
| rs1863651 | 8 | 82066998 | T | A | 0.699 | -0.017 | 0.002 | 1.1e-13 |
| rs72673751 | 8 | 106578940 | C | T | 0.190 | -0.017 | 0.003 | 3.4e-10 |
| rs1954735 | 8 | 108239325 | C | T | 0.169 | 0.022 | 0.003 | 1.2e-15 |
| rs3808461 | 8 | 116597716 | T | C | 0.566 | -0.031 | 0.002 | 1e-47 |
| rs12334935 | 8 | 126617990 | A | G | 0.472 | -0.015 | 0.002 | 8.5e-13 |
| rs7843207 | 8 | 130570063 | A | C | 0.506 | -0.072 | 0.002 | 1e-200 |
| rs10101101 | 8 | 130691767 | A | G | 0.139 | -0.041 | 0.003 | 3.5e-40 |
| rs4397371 | 8 | 130701417 | T | C | 0.733 | -0.018 | 0.002 | 9e-14 |
| rs837227 | 8 | 130964654 | T | C | 0.744 | -0.017 | 0.002 | 4.2e-13 |
| rs10099546 | 8 | 144638734 | G | A | 0.142 | -0.021 | 0.003 | 1.8e-12 |
| rs34881325 | 9 | 2622134 | T | C | 0.376 | 0.018 | 0.002 | 1.1e-16 |
| rs7036656 | 9 | 21990457 | T | C | 0.722 | 0.029 | 0.002 | 4.2e-35 |
| rs10757287 | 9 | 22143570 | T | A | 0.162 | -0.051 | 0.003 | 2e-71 |
| rs11557154 | 9 | 34107505 | T | C | 0.127 | 0.033 | 0.003 | 2.1e-25 |
| rs679787 | 9 | 35040449 | C | T | 0.717 | 0.018 | 0.002 | 1.7e-14 |
| rs10758481 | 9 | 38195756 | C | T | 0.504 | -0.015 | 0.002 | 3.2e-13 |
| rs10780209 | 9 | 91472127 | A | G | 0.477 | 0.045 | 0.002 | 2.5e-105 |
| rs9410425 | 9 | 91562311 | A | G | 0.323 | -0.029 | 0.002 | 1e-37 |
| rs575107 | 9 | 113583908 | A | G | 0.070 | -0.034 | 0.004 | 5.7e-17 |
| rs1008158 | 9 | 113828811 | G | A | 0.342 | -0.071 | 0.002 | 1e-200 |
| rs13296236 | 9 | 113877078 | G | T | 0.362 | -0.071 | 0.002 | 1e-200 |
| rs13296201 | 9 | 114341971 | T | C | 0.192 | 0.050 | 0.003 | 2.6e-78 |
| rs2181148 | 9 | 114467450 | C | T | 0.652 | 0.027 | 0.002 | 6.5e-32 |
| rs72759267 | 9 | 126971204 | T | C | 0.157 | 0.024 | 0.003 | 5.1e-16 |
| rs3824527 | 9 | 130702642 | T | C | 0.099 | 0.025 | 0.003 | 7.6e-13 |
| rs687289 | 9 | 136137106 | A | G | 0.318 | -0.047 | 0.002 | 9e-97 |
| rs71477838 | 10 | 3827920 | T | C | 0.148 | -0.020 | 0.003 | 1.3e-11 |
| rs7918233 | 10 | 3919071 | A | G | 0.489 | 0.013 | 0.002 | 1.4e-10 |
| rs7919913 | 10 | 5926216 | A | G | 0.346 | 0.020 | 0.002 | 6.9e-19 |
| rs2646420 | 10 | 8467037 | T | C | 0.380 | -0.026 | 0.002 | 2.8e-33 |
| rs112955547 | 10 | 25217783 | A | G | 0.369 | -0.046 | 0.002 | 4.2e-100 |
| rs475616 | 10 | 30496905 | G | A | 0.656 | 0.015 | 0.002 | 5.1e-12 |
| rs72790861 | 10 | 44880207 | G | C | 0.308 | -0.021 | 0.002 | 1.1e-20 |
| rs61850681 | 10 | 50248924 | A | G | 0.237 | -0.023 | 0.002 | 5.7e-21 |
| rs2790180 | 10 | 60036246 | G | T | 0.112 | -0.021 | 0.003 | 2.6e-10 |
| rs2393573 | 10 | 61657500 | C | G | 0.327 | 0.016 | 0.002 | 2.6e-12 |
| rs224101 | 10 | 64543777 | A | T | 0.605 | 0.022 | 0.002 | 1.4e-24 |
| rs7919533 | 10 | 73525525 | C | T | 0.486 | 0.020 | 0.002 | 2.8e-22 |
| rs1870148 | 10 | 82271341 | A | G | 0.789 | 0.021 | 0.003 | 8.6e-17 |
| rs11189167 | 10 | 99125722 | G | A | 0.255 | 0.054 | 0.002 | 1.4e-112 |
| rs11190141 | 10 | 101292390 | T | C | 0.372 | -0.044 | 0.002 | 2.7e-91 |
| rs185575325 | 10 | 104665997 | G | C | 0.075 | -0.026 | 0.004 | 4.9e-11 |
| rs10885396 | 10 | 114711755 | T | C | 0.541 | -0.016 | 0.002 | 6.9e-14 |
| rs4980325 | 11 | 234451 | T | G | 0.543 | 0.014 | 0.002 | 6.2e-11 |
| rs11246065 | 11 | 324170 | G | A | 0.339 | -0.023 | 0.002 | 3.3e-26 |
| rs907612 | 11 | 1874221 | T | C | 0.380 | -0.035 | 0.002 | 4.3e-57 |
| rs9783374 | 11 | 2340619 | C | T | 0.851 | 0.021 | 0.003 | 1.1e-12 |
| rs7120300 | 11 | 8823493 | T | C | 0.737 | -0.016 | 0.002 | 4.7e-12 |
| rs1885523 | 11 | 33903814 | T | C | 0.507 | 0.014 | 0.002 | 1.3e-10 |
| rs2596397 | 11 | 47247621 | A | G | 0.808 | -0.022 | 0.003 | 7.9e-17 |
| rs534903026 | 11 | 57309610 | A | C | 0.123 | 0.020 | 0.003 | 3.1e-09 |
| rs6591578 | 11 | 60158649 | A | G | 0.630 | 0.023 | 0.002 | 3.2e-25 |
| rs11227332 | 11 | 65623739 | G | A | 0.193 | -0.016 | 0.003 | 3.1e-09 |
| rs4441044 | 11 | 69500363 | A | G | 0.645 | -0.019 | 0.002 | 2.3e-17 |
| rs4944832 | 11 | 72949172 | A | G | 0.414 | -0.016 | 0.002 | 2.7e-14 |
| rs17756204 | 11 | 88083639 | A | G | 0.093 | -0.029 | 0.004 | 7.3e-16 |
| rs61890767 | 11 | 100645477 | C | A | 0.262 | 0.017 | 0.002 | 9e-13 |
| rs17684670 | 11 | 101676821 | A | G | 0.065 | 0.035 | 0.004 | 2.6e-16 |
| rs227043 | 11 | 108231309 | A | C | 0.553 | 0.017 | 0.002 | 4.4e-16 |
| rs3782004 | 11 | 114070250 | C | T | 0.188 | -0.017 | 0.003 | 5.9e-10 |
| rs7110984 | 11 | 118442234 | A | G | 0.047 | 0.034 | 0.005 | 4.4e-12 |
| rs3794073 | 11 | 119146129 | C | T | 0.273 | -0.018 | 0.002 | 1.3e-14 |
| rs12577260 | 11 | 122506129 | G | C | 0.124 | 0.065 | 0.003 | 1.1e-92 |
| rs7108562 | 11 | 128350653 | C | T | 0.334 | 0.021 | 0.002 | 1.4e-20 |
| rs74400777 | 11 | 128634760 | T | C | 0.156 | -0.019 | 0.003 | 2e-09 |
| rs2535393 | 12 | 711662 | C | T | 0.600 | -0.017 | 0.002 | 1.4e-15 |
| rs7955734 | 12 | 4333159 | G | C | 0.210 | -0.024 | 0.003 | 6.7e-21 |
| rs10849448 | 12 | 6493351 | G | A | 0.753 | -0.054 | 0.002 | 5.5e-106 |
| rs78326895 | 12 | 10584254 | C | T | 0.144 | 0.044 | 0.003 | 1.3e-46 |
| rs4082413 | 12 | 30783184 | G | C | 0.511 | 0.017 | 0.002 | 2.1e-15 |
| rs2651369 | 12 | 32552769 | G | C | 0.323 | 0.014 | 0.002 | 1.2e-09 |
| rs1700159 | 12 | 52305786 | T | C | 0.772 | 0.019 | 0.002 | 7.6e-15 |
| rs11170652 | 12 | 54099076 | A | G | 0.194 | -0.019 | 0.003 | 2.3e-13 |
| rs35979828 | 12 | 54685880 | T | C | 0.070 | -0.044 | 0.004 | 4.3e-27 |
| rs73120022 | 12 | 57613780 | A | G | 0.248 | -0.014 | 0.002 | 3.2e-09 |
| rs1800973 | 12 | 69744014 | A | C | 0.061 | 0.126 | 0.004 | 1.4e-182 |
| rs7487314 | 12 | 88836215 | T | G | 0.701 | -0.015 | 0.002 | 5.8e-11 |
| rs4254123 | 12 | 89908218 | A | C | 0.707 | 0.019 | 0.002 | 4.5e-16 |
| rs147780246 | 12 | 101884399 | T | G | 0.019 | 0.045 | 0.008 | 4.5e-09 |
| rs12372532 | 12 | 108726376 | T | C | 0.079 | 0.023 | 0.004 | 2.8e-09 |
| rs7314538 | 12 | 110050110 | G | A | 0.441 | 0.017 | 0.002 | 3.9e-15 |
| rs3184504 | 12 | 111884608 | C | T | 0.519 | -0.047 | 0.002 | 3.4e-112 |
| rs28432783 | 12 | 124857823 | C | T | 0.606 | 0.014 | 0.002 | 8.9e-11 |
| rs10846740 | 12 | 125303282 | C | T | 0.858 | -0.023 | 0.003 | 4.6e-14 |
| rs2504209 | 13 | 28546766 | G | A | 0.811 | 0.019 | 0.003 | 6.6e-13 |
| rs188175496 | 13 | 28595940 | T | C | 0.024 | -0.099 | 0.007 | 7.2e-41 |
| rs76428106 | 13 | 28604007 | C | T | 0.013 | 0.555 | 0.010 | 1e-200 |
| rs17086242 | 13 | 28611221 | C | T | 0.041 | -0.070 | 0.005 | 2e-40 |
| rs138028125 | 13 | 28712689 | G | C | 0.035 | 0.139 | 0.006 | 9.9e-122 |
| rs1892548 | 13 | 41002641 | C | T | 0.657 | -0.064 | 0.002 | 3.1e-182 |
| rs9532580 | 13 | 41244260 | C | T | 0.264 | -0.030 | 0.002 | 1.4e-37 |
| rs9316470 | 13 | 50203763 | A | G | 0.126 | -0.020 | 0.003 | 6.1e-10 |
| rs809845 | 13 | 50724055 | T | C | 0.443 | 0.017 | 0.002 | 1.6e-16 |
| rs73217470 | 13 | 72513420 | G | A | 0.047 | 0.064 | 0.005 | 5.1e-38 |
| rs1146933 | 13 | 78404049 | C | T | 0.785 | -0.015 | 0.003 | 1.4e-09 |
| rs60699901 | 13 | 92009193 | C | T | 0.117 | 0.024 | 0.003 | 7.4e-13 |
| rs374039502 | 13 | 108960385 | A | T | 0.021 | -0.113 | 0.008 | 3.5e-48 |
| rs9514999 | 13 | 109827299 | T | C | 0.444 | 0.013 | 0.002 | 7.2e-10 |
| rs9555596 | 13 | 110011839 | C | T | 0.385 | 0.023 | 0.002 | 1.2e-25 |
| rs7983334 | 13 | 110303919 | A | G | 0.872 | -0.019 | 0.003 | 3.5e-09 |
| rs58814158 | 13 | 110820255 | G | T | 0.156 | -0.024 | 0.003 | 1.4e-16 |
| rs2239630 | 14 | 23589349 | G | A | 0.558 | 0.046 | 0.002 | 1.4e-106 |
| rs1052484 | 14 | 25281444 | C | G | 0.217 | 0.015 | 0.003 | 1.8e-09 |
| rs2038700 | 14 | 25461989 | C | T | 0.394 | 0.040 | 0.002 | 1.3e-77 |
| rs1400745 | 14 | 35355300 | G | A | 0.528 | 0.019 | 0.002 | 1.1e-18 |
| rs113670117 | 14 | 36099366 | T | C | 0.088 | -0.024 | 0.004 | 1.2e-10 |
| rs12147629 | 14 | 75307588 | G | A | 0.311 | 0.019 | 0.002 | 2.6e-17 |
| rs754388 | 14 | 93115410 | C | G | 0.811 | 0.029 | 0.003 | 9.2e-27 |
| rs4905043 | 14 | 93550009 | A | G | 0.399 | 0.018 | 0.002 | 7.1e-17 |
| rs62007660 | 14 | 103843344 | A | G | 0.737 | 0.064 | 0.002 | 3.9e-159 |
| rs11856829 | 15 | 39277781 | T | C | 0.438 | 0.014 | 0.002 | 1.2e-10 |
| rs72726037 | 15 | 42263979 | A | G | 0.111 | -0.056 | 0.003 | 5.4e-62 |
| rs3101851 | 15 | 50846896 | T | C | 0.214 | -0.017 | 0.003 | 1.8e-11 |
| rs8034610 | 15 | 64628917 | A | G | 0.879 | 0.061 | 0.003 | 2.6e-79 |
| rs2572207 | 15 | 66070693 | T | C | 0.240 | -0.025 | 0.002 | 4e-24 |
| rs8037798 | 15 | 75240030 | G | T | 0.231 | 0.023 | 0.003 | 1.8e-20 |
| rs1077965 | 15 | 80065337 | G | A | 0.425 | 0.017 | 0.002 | 1.3e-15 |
| rs2562756 | 15 | 80226326 | G | A | 0.329 | -0.026 | 0.002 | 2.9e-32 |
| rs8033923 | 15 | 80257747 | G | A | 0.210 | -0.078 | 0.003 | 1e-200 |
| rs2061822 | 15 | 86122779 | C | T | 0.673 | -0.016 | 0.002 | 3.8e-12 |
| rs3803478 | 15 | 99090692 | A | C | 0.198 | 0.021 | 0.003 | 3.1e-15 |
| rs1967309 | 16 | 4065583 | G | A | 0.605 | -0.023 | 0.002 | 2.1e-27 |
| rs8049116 | 16 | 4150530 | T | C | 0.117 | 0.025 | 0.003 | 3.3e-14 |
| rs11641185 | 16 | 10986596 | T | C | 0.733 | -0.016 | 0.002 | 4.3e-12 |
| rs10871483 | 16 | 29592725 | A | T | 0.517 | -0.015 | 0.002 | 3.2e-11 |
| rs7196129 | 16 | 30471109 | C | T | 0.528 | 0.027 | 0.002 | 2.2e-36 |
| rs35675346 | 16 | 30936081 | A | G | 0.240 | -0.019 | 0.002 | 5.3e-15 |
| rs11644125 | 16 | 57058974 | T | C | 0.600 | -0.020 | 0.002 | 8.7e-21 |
| rs34746889 | 16 | 67221444 | C | G | 0.018 | -0.055 | 0.008 | 6e-12 |
| rs12924604 | 16 | 74601486 | A | G | 0.198 | -0.018 | 0.003 | 4.3e-12 |
| rs11640701 | 16 | 84603083 | A | G | 0.303 | 0.015 | 0.002 | 1.3e-10 |
| rs428578 | 16 | 85917944 | G | A | 0.348 | -0.071 | 0.002 | 1e-200 |
| rs305071 | 16 | 85949271 | A | G | 0.120 | -0.083 | 0.003 | 1.9e-141 |
| rs2176777 | 16 | 86012521 | C | G | 0.813 | 0.056 | 0.003 | 8.1e-98 |
| rs4843885 | 16 | 86033186 | G | T | 0.183 | -0.023 | 0.003 | 7.4e-17 |
| rs16940328 | 16 | 86076108 | A | G | 0.113 | 0.039 | 0.003 | 4e-31 |
| rs58912472 | 17 | 1626018 | G | A | 0.527 | -0.012 | 0.002 | 3.1e-09 |
| rs10459917 | 17 | 2009752 | C | A | 0.203 | -0.027 | 0.003 | 1.9e-24 |
| rs9898876 | 17 | 7526962 | T | G | 0.199 | 0.019 | 0.003 | 2.4e-13 |
| rs118083884 | 17 | 16522922 | A | G | 0.016 | -0.096 | 0.009 | 6.4e-26 |
| rs4925114 | 17 | 17711270 | G | A | 0.654 | -0.019 | 0.002 | 1.9e-18 |
| rs62057793 | 17 | 25856325 | A | G | 0.111 | -0.021 | 0.003 | 7.8e-10 |
| rs880749 | 17 | 28059970 | C | T | 0.527 | -0.034 | 0.002 | 8.4e-59 |
| rs7223589 | 17 | 29728284 | C | T | 0.615 | 0.015 | 0.002 | 1.7e-11 |
| rs138899494 | 17 | 32540836 | A | G | 0.139 | 0.022 | 0.003 | 1.2e-12 |
| rs1016680 | 17 | 35874385 | C | A | 0.635 | -0.027 | 0.002 | 2.8e-34 |
| rs2302774 | 17 | 38183090 | T | G | 0.382 | -0.020 | 0.002 | 2.5e-20 |
| rs2074163 | 17 | 40270957 | A | G | 0.176 | 0.019 | 0.003 | 9.9e-12 |
| rs62054807 | 17 | 43915497 | T | C | 0.293 | 0.022 | 0.002 | 6.1e-22 |
| rs34097845 | 17 | 56358429 | T | C | 0.057 | -0.114 | 0.005 | 4.3e-133 |
| rs7212018 | 17 | 56411460 | A | G | 0.131 | 0.025 | 0.003 | 4e-16 |
| rs2526359 | 17 | 57933508 | G | A | 0.436 | -0.039 | 0.002 | 2.4e-75 |
| rs236513 | 17 | 68169458 | C | T | 0.118 | -0.021 | 0.003 | 1.1e-10 |
| rs2084312 | 17 | 72695211 | T | C | 0.800 | 0.036 | 0.003 | 4.2e-44 |
| rs16978176 | 17 | 72761242 | C | T | 0.469 | -0.025 | 0.002 | 8.3e-34 |
| rs745570 | 17 | 77781725 | G | A | 0.514 | 0.015 | 0.002 | 1.7e-12 |
| rs9330536 | 17 | 81083854 | C | T | 0.487 | 0.064 | 0.002 | 1.9e-172 |
| rs238135 | 18 | 3448979 | C | T | 0.290 | -0.015 | 0.002 | 1.5e-10 |
| rs4006418 | 18 | 19710333 | T | C | 0.084 | 0.027 | 0.004 | 7.4e-13 |
| rs12327253 | 18 | 20719164 | G | A | 0.288 | 0.020 | 0.002 | 4.4e-18 |
| rs718515 | 18 | 43856297 | A | G | 0.553 | -0.022 | 0.002 | 2.2e-25 |
| rs8084255 | 18 | 48144754 | T | C | 0.373 | 0.015 | 0.002 | 1.3e-11 |
| rs954954 | 18 | 60902328 | C | A | 0.105 | -0.030 | 0.003 | 2.8e-18 |
| rs17758695 | 18 | 60920854 | T | C | 0.029 | -0.113 | 0.006 | 6e-74 |
| rs10460159 | 18 | 60996360 | C | T | 0.429 | -0.016 | 0.002 | 8.1e-15 |
| rs3177609 | 18 | 74071078 | C | T | 0.077 | 0.031 | 0.004 | 1.2e-15 |
| rs2007483 | 18 | 77476131 | A | T | 0.382 | -0.025 | 0.002 | 4.4e-30 |
| rs61242663 | 19 | 836059 | T | C | 0.258 | 0.027 | 0.002 | 3.7e-28 |
| rs113772652 | 19 | 1031550 | T | C | 0.459 | 0.021 | 0.002 | 2.2e-22 |
| rs2873040 | 19 | 3172494 | T | C | 0.022 | -0.050 | 0.007 | 5.4e-12 |
| rs12980774 | 19 | 3410135 | C | T | 0.222 | -0.017 | 0.003 | 4.9e-11 |
| rs62107586 | 19 | 6468331 | C | T | 0.448 | -0.014 | 0.002 | 6.3e-12 |
| rs413141 | 19 | 6675989 | G | A | 0.863 | 0.045 | 0.003 | 3.8e-48 |
| rs4239552 | 19 | 7848679 | T | C | 0.219 | -0.021 | 0.003 | 5.4e-17 |
| rs35928002 | 19 | 18116144 | G | A | 0.306 | -0.034 | 0.002 | 3.7e-49 |
| rs10406080 | 19 | 18402142 | G | C | 0.616 | 0.029 | 0.002 | 7.9e-42 |
| rs79802726 | 19 | 18654587 | T | G | 0.187 | 0.020 | 0.003 | 4.1e-13 |
| rs7249692 | 19 | 19670688 | C | T | 0.671 | -0.018 | 0.002 | 1.3e-15 |
| rs62126615 | 19 | 33749710 | T | C | 0.167 | 0.023 | 0.003 | 3.6e-16 |
| rs12151289 | 19 | 33751852 | C | G | 0.030 | -0.088 | 0.006 | 1.3e-45 |
| rs8111664 | 19 | 44295092 | A | G | 0.424 | -0.041 | 0.002 | 2.9e-82 |
| rs13344267 | 19 | 45137012 | C | A | 0.200 | 0.016 | 0.003 | 1.8e-09 |
| rs439401 | 19 | 45414451 | C | T | 0.638 | -0.016 | 0.002 | 1.9e-13 |
| rs344812 | 19 | 45791838 | C | A | 0.306 | -0.051 | 0.002 | 9e-111 |
| rs11669500 | 19 | 52144572 | C | T | 0.383 | -0.019 | 0.002 | 1.3e-16 |
| rs62143206 | 19 | 54326212 | T | G | 0.211 | -0.073 | 0.003 | 2.1e-179 |
| rs11666987 | 19 | 55698477 | T | G | 0.161 | 0.018 | 0.003 | 8.2e-10 |
| rs34600126 | 20 | 1597433 | T | G | 0.219 | 0.023 | 0.003 | 2.1e-19 |
| rs4813619 | 20 | 2815715 | T | G | 0.511 | 0.017 | 0.002 | 1.5e-15 |
| rs628977 | 20 | 3649721 | C | T | 0.629 | -0.014 | 0.002 | 4e-10 |
| rs1883932 | 20 | 8609588 | T | A | 0.508 | -0.029 | 0.002 | 3.7e-44 |
| rs192009275 | 20 | 30250942 | T | C | 0.255 | -0.021 | 0.002 | 1.9e-17 |
| rs4911236 | 20 | 31082056 | A | G | 0.308 | -0.032 | 0.002 | 4.4e-42 |
| rs61114499 | 20 | 31192357 | A | G | 0.259 | 0.055 | 0.002 | 6.4e-115 |
| rs4142441 | 20 | 42839620 | G | A | 0.151 | 0.035 | 0.003 | 1.2e-31 |
| rs6125961 | 20 | 48884124 | A | C | 0.200 | -0.072 | 0.003 | 2.6e-167 |
| rs4350814 | 20 | 48969177 | A | G | 0.176 | -0.047 | 0.003 | 9e-65 |
| rs6091164 | 20 | 49073733 | T | G | 0.063 | -0.036 | 0.004 | 1.8e-16 |
| rs3787513 | 20 | 61573736 | A | G | 0.209 | -0.017 | 0.003 | 5.2e-11 |
| rs1997577 | 21 | 16371102 | T | A | 0.155 | -0.033 | 0.003 | 7.9e-30 |
| rs11088296 | 21 | 36237546 | T | C | 0.263 | 0.017 | 0.002 | 1.1e-12 |
| rs10460712 | 21 | 36292228 | C | A | 0.898 | -0.024 | 0.003 | 5.3e-12 |
| rs34825969 | 21 | 39846142 | A | G | 0.135 | -0.026 | 0.003 | 5.4e-17 |
| rs41409548 | 22 | 17579495 | A | G | 0.033 | -0.103 | 0.006 | 3.1e-60 |
| rs5748937 | 22 | 17675324 | T | C | 0.054 | -0.069 | 0.005 | 2.1e-44 |
| rs138632530 | 22 | 17676901 | T | C | 0.108 | 0.025 | 0.004 | 1.9e-12 |
| rs2019180 | 22 | 17712142 | A | G | 0.092 | -0.035 | 0.004 | 2.7e-20 |
| rs35720801 | 22 | 24639878 | A | G | 0.275 | -0.029 | 0.002 | 1e-33 |
| rs9625746 | 22 | 29637658 | C | G | 0.416 | -0.021 | 0.002 | 2.5e-23 |
| rs3091374 | 22 | 39480245 | T | C | 0.558 | 0.016 | 0.002 | 5.8e-15 |
| rs5758364 | 22 | 41855912 | C | A | 0.776 | 0.016 | 0.003 | 1.6e-09 |
| rs138918 | 22 | 43559451 | A | G | 0.392 | 0.031 | 0.002 | 5.8e-47 |
| rs739239 | 22 | 50050468 | A | G | 0.613 | 0.018 | 0.002 | 1.4e-17 |

**Table S10. 469 SNPs significantly associated with monocyte count used IVs in forward MR analyses derived from Chen MH et al.** Chr: Chromosome; EA: Effect allele; NEA: Non-effect allele; EAF: Effect allele frequency.

| SNP | Chr | Pos | EA | NEA | EAF | Beta | SE | P-value |
| --- | --- | --- | --- | --- | --- | --- | --- | --- |
| rs11190141 | 10 | 101292390 | T | C | 0.372 | -0.044 | 0.002 | 4.02e-113 |
| rs11191514 | 10 | 104773364 | T | C | 0.080 | -0.028 | 0.003 | 1.55e-15 |
| rs7095778 | 10 | 111929369 | G | A | 0.091 | -0.020 | 0.003 | 3.56e-10 |
| rs10885396 | 10 | 114711755 | T | C | 0.542 | -0.014 | 0.002 | 2.71e-14 |
| rs10828725 | 10 | 25218243 | T | G | 0.368 | -0.047 | 0.002 | 3.56e-121 |
| rs475616 | 10 | 30496905 | G | A | 0.656 | 0.016 | 0.002 | 2.97e-15 |
| rs1043009 | 10 | 3819714 | T | C | 0.377 | -0.013 | 0.002 | 5.81e-12 |
| rs7918233 | 10 | 3919071 | A | G | 0.489 | 0.012 | 0.002 | 1.54e-10 |
| rs72790862 | 10 | 44880260 | C | T | 0.307 | -0.020 | 0.002 | 1.92e-23 |
| rs17011726 | 10 | 50264204 | G | C | 0.233 | -0.023 | 0.002 | 1.23e-24 |
| rs10795595 | 10 | 5895923 | T | A | 0.425 | 0.020 | 0.002 | 3.17e-25 |
| rs1180658 | 10 | 61645945 | A | C | 0.634 | -0.013 | 0.002 | 1.19e-10 |
| rs224111 | 10 | 64552010 | A | G | 0.390 | -0.021 | 0.002 | 1.16e-27 |
| rs7919533 | 10 | 73525525 | C | T | 0.488 | 0.020 | 0.002 | 1.55e-26 |
| rs77637782 | 10 | 80924258 | T | G | 0.433 | -0.012 | 0.002 | 7.39e-10 |
| rs2646421 | 10 | 8467155 | C | G | 0.378 | -0.026 | 0.002 | 3.28e-41 |
| rs2184697 | 10 | 89851815 | G | T | 0.293 | -0.015 | 0.002 | 1.23e-12 |
| rs11189154 | 10 | 99108922 | A | G | 0.256 | 0.051 | 0.002 | 1.25e-122 |
| rs71482156 | 11 | 118734516 | T | A | 0.016 | 0.056 | 0.008 | 4.71e-12 |
| rs10892342 | 11 | 119135814 | C | T | 0.276 | -0.018 | 0.002 | 7.02e-18 |
| rs662333 | 11 | 121578945 | A | G | 0.780 | -0.014 | 0.002 | 1.05e-09 |
| rs11602323 | 11 | 122519281 | G | T | 0.123 | 0.065 | 0.003 | 9.22e-115 |
| rs695113 | 11 | 128562098 | T | C | 0.697 | 0.021 | 0.002 | 1.04e-24 |
| rs907612 | 11 | 1874221 | T | C | 0.380 | -0.033 | 0.002 | 5.19e-60 |
| rs4980325 | 11 | 234451 | T | G | 0.541 | 0.014 | 0.002 | 5.73e-13 |
| rs11246065 | 11 | 324170 | G | A | 0.339 | -0.022 | 0.002 | 1.6e-26 |
| rs1885525 | 11 | 33904180 | A | G | 0.506 | 0.013 | 0.002 | 7.62e-12 |
| rs2957873 | 11 | 47249294 | A | G | 0.809 | -0.021 | 0.002 | 9.44e-19 |
| rs573790 | 11 | 59855385 | C | T | 0.614 | -0.016 | 0.002 | 1.48e-16 |
| rs6591578 | 11 | 60158649 | A | G | 0.634 | 0.022 | 0.002 | 2.03e-29 |
| rs10796828 | 11 | 69490346 | G | T | 0.635 | -0.018 | 0.002 | 3.26e-20 |
| rs11235689 | 11 | 72949747 | T | C | 0.414 | -0.016 | 0.002 | 4.79e-16 |
| rs72966841 | 11 | 88070110 | T | A | 0.094 | -0.027 | 0.003 | 1.61e-16 |
| rs7120300 | 11 | 8823493 | T | C | 0.735 | -0.017 | 0.002 | 6.41e-16 |
| rs7975680 | 12 | 101864287 | C | A | 0.604 | 0.013 | 0.002 | 2.12e-10 |
| rs2734442 | 12 | 10600368 | A | G | 0.876 | -0.042 | 0.003 | 3e-49 |
| rs10744732 | 12 | 1078076 | T | C | 0.378 | 0.012 | 0.002 | 2.49e-09 |
| rs4365101 | 12 | 108707583 | A | G | 0.068 | -0.027 | 0.004 | 1.14e-12 |
| rs9943753 | 12 | 109840940 | G | A | 0.628 | 0.014 | 0.002 | 4.79e-12 |
| rs3184504 | 12 | 111884608 | C | T | 0.517 | -0.047 | 0.002 | 1.58e-139 |
| rs73201961 | 12 | 116830960 | C | A | 0.089 | 0.027 | 0.003 | 1.44e-16 |
| rs7308348 | 12 | 122225420 | C | T | 0.182 | 0.015 | 0.002 | 4.78e-10 |
| rs11057841 | 12 | 125316743 | T | C | 0.144 | 0.020 | 0.003 | 8.16e-14 |
| rs4082413 | 12 | 30783184 | G | C | 0.512 | 0.015 | 0.002 | 2.06e-16 |
| rs2651369 | 12 | 32552769 | G | C | 0.326 | 0.014 | 0.002 | 1.45e-12 |
| rs10849020 | 12 | 4332009 | G | C | 0.210 | -0.024 | 0.002 | 1.28e-25 |
| rs1700159 | 12 | 52305786 | T | C | 0.774 | 0.020 | 0.002 | 1.83e-19 |
| rs12309572 | 12 | 54074686 | T | C | 0.168 | 0.019 | 0.003 | 2.44e-14 |
| rs35979828 | 12 | 54685880 | T | C | 0.071 | -0.046 | 0.004 | 1.28e-35 |
| rs7485577 | 12 | 57616061 | A | G | 0.269 | -0.015 | 0.002 | 9.09e-12 |
| rs10849448 | 12 | 6493351 | G | A | 0.753 | -0.050 | 0.002 | 2.77e-111 |
| rs1800973 | 12 | 69744014 | A | C | 0.062 | 0.113 | 0.004 | 1.64e-187 |
| rs11104881 | 12 | 88843474 | C | T | 0.702 | -0.016 | 0.002 | 3.35e-14 |
| rs12306790 | 12 | 89864047 | T | C | 0.715 | 0.019 | 0.002 | 2.22e-20 |
| rs9555596 | 13 | 110011839 | C | T | 0.383 | 0.023 | 0.002 | 1.55e-32 |
| rs7983334 | 13 | 110303919 | A | G | 0.872 | -0.019 | 0.003 | 3.38e-11 |
| rs58814158 | 13 | 110820255 | G | T | 0.157 | -0.024 | 0.003 | 3.24e-19 |
| rs9583493 | 13 | 111091632 | A | C | 0.427 | 0.011 | 0.002 | 3.12e-09 |
| rs2183246 | 13 | 114901188 | T | C | 0.683 | -0.014 | 0.002 | 1.94e-11 |
| rs2504209 | 13 | 28546766 | G | A | 0.811 | 0.018 | 0.002 | 3.58e-14 |
| rs188175496 | 13 | 28595940 | T | C | 0.024 | -0.093 | 0.007 | 2.93e-43 |
| rs76428106 | 13 | 28604007 | C | T | 0.013 | 0.536 | 0.009 | 1e-200 |
| rs17086242 | 13 | 28611221 | C | T | 0.041 | -0.069 | 0.005 | 1.71e-47 |
| rs138028125 | 13 | 28712689 | G | C | 0.035 | 0.135 | 0.006 | 6.19e-131 |
| rs1892548 | 13 | 41002641 | C | T | 0.655 | -0.063 | 0.002 | 1e-200 |
| rs9532580 | 13 | 41244260 | C | T | 0.264 | -0.028 | 0.002 | 9.72e-40 |
| rs9316470 | 13 | 50203763 | A | G | 0.127 | -0.017 | 0.003 | 1.72e-09 |
| rs809845 | 13 | 50724055 | T | C | 0.443 | 0.015 | 0.002 | 3.57e-16 |
| rs73217470 | 13 | 72513420 | G | A | 0.047 | 0.065 | 0.004 | 2.11e-47 |
| rs1146933 | 13 | 78404049 | C | T | 0.786 | -0.014 | 0.002 | 8.88e-10 |
| rs60699901 | 13 | 92009193 | C | T | 0.118 | 0.022 | 0.003 | 2.02e-13 |
| rs8016326 | 14 | 103846716 | A | G | 0.734 | 0.061 | 0.002 | 3.81e-178 |
| rs4983387 | 14 | 105268228 | A | G | 0.899 | 0.019 | 0.003 | 2.2e-09 |
| rs2239630 | 14 | 23589349 | G | A | 0.556 | 0.049 | 0.002 | 4.68e-146 |
| rs1052484 | 14 | 25281444 | C | G | 0.216 | 0.014 | 0.002 | 1.28e-09 |
| rs2038700 | 14 | 25461989 | C | T | 0.394 | 0.040 | 0.002 | 2.81e-96 |
| rs754388 | 14 | 93115410 | C | G | 0.811 | 0.030 | 0.002 | 6.43e-36 |
| rs4905043 | 14 | 93550009 | A | G | 0.400 | 0.019 | 0.002 | 1.31e-20 |
| rs62018159 | 15 | 40397421 | A | G | 0.209 | 0.015 | 0.002 | 1.42e-10 |
| rs1002774 | 15 | 42261781 | A | G | 0.112 | -0.054 | 0.003 | 4.48e-73 |
| rs60695341 | 15 | 51010271 | T | C | 0.197 | -0.018 | 0.002 | 1.14e-14 |
| rs6493575 | 15 | 53013475 | T | C | 0.396 | -0.012 | 0.002 | 1.86e-10 |
| rs62011334 | 15 | 63837491 | G | A | 0.355 | 0.012 | 0.002 | 2.1e-09 |
| rs7180079 | 15 | 64629873 | G | A | 0.877 | 0.059 | 0.003 | 3.41e-95 |
| rs11857609 | 15 | 66095270 | C | T | 0.760 | 0.023 | 0.002 | 3.99e-26 |
| rs1081230 | 15 | 80095646 | G | A | 0.577 | -0.020 | 0.002 | 8.43e-25 |
| rs2061822 | 15 | 86122779 | C | T | 0.674 | -0.014 | 0.002 | 8.67e-12 |
| rs73467599 | 15 | 99088453 | C | T | 0.199 | 0.018 | 0.002 | 5.95e-15 |
| rs11641185 | 16 | 10986596 | T | C | 0.732 | -0.015 | 0.002 | 5.06e-13 |
| rs7196129 | 16 | 30471109 | C | T | 0.530 | 0.025 | 0.002 | 2.45e-39 |
| rs7185007 | 16 | 30927509 | T | C | 0.240 | -0.017 | 0.002 | 2.4e-14 |
| rs1967309 | 16 | 4065583 | G | A | 0.603 | -0.022 | 0.002 | 1.43e-30 |
| rs11508026 | 16 | 56999328 | T | C | 0.431 | -0.012 | 0.002 | 2.7e-10 |
| rs11644125 | 16 | 57058974 | T | C | 0.598 | -0.019 | 0.002 | 4.73e-24 |
| rs9937847 | 16 | 85917551 | C | T | 0.071 | -0.112 | 0.004 | 1e-200 |
| rs366078 | 16 | 85964563 | C | T | 0.158 | 0.107 | 0.003 | 1e-200 |
| rs76209847 | 16 | 85967249 | T | C | 0.037 | -0.075 | 0.005 | 7.35e-49 |
| rs2885363 | 16 | 86009261 | C | G | 0.079 | 0.117 | 0.004 | 1e-200 |
| rs140634734 | 16 | 86018831 | T | C | 0.028 | 0.047 | 0.006 | 4.27e-15 |
| rs79252394 | 16 | 86021968 | T | G | 0.030 | -0.043 | 0.006 | 2.91e-14 |
| rs12942576 | 17 | 1312461 | C | T | 0.683 | 0.012 | 0.002 | 2.19e-09 |
| rs118083884 | 17 | 16522922 | A | G | 0.017 | -0.097 | 0.009 | 1.01e-29 |
| rs12941356 | 17 | 17716531 | G | A | 0.588 | -0.017 | 0.002 | 2.45e-18 |
| rs9915112 | 17 | 2007826 | G | A | 0.203 | -0.027 | 0.002 | 4.21e-30 |
| rs62057793 | 17 | 25856325 | A | G | 0.112 | -0.019 | 0.003 | 8e-11 |
| rs71368117 | 17 | 27329227 | C | T | 0.087 | -0.024 | 0.003 | 1.58e-11 |
| rs2729450 | 17 | 28088459 | C | T | 0.524 | -0.031 | 0.002 | 1.36e-59 |
| rs17614093 | 17 | 32509256 | G | C | 0.430 | -0.015 | 0.002 | 7.17e-15 |
| rs1016680 | 17 | 35874385 | C | A | 0.637 | -0.026 | 0.002 | 4.31e-39 |
| rs2302774 | 17 | 38183090 | T | G | 0.383 | -0.024 | 0.002 | 5.68e-36 |
| rs2074163 | 17 | 40270957 | A | G | 0.177 | 0.019 | 0.002 | 4.21e-14 |
| rs62054807 | 17 | 43915497 | T | C | 0.295 | 0.020 | 0.002 | 2.45e-22 |
| rs34097845 | 17 | 56358429 | T | C | 0.056 | -0.101 | 0.004 | 2.99e-124 |
| rs11656162 | 17 | 56404625 | C | G | 0.034 | 0.035 | 0.005 | 7.65e-11 |
| rs2665405 | 17 | 57875292 | A | G | 0.548 | 0.038 | 0.002 | 5.37e-93 |
| rs236513 | 17 | 68169458 | C | T | 0.118 | -0.020 | 0.003 | 6.64e-12 |
| rs2084312 | 17 | 72695211 | T | C | 0.800 | 0.038 | 0.002 | 8.97e-58 |
| rs16978176 | 17 | 72761242 | C | T | 0.465 | -0.025 | 0.002 | 2.89e-40 |
| rs9898876 | 17 | 7526962 | T | G | 0.197 | 0.018 | 0.002 | 9.89e-14 |
| rs745570 | 17 | 77781725 | G | A | 0.514 | 0.014 | 0.002 | 5.38e-13 |
| rs9674881 | 17 | 81078568 | T | C | 0.486 | 0.063 | 0.002 | 2.16e-189 |
| rs238136 | 18 | 3448755 | G | A | 0.250 | -0.015 | 0.002 | 1.68e-11 |
| rs11082397 | 18 | 42094299 | G | T | 0.129 | 0.019 | 0.003 | 1.36e-11 |
| rs718515 | 18 | 43856297 | A | G | 0.552 | -0.020 | 0.002 | 5.66e-26 |
| rs9963693 | 18 | 45599257 | C | T | 0.261 | 0.013 | 0.002 | 5.39e-10 |
| rs745822 | 18 | 48142822 | G | T | 0.376 | 0.016 | 0.002 | 3.16e-17 |
| rs954954 | 18 | 60902328 | C | A | 0.105 | -0.029 | 0.003 | 4.2e-21 |
| rs141390096 | 18 | 60911544 | C | G | 0.037 | -0.033 | 0.005 | 4.2e-11 |
| rs17758695 | 18 | 60920854 | T | C | 0.029 | -0.109 | 0.006 | 3.89e-80 |
| rs10460159 | 18 | 60996360 | C | T | 0.429 | -0.016 | 0.002 | 9.88e-17 |
| rs3177609 | 18 | 74071078 | C | T | 0.078 | 0.031 | 0.004 | 8.26e-19 |
| rs2007483 | 18 | 77476131 | A | T | 0.382 | -0.024 | 0.002 | 1.63e-34 |
| rs10403909 | 19 | 1008358 | G | A | 0.724 | 0.019 | 0.002 | 1.95e-17 |
| rs11086102 | 19 | 18398628 | C | G | 0.633 | 0.031 | 0.002 | 2.26e-54 |
| rs3859570 | 19 | 18510925 | C | T | 0.425 | -0.028 | 0.002 | 3.97e-44 |
| rs7249692 | 19 | 19670688 | C | T | 0.671 | -0.017 | 0.002 | 8.83e-18 |
| rs72987040 | 19 | 3251116 | A | C | 0.124 | -0.025 | 0.003 | 2.71e-18 |
| rs62126615 | 19 | 33749710 | T | C | 0.168 | 0.020 | 0.003 | 3.69e-16 |
| rs74259566 | 19 | 42735177 | G | C | 0.101 | 0.029 | 0.003 | 7.51e-20 |
| rs56344893 | 19 | 44282529 | A | C | 0.373 | -0.044 | 0.002 | 2.12e-114 |
| rs10425556 | 19 | 45259871 | T | G | 0.062 | 0.026 | 0.004 | 1.94e-10 |
| rs11669910 | 19 | 45741333 | T | A | 0.258 | -0.067 | 0.002 | 1e-200 |
| rs33978622 | 19 | 51736383 | C | G | 0.329 | -0.018 | 0.002 | 7.51e-19 |
| rs3803904 | 19 | 55699646 | T | C | 0.134 | 0.017 | 0.003 | 1.96e-09 |
| rs413141 | 19 | 6675989 | G | A | 0.863 | 0.043 | 0.003 | 1.42e-54 |
| rs571497 | 19 | 7827830 | A | G | 0.155 | -0.028 | 0.003 | 8.94e-27 |
| rs6687430 | 1 | 10633245 | A | G | 0.543 | 0.012 | 0.002 | 2.41e-10 |
| rs284324 | 1 | 10724236 | A | G | 0.495 | 0.021 | 0.002 | 5.94e-27 |
| rs333947 | 1 | 110470764 | A | G | 0.150 | 0.037 | 0.003 | 2.21e-44 |
| rs11102144 | 1 | 111195493 | G | A | 0.166 | -0.016 | 0.003 | 1.43e-10 |
| rs67224956 | 1 | 118154575 | C | T | 0.170 | 0.027 | 0.002 | 2.01e-27 |
| rs6665912 | 1 | 150544093 | C | T | 0.223 | -0.072 | 0.002 | 1e-200 |
| rs60752752 | 1 | 153339782 | G | A | 0.118 | 0.025 | 0.003 | 2.95e-17 |
| rs3027012 | 1 | 159174123 | T | C | 0.183 | 0.028 | 0.002 | 4.37e-31 |
| rs4987353 | 1 | 169666987 | A | G | 0.310 | -0.022 | 0.002 | 1.93e-27 |
| rs3795503 | 1 | 180905694 | T | C | 0.315 | -0.019 | 0.002 | 1.68e-20 |
| rs11590380 | 1 | 185400939 | C | T | 0.679 | 0.025 | 0.002 | 1.69e-32 |
| rs12747432 | 1 | 198566554 | T | G | 0.067 | -0.024 | 0.004 | 1.31e-10 |
| rs9787298 | 1 | 204276040 | C | A | 0.211 | -0.016 | 0.002 | 2.66e-12 |
| rs1772143 | 1 | 205799987 | A | T | 0.413 | 0.020 | 0.002 | 8.11e-26 |
| rs7522307 | 1 | 207998783 | C | G | 0.124 | -0.023 | 0.003 | 4.36e-16 |
| rs701905 | 1 | 212553111 | C | G | 0.352 | 0.022 | 0.002 | 1.63e-28 |
| rs2784250 | 1 | 221060393 | G | T | 0.140 | -0.017 | 0.003 | 3.11e-10 |
| rs1335929 | 1 | 221136787 | C | T | 0.571 | 0.017 | 0.002 | 1.77e-18 |
| rs113292043 | 1 | 223231657 | T | C | 0.066 | -0.025 | 0.004 | 5.45e-11 |
| rs6696074 | 1 | 225962690 | T | C | 0.563 | 0.018 | 0.002 | 1.49e-21 |
| rs4626924 | 1 | 234909298 | T | C | 0.551 | -0.018 | 0.002 | 8.2e-22 |
| rs4269828 | 1 | 235257838 | G | A | 0.560 | 0.012 | 0.002 | 1.89e-10 |
| rs6429432 | 1 | 236107241 | C | A | 0.892 | -0.080 | 0.003 | 6.29e-156 |
| rs1212036 | 1 | 23690216 | T | C | 0.114 | -0.018 | 0.003 | 1.72e-09 |
| rs61165644 | 1 | 244487940 | G | A | 0.141 | -0.026 | 0.003 | 2.59e-22 |
| rs4335411 | 1 | 249191706 | A | G | 0.762 | -0.017 | 0.002 | 5.51e-13 |
| rs11247908 | 1 | 26638222 | A | G | 0.173 | -0.029 | 0.002 | 1.92e-32 |
| rs188393352 | 1 | 28254136 | G | C | 0.010 | 0.087 | 0.010 | 2.36e-17 |
| rs35351292 | 1 | 31208042 | T | C | 0.265 | 0.014 | 0.002 | 6.97e-11 |
| rs41268099 | 1 | 41328644 | A | G | 0.111 | 0.029 | 0.003 | 2.47e-22 |
| rs2786487 | 1 | 42368339 | C | G | 0.559 | 0.015 | 0.002 | 3.33e-15 |
| rs17387886 | 1 | 43419705 | C | G | 0.174 | -0.028 | 0.003 | 1.45e-28 |
| rs55684236 | 1 | 44080009 | A | G | 0.348 | 0.012 | 0.002 | 3.89e-09 |
| rs2274664 | 1 | 54704629 | T | C | 0.580 | -0.015 | 0.002 | 2.75e-15 |
| rs12738019 | 1 | 54876199 | G | A | 0.556 | -0.013 | 0.002 | 2.21e-12 |
| rs72675573 | 1 | 56636881 | T | C | 0.366 | 0.016 | 0.002 | 1.76e-16 |
| rs1933295 | 1 | 62107021 | G | A | 0.777 | -0.024 | 0.002 | 1.74e-25 |
| rs6664626 | 1 | 66128239 | T | G | 0.143 | 0.019 | 0.003 | 4.38e-11 |
| rs41313381 | 1 | 79411968 | A | C | 0.030 | 0.043 | 0.005 | 8.85e-16 |
| rs7532966 | 1 | 89955188 | C | T | 0.499 | 0.017 | 0.002 | 1.6e-19 |
| rs12144117 | 1 | 9182141 | T | C | 0.215 | -0.014 | 0.002 | 7.63e-10 |
| rs114425738 | 1 | 92171673 | C | G | 0.013 | 0.268 | 0.008 | 1e-200 |
| rs115340020 | 1 | 92766438 | A | G | 0.027 | -0.094 | 0.006 | 2.22e-59 |
| rs7524046 | 1 | 9307033 | A | G | 0.251 | 0.015 | 0.002 | 1.11e-11 |
| rs2433279 | 1 | 93907909 | G | T | 0.879 | 0.023 | 0.003 | 2.08e-15 |
| rs80036648 | 1 | 93912771 | G | A | 0.017 | 0.155 | 0.008 | 1.16e-92 |
| rs7516138 | 1 | 9711642 | G | A | 0.391 | -0.023 | 0.002 | 1.31e-31 |
| rs34600126 | 20 | 1597433 | T | G | 0.218 | 0.022 | 0.002 | 6.03e-23 |
| rs4813619 | 20 | 2815715 | T | G | 0.511 | 0.017 | 0.002 | 4.24e-18 |
| rs12480462 | 20 | 31065178 | T | C | 0.316 | -0.031 | 0.002 | 1.44e-52 |
| rs12480732 | 20 | 31191015 | T | C | 0.258 | 0.054 | 0.002 | 8.58e-136 |
| rs628977 | 20 | 3649721 | C | T | 0.631 | -0.013 | 0.002 | 1.21e-10 |
| rs4142441 | 20 | 42839620 | G | A | 0.152 | 0.031 | 0.003 | 1.22e-31 |
| rs34414028 | 20 | 47212868 | T | A | 0.209 | 0.014 | 0.002 | 2.47e-09 |
| rs932905 | 20 | 48796131 | A | G | 0.282 | 0.021 | 0.002 | 6.11e-24 |
| rs17196752 | 20 | 48887268 | T | C | 0.190 | -0.072 | 0.002 | 8.1e-199 |
| rs6512627 | 20 | 48967591 | A | G | 0.194 | -0.047 | 0.002 | 1.83e-88 |
| rs3787513 | 20 | 61573736 | A | G | 0.207 | -0.015 | 0.002 | 6.32e-11 |
| rs4432538 | 20 | 8607393 | A | G | 0.509 | -0.029 | 0.002 | 4.75e-54 |
| rs1997577 | 21 | 16371102 | T | A | 0.155 | -0.030 | 0.003 | 5.8e-30 |
| rs57221391 | 21 | 16583936 | G | T | 0.071 | -0.022 | 0.004 | 1.51e-09 |
| rs35068491 | 21 | 36238307 | C | T | 0.209 | 0.015 | 0.002 | 5.68e-10 |
| rs2834670 | 21 | 36280376 | G | A | 0.176 | 0.027 | 0.003 | 1.11e-19 |
| rs73203055 | 21 | 36407530 | G | C | 0.072 | -0.026 | 0.004 | 1.27e-12 |
| rs192498589 | 22 | 17516047 | G | A | 0.017 | 0.070 | 0.008 | 8.24e-18 |
| rs41409548 | 22 | 17579495 | A | G | 0.034 | -0.096 | 0.006 | 1.49e-64 |
| rs9606614 | 22 | 17579956 | T | C | 0.033 | 0.034 | 0.005 | 1.19e-10 |
| rs2159068 | 22 | 17653955 | G | C | 0.798 | 0.017 | 0.002 | 3.28e-13 |
| rs2019180 | 22 | 17712142 | A | G | 0.093 | -0.037 | 0.003 | 2.57e-27 |
| rs35477714 | 22 | 18202530 | C | G | 0.505 | 0.013 | 0.002 | 1.39e-11 |
| rs34505104 | 22 | 24624609 | G | A | 0.305 | -0.033 | 0.002 | 6.38e-57 |
| rs9625746 | 22 | 29637658 | C | G | 0.413 | -0.021 | 0.002 | 7.42e-28 |
| rs62236123 | 22 | 31474019 | C | G | 0.042 | -0.028 | 0.005 | 4.75e-09 |
| rs139271 | 22 | 39487595 | C | T | 0.442 | -0.015 | 0.002 | 1.11e-15 |
| rs47341 | 22 | 43560763 | T | C | 0.396 | 0.031 | 0.002 | 1.46e-54 |
| rs739241 | 22 | 50052921 | A | G | 0.607 | 0.018 | 0.002 | 1.72e-20 |
| rs140763648 | 22 | 50767351 | G | A | 0.412 | -0.012 | 0.002 | 4.34e-09 |
| rs6745920 | 2 | 102145661 | C | T | 0.619 | 0.013 | 0.002 | 2.02e-10 |
| rs150449635 | 2 | 111752151 | C | T | 0.022 | 0.140 | 0.007 | 4.24e-97 |
| rs72836307 | 2 | 111776154 | T | C | 0.156 | -0.041 | 0.003 | 1.37e-56 |
| rs3789062 | 2 | 111917317 | T | C | 0.245 | -0.020 | 0.002 | 3.22e-20 |
| rs4669869 | 2 | 12898460 | C | T | 0.445 | 0.020 | 0.002 | 3.76e-27 |
| rs7574456 | 2 | 136890059 | T | C | 0.740 | -0.040 | 0.002 | 5.2e-80 |
| rs7608128 | 2 | 137007943 | T | G | 0.181 | -0.019 | 0.002 | 1.39e-15 |
| rs3856364 | 2 | 145477217 | G | C | 0.680 | 0.013 | 0.002 | 3.45e-10 |
| rs13032786 | 2 | 148803672 | G | C | 0.300 | 0.020 | 0.002 | 2.49e-23 |
| rs1863219 | 2 | 160418171 | G | C | 0.512 | 0.018 | 0.002 | 5.95e-23 |
| rs13032491 | 2 | 161080233 | T | C | 0.139 | 0.016 | 0.003 | 4.72e-09 |
| rs6733162 | 2 | 162913498 | C | G | 0.624 | 0.012 | 0.002 | 1.4e-09 |
| rs3931 | 2 | 169721381 | G | A | 0.320 | 0.023 | 0.002 | 2.02e-30 |
| rs8207 | 2 | 170493863 | G | A | 0.267 | 0.014 | 0.002 | 1.29e-10 |
| rs11680095 | 2 | 181825956 | T | C | 0.592 | 0.022 | 0.002 | 1.35e-31 |
| rs78655702 | 2 | 182303729 | T | C | 0.050 | -0.059 | 0.004 | 3.58e-43 |
| rs155127 | 2 | 182305701 | C | G | 0.348 | 0.060 | 0.002 | 1e-200 |
| rs155109 | 2 | 182352679 | G | A | 0.321 | -0.021 | 0.002 | 7.19e-26 |
| rs17860428 | 2 | 202151400 | A | G | 0.180 | -0.016 | 0.002 | 1.39e-10 |
| rs6736362 | 2 | 219115108 | T | C | 0.559 | 0.022 | 0.002 | 6.45e-32 |
| rs6436124 | 2 | 220041433 | A | C | 0.608 | 0.015 | 0.002 | 8.1e-15 |
| rs10197805 | 2 | 225750753 | C | T | 0.157 | -0.042 | 0.003 | 4.7e-59 |
| rs4470337 | 2 | 228293381 | G | A | 0.596 | -0.012 | 0.002 | 4.33e-10 |
| rs13002735 | 2 | 232268884 | C | A | 0.241 | -0.014 | 0.002 | 3.3e-10 |
| rs30102 | 2 | 237817538 | C | G | 0.317 | -0.013 | 0.002 | 9.89e-11 |
| rs34236350 | 2 | 241568326 | T | C | 0.183 | -0.032 | 0.003 | 8.44e-38 |
| rs2950835 | 2 | 27750545 | G | A | 0.557 | 0.013 | 0.002 | 7.45e-12 |
| rs649729 | 2 | 31464385 | A | T | 0.698 | 0.021 | 0.002 | 5.04e-24 |
| rs28498283 | 2 | 43360065 | T | A | 0.258 | 0.021 | 0.002 | 2.41e-22 |
| rs6706095 | 2 | 46075677 | G | T | 0.760 | 0.019 | 0.002 | 9.2e-19 |
| rs75475627 | 2 | 54787592 | G | C | 0.077 | 0.024 | 0.004 | 1.8e-11 |
| rs1562309 | 2 | 61770126 | A | C | 0.615 | -0.015 | 0.002 | 1.04e-15 |
| rs1968179 | 2 | 64929212 | T | C | 0.243 | 0.018 | 0.002 | 3.81e-17 |
| rs13385171 | 2 | 65661843 | C | T | 0.583 | 0.028 | 0.002 | 5.14e-49 |
| rs3111414 | 2 | 8443859 | G | C | 0.794 | 0.017 | 0.002 | 5.62e-14 |
| rs7572278 | 2 | 8563029 | A | T | 0.207 | 0.022 | 0.002 | 3.44e-20 |
| rs4907230 | 2 | 96855241 | A | G | 0.324 | 0.016 | 0.002 | 7.24e-15 |
| rs62261974 | 3 | 107296969 | G | A | 0.183 | -0.017 | 0.002 | 7.01e-12 |
| rs1822534 | 3 | 12266804 | G | A | 0.394 | -0.031 | 0.002 | 4.69e-59 |
| rs62263808 | 3 | 123086587 | A | G | 0.217 | -0.022 | 0.002 | 2.37e-22 |
| rs2291079 | 3 | 124483089 | G | C | 0.219 | -0.015 | 0.002 | 1.97e-11 |
| rs11713343 | 3 | 128185399 | A | G | 0.197 | -0.026 | 0.002 | 5.24e-28 |
| rs2245626 | 3 | 128287687 | G | A | 0.676 | 0.075 | 0.002 | 1e-200 |
| rs2669833 | 3 | 129604169 | A | G | 0.160 | -0.019 | 0.003 | 2.54e-13 |
| rs7434143 | 3 | 136600667 | T | G | 0.680 | 0.017 | 0.002 | 6.49e-17 |
| rs6800122 | 3 | 141249398 | T | C | 0.400 | -0.022 | 0.002 | 4.75e-30 |
| rs10804681 | 3 | 141660675 | T | A | 0.843 | -0.018 | 0.003 | 9.56e-13 |
| rs1472121 | 3 | 151036229 | T | C | 0.776 | 0.014 | 0.002 | 3.65e-10 |
| rs16831132 | 3 | 159923303 | T | C | 0.397 | -0.014 | 0.002 | 2.07e-12 |
| rs7633965 | 3 | 168864643 | C | A | 0.929 | 0.025 | 0.004 | 8.35e-12 |
| rs231988 | 3 | 172276979 | C | T | 0.868 | 0.026 | 0.003 | 4.8e-19 |
| rs2018092 | 3 | 185903842 | C | T | 0.615 | -0.012 | 0.002 | 1.97e-09 |
| rs56925909 | 3 | 187429202 | G | C | 0.219 | 0.014 | 0.002 | 1.33e-09 |
| rs9815073 | 3 | 188115682 | A | C | 0.346 | -0.022 | 0.002 | 4.16e-26 |
| rs7626444 | 3 | 196504902 | C | G | 0.421 | -0.029 | 0.002 | 5.72e-52 |
| rs869785 | 3 | 24347800 | C | T | 0.670 | -0.017 | 0.002 | 3.97e-18 |
| rs3732378 | 3 | 39307162 | A | G | 0.173 | 0.055 | 0.002 | 1.07e-108 |
| rs4683345 | 3 | 42886642 | A | G | 0.385 | -0.058 | 0.002 | 1e-200 |
| rs73083231 | 3 | 43172180 | G | A | 0.045 | 0.029 | 0.005 | 1.04e-10 |
| rs28677778 | 3 | 46322171 | G | A | 0.067 | -0.065 | 0.004 | 5.53e-68 |
| rs112313229 | 3 | 46364860 | A | G | 0.045 | 0.057 | 0.005 | 6.64e-36 |
| rs7429588 | 3 | 46835250 | C | T | 0.293 | 0.015 | 0.002 | 7.84e-11 |
| rs11130612 | 3 | 58029016 | T | C | 0.334 | -0.012 | 0.002 | 4.71e-10 |
| rs13079034 | 3 | 71813219 | T | C | 0.113 | -0.021 | 0.003 | 1.05e-12 |
| rs9809116 | 3 | 72397279 | G | A | 0.409 | 0.017 | 0.002 | 8.21e-20 |
| rs10935473 | 3 | 98416900 | T | G | 0.442 | 0.025 | 0.002 | 2.3e-40 |
| rs141936164 | 4 | 103401723 | G | A | 0.351 | -0.019 | 0.002 | 2.29e-21 |
| rs144317085 | 4 | 105806108 | T | A | 0.034 | 0.062 | 0.005 | 3.1e-32 |
| rs6533483 | 4 | 110887989 | A | G | 0.403 | -0.018 | 0.002 | 8.82e-22 |
| rs7692994 | 4 | 120427489 | A | C | 0.330 | -0.014 | 0.002 | 5.59e-12 |
| rs4240356 | 4 | 145042250 | G | C | 0.550 | -0.024 | 0.002 | 1.94e-35 |
| rs7687559 | 4 | 154385925 | C | G | 0.398 | 0.014 | 0.002 | 1.58e-12 |
| rs113862443 | 4 | 157725371 | G | C | 0.085 | -0.025 | 0.003 | 7.94e-14 |
| rs56058420 | 4 | 185237116 | G | A | 0.356 | 0.012 | 0.002 | 2.96e-09 |
| rs2269486 | 4 | 2267566 | T | A | 0.164 | 0.021 | 0.003 | 8.55e-16 |
| rs73809166 | 4 | 36305937 | C | T | 0.023 | 0.046 | 0.006 | 3.78e-13 |
| rs2711981 | 4 | 39039258 | T | C | 0.663 | -0.022 | 0.002 | 1.59e-27 |
| rs723585 | 4 | 55503194 | G | A | 0.483 | -0.017 | 0.002 | 9.43e-19 |
| rs9637714 | 4 | 57731446 | C | T | 0.289 | -0.016 | 0.002 | 5.42e-15 |
| rs871134 | 4 | 7044380 | T | C | 0.571 | 0.038 | 0.002 | 7.06e-89 |
| rs11723621 | 4 | 72615362 | G | A | 0.291 | -0.013 | 0.002 | 1.48e-10 |
| rs113172748 | 4 | 79639615 | G | A | 0.671 | 0.016 | 0.002 | 4.59e-15 |
| rs17005891 | 4 | 83547862 | A | G | 0.184 | -0.051 | 0.002 | 1.15e-98 |
| rs4566648 | 4 | 84159219 | G | T | 0.342 | -0.025 | 0.002 | 8.14e-38 |
| rs2548257 | 5 | 100162317 | A | C | 0.653 | 0.013 | 0.002 | 5.35e-11 |
| rs10478058 | 5 | 110962584 | G | A | 0.197 | 0.015 | 0.002 | 3.01e-10 |
| rs6869021 | 5 | 111039017 | T | C | 0.313 | -0.017 | 0.002 | 5.47e-17 |
| rs11242109 | 5 | 131677047 | T | G | 0.478 | -0.022 | 0.002 | 4.75e-33 |
| rs12332674 | 5 | 132322852 | G | T | 0.152 | -0.017 | 0.003 | 3.33e-10 |
| rs329125 | 5 | 133871101 | A | C | 0.207 | -0.017 | 0.002 | 1.38e-12 |
| rs6579771 | 5 | 149482262 | T | C | 0.269 | 0.034 | 0.002 | 6.13e-57 |
| rs17656204 | 5 | 149501803 | T | C | 0.322 | -0.054 | 0.002 | 6.78e-156 |
| rs360017 | 5 | 173207353 | G | A | 0.775 | -0.024 | 0.002 | 1.62e-27 |
| rs964752 | 5 | 17399899 | G | C | 0.540 | -0.012 | 0.002 | 9.61e-11 |
| rs6883116 | 5 | 179230321 | C | T | 0.417 | -0.018 | 0.002 | 8.21e-21 |
| rs116619972 | 5 | 32214314 | A | G | 0.026 | -0.040 | 0.006 | 6.73e-12 |
| rs445611 | 5 | 35396119 | G | A | 0.845 | 0.016 | 0.003 | 8.58e-10 |
| rs4865956 | 5 | 54882505 | A | T | 0.696 | -0.019 | 0.002 | 1.97e-20 |
| rs28722705 | 5 | 55453942 | T | A | 0.149 | 0.034 | 0.003 | 1e-38 |
| rs74735005 | 5 | 57202727 | C | T | 0.206 | -0.017 | 0.002 | 3.14e-13 |
| rs2338021 | 5 | 71749094 | C | T | 0.880 | -0.032 | 0.003 | 3.64e-28 |
| rs31243 | 5 | 75594360 | G | A | 0.096 | -0.023 | 0.003 | 1.35e-12 |
| rs244760 | 5 | 88111209 | G | C | 0.764 | -0.014 | 0.002 | 1.14e-10 |
| rs707794 | 6 | 10519890 | A | C | 0.532 | -0.030 | 0.002 | 7.79e-59 |
| rs9480737 | 6 | 107442277 | G | A | 0.321 | -0.022 | 0.002 | 2.74e-27 |
| rs1970364 | 6 | 113969527 | T | C | 0.265 | 0.019 | 0.002 | 1.32e-19 |
| rs9375150 | 6 | 122887555 | G | T | 0.447 | -0.018 | 0.002 | 1.84e-22 |
| rs761841 | 6 | 131114386 | C | T | 0.577 | 0.012 | 0.002 | 1.31e-10 |
| rs12055642 | 6 | 135079040 | C | T | 0.199 | 0.018 | 0.002 | 1.04e-14 |
| rs7776054 | 6 | 135418916 | G | A | 0.261 | -0.038 | 0.002 | 6.09e-71 |
| rs55666429 | 6 | 137113137 | A | G | 0.588 | -0.016 | 0.002 | 1.96e-16 |
| rs2797681 | 6 | 137593036 | C | G | 0.256 | -0.015 | 0.002 | 1.14e-10 |
| rs149110519 | 6 | 144385777 | T | C | 0.036 | 0.075 | 0.005 | 1.51e-48 |
| rs9390460 | 6 | 147694334 | C | T | 0.538 | 0.012 | 0.002 | 3.01e-10 |
| rs11155787 | 6 | 151686905 | T | C | 0.638 | 0.014 | 0.002 | 3.92e-12 |
| rs2817441 | 6 | 156938699 | T | C | 0.279 | -0.022 | 0.002 | 4.67e-25 |
| rs73784287 | 6 | 160980401 | G | A | 0.149 | 0.015 | 0.003 | 4.93e-09 |
| rs707845 | 6 | 16711855 | G | A | 0.511 | -0.017 | 0.002 | 6.09e-19 |
| rs3012415 | 6 | 170489464 | T | C | 0.823 | 0.020 | 0.003 | 1.37e-15 |
| rs2273215 | 6 | 170586082 | A | G | 0.458 | -0.018 | 0.002 | 2.72e-20 |
| rs6940465 | 6 | 21207748 | A | G | 0.151 | 0.016 | 0.003 | 1.61e-09 |
| rs78912080 | 6 | 25616453 | G | A | 0.075 | 0.026 | 0.004 | 5.55e-13 |
| rs12530071 | 6 | 2886067 | C | T | 0.225 | -0.015 | 0.002 | 5.78e-11 |
| rs2853999 | 6 | 31326074 | T | A | 0.138 | -0.076 | 0.003 | 2.93e-171 |
| rs17207748 | 6 | 31863168 | C | T | 0.047 | -0.035 | 0.004 | 2.3e-15 |
| rs56007794 | 6 | 41990827 | T | A | 0.244 | 0.039 | 0.002 | 4.77e-72 |
| rs11963621 | 6 | 44591006 | C | T | 0.101 | -0.048 | 0.003 | 6.15e-54 |
| rs115202835 | 6 | 45533239 | A | G | 0.068 | 0.027 | 0.004 | 4.52e-13 |
| rs10948314 | 6 | 46911131 | G | A | 0.103 | -0.034 | 0.003 | 1.04e-27 |
| rs644492 | 6 | 53444319 | G | A | 0.196 | -0.020 | 0.002 | 1.84e-17 |
| rs3761986 | 6 | 6680510 | C | T | 0.394 | -0.012 | 0.002 | 4.88e-10 |
| rs9379077 | 6 | 7167170 | G | A | 0.201 | 0.029 | 0.002 | 1.46e-34 |
| rs915125 | 6 | 82463376 | T | C | 0.282 | 0.034 | 0.002 | 4.34e-59 |
| rs62470670 | 7 | 114651694 | G | A | 0.092 | -0.023 | 0.003 | 5.3e-13 |
| rs7803075 | 7 | 130742066 | G | A | 0.735 | -0.014 | 0.002 | 2.28e-11 |
| rs7785014 | 7 | 137881690 | C | T | 0.671 | -0.025 | 0.002 | 2.04e-36 |
| rs7787179 | 7 | 143081833 | G | A | 0.207 | 0.021 | 0.002 | 3.31e-19 |
| rs62491927 | 7 | 149172894 | A | C | 0.257 | -0.013 | 0.002 | 1.42e-09 |
| rs17700436 | 7 | 17167825 | T | C | 0.057 | -0.028 | 0.004 | 1.53e-12 |
| rs34947605 | 7 | 2581594 | A | G | 0.172 | 0.015 | 0.002 | 3.39e-09 |
| rs62454420 | 7 | 27191804 | G | A | 0.070 | -0.035 | 0.004 | 2.39e-21 |
| rs798563 | 7 | 2757867 | C | A | 0.297 | 0.018 | 0.002 | 3.23e-17 |
| rs17156536 | 7 | 28279488 | A | C | 0.159 | 0.025 | 0.003 | 8.66e-22 |
| rs10238435 | 7 | 28724491 | T | C | 0.240 | -0.024 | 0.002 | 4.23e-25 |
| rs112248289 | 7 | 44770213 | C | A | 0.129 | 0.022 | 0.003 | 4.83e-15 |
| rs2136769 | 7 | 47469152 | C | A | 0.449 | 0.011 | 0.002 | 2.34e-09 |
| rs4385425 | 7 | 50307334 | G | A | 0.324 | -0.060 | 0.002 | 1.28e-198 |
| rs149007767 | 7 | 50370254 | T | C | 0.162 | -0.072 | 0.003 | 1.97e-163 |
| rs11766800 | 7 | 50435617 | T | A | 0.322 | 0.019 | 0.002 | 2.26e-21 |
| rs41430449 | 7 | 50798525 | G | C | 0.066 | -0.030 | 0.004 | 1.82e-15 |
| rs6796 | 7 | 6502367 | C | T | 0.274 | 0.046 | 0.002 | 1.66e-102 |
| rs7786376 | 7 | 73042614 | G | A | 0.276 | 0.012 | 0.002 | 4.73e-09 |
| rs178405 | 7 | 73513045 | T | G | 0.784 | -0.015 | 0.002 | 1.6e-10 |
| rs445 | 7 | 92408370 | T | C | 0.098 | -0.094 | 0.003 | 3.59e-196 |
| rs6465661 | 7 | 97885213 | T | C | 0.492 | -0.016 | 0.002 | 5.49e-18 |
| rs7792525 | 7 | 99972122 | G | A | 0.186 | 0.019 | 0.002 | 4.21e-15 |
| rs72673751 | 8 | 106578940 | C | T | 0.190 | -0.017 | 0.002 | 4.35e-12 |
| rs1954735 | 8 | 108239325 | C | T | 0.168 | 0.021 | 0.003 | 1.01e-16 |
| rs13271228 | 8 | 116597409 | G | T | 0.567 | -0.031 | 0.002 | 4.35e-59 |
| rs12334935 | 8 | 126617990 | A | G | 0.471 | -0.015 | 0.002 | 2.75e-15 |
| rs7843207 | 8 | 130570063 | A | C | 0.506 | -0.069 | 0.002 | 1e-200 |
| rs10101101 | 8 | 130691767 | A | G | 0.139 | -0.041 | 0.003 | 1.44e-50 |
| rs4397371 | 8 | 130701417 | T | C | 0.734 | -0.018 | 0.002 | 1.17e-17 |
| rs837227 | 8 | 130964654 | T | C | 0.745 | -0.015 | 0.002 | 1.11e-11 |
| rs10099546 | 8 | 144638734 | G | A | 0.143 | -0.020 | 0.003 | 4.33e-13 |
| rs4871844 | 8 | 22879734 | C | T | 0.343 | 0.016 | 0.002 | 1.27e-14 |
| rs62501136 | 8 | 23089429 | A | G | 0.221 | 0.020 | 0.002 | 1.72e-19 |
| rs62502392 | 8 | 27190945 | A | G | 0.143 | -0.018 | 0.003 | 2.08e-11 |
| rs4737009 | 8 | 41630405 | A | G | 0.236 | 0.015 | 0.002 | 3.44e-12 |
| rs113155021 | 8 | 43719739 | A | G | 0.015 | -0.048 | 0.008 | 2.21e-09 |
| rs145718079 | 8 | 48641192 | A | G | 0.010 | -0.080 | 0.010 | 4.68e-16 |
| rs45577137 | 8 | 48651633 | G | A | 0.044 | -0.065 | 0.005 | 9.05e-39 |
| rs113015223 | 8 | 57037166 | T | C | 0.211 | 0.019 | 0.002 | 1.06e-15 |
| rs3808609 | 8 | 59465377 | C | G | 0.318 | 0.017 | 0.002 | 3.86e-17 |
| rs7824937 | 8 | 61392724 | G | A | 0.369 | 0.018 | 0.002 | 1.03e-19 |
| rs7826487 | 8 | 6880925 | G | A | 0.116 | -0.041 | 0.003 | 2.17e-43 |
| rs12542907 | 8 | 68813188 | G | C | 0.399 | -0.024 | 0.002 | 1.01e-36 |
| rs13267464 | 8 | 6906925 | T | C | 0.399 | -0.020 | 0.002 | 8.9e-27 |
| rs16939607 | 8 | 79013333 | A | G | 0.146 | -0.025 | 0.003 | 2.43e-21 |
| rs1863651 | 8 | 82066998 | T | A | 0.699 | -0.017 | 0.002 | 4.32e-16 |
| rs13292863 | 9 | 113316712 | G | C | 0.155 | -0.016 | 0.003 | 5.77e-10 |
| rs1008158 | 9 | 113828811 | G | A | 0.343 | -0.067 | 0.002 | 1e-200 |
| rs10817147 | 9 | 113864075 | T | C | 0.377 | -0.059 | 0.002 | 1e-200 |
| rs77155602 | 9 | 114153208 | T | C | 0.180 | 0.029 | 0.002 | 3.25e-33 |
| rs13293713 | 9 | 114353573 | A | G | 0.081 | 0.052 | 0.004 | 1.83e-50 |
| rs72759267 | 9 | 126971204 | T | C | 0.159 | 0.021 | 0.003 | 5.36e-16 |
| rs2491104 | 9 | 130313823 | A | T | 0.242 | -0.014 | 0.002 | 3.17e-10 |
| rs79086185 | 9 | 130666538 | C | A | 0.039 | 0.031 | 0.005 | 1.17e-09 |
| rs11792030 | 9 | 130732588 | A | G | 0.104 | 0.022 | 0.003 | 1e-12 |
| rs676996 | 9 | 136146077 | G | T | 0.320 | -0.044 | 0.002 | 5.13e-106 |
| rs3731211 | 9 | 21986847 | A | T | 0.720 | 0.028 | 0.002 | 2.51e-42 |
| rs12376511 | 9 | 22142756 | C | T | 0.164 | -0.049 | 0.003 | 3.44e-82 |
| rs34881325 | 9 | 2622134 | T | C | 0.379 | 0.010 | 0.002 | 8.15e-11 |
| rs11557154 | 9 | 34107505 | T | C | 0.128 | 0.032 | 0.003 | 2.64e-30 |
| rs10814193 | 9 | 35018686 | C | A | 0.716 | 0.015 | 0.002 | 1.07e-12 |
| rs2236288 | 9 | 35749845 | C | G | 0.229 | 0.013 | 0.002 | 1.78e-09 |
| rs10758481 | 9 | 38195756 | C | T | 0.502 | -0.014 | 0.002 | 4.63e-14 |
| rs10780209 | 9 | 91472127 | A | G | 0.479 | 0.045 | 0.002 | 5.69e-131 |
| rs9410425 | 9 | 91562311 | A | G | 0.325 | -0.030 | 0.002 | 3.23e-51 |
| rs290243 | 9 | 93571337 | A | G | 0.215 | 0.014 | 0.002 | 3e-10 |
| rs34746889 | 16 | 67221444 | G | C | 0.983 | 0.058 | 0.007 | 2e-15 |
| rs75726056 | 1 | 150859753 | A | G | 0.988 | -0.072 | 0.008 | 2.11e-17 |
| rs115240699 | 6 | 33743079 | A | G | 0.993 | -0.067 | 0.011 | 2.97e-09 |
| rs10909370 | 10 | 115847890 | T | G | 0.464 | -0.012 | 0.002 | 4.76e-09 |
| rs2343551 | 10 | 82267945 | C | A | 0.795 | 0.021 | 0.002 | 1.29e-18 |
| rs2651780 | 11 | 2345261 | A | G | 0.850 | 0.022 | 0.003 | 5.51e-15 |
| rs11227312 | 11 | 65586597 | G | C | 0.485 | 0.012 | 0.002 | 3.87e-10 |
| rs113681054 | 12 | 21402979 | C | T | 0.161 | -0.016 | 0.003 | 3.31e-10 |
| rs2535403 | 12 | 718885 | C | T | 0.557 | -0.016 | 0.002 | 7.7e-16 |
| rs374039502 | 13 | 108960385 | A | T | 0.022 | -0.116 | 0.007 | 4.16e-61 |
| rs113235072 | 14 | 35344040 | C | G | 0.071 | 0.036 | 0.004 | 2.06e-21 |
| rs138841053 | 14 | 59985478 | C | T | 0.406 | 0.012 | 0.002 | 4.54e-10 |
| rs2300598 | 14 | 75359229 | C | T | 0.313 | 0.017 | 0.002 | 1.29e-16 |
| rs8037798 | 15 | 75240030 | G | T | 0.231 | 0.022 | 0.002 | 5.84e-22 |
| rs2562750 | 15 | 80250606 | A | T | 0.759 | 0.070 | 0.002 | 1e-200 |
| rs10871483 | 16 | 29592725 | A | T | 0.521 | -0.015 | 0.002 | 1.49e-11 |
| rs8054439 | 16 | 4146007 | C | G | 0.117 | 0.024 | 0.003 | 1.78e-15 |
| rs12924604 | 16 | 74601486 | A | G | 0.198 | -0.017 | 0.002 | 8.24e-13 |
| rs181791 | 16 | 84558889 | T | C | 0.164 | 0.026 | 0.003 | 2.94e-18 |
| rs7223589 | 17 | 29728284 | C | T | 0.616 | 0.014 | 0.002 | 1.93e-12 |
| rs113232639 | 18 | 20715656 | A | G | 0.489 | -0.018 | 0.002 | 5.66e-19 |
| rs12151289 | 19 | 33751852 | C | G | 0.031 | -0.085 | 0.006 | 1.13e-46 |
| rs62113155 | 19 | 36752859 | T | G | 0.201 | -0.019 | 0.002 | 1.33e-14 |
| rs12972849 | 19 | 54338067 | A | G | 0.652 | 0.051 | 0.002 | 7.41e-95 |
| rs61242663 | 19 | 836059 | T | C | 0.260 | 0.026 | 0.002 | 5.69e-27 |
| rs28549287 | 1 | 110230138 | A | G | 0.761 | -0.017 | 0.003 | 1.17e-11 |
| rs7512283 | 1 | 161000113 | G | A | 0.795 | 0.017 | 0.002 | 2.51e-12 |
| rs61804163 | 1 | 161622861 | C | T | 0.296 | -0.020 | 0.002 | 3.69e-19 |
| rs7542018 | 1 | 243972240 | A | G | 0.887 | 0.022 | 0.003 | 1.48e-12 |
| rs7529978 | 1 | 53204705 | A | G | 0.470 | -0.017 | 0.002 | 5.28e-19 |
| rs2815358 | 1 | 67493778 | C | T | 0.650 | -0.018 | 0.002 | 9.15e-18 |
| rs192009275 | 20 | 30250942 | T | C | 0.257 | -0.021 | 0.002 | 1.08e-20 |
| rs6055629 | 20 | 8178309 | A | C | 0.485 | -0.012 | 0.002 | 7.12e-10 |
| rs180765893 | 2 | 182372755 | A | T | 0.017 | 0.048 | 0.008 | 1.48e-09 |
| rs185880215 | 2 | 43549887 | T | C | 0.069 | 0.023 | 0.004 | 2.72e-09 |
| rs4683611 | 3 | 141290903 | C | T | 0.203 | -0.016 | 0.003 | 4.7e-09 |
| rs549046531 | 4 | 145975794 | T | C | 0.272 | -0.019 | 0.002 | 4.49e-16 |
| rs565245327 | 6 | 31614543 | T | G | 0.471 | -0.045 | 0.003 | 8.78e-70 |
| rs28474755 | 6 | 32623919 | T | C | 0.121 | 0.042 | 0.003 | 2.23e-45 |
| rs6968441 | 7 | 107151877 | C | G | 0.467 | 0.012 | 0.002 | 2.8e-10 |
| rs6950899 | 7 | 65286649 | A | G | 0.370 | 0.015 | 0.002 | 3.74e-12 |
| rs201446427 | 7 | 6851361 | C | A | 0.197 | 0.019 | 0.003 | 1.86e-11 |
| rs71518544 | 8 | 11782349 | T | G | 0.404 | 0.016 | 0.002 | 1.09e-14 |
| rs2978891 | 8 | 6697258 | A | T | 0.505 | -0.020 | 0.002 | 1.51e-25 |
| rs7020171 | 9 | 91776952 | G | T | 0.248 | -0.014 | 0.002 | 9.16e-10 |
| rs549758081 | 1 | 93691855 | D | I | 0.433 | -0.016 | 0.002 | 2.22e-12 |

**Table S11. 393 SNPs significantly associated with lymphocyte count used IVs in forward MR analyses derived from Vuckovic D et al.** Chr: Chromosome; EA: Effect allele; NEA: Non-effect allele; EAF: Effect allele frequency.

| SNP | Chr | Pos | EA | NEA | EAF | Beta | SE | P-value |
| --- | --- | --- | --- | --- | --- | --- | --- | --- |
| rs159963 | 1 | 8504421 | A | C | 0.583 | -0.019 | 0.002 | 3.8e-19 |
| rs753612 | 1 | 19978895 | G | A | 0.611 | 0.014 | 0.002 | 4.9e-11 |
| rs7517330 | 1 | 23830340 | A | C | 0.152 | 0.020 | 0.003 | 9.1e-11 |
| rs12750971 | 1 | 24219590 | T | C | 0.758 | -0.022 | 0.003 | 2.4e-16 |
| rs9438860 | 1 | 25224957 | C | G | 0.410 | -0.016 | 0.002 | 3.4e-14 |
| rs11247712 | 1 | 28229681 | T | C | 0.369 | -0.020 | 0.002 | 8.8e-21 |
| rs77425607 | 1 | 39544876 | A | G | 0.333 | -0.014 | 0.002 | 1.5e-09 |
| rs4068540 | 1 | 40398941 | G | A | 0.267 | 0.017 | 0.002 | 1.8e-12 |
| rs5011302 | 1 | 56907563 | C | T | 0.544 | -0.020 | 0.002 | 2.2e-21 |
| rs3014983 | 1 | 65460691 | T | C | 0.761 | -0.019 | 0.003 | 6e-14 |
| rs6700896 | 1 | 66089782 | T | C | 0.378 | -0.018 | 0.002 | 1e-16 |
| rs2755253 | 1 | 67470843 | T | C | 0.707 | -0.032 | 0.002 | 6.5e-43 |
| rs2635119 | 1 | 79352340 | C | T | 0.688 | 0.017 | 0.002 | 6.5e-14 |
| rs12741113 | 1 | 101147149 | C | T | 0.447 | -0.018 | 0.002 | 1e-16 |
| rs6696259 | 1 | 101221482 | C | G | 0.410 | 0.035 | 0.002 | 3.5e-59 |
| rs377345022 | 1 | 101653213 | T | A | 0.265 | -0.019 | 0.002 | 1e-13 |
| rs10875371 | 1 | 101762529 | T | C | 0.420 | -0.022 | 0.002 | 2.4e-24 |
| rs2800883 | 1 | 112143035 | C | T | 0.378 | -0.014 | 0.002 | 1.2e-10 |
| rs2476601 | 1 | 114377568 | G | A | 0.899 | 0.058 | 0.003 | 4.4e-62 |
| rs4970926 | 1 | 150673684 | C | T | 0.513 | -0.027 | 0.002 | 3.1e-38 |
| rs2157691 | 1 | 158582838 | C | G | 0.266 | 0.032 | 0.002 | 3.3e-42 |
| rs12405457 | 1 | 160759052 | G | A | 0.215 | 0.026 | 0.003 | 7.1e-25 |
| rs11746 | 1 | 161683136 | A | G | 0.487 | 0.019 | 0.002 | 7.4e-20 |
| rs10918048 | 1 | 164558512 | T | A | 0.122 | -0.020 | 0.003 | 7.9e-10 |
| rs1689796 | 1 | 182151236 | G | T | 0.334 | 0.018 | 0.002 | 2.5e-15 |
| rs188626852 | 1 | 198498784 | A | C | 0.013 | 0.074 | 0.010 | 7.4e-14 |
| rs10494783 | 1 | 198663661 | A | G | 0.051 | -0.054 | 0.005 | 3.9e-29 |
| rs17612648 | 1 | 198665917 | G | C | 0.015 | 0.061 | 0.009 | 5.1e-11 |
| rs3861929 | 1 | 201025850 | T | C | 0.605 | -0.016 | 0.002 | 3.7e-14 |
| rs4531346 | 1 | 204269185 | A | G | 0.268 | -0.020 | 0.002 | 3.5e-16 |
| rs7548606 | 1 | 212388607 | G | A | 0.626 | 0.014 | 0.002 | 2.1e-10 |
| rs12745411 | 1 | 214405704 | T | G | 0.599 | 0.018 | 0.002 | 3.2e-16 |
| rs12021809 | 1 | 225941782 | A | G | 0.109 | -0.027 | 0.003 | 1.3e-15 |
| rs4659611 | 1 | 236033115 | T | A | 0.744 | -0.018 | 0.002 | 3.5e-13 |
| rs74227709 | 1 | 247722588 | A | G | 0.072 | -0.030 | 0.004 | 2.8e-13 |
| rs6721663 | 2 | 7615061 | A | G | 0.159 | -0.026 | 0.003 | 1.2e-19 |
| rs7572278 | 2 | 8563029 | A | T | 0.203 | 0.023 | 0.003 | 1.2e-18 |
| rs56243142 | 2 | 11333606 | C | T | 0.547 | 0.015 | 0.002 | 3.9e-13 |
| rs730126 | 2 | 12891476 | C | A | 0.414 | 0.015 | 0.002 | 1.2e-12 |
| rs72781679 | 2 | 24237712 | A | G | 0.135 | 0.061 | 0.003 | 3.1e-87 |
| rs4407213 | 2 | 25541636 | C | G | 0.032 | 0.045 | 0.006 | 2.2e-13 |
| rs1260326 | 2 | 27730940 | C | T | 0.605 | -0.024 | 0.002 | 5.8e-28 |
| rs113542380 | 2 | 43464818 | A | G | 0.075 | -0.070 | 0.004 | 2.6e-66 |
| rs805316 | 2 | 54133744 | C | T | 0.276 | -0.015 | 0.002 | 1.4e-10 |
| rs2920880 | 2 | 55297818 | T | C | 0.594 | 0.016 | 0.002 | 3.5e-13 |
| rs12991188 | 2 | 62554797 | G | A | 0.599 | 0.014 | 0.002 | 2.3e-10 |
| rs4852777 | 2 | 71534161 | C | G | 0.595 | 0.013 | 0.002 | 1.4e-09 |
| rs6755786 | 2 | 103048103 | T | C | 0.774 | 0.028 | 0.003 | 1.1e-27 |
| rs13019266 | 2 | 111604642 | C | T | 0.679 | 0.031 | 0.002 | 1e-41 |
| rs1976055 | 2 | 111787853 | A | C | 0.157 | -0.017 | 0.003 | 3.1e-09 |
| rs199898421 | 2 | 112193161 | G | A | 0.059 | 0.038 | 0.006 | 1.1e-09 |
| rs55706446 | 2 | 112239082 | T | C | 0.075 | -0.074 | 0.004 | 1.7e-66 |
| rs2241978 | 2 | 113954006 | G | A | 0.482 | 0.021 | 0.002 | 1.4e-23 |
| rs11688303 | 2 | 128410244 | T | C | 0.118 | 0.020 | 0.003 | 2e-09 |
| rs71420836 | 2 | 129060871 | T | C | 0.096 | 0.033 | 0.004 | 2.8e-20 |
| rs73961715 | 2 | 143803847 | C | T | 0.129 | -0.051 | 0.003 | 3.9e-59 |
| rs34030812 | 2 | 144248905 | C | T | 0.367 | -0.019 | 0.002 | 3.2e-18 |
| rs10173412 | 2 | 161330940 | C | T | 0.201 | -0.042 | 0.003 | 2.4e-56 |
| rs2075302 | 2 | 163076146 | C | T | 0.453 | -0.015 | 0.002 | 2.1e-12 |
| rs74401397 | 2 | 181944580 | G | C | 0.042 | -0.038 | 0.005 | 3.9e-13 |
| rs62180252 | 2 | 182175221 | C | T | 0.250 | 0.052 | 0.002 | 6.6e-101 |
| rs10179705 | 2 | 188315234 | A | G | 0.292 | -0.022 | 0.002 | 4.2e-21 |
| rs3769818 | 2 | 202151163 | G | A | 0.732 | 0.033 | 0.002 | 1.1e-43 |
| rs11888947 | 2 | 204642460 | A | T | 0.730 | -0.015 | 0.002 | 1.2e-10 |
| rs715 | 2 | 211543055 | C | T | 0.311 | -0.015 | 0.002 | 3.3e-11 |
| rs13018173 | 2 | 216248606 | C | T | 0.232 | -0.015 | 0.002 | 2.9e-09 |
| rs78058190 | 2 | 219699999 | A | G | 0.050 | 0.048 | 0.005 | 1e-18 |
| rs978917 | 2 | 223974443 | A | G | 0.616 | 0.017 | 0.002 | 2.1e-15 |
| rs11676298 | 2 | 227291731 | G | C | 0.192 | 0.026 | 0.003 | 6.5e-22 |
| rs4973318 | 2 | 231222675 | C | T | 0.106 | 0.022 | 0.003 | 9e-11 |
| rs58106596 | 2 | 232579379 | A | G | 0.222 | -0.024 | 0.003 | 2e-21 |
| rs13016839 | 2 | 234357513 | G | A | 0.106 | -0.026 | 0.003 | 6.8e-14 |
| rs9826367 | 3 | 12294202 | G | A | 0.449 | -0.035 | 0.002 | 3.3e-61 |
| rs4261897 | 3 | 18764304 | C | T | 0.296 | 0.021 | 0.002 | 8.9e-19 |
| rs13096529 | 3 | 24232035 | T | C | 0.191 | 0.020 | 0.003 | 1.5e-13 |
| rs2887944 | 3 | 27758275 | T | G | 0.435 | 0.024 | 0.002 | 2e-28 |
| rs75033378 | 3 | 28295653 | T | C | 0.175 | 0.037 | 0.003 | 4.2e-39 |
| rs4676482 | 3 | 39237190 | T | A | 0.190 | 0.016 | 0.003 | 3.8e-09 |
| rs11706384 | 3 | 39296881 | T | G | 0.236 | 0.026 | 0.002 | 4.4e-25 |
| rs13063578 | 3 | 47087837 | A | T | 0.401 | 0.054 | 0.002 | 2.6e-125 |
| rs7637449 | 3 | 56628031 | A | G | 0.529 | 0.015 | 0.002 | 3.4e-13 |
| rs12485738 | 3 | 56865776 | G | A | 0.629 | 0.017 | 0.002 | 5.5e-15 |
| rs35592432 | 3 | 71355240 | C | G | 0.027 | 0.107 | 0.007 | 1.6e-55 |
| rs6809697 | 3 | 71414864 | A | G | 0.135 | -0.020 | 0.003 | 2.5e-10 |
| rs6782672 | 3 | 108506049 | G | A | 0.659 | 0.016 | 0.002 | 8.4e-13 |
| rs59107033 | 3 | 123105643 | T | C | 0.225 | -0.025 | 0.003 | 8.6e-24 |
| rs332507 | 3 | 124435110 | C | T | 0.220 | -0.018 | 0.003 | 2e-12 |
| rs11712335 | 3 | 128210550 | C | T | 0.270 | 0.016 | 0.002 | 2.3e-11 |
| rs9834250 | 3 | 136561899 | G | C | 0.687 | 0.020 | 0.002 | 1.5e-18 |
| rs7625643 | 3 | 141150026 | G | A | 0.446 | 0.025 | 0.002 | 1.9e-31 |
| rs9863821 | 3 | 151007175 | T | C | 0.775 | 0.026 | 0.003 | 9.7e-24 |
| rs2647929 | 3 | 159693570 | C | T | 0.567 | 0.015 | 0.002 | 1e-11 |
| rs2291900 | 3 | 170825920 | A | G | 0.293 | 0.017 | 0.002 | 1.3e-12 |
| rs7627511 | 3 | 171178037 | C | G | 0.217 | -0.018 | 0.003 | 2.1e-11 |
| rs10937106 | 3 | 182738287 | A | G | 0.713 | -0.015 | 0.002 | 5.3e-10 |
| rs77265382 | 3 | 185546150 | T | C | 0.045 | -0.036 | 0.005 | 2.8e-12 |
| rs2089979 | 3 | 196501413 | G | A | 0.416 | -0.036 | 0.002 | 3.2e-61 |
| rs1344603 | 4 | 38109847 | C | T | 0.675 | 0.031 | 0.002 | 2.6e-42 |
| rs1386623 | 4 | 38372043 | T | G | 0.681 | 0.068 | 0.002 | 8.5e-198 |
| rs13105987 | 4 | 38602467 | A | G | 0.277 | 0.050 | 0.002 | 3.6e-98 |
| rs73142294 | 4 | 40301624 | T | C | 0.249 | 0.016 | 0.002 | 1.7e-10 |
| rs7437961 | 4 | 55066758 | C | T | 0.236 | 0.015 | 0.002 | 1.7e-09 |
| rs7665147 | 4 | 57767327 | A | T | 0.186 | -0.023 | 0.003 | 1.1e-17 |
| rs2298850 | 4 | 72614267 | C | G | 0.279 | -0.016 | 0.002 | 1.4e-11 |
| rs7662083 | 4 | 80933308 | A | G | 0.529 | -0.031 | 0.002 | 1.2e-48 |
| rs17005891 | 4 | 83547862 | A | G | 0.184 | -0.047 | 0.003 | 7.5e-66 |
| rs17703261 | 4 | 88058337 | T | A | 0.187 | 0.020 | 0.003 | 2.4e-13 |
| rs1431545 | 4 | 90220659 | T | C | 0.654 | 0.021 | 0.002 | 1.6e-20 |
| rs909349 | 4 | 103556316 | C | T | 0.521 | 0.039 | 0.002 | 6.4e-76 |
| rs62332762 | 4 | 106143492 | T | C | 0.399 | -0.027 | 0.002 | 2.4e-35 |
| rs2237040 | 4 | 110838862 | T | C | 0.404 | -0.019 | 0.002 | 2.1e-18 |
| rs72684793 | 4 | 123024769 | A | C | 0.300 | 0.015 | 0.002 | 1.7e-10 |
| rs309375 | 4 | 123681156 | A | C | 0.572 | 0.018 | 0.002 | 1.5e-16 |
| rs112737912 | 4 | 149366675 | T | G | 0.360 | -0.014 | 0.002 | 1.6e-10 |
| rs4696314 | 4 | 153005716 | C | T | 0.512 | 0.016 | 0.002 | 1.3e-13 |
| rs419470 | 5 | 16625591 | G | A | 0.716 | 0.016 | 0.002 | 2.8e-11 |
| rs6859682 | 5 | 34668797 | G | A | 0.331 | -0.014 | 0.002 | 2.1e-09 |
| rs11567699 | 5 | 35859011 | G | C | 0.260 | 0.056 | 0.002 | 2.8e-117 |
| rs2548499 | 5 | 52072544 | G | A | 0.208 | 0.020 | 0.003 | 3.7e-14 |
| rs2408014 | 5 | 54864017 | T | A | 0.681 | -0.016 | 0.002 | 7.9e-12 |
| rs151912 | 5 | 57607142 | T | A | 0.596 | -0.013 | 0.002 | 1.1e-09 |
| rs251398 | 5 | 67509801 | C | T | 0.451 | -0.017 | 0.002 | 2.5e-16 |
| rs678393 | 5 | 72418309 | G | T | 0.261 | -0.017 | 0.002 | 5.1e-12 |
| rs2544920 | 5 | 100141374 | T | A | 0.617 | 0.014 | 0.002 | 6.6e-11 |
| rs78270096 | 5 | 110517419 | A | G | 0.070 | -0.032 | 0.004 | 2.5e-13 |
| rs73239278 | 5 | 118695051 | A | G | 0.202 | -0.032 | 0.003 | 7.4e-34 |
| rs251390 | 5 | 127538346 | A | G | 0.749 | 0.021 | 0.002 | 4.7e-18 |
| rs58835386 | 5 | 131600559 | G | A | 0.567 | -0.023 | 0.002 | 4.1e-26 |
| rs75925066 | 5 | 134722833 | A | C | 0.023 | 0.087 | 0.007 | 1.5e-35 |
| rs60580948 | 5 | 148340270 | G | A | 0.139 | -0.020 | 0.003 | 5.4e-11 |
| rs3815725 | 5 | 150518532 | G | A | 0.519 | -0.016 | 0.002 | 5.4e-14 |
| rs17055818 | 5 | 157969475 | C | T | 0.254 | -0.032 | 0.002 | 5.5e-39 |
| rs34066608 | 5 | 158360881 | C | A | 0.098 | -0.035 | 0.004 | 2e-22 |
| rs6882776 | 5 | 172664163 | A | G | 0.283 | 0.014 | 0.002 | 3.5e-09 |
| rs360017 | 5 | 173207353 | G | A | 0.775 | -0.022 | 0.003 | 4.7e-18 |
| rs10063803 | 5 | 176667327 | A | C | 0.832 | 0.025 | 0.003 | 3.3e-19 |
| rs872071 | 6 | 411064 | G | A | 0.528 | 0.025 | 0.002 | 2.2e-32 |
| rs9392840 | 6 | 6901582 | T | C | 0.272 | 0.026 | 0.002 | 2.9e-26 |
| rs16870458 | 6 | 10545728 | A | G | 0.221 | -0.016 | 0.003 | 4.1e-10 |
| rs17119 | 6 | 14719496 | A | G | 0.804 | -0.018 | 0.003 | 3e-11 |
| rs52484 | 6 | 23564366 | G | A | 0.532 | -0.014 | 0.002 | 5.3e-11 |
| rs6927418 | 6 | 24827912 | T | C | 0.556 | -0.025 | 0.002 | 6.4e-32 |
| rs2523855 | 6 | 31022158 | C | G | 0.331 | 0.071 | 0.002 | 1e-200 |
| rs9265552 | 6 | 31298296 | G | C | 0.249 | -0.070 | 0.003 | 6.3e-142 |
| rs377181093 | 6 | 31422893 | G | A | 0.092 | 0.051 | 0.004 | 5.4e-39 |
| rs200801362 | 6 | 31555480 | C | T | 0.161 | -0.081 | 0.004 | 2.5e-94 |
| rs9272557 | 6 | 32607048 | C | T | 0.242 | -0.057 | 0.003 | 1.9e-111 |
| rs9942489 | 6 | 35323709 | A | T | 0.037 | -0.076 | 0.006 | 5.2e-42 |
| rs9381130 | 6 | 42040606 | A | G | 0.107 | 0.027 | 0.003 | 6.1e-15 |
| rs4711790 | 6 | 44573822 | T | C | 0.341 | -0.019 | 0.002 | 7.4e-18 |
| rs2294861 | 6 | 53136516 | T | C | 0.294 | 0.018 | 0.002 | 1.7e-14 |
| rs72928038 | 6 | 90976768 | A | G | 0.179 | -0.036 | 0.003 | 4e-38 |
| rs12526696 | 6 | 108053364 | A | G | 0.161 | 0.039 | 0.003 | 1.4e-41 |
| rs6929208 | 6 | 109720405 | C | G | 0.298 | -0.015 | 0.002 | 1.1e-10 |
| rs6926219 | 6 | 122720806 | A | G | 0.547 | 0.025 | 0.002 | 1.7e-32 |
| rs9375486 | 6 | 127388186 | T | C | 0.377 | -0.013 | 0.002 | 1.6e-09 |
| rs2327531 | 6 | 135048564 | G | A | 0.208 | 0.017 | 0.003 | 2.7e-11 |
| rs35786788 | 6 | 135419042 | A | G | 0.253 | -0.041 | 0.002 | 8.6e-62 |
| rs7763994 | 6 | 151912354 | T | A | 0.290 | 0.014 | 0.002 | 1.5e-09 |
| rs17710008 | 6 | 153043035 | A | G | 0.184 | 0.020 | 0.003 | 1.7e-13 |
| rs41267765 | 6 | 159462424 | T | C | 0.021 | -0.048 | 0.007 | 3.3e-11 |
| rs2273215 | 6 | 170586082 | A | G | 0.459 | -0.016 | 0.002 | 6e-14 |
| rs10480059 | 7 | 970035 | T | C | 0.518 | -0.016 | 0.002 | 1.4e-13 |
| rs4719654 | 7 | 2760008 | C | G | 0.442 | -0.015 | 0.002 | 1e-11 |
| rs852432 | 7 | 5562740 | G | A | 0.367 | 0.014 | 0.002 | 3.4e-10 |
| rs6796 | 7 | 6502367 | C | T | 0.278 | 0.028 | 0.002 | 2.3e-33 |
| rs35345753 | 7 | 22740513 | G | C | 0.209 | -0.027 | 0.003 | 5e-25 |
| rs4719922 | 7 | 28158058 | T | C | 0.201 | -0.025 | 0.003 | 8.4e-21 |
| rs2710804 | 7 | 36084529 | C | T | 0.377 | 0.015 | 0.002 | 1.4e-11 |
| rs73112675 | 7 | 37379030 | G | A | 0.180 | 0.019 | 0.003 | 5.9e-12 |
| rs3735485 | 7 | 45009341 | G | A | 0.845 | 0.045 | 0.003 | 2.8e-54 |
| rs150640087 | 7 | 50444152 | T | G | 0.016 | 0.061 | 0.009 | 1.1e-12 |
| rs10237524 | 7 | 56036024 | A | G | 0.330 | -0.019 | 0.002 | 6.4e-18 |
| rs4718976 | 7 | 70113409 | C | T | 0.695 | -0.016 | 0.002 | 5.2e-12 |
| rs2530482 | 7 | 74300890 | C | A | 0.285 | 0.016 | 0.002 | 1e-10 |
| rs7780424 | 7 | 77211703 | T | A | 0.609 | -0.014 | 0.002 | 4.9e-10 |
| rs10230506 | 7 | 92471518 | A | T | 0.160 | 0.019 | 0.003 | 3.1e-11 |
| rs11551890 | 7 | 97845713 | A | G | 0.489 | 0.013 | 0.002 | 7.8e-10 |
| rs7780328 | 7 | 99256844 | G | A | 0.067 | 0.040 | 0.004 | 1.2e-21 |
| rs9920 | 7 | 116200092 | C | T | 0.105 | 0.033 | 0.003 | 9.5e-22 |
| rs62621812 | 7 | 127015083 | A | G | 0.020 | 0.058 | 0.008 | 5e-14 |
| rs2727487 | 7 | 129513820 | C | A | 0.629 | -0.014 | 0.002 | 4.8e-10 |
| rs7793008 | 7 | 130747306 | G | A | 0.205 | 0.022 | 0.003 | 2.6e-17 |
| rs62485905 | 7 | 138741586 | T | C | 0.551 | -0.018 | 0.002 | 2e-17 |
| rs55700285 | 7 | 150209636 | T | G | 0.231 | 0.025 | 0.003 | 8.1e-23 |
| rs9886191 | 7 | 156656661 | T | C | 0.431 | -0.014 | 0.002 | 7.5e-11 |
| rs28588745 | 8 | 10647044 | T | A | 0.206 | 0.024 | 0.003 | 3.4e-20 |
| rs6586777 | 8 | 18702471 | A | C | 0.436 | -0.013 | 0.002 | 6.8e-10 |
| rs533703709 | 8 | 21736272 | T | A | 0.042 | -0.043 | 0.005 | 4.8e-16 |
| rs2313571 | 8 | 22789184 | T | C | 0.348 | 0.019 | 0.002 | 4.2e-17 |
| rs1567124 | 8 | 25244326 | A | C | 0.310 | 0.015 | 0.002 | 5.5e-11 |
| rs6474359 | 8 | 41549194 | C | T | 0.038 | -0.048 | 0.006 | 3.9e-18 |
| rs4737010 | 8 | 41630447 | A | G | 0.229 | 0.028 | 0.003 | 2.2e-28 |
| rs1976451 | 8 | 49456791 | G | T | 0.133 | -0.019 | 0.003 | 3.5e-09 |
| rs200523234 | 8 | 59501137 | C | A | 0.296 | 0.014 | 0.002 | 1.6e-09 |
| rs948421 | 8 | 61403965 | C | T | 0.376 | 0.014 | 0.002 | 4.7e-11 |
| rs6982175 | 8 | 61812553 | C | T | 0.879 | -0.023 | 0.003 | 5.1e-12 |
| rs894885 | 8 | 72908283 | T | G | 0.238 | -0.019 | 0.002 | 4.4e-14 |
| rs75653581 | 8 | 78363128 | T | C | 0.013 | 0.068 | 0.010 | 2.4e-12 |
| rs117934175 | 8 | 79069520 | A | G | 0.040 | -0.043 | 0.005 | 3.1e-15 |
| rs1441850 | 8 | 79657666 | C | T | 0.249 | 0.052 | 0.002 | 2.1e-99 |
| rs17424046 | 8 | 101372316 | T | C | 0.135 | 0.030 | 0.003 | 1.4e-21 |
| rs543427 | 8 | 102468599 | C | G | 0.491 | 0.016 | 0.002 | 1.9e-14 |
| rs6995717 | 8 | 106229410 | C | T | 0.449 | -0.013 | 0.002 | 4.8e-10 |
| rs72680380 | 8 | 119932741 | A | T | 0.466 | -0.013 | 0.002 | 3.2e-10 |
| rs2954025 | 8 | 126484463 | T | C | 0.649 | -0.015 | 0.002 | 5.2e-12 |
| rs10956401 | 8 | 129002419 | A | G | 0.343 | -0.025 | 0.002 | 9.3e-30 |
| rs837223 | 8 | 130971463 | T | C | 0.343 | -0.017 | 0.002 | 6.1e-14 |
| rs7007986 | 8 | 142217115 | A | G | 0.602 | -0.022 | 0.002 | 1.1e-24 |
| rs7033677 | 9 | 4049942 | G | A | 0.640 | -0.020 | 0.002 | 1.5e-18 |
| rs10815097 | 9 | 4858114 | G | A | 0.566 | -0.013 | 0.002 | 4.8e-10 |
| rs12336678 | 9 | 21785894 | T | C | 0.098 | -0.023 | 0.004 | 1.1e-10 |
| rs3731211 | 9 | 21986847 | A | T | 0.722 | 0.046 | 0.002 | 4.5e-86 |
| rs2065500 | 9 | 22145694 | G | A | 0.158 | -0.051 | 0.003 | 1.4e-69 |
| rs11541908 | 9 | 35705759 | A | G | 0.279 | 0.022 | 0.002 | 2.4e-21 |
| rs12343705 | 9 | 38197187 | T | A | 0.490 | -0.020 | 0.002 | 9.3e-22 |
| rs2807303 | 9 | 82187095 | A | G | 0.346 | 0.022 | 0.002 | 1.2e-22 |
| rs35010780 | 9 | 86499923 | A | G | 0.129 | -0.022 | 0.003 | 7.7e-12 |
| rs61750929 | 9 | 91495135 | T | C | 0.056 | -0.076 | 0.005 | 6e-60 |
| rs6477734 | 9 | 112728014 | C | T | 0.490 | -0.015 | 0.002 | 1e-12 |
| rs59364245 | 9 | 113897765 | C | T | 0.559 | -0.013 | 0.002 | 3.6e-09 |
| rs3761846 | 9 | 123689597 | T | C | 0.569 | 0.021 | 0.002 | 3.3e-23 |
| rs10985915 | 9 | 126018715 | G | A | 0.112 | -0.023 | 0.003 | 1.6e-11 |
| rs2791737 | 9 | 133632758 | G | A | 0.398 | -0.015 | 0.002 | 3.2e-12 |
| rs2427964 | 9 | 136931957 | C | T | 0.302 | 0.015 | 0.002 | 2e-10 |
| rs17250351 | 9 | 139801262 | A | G | 0.011 | 0.088 | 0.010 | 9.1e-18 |
| rs61839660 | 10 | 6094697 | T | C | 0.098 | -0.046 | 0.004 | 8.5e-39 |
| rs3824667 | 10 | 8100125 | G | A | 0.827 | 0.027 | 0.003 | 3.1e-21 |
| rs11593057 | 10 | 14887979 | T | C | 0.161 | -0.018 | 0.003 | 1.7e-10 |
| rs10828725 | 10 | 25218243 | T | G | 0.369 | -0.033 | 0.002 | 1.7e-50 |
| rs2992263 | 10 | 26741207 | A | G | 0.609 | -0.020 | 0.002 | 2.9e-20 |
| rs2091084 | 10 | 30492267 | C | T | 0.653 | 0.014 | 0.002 | 1.3e-09 |
| rs72790861 | 10 | 44880207 | G | C | 0.308 | -0.018 | 0.002 | 7e-15 |
| rs61850684 | 10 | 50252936 | A | G | 0.235 | -0.034 | 0.003 | 6.4e-42 |
| rs4948492 | 10 | 63719739 | T | C | 0.660 | -0.024 | 0.002 | 5.7e-26 |
| rs224082 | 10 | 64566258 | T | C | 0.377 | -0.015 | 0.002 | 4.1e-12 |
| rs10762264 | 10 | 70976833 | A | G | 0.685 | -0.021 | 0.002 | 1.5e-20 |
| rs10823631 | 10 | 72625741 | G | A | 0.326 | -0.019 | 0.002 | 2.1e-16 |
| rs748113 | 10 | 73508791 | C | T | 0.436 | -0.035 | 0.002 | 1.3e-61 |
| rs1878036 | 10 | 82280137 | G | T | 0.794 | 0.018 | 0.003 | 2.4e-11 |
| rs11187162 | 10 | 94508636 | A | G | 0.161 | 0.021 | 0.003 | 7.7e-13 |
| rs58667319 | 10 | 96524395 | T | C | 0.123 | -0.024 | 0.003 | 1.8e-13 |
| rs41317270 | 10 | 98369755 | T | C | 0.177 | -0.018 | 0.003 | 3.2e-11 |
| rs11189058 | 10 | 99001258 | A | G | 0.661 | 0.014 | 0.002 | 2.7e-10 |
| rs603424 | 10 | 102075479 | A | G | 0.171 | 0.021 | 0.003 | 4.1e-14 |
| rs35896146 | 10 | 116024458 | A | G | 0.159 | 0.024 | 0.003 | 4e-16 |
| rs7071131 | 10 | 121026712 | G | A | 0.495 | -0.017 | 0.002 | 4.1e-15 |
| rs10794175 | 10 | 126358073 | T | G | 0.415 | -0.022 | 0.002 | 3.3e-24 |
| rs574185 | 11 | 221322 | T | G | 0.538 | 0.017 | 0.002 | 2.8e-16 |
| rs7945319 | 11 | 405197 | A | G | 0.553 | 0.014 | 0.002 | 1.2e-10 |
| rs61867141 | 11 | 1874892 | A | G | 0.195 | -0.020 | 0.003 | 1.1e-13 |
| rs4756265 | 11 | 36078171 | G | A | 0.418 | 0.020 | 0.002 | 1.1e-19 |
| rs10838634 | 11 | 46929954 | G | A | 0.902 | -0.026 | 0.004 | 5.8e-13 |
| rs7123436 | 11 | 48013484 | A | G | 0.201 | -0.038 | 0.003 | 7e-48 |
| rs547957008 | 11 | 60258999 | T | A | 0.517 | 0.017 | 0.002 | 4.7e-16 |
| rs74679312 | 11 | 62194434 | G | A | 0.058 | 0.032 | 0.005 | 5.4e-12 |
| rs949349 | 11 | 86297189 | C | T | 0.270 | 0.014 | 0.002 | 4.9e-09 |
| rs1111890 | 11 | 100501571 | G | C | 0.368 | 0.023 | 0.002 | 1.7e-25 |
| rs1800057 | 11 | 108143456 | G | C | 0.027 | 0.040 | 0.007 | 1.1e-09 |
| rs58432776 | 11 | 122514281 | A | C | 0.380 | 0.032 | 0.002 | 1.8e-49 |
| rs35871881 | 11 | 123356451 | G | A | 0.225 | 0.016 | 0.003 | 2.1e-10 |
| rs10790962 | 11 | 128385169 | A | G | 0.469 | 0.023 | 0.002 | 2.5e-26 |
| rs34038797 | 12 | 740009 | G | C | 0.479 | -0.021 | 0.002 | 8.8e-22 |
| rs10466905 | 12 | 6502832 | A | G | 0.190 | 0.060 | 0.003 | 2.7e-110 |
| rs768908 | 12 | 10035304 | T | C | 0.125 | -0.020 | 0.003 | 2.5e-10 |
| rs34325 | 12 | 12877692 | C | T | 0.524 | -0.027 | 0.002 | 4.1e-36 |
| rs7134738 | 12 | 26358170 | C | T | 0.351 | -0.017 | 0.002 | 4.7e-14 |
| rs1900942 | 12 | 26831928 | G | C | 0.839 | 0.018 | 0.003 | 2e-10 |
| rs11168249 | 12 | 48208368 | C | T | 0.461 | -0.018 | 0.002 | 8e-18 |
| rs73108566 | 12 | 49080698 | A | T | 0.329 | -0.019 | 0.002 | 1.4e-16 |
| rs1131017 | 12 | 56435929 | G | C | 0.572 | 0.014 | 0.002 | 3.6e-11 |
| rs4913407 | 12 | 68560246 | T | G | 0.705 | 0.016 | 0.002 | 5.2e-12 |
| rs6538189 | 12 | 89885923 | A | C | 0.239 | 0.016 | 0.002 | 7.5e-11 |
| rs17041439 | 12 | 101873240 | C | A | 0.056 | 0.043 | 0.005 | 2e-20 |
| rs4764803 | 12 | 101898230 | G | C | 0.616 | 0.015 | 0.002 | 7.5e-12 |
| rs1265564 | 12 | 111708458 | C | A | 0.441 | 0.073 | 0.002 | 1e-200 |
| rs73207626 | 12 | 112686873 | G | A | 0.043 | -0.035 | 0.005 | 1.9e-11 |
| rs373582220 | 12 | 112847812 | A | G | 0.372 | -0.035 | 0.003 | 6e-38 |
| rs1293782 | 12 | 113463594 | A | C | 0.643 | -0.015 | 0.002 | 1.4e-11 |
| rs6488868 | 12 | 123799974 | G | A | 0.738 | -0.027 | 0.002 | 5.3e-29 |
| rs67516712 | 12 | 124201524 | A | G | 0.425 | 0.016 | 0.002 | 4.1e-14 |
| rs3768 | 12 | 124499839 | T | C | 0.194 | -0.018 | 0.003 | 5.2e-11 |
| rs76428106 | 13 | 28604007 | C | T | 0.013 | 0.063 | 0.010 | 7.7e-11 |
| rs9532679 | 13 | 41522338 | C | A | 0.150 | -0.032 | 0.003 | 1.7e-26 |
| rs9533100 | 13 | 42996548 | T | G | 0.536 | -0.021 | 0.002 | 9.3e-23 |
| rs2146880 | 13 | 46706598 | A | C | 0.461 | 0.015 | 0.002 | 3.4e-13 |
| rs9316484 | 13 | 50645164 | C | T | 0.220 | -0.028 | 0.003 | 3.8e-27 |
| rs67483792 | 13 | 72503638 | T | C | 0.043 | -0.036 | 0.005 | 1.3e-11 |
| rs9592965 | 13 | 74615925 | C | A | 0.327 | -0.015 | 0.002 | 1.3e-10 |
| rs3812849 | 13 | 74701736 | C | A | 0.264 | 0.045 | 0.002 | 2.7e-76 |
| rs9513573 | 13 | 99815898 | A | G | 0.279 | -0.015 | 0.002 | 3.2e-10 |
| rs1320472 | 13 | 114843162 | C | T | 0.460 | -0.014 | 0.002 | 4.2e-11 |
| rs9590390 | 13 | 114920867 | A | G | 0.294 | 0.030 | 0.002 | 1.4e-37 |
| rs45528934 | 14 | 23793305 | T | C | 0.162 | -0.019 | 0.003 | 7.9e-11 |
| rs2057340 | 14 | 35848774 | G | A | 0.655 | 0.016 | 0.002 | 1.2e-12 |
| rs696 | 14 | 35871093 | T | C | 0.363 | 0.027 | 0.002 | 2.3e-34 |
| rs11848179 | 14 | 64977053 | G | A | 0.318 | 0.030 | 0.002 | 2.1e-38 |
| rs10138752 | 14 | 69179971 | T | C | 0.078 | -0.029 | 0.004 | 3.2e-13 |
| rs175714 | 14 | 75981856 | C | T | 0.577 | 0.027 | 0.002 | 5.1e-36 |
| rs7157267 | 14 | 103350813 | C | T | 0.221 | 0.030 | 0.003 | 9.5e-33 |
| rs12882130 | 14 | 103878774 | G | C | 0.384 | 0.017 | 0.002 | 5e-15 |
| rs11855923 | 15 | 40892601 | A | G | 0.392 | -0.017 | 0.002 | 1.5e-15 |
| rs139974673 | 15 | 44027885 | C | T | 0.026 | 0.054 | 0.007 | 1.8e-15 |
| rs12593807 | 15 | 48546727 | C | T | 0.135 | -0.022 | 0.003 | 6.8e-13 |
| rs7161799 | 15 | 58770523 | T | C | 0.075 | 0.038 | 0.004 | 1.3e-20 |
| rs149453951 | 15 | 69583434 | T | C | 0.024 | 0.048 | 0.007 | 3.6e-11 |
| rs34025077 | 15 | 70033578 | G | A | 0.103 | -0.033 | 0.003 | 2.1e-21 |
| rs631864 | 15 | 70376441 | C | T | 0.472 | 0.014 | 0.002 | 5.9e-11 |
| rs17507693 | 15 | 83793622 | T | A | 0.222 | 0.017 | 0.003 | 2.6e-11 |
| rs17832339 | 15 | 90557494 | T | G | 0.039 | 0.050 | 0.005 | 1.4e-19 |
| rs8030465 | 15 | 90656372 | G | C | 0.573 | -0.013 | 0.002 | 2e-09 |
| rs28539372 | 15 | 91021412 | A | T | 0.319 | 0.040 | 0.002 | 1.3e-69 |
| rs447361 | 16 | 4148240 | G | T | 0.669 | 0.020 | 0.002 | 2.7e-18 |
| rs1677490 | 16 | 9028541 | C | G | 0.794 | 0.016 | 0.003 | 1.7e-09 |
| rs17229044 | 16 | 11062936 | T | C | 0.208 | -0.019 | 0.003 | 7.2e-13 |
| rs11643225 | 16 | 24046655 | T | G | 0.059 | -0.033 | 0.005 | 2e-13 |
| rs11574938 | 16 | 30485393 | C | G | 0.517 | 0.057 | 0.002 | 1.1e-155 |
| rs28853644 | 16 | 30801027 | T | C | 0.270 | -0.025 | 0.002 | 2.3e-26 |
| rs9746394 | 16 | 31300769 | C | T | 0.270 | -0.016 | 0.002 | 2.9e-11 |
| rs7192652 | 16 | 57075180 | G | A | 0.478 | 0.022 | 0.002 | 1.1e-25 |
| rs80180406 | 16 | 67435558 | A | G | 0.023 | -0.047 | 0.007 | 3.6e-11 |
| rs117556162 | 16 | 67680806 | A | G | 0.059 | 0.044 | 0.004 | 1.4e-22 |
| rs8058619 | 16 | 75182354 | C | G | 0.308 | -0.018 | 0.002 | 2.7e-14 |
| rs11150084 | 16 | 78561820 | G | C | 0.669 | 0.015 | 0.002 | 1.9e-11 |
| rs247826 | 16 | 84582965 | T | C | 0.222 | 0.037 | 0.003 | 7.9e-48 |
| rs8060375 | 16 | 88507538 | T | C | 0.321 | 0.014 | 0.002 | 6.5e-10 |
| rs8052370 | 16 | 88837298 | T | C | 0.610 | -0.023 | 0.002 | 3.9e-25 |
| rs7214308 | 17 | 1965085 | T | C | 0.742 | -0.015 | 0.002 | 3.6e-10 |
| rs6502555 | 17 | 2729652 | C | T | 0.269 | 0.029 | 0.002 | 7.5e-34 |
| rs62091998 | 17 | 2881625 | G | A | 0.308 | -0.025 | 0.002 | 5.1e-27 |
| rs34230287 | 17 | 4613630 | T | C | 0.239 | 0.031 | 0.002 | 2.5e-36 |
| rs62073635 | 17 | 4770197 | C | T | 0.346 | -0.027 | 0.002 | 4.3e-33 |
| rs77234976 | 17 | 7141140 | G | C | 0.145 | -0.024 | 0.003 | 5.3e-15 |
| rs11652760 | 17 | 16786819 | G | T | 0.101 | -0.025 | 0.004 | 2.1e-12 |
| rs2297508 | 17 | 17715317 | G | C | 0.649 | -0.023 | 0.002 | 8.7e-26 |
| rs2453582 | 17 | 19439066 | T | C | 0.386 | 0.018 | 0.002 | 6.6e-17 |
| rs3760456 | 17 | 27948844 | T | C | 0.441 | -0.016 | 0.002 | 9.8e-14 |
| rs2522972 | 17 | 35839021 | T | C | 0.639 | -0.022 | 0.002 | 7.4e-23 |
| rs4795397 | 17 | 38023745 | G | A | 0.482 | -0.031 | 0.002 | 2.1e-48 |
| rs3136685 | 17 | 38719799 | T | C | 0.161 | -0.021 | 0.003 | 8.7e-13 |
| rs72836561 | 17 | 41926126 | T | C | 0.031 | -0.037 | 0.006 | 7.3e-10 |
| rs1295927 | 17 | 57929535 | G | A | 0.447 | -0.022 | 0.002 | 1.2e-25 |
| rs9893518 | 17 | 62006997 | C | T | 0.258 | -0.014 | 0.002 | 3.8e-09 |
| rs16978167 | 17 | 72673667 | T | A | 0.764 | 0.024 | 0.003 | 2.4e-21 |
| rs34891561 | 17 | 73027851 | T | G | 0.333 | -0.015 | 0.002 | 4.4e-11 |
| rs2665967 | 17 | 74016834 | C | T | 0.732 | 0.015 | 0.002 | 1.2e-09 |
| rs4789299 | 17 | 74452010 | A | G | 0.203 | 0.030 | 0.003 | 2.6e-29 |
| rs8096327 | 18 | 12887750 | G | A | 0.391 | -0.013 | 0.002 | 2.1e-09 |
| rs3851820 | 18 | 20461322 | T | A | 0.744 | -0.016 | 0.002 | 1.2e-10 |
| rs77551289 | 18 | 60788745 | G | A | 0.098 | -0.028 | 0.004 | 9.2e-15 |
| rs4940572 | 18 | 60826343 | A | G | 0.134 | 0.024 | 0.003 | 2.8e-14 |
| rs111304587 | 18 | 77271669 | G | T | 0.085 | 0.029 | 0.004 | 2.5e-14 |
| rs77232119 | 19 | 300952 | G | T | 0.273 | 0.015 | 0.002 | 1.9e-10 |
| rs35140707 | 19 | 1078297 | T | C | 0.100 | -0.059 | 0.004 | 1.8e-63 |
| rs28841391 | 19 | 1193965 | G | T | 0.759 | 0.028 | 0.003 | 2.8e-27 |
| rs35251378 | 19 | 10459969 | A | G | 0.293 | 0.044 | 0.002 | 2.8e-78 |
| rs56252104 | 19 | 13123188 | T | C | 0.261 | 0.019 | 0.002 | 1.1e-15 |
| rs62124251 | 19 | 13915213 | T | G | 0.348 | 0.032 | 0.002 | 2.6e-47 |
| rs35024802 | 19 | 16388636 | T | C | 0.148 | -0.023 | 0.003 | 1.9e-14 |
| rs56133626 | 19 | 16441973 | A | G | 0.328 | -0.070 | 0.002 | 1e-200 |
| rs183685887 | 19 | 16598721 | C | T | 0.039 | 0.116 | 0.006 | 6.7e-97 |
| rs4530278 | 19 | 33752994 | T | G | 0.601 | 0.019 | 0.002 | 4.3e-18 |
| rs4805881 | 19 | 33896432 | C | A | 0.665 | -0.015 | 0.002 | 3.3e-11 |
| rs75018496 | 19 | 40785683 | G | C | 0.077 | 0.024 | 0.004 | 1.6e-09 |
| rs61387190 | 19 | 44260929 | T | C | 0.157 | 0.030 | 0.003 | 1.1e-25 |
| rs34159938 | 19 | 47690811 | A | G | 0.259 | 0.031 | 0.002 | 4.3e-38 |
| rs11084096 | 19 | 52128795 | A | G | 0.296 | -0.018 | 0.002 | 1.4e-14 |
| rs2070745 | 19 | 52249947 | G | C | 0.365 | -0.017 | 0.002 | 6.3e-14 |
| rs78487206 | 19 | 55347738 | T | G | 0.165 | -0.019 | 0.003 | 2.8e-10 |
| rs4630833 | 20 | 1930798 | C | T | 0.263 | -0.045 | 0.002 | 5.1e-79 |
| rs1883932 | 20 | 8609588 | T | A | 0.508 | -0.028 | 0.002 | 6.3e-40 |
| rs34952318 | 20 | 11177055 | A | G | 0.049 | -0.037 | 0.005 | 1.3e-13 |
| rs1997797 | 20 | 31387954 | G | C | 0.447 | 0.018 | 0.002 | 7.6e-17 |
| rs7273174 | 20 | 34350306 | C | T | 0.110 | -0.020 | 0.003 | 2.9e-09 |
| rs6072080 | 20 | 39260927 | T | C | 0.585 | 0.021 | 0.002 | 2e-21 |
| rs4812803 | 20 | 42850827 | T | C | 0.285 | 0.015 | 0.002 | 2.9e-10 |
| rs13045492 | 20 | 49134535 | T | A | 0.084 | 0.031 | 0.004 | 1.2e-15 |
| rs34323943 | 20 | 52192637 | C | T | 0.111 | 0.023 | 0.003 | 2e-11 |
| rs343829 | 20 | 57553504 | C | G | 0.929 | -0.027 | 0.004 | 4.6e-11 |
| rs259981 | 20 | 57748369 | A | T | 0.359 | -0.028 | 0.002 | 9.2e-36 |
| rs6064762 | 20 | 57867578 | C | T | 0.539 | -0.013 | 0.002 | 2.9e-10 |
| rs1997577 | 21 | 16371102 | T | A | 0.155 | -0.018 | 0.003 | 1.5e-09 |
| rs150797 | 21 | 43465831 | G | C | 0.454 | -0.015 | 0.002 | 8.6e-12 |
| rs1893592 | 21 | 43855067 | C | A | 0.300 | 0.016 | 0.002 | 2.7e-12 |
| rs721131 | 21 | 47957767 | C | T | 0.320 | -0.020 | 0.002 | 3.9e-19 |
| rs1034564 | 22 | 19984013 | T | C | 0.286 | 0.022 | 0.002 | 3.5e-21 |
| rs4821124 | 22 | 21979289 | C | T | 0.189 | -0.043 | 0.003 | 6e-56 |
| rs552292854 | 22 | 24654627 | T | C | 0.254 | -0.017 | 0.003 | 6.4e-11 |
| rs4583569 | 22 | 28933106 | T | C | 0.878 | 0.033 | 0.003 | 7.1e-24 |
| rs6006356 | 22 | 30503171 | A | G | 0.446 | 0.035 | 0.002 | 1e-61 |
| rs139425 | 22 | 39559742 | C | G | 0.451 | -0.027 | 0.002 | 6.2e-36 |
| rs202638 | 22 | 41855458 | G | A | 0.795 | -0.027 | 0.003 | 2.5e-24 |
| rs9607869 | 22 | 42418110 | A | T | 0.299 | 0.015 | 0.002 | 6.2e-11 |

**Table S12. 456 SNPs significantly associated with lymphocyte count used IVs in forward MR analyses derived from Chen MH et al.** Chr: Chromosome; EA: Effect allele; NEA: Non-effect allele; EAF: Effect allele frequency.

| SNP | Chr | Pos | EA | NEA | EAF | Beta | SE | P-value |
| --- | --- | --- | --- | --- | --- | --- | --- | --- |
| rs603424 | 10 | 102075479 | A | G | 0.175 | 0.020 | 0.002 | 2.78e-16 |
| rs35896146 | 10 | 116024458 | A | G | 0.159 | 0.022 | 0.003 | 8.15e-17 |
| rs7071131 | 10 | 121026712 | G | A | 0.495 | -0.016 | 0.002 | 3.85e-16 |
| rs10794175 | 10 | 126358073 | T | G | 0.416 | -0.021 | 0.002 | 2.5e-26 |
| rs35001362 | 10 | 14958758 | A | C | 0.236 | -0.015 | 0.002 | 1.52e-11 |
| rs10828725 | 10 | 25218243 | T | G | 0.366 | -0.032 | 0.002 | 1.29e-59 |
| rs2992263 | 10 | 26741207 | A | G | 0.610 | -0.020 | 0.002 | 1.58e-25 |
| rs2462017 | 10 | 30486133 | A | G | 0.652 | 0.014 | 0.002 | 2.87e-12 |
| rs7093481 | 10 | 44879711 | G | A | 0.307 | -0.017 | 0.002 | 6.84e-16 |
| rs61850684 | 10 | 50252936 | A | G | 0.234 | -0.032 | 0.002 | 2.59e-47 |
| rs3118469 | 10 | 6101129 | T | A | 0.300 | 0.029 | 0.002 | 1.5e-45 |
| rs4948492 | 10 | 63719739 | T | C | 0.662 | -0.022 | 0.002 | 5.69e-28 |
| rs224082 | 10 | 64566258 | T | C | 0.378 | -0.016 | 0.002 | 1.79e-16 |
| rs10762264 | 10 | 70976833 | A | G | 0.681 | -0.020 | 0.002 | 1.06e-23 |
| rs10823631 | 10 | 72625741 | G | A | 0.325 | -0.017 | 0.002 | 3.11e-17 |
| rs703257 | 10 | 73119135 | T | C | 0.888 | -0.021 | 0.003 | 3.71e-12 |
| rs748113 | 10 | 73508791 | C | T | 0.436 | -0.034 | 0.002 | 1.53e-65 |
| rs10748526 | 10 | 82273079 | C | T | 0.794 | 0.015 | 0.002 | 7.88e-11 |
| rs11187162 | 10 | 94508636 | A | G | 0.162 | 0.020 | 0.003 | 2.61e-15 |
| rs58667319 | 10 | 96524395 | T | C | 0.123 | -0.023 | 0.003 | 9.91e-16 |
| rs41317270 | 10 | 98369755 | T | C | 0.177 | -0.018 | 0.002 | 9.82e-14 |
| rs11189058 | 10 | 99001258 | A | G | 0.660 | 0.012 | 0.002 | 7.31e-10 |
| rs11224302 | 11 | 100456604 | T | C | 0.099 | -0.038 | 0.003 | 1.73e-33 |
| rs17116384 | 11 | 113981241 | G | A | 0.317 | 0.013 | 0.002 | 4.5e-10 |
| rs1893033 | 11 | 119089055 | T | C | 0.075 | -0.028 | 0.004 | 5.7e-15 |
| rs11218725 | 11 | 122509237 | G | A | 0.377 | 0.033 | 0.002 | 1.03e-63 |
| rs79805393 | 11 | 128412297 | C | T | 0.173 | 0.017 | 0.003 | 1.04e-11 |
| rs7127911 | 11 | 128492549 | T | G | 0.266 | 0.024 | 0.002 | 3.44e-29 |
| rs2403246 | 11 | 18101078 | G | C | 0.414 | 0.012 | 0.002 | 4.81e-10 |
| rs61867141 | 11 | 1874892 | A | G | 0.195 | -0.017 | 0.003 | 1.37e-11 |
| rs11033388 | 11 | 36074218 | T | A | 0.419 | 0.020 | 0.002 | 2.07e-24 |
| rs74045276 | 11 | 403484 | C | G | 0.565 | 0.013 | 0.002 | 2.85e-11 |
| rs10838634 | 11 | 46929954 | G | A | 0.902 | -0.026 | 0.003 | 3e-16 |
| rs1017875 | 11 | 47999218 | T | C | 0.202 | -0.036 | 0.002 | 5.01e-53 |
| rs74679312 | 11 | 62194434 | G | A | 0.057 | 0.031 | 0.004 | 2.28e-13 |
| rs34662054 | 11 | 68908815 | C | G | 0.299 | 0.014 | 0.002 | 4.8e-12 |
| rs949349 | 11 | 86297189 | C | T | 0.271 | 0.014 | 0.002 | 3.26e-11 |
| rs768908 | 12 | 10035304 | T | C | 0.125 | -0.020 | 0.003 | 2.4e-12 |
| rs17041439 | 12 | 101873240 | C | A | 0.057 | 0.041 | 0.004 | 2.11e-23 |
| rs4764803 | 12 | 101898230 | G | C | 0.617 | 0.014 | 0.002 | 4.87e-13 |
| rs1265564 | 12 | 111708458 | C | A | 0.443 | 0.072 | 0.002 | 1e-200 |
| rs73207626 | 12 | 112686873 | G | A | 0.042 | -0.036 | 0.005 | 9.72e-14 |
| rs1293782 | 12 | 113463594 | A | C | 0.644 | -0.015 | 0.002 | 3.74e-14 |
| rs28533432 | 12 | 123873242 | T | C | 0.700 | -0.025 | 0.002 | 7.17e-33 |
| rs67516712 | 12 | 124201524 | A | G | 0.423 | 0.015 | 0.002 | 9.11e-15 |
| rs10846577 | 12 | 124400261 | C | T | 0.442 | -0.014 | 0.002 | 2.37e-13 |
| rs34327 | 12 | 12873748 | C | T | 0.523 | -0.026 | 0.002 | 5.91e-42 |
| rs6487543 | 12 | 26438189 | A | G | 0.771 | 0.020 | 0.002 | 6.2e-18 |
| rs11611710 | 12 | 4340527 | C | A | 0.269 | -0.013 | 0.002 | 2.45e-09 |
| rs11168249 | 12 | 48208368 | C | T | 0.461 | -0.019 | 0.002 | 6.04e-24 |
| rs73108566 | 12 | 49080698 | A | T | 0.332 | -0.017 | 0.002 | 1.23e-16 |
| rs4761790 | 12 | 51749966 | A | G | 0.763 | 0.014 | 0.002 | 8.26e-10 |
| rs706819 | 12 | 52315923 | C | T | 0.738 | 0.014 | 0.002 | 3.61e-10 |
| rs1131017 | 12 | 56435929 | G | C | 0.573 | 0.014 | 0.002 | 9.76e-13 |
| rs10466905 | 12 | 6502832 | A | G | 0.190 | 0.062 | 0.002 | 7.09e-140 |
| rs1118866 | 12 | 68520751 | T | C | 0.704 | 0.015 | 0.002 | 1.1e-12 |
| rs34038797 | 12 | 740009 | G | C | 0.481 | -0.019 | 0.002 | 9.44e-23 |
| rs7308380 | 12 | 89851232 | C | A | 0.751 | 0.016 | 0.002 | 2.47e-13 |
| rs1371076 | 12 | 90561450 | C | T | 0.740 | 0.015 | 0.002 | 4.22e-12 |
| rs374053567 | 13 | 114454143 | G | A | 0.042 | 0.031 | 0.005 | 2.05e-09 |
| rs2001388 | 13 | 114843054 | C | T | 0.461 | -0.014 | 0.002 | 1.55e-12 |
| rs9590390 | 13 | 114920867 | A | G | 0.292 | 0.030 | 0.002 | 2.51e-46 |
| rs76428106 | 13 | 28604007 | C | T | 0.013 | 0.063 | 0.009 | 5.49e-13 |
| rs9532679 | 13 | 41522338 | C | A | 0.149 | -0.032 | 0.003 | 6.33e-33 |
| rs9525619 | 13 | 42992135 | T | C | 0.534 | -0.020 | 0.002 | 1.08e-26 |
| rs9534338 | 13 | 46707137 | C | T | 0.460 | 0.015 | 0.002 | 1.38e-15 |
| rs9568054 | 13 | 49179386 | A | C | 0.323 | 0.014 | 0.002 | 1.41e-11 |
| rs1523178 | 13 | 50652456 | C | A | 0.221 | -0.026 | 0.002 | 3.86e-29 |
| rs912416 | 13 | 50933553 | C | A | 0.445 | -0.012 | 0.002 | 3.58e-10 |
| rs67483792 | 13 | 72503638 | T | C | 0.043 | -0.035 | 0.005 | 8.06e-14 |
| rs9592965 | 13 | 74615925 | C | A | 0.327 | -0.014 | 0.002 | 8.83e-13 |
| rs3812849 | 13 | 74701736 | C | A | 0.266 | 0.043 | 0.002 | 1.32e-91 |
| rs7157267 | 14 | 103350813 | C | T | 0.222 | 0.030 | 0.002 | 1.99e-40 |
| rs4900575 | 14 | 103899169 | C | G | 0.366 | 0.018 | 0.002 | 2.03e-20 |
| rs45528934 | 14 | 23793305 | T | C | 0.162 | -0.021 | 0.003 | 1.85e-15 |
| rs2057340 | 14 | 35848774 | G | A | 0.653 | 0.014 | 0.002 | 4.66e-12 |
| rs696 | 14 | 35871093 | T | C | 0.365 | 0.025 | 0.002 | 4.94e-38 |
| rs1966865 | 14 | 64975861 | A | G | 0.317 | 0.027 | 0.002 | 1.36e-40 |
| rs10138752 | 14 | 69179971 | T | C | 0.077 | -0.030 | 0.004 | 4.89e-17 |
| rs175714 | 14 | 75981856 | C | T | 0.577 | 0.026 | 0.002 | 9.2e-42 |
| rs2180369 | 14 | 93516465 | C | T | 0.111 | 0.020 | 0.003 | 1.47e-10 |
| rs10145277 | 14 | 99793449 | A | T | 0.625 | -0.015 | 0.002 | 3.19e-14 |
| rs2412544 | 15 | 40949526 | T | C | 0.394 | -0.016 | 0.002 | 1.13e-16 |
| rs139974673 | 15 | 44027885 | C | T | 0.026 | 0.054 | 0.006 | 1.89e-19 |
| rs12593807 | 15 | 48546727 | C | T | 0.136 | -0.021 | 0.003 | 7.57e-14 |
| rs7161799 | 15 | 58770523 | T | C | 0.076 | 0.034 | 0.004 | 2.79e-21 |
| rs2439408 | 15 | 66925163 | C | G | 0.561 | -0.012 | 0.002 | 9.72e-11 |
| rs149453951 | 15 | 69583434 | T | C | 0.024 | 0.049 | 0.007 | 4.04e-13 |
| rs34025077 | 15 | 70033578 | G | A | 0.101 | -0.033 | 0.003 | 1.25e-25 |
| rs631864 | 15 | 70376441 | C | T | 0.472 | 0.014 | 0.002 | 1.48e-12 |
| rs17507693 | 15 | 83793622 | T | A | 0.223 | 0.016 | 0.002 | 9.86e-13 |
| rs187856913 | 15 | 90555181 | A | G | 0.039 | 0.048 | 0.005 | 6.24e-22 |
| rs11073903 | 15 | 90653194 | A | G | 0.567 | -0.013 | 0.002 | 4.41e-11 |
| rs28539372 | 15 | 91021412 | A | T | 0.319 | 0.038 | 0.002 | 3.38e-78 |
| rs17229044 | 16 | 11062936 | T | C | 0.210 | -0.019 | 0.002 | 1.78e-16 |
| rs145719494 | 16 | 24044921 | G | A | 0.059 | -0.031 | 0.004 | 3.71e-14 |
| rs12598978 | 16 | 30482540 | G | T | 0.520 | 0.055 | 0.002 | 1.58e-181 |
| rs28853644 | 16 | 30801027 | T | C | 0.271 | -0.024 | 0.002 | 3.77e-30 |
| rs78487935 | 16 | 3651579 | G | A | 0.078 | -0.024 | 0.004 | 3.2e-11 |
| rs447361 | 16 | 4148240 | G | T | 0.666 | 0.018 | 0.002 | 6.1e-19 |
| rs7192652 | 16 | 57075180 | G | A | 0.481 | 0.022 | 0.002 | 4.32e-31 |
| rs149012671 | 16 | 67596482 | G | C | 0.044 | -0.030 | 0.005 | 1.89e-10 |
| rs117556162 | 16 | 67680806 | A | G | 0.058 | 0.044 | 0.004 | 3.31e-27 |
| rs9939124 | 16 | 75185764 | T | C | 0.307 | -0.017 | 0.002 | 3.6e-16 |
| rs247826 | 16 | 84582965 | T | C | 0.221 | 0.039 | 0.002 | 1.19e-63 |
| rs56292801 | 16 | 88535341 | A | G | 0.273 | 0.014 | 0.002 | 1.02e-09 |
| rs8052370 | 16 | 88837298 | T | C | 0.609 | -0.021 | 0.002 | 5.82e-25 |
| rs1677490 | 16 | 9028541 | C | G | 0.791 | 0.016 | 0.002 | 1.25e-11 |
| rs11652968 | 17 | 16813349 | T | C | 0.190 | -0.016 | 0.002 | 8.04e-11 |
| rs2297508 | 17 | 17715317 | G | C | 0.648 | -0.022 | 0.002 | 2.2e-27 |
| rs2453582 | 17 | 19439066 | T | C | 0.391 | 0.018 | 0.002 | 4.88e-19 |
| rs6502555 | 17 | 2729652 | C | T | 0.269 | 0.029 | 0.002 | 9.26e-38 |
| rs3760456 | 17 | 27948844 | T | C | 0.439 | -0.015 | 0.002 | 1.4e-14 |
| rs62091998 | 17 | 2881625 | G | A | 0.307 | -0.022 | 0.002 | 1.33e-27 |
| rs2522972 | 17 | 35839021 | T | C | 0.640 | -0.020 | 0.002 | 6.54e-25 |
| rs4795397 | 17 | 38023745 | G | A | 0.479 | -0.030 | 0.002 | 3.62e-57 |
| rs2072081 | 17 | 42327493 | T | G | 0.255 | -0.017 | 0.002 | 2.53e-14 |
| rs35186877 | 17 | 4625892 | A | G | 0.214 | 0.032 | 0.002 | 1.94e-42 |
| rs8075090 | 17 | 4969108 | C | T | 0.524 | 0.021 | 0.002 | 2.01e-28 |
| rs9898958 | 17 | 55242242 | G | T | 0.153 | 0.016 | 0.003 | 3.56e-09 |
| rs1292069 | 17 | 57928290 | C | T | 0.450 | -0.023 | 0.002 | 3.09e-32 |
| rs11657448 | 17 | 62023128 | T | C | 0.322 | 0.013 | 0.002 | 8.47e-10 |
| rs1215 | 17 | 7163350 | G | A | 0.144 | -0.023 | 0.003 | 3.67e-17 |
| rs9906320 | 17 | 72690829 | A | G | 0.775 | 0.025 | 0.002 | 8.67e-27 |
| rs1491765 | 17 | 73028923 | A | G | 0.293 | -0.017 | 0.002 | 1.67e-16 |
| rs2665960 | 17 | 74024711 | G | A | 0.669 | 0.012 | 0.002 | 4.03e-09 |
| rs4789294 | 17 | 74419177 | G | A | 0.219 | 0.027 | 0.002 | 1.05e-30 |
| rs116876036 | 17 | 76393762 | T | C | 0.090 | 0.021 | 0.003 | 9.57e-10 |
| rs8096327 | 18 | 12887750 | G | A | 0.391 | -0.013 | 0.002 | 1.59e-10 |
| rs3851820 | 18 | 20461322 | T | A | 0.745 | -0.018 | 0.002 | 7.37e-16 |
| rs4987855 | 18 | 60793549 | T | C | 0.096 | -0.024 | 0.003 | 1.16e-13 |
| rs4940572 | 18 | 60826343 | A | G | 0.136 | 0.023 | 0.003 | 1.17e-16 |
| rs9676181 | 18 | 74769175 | A | T | 0.574 | 0.012 | 0.002 | 9.73e-10 |
| rs111626441 | 18 | 77250669 | A | G | 0.097 | 0.028 | 0.003 | 8.97e-17 |
| rs36084354 | 19 | 1079959 | A | G | 0.092 | -0.061 | 0.003 | 6.6e-71 |
| rs3817621 | 19 | 12998205 | C | G | 0.238 | -0.017 | 0.002 | 1.11e-13 |
| rs345627 | 19 | 13917264 | T | C | 0.349 | 0.034 | 0.002 | 3.13e-60 |
| rs897791 | 19 | 16490859 | G | A | 0.967 | -0.096 | 0.005 | 3.3e-69 |
| rs138243594 | 19 | 16495586 | A | G | 0.013 | -0.076 | 0.009 | 4.57e-16 |
| rs148213173 | 19 | 16647026 | C | A | 0.054 | 0.108 | 0.004 | 5.63e-146 |
| rs4530278 | 19 | 33752994 | T | G | 0.598 | 0.017 | 0.002 | 5.24e-18 |
| rs4805881 | 19 | 33896432 | C | A | 0.665 | -0.015 | 0.002 | 1.41e-13 |
| rs75018496 | 19 | 40785683 | G | C | 0.076 | 0.025 | 0.004 | 1.6e-11 |
| rs60505738 | 19 | 4089811 | T | C | 0.090 | 0.026 | 0.004 | 1.2e-13 |
| rs61387190 | 19 | 44260929 | T | C | 0.157 | 0.032 | 0.003 | 5.81e-34 |
| rs56179616 | 19 | 47695751 | T | G | 0.259 | 0.031 | 0.002 | 6.22e-47 |
| rs3810276 | 19 | 50934939 | A | G | 0.405 | -0.012 | 0.002 | 1.47e-09 |
| rs11084096 | 19 | 52128795 | A | G | 0.296 | -0.018 | 0.002 | 6.42e-19 |
| rs2070745 | 19 | 52249947 | G | C | 0.367 | -0.016 | 0.002 | 3.94e-16 |
| rs12088882 | 1 | 101224620 | T | C | 0.408 | 0.034 | 0.002 | 1.3e-70 |
| rs10875371 | 1 | 101762529 | T | C | 0.418 | -0.021 | 0.002 | 3.37e-27 |
| rs77954449 | 1 | 10242292 | G | A | 0.028 | -0.037 | 0.006 | 9.78e-11 |
| rs2476601 | 1 | 114377568 | G | A | 0.900 | 0.054 | 0.003 | 1.47e-67 |
| rs11204702 | 1 | 150663507 | A | C | 0.510 | -0.026 | 0.002 | 3.37e-42 |
| rs967072 | 1 | 156972485 | T | C | 0.137 | -0.017 | 0.003 | 3.88e-10 |
| rs12045893 | 1 | 158530416 | T | C | 0.248 | 0.027 | 0.002 | 9.7e-35 |
| rs1633267 | 1 | 159010220 | G | T | 0.865 | -0.018 | 0.003 | 1.46e-10 |
| rs72634819 | 1 | 1596500 | T | C | 0.264 | -0.014 | 0.002 | 4.67e-10 |
| rs494091 | 1 | 160791892 | C | T | 0.426 | 0.027 | 0.002 | 6.35e-46 |
| rs60220286 | 1 | 161639559 | A | G | 0.175 | -0.021 | 0.003 | 8.03e-15 |
| rs6427618 | 1 | 161680258 | A | G | 0.621 | 0.014 | 0.002 | 1.55e-12 |
| rs2489279 | 1 | 182144119 | G | C | 0.334 | 0.018 | 0.002 | 1.28e-18 |
| rs72740550 | 1 | 197342380 | A | G | 0.222 | -0.019 | 0.002 | 1.58e-16 |
| rs10494783 | 1 | 198663661 | A | G | 0.052 | -0.054 | 0.004 | 8.95e-37 |
| rs113116201 | 1 | 198830942 | C | T | 0.020 | 0.041 | 0.007 | 1.93e-09 |
| rs2294630 | 1 | 19981791 | G | A | 0.493 | 0.013 | 0.002 | 8.15e-12 |
| rs11240335 | 1 | 203714629 | T | C | 0.067 | 0.029 | 0.004 | 6.49e-13 |
| rs4619033 | 1 | 204269242 | G | A | 0.268 | -0.019 | 0.002 | 5.73e-19 |
| rs1967115 | 1 | 212375359 | A | G | 0.629 | 0.017 | 0.002 | 1.04e-17 |
| rs12745411 | 1 | 214405704 | T | G | 0.600 | 0.017 | 0.002 | 6.32e-18 |
| rs12021809 | 1 | 225941782 | A | G | 0.110 | -0.026 | 0.003 | 2.69e-18 |
| rs533483 | 1 | 234765256 | A | G | 0.244 | -0.014 | 0.002 | 1.58e-10 |
| rs4660128 | 1 | 236087992 | C | T | 0.897 | -0.025 | 0.003 | 3.12e-15 |
| rs61778219 | 1 | 24235932 | T | C | 0.141 | 0.023 | 0.003 | 2.67e-17 |
| rs4925547 | 1 | 247612435 | T | A | 0.402 | 0.012 | 0.002 | 4.55e-10 |
| rs74227709 | 1 | 247722588 | A | G | 0.072 | -0.029 | 0.004 | 2.51e-15 |
| rs2282718 | 1 | 25241056 | A | G | 0.372 | -0.018 | 0.002 | 3.25e-18 |
| rs4274112 | 1 | 26746199 | G | A | 0.373 | 0.014 | 0.002 | 1.51e-12 |
| rs11247712 | 1 | 28229681 | T | C | 0.369 | -0.020 | 0.002 | 3.38e-24 |
| rs12142033 | 1 | 39542129 | A | G | 0.333 | -0.013 | 0.002 | 1.73e-10 |
| rs59543294 | 1 | 51495673 | C | T | 0.107 | 0.019 | 0.003 | 3.46e-10 |
| rs4319365 | 1 | 56907720 | A | G | 0.545 | -0.019 | 0.002 | 3.26e-24 |
| rs1933295 | 1 | 62107021 | G | A | 0.777 | -0.014 | 0.002 | 1.22e-09 |
| rs12127284 | 1 | 65431599 | T | C | 0.710 | -0.017 | 0.002 | 1.11e-15 |
| rs6700896 | 1 | 66089782 | T | C | 0.379 | -0.017 | 0.002 | 3.67e-19 |
| rs2755253 | 1 | 67470843 | T | C | 0.709 | -0.031 | 0.002 | 2.87e-50 |
| rs2275902 | 1 | 79357360 | C | G | 0.298 | -0.016 | 0.002 | 5.22e-15 |
| rs12131864 | 1 | 8591162 | G | A | 0.631 | -0.019 | 0.002 | 3.34e-23 |
| rs462579 | 1 | 91511604 | T | A | 0.109 | -0.021 | 0.003 | 2.87e-12 |
| rs34952318 | 20 | 11177055 | A | G | 0.050 | -0.035 | 0.004 | 1.11e-14 |
| rs4411786 | 20 | 1930897 | C | T | 0.265 | -0.045 | 0.002 | 6.39e-98 |
| rs1997797 | 20 | 31387954 | G | C | 0.446 | 0.017 | 0.002 | 4.44e-19 |
| rs7273174 | 20 | 34350306 | C | T | 0.110 | -0.022 | 0.003 | 1.17e-12 |
| rs6072080 | 20 | 39260927 | T | C | 0.585 | 0.020 | 0.002 | 1.77e-24 |
| rs6103270 | 20 | 42015635 | G | A | 0.226 | -0.014 | 0.002 | 1.91e-09 |
| rs968478 | 20 | 47339742 | G | A | 0.397 | 0.015 | 0.002 | 3.11e-14 |
| rs2230604 | 20 | 49196284 | T | C | 0.084 | 0.030 | 0.003 | 3.65e-18 |
| rs6021231 | 20 | 50092287 | C | T | 0.471 | -0.011 | 0.002 | 2.21e-09 |
| rs6063965 | 20 | 52191200 | A | G | 0.114 | 0.023 | 0.003 | 3.46e-14 |
| rs259981 | 20 | 57748369 | A | T | 0.360 | -0.027 | 0.002 | 3.91e-42 |
| rs2738783 | 20 | 62308612 | G | T | 0.790 | -0.016 | 0.002 | 1.24e-10 |
| rs1297265 | 21 | 16817051 | G | A | 0.443 | 0.012 | 0.002 | 7.44e-10 |
| rs150797 | 21 | 43465831 | G | C | 0.447 | -0.014 | 0.002 | 2.19e-12 |
| rs1893592 | 21 | 43855067 | C | A | 0.296 | 0.015 | 0.002 | 2.13e-12 |
| rs721131 | 21 | 47957767 | C | T | 0.320 | -0.019 | 0.002 | 1.36e-20 |
| rs9605047 | 22 | 20002848 | T | G | 0.328 | 0.020 | 0.002 | 2.99e-21 |
| rs5998509 | 22 | 21917479 | T | C | 0.188 | -0.041 | 0.002 | 2.19e-61 |
| rs71318973 | 22 | 24658265 | T | G | 0.241 | -0.015 | 0.002 | 7.29e-12 |
| rs16986308 | 22 | 28717032 | A | G | 0.116 | -0.032 | 0.003 | 9.95e-28 |
| rs714027 | 22 | 30577771 | G | A | 0.552 | -0.035 | 0.002 | 5.96e-75 |
| rs139425 | 22 | 39559742 | C | G | 0.447 | -0.027 | 0.002 | 1.85e-45 |
| rs2899319 | 22 | 39874314 | C | A | 0.619 | 0.012 | 0.002 | 3.13e-09 |
| rs5758452 | 22 | 42144927 | A | G | 0.793 | -0.027 | 0.002 | 9.92e-31 |
| rs62241216 | 22 | 50744821 | G | A | 0.492 | -0.012 | 0.002 | 3.88e-10 |
| rs6755786 | 2 | 103048103 | T | C | 0.773 | 0.027 | 0.002 | 4.2e-32 |
| rs1533298 | 2 | 111616304 | T | C | 0.715 | 0.031 | 0.002 | 1.19e-49 |
| rs1976055 | 2 | 111787853 | A | C | 0.156 | -0.017 | 0.003 | 4.05e-11 |
| rs144942718 | 2 | 112238447 | A | G | 0.075 | -0.072 | 0.004 | 3.47e-73 |
| rs17366643 | 2 | 11344504 | C | T | 0.538 | 0.015 | 0.002 | 1.35e-14 |
| rs4849169 | 2 | 113953657 | C | A | 0.482 | 0.021 | 0.002 | 4.14e-28 |
| rs35002856 | 2 | 12878229 | C | G | 0.373 | 0.012 | 0.002 | 4.03e-10 |
| rs71420836 | 2 | 129060871 | T | C | 0.097 | 0.030 | 0.003 | 1.67e-19 |
| rs62160910 | 2 | 136919140 | C | G | 0.058 | -0.024 | 0.004 | 4.53e-09 |
| rs73961715 | 2 | 143803847 | C | T | 0.128 | -0.049 | 0.003 | 2.79e-67 |
| rs148601577 | 2 | 143847602 | G | A | 0.020 | -0.049 | 0.007 | 3.79e-12 |
| rs34030812 | 2 | 144248905 | C | T | 0.367 | -0.018 | 0.002 | 1.49e-20 |
| rs10173412 | 2 | 161330940 | C | T | 0.201 | -0.039 | 0.002 | 1.45e-62 |
| rs10180049 | 2 | 162993472 | A | G | 0.385 | 0.015 | 0.002 | 5.21e-14 |
| rs13432797 | 2 | 165526587 | C | G | 0.345 | -0.012 | 0.002 | 2.12e-09 |
| rs529002 | 2 | 169703896 | G | T | 0.551 | -0.011 | 0.002 | 4.46e-09 |
| rs11684463 | 2 | 182077535 | T | A | 0.327 | -0.047 | 0.002 | 1.3e-121 |
| rs62180255 | 2 | 182188444 | A | T | 0.441 | 0.034 | 0.002 | 1.81e-72 |
| rs10179705 | 2 | 188315234 | A | G | 0.294 | -0.021 | 0.002 | 1.29e-24 |
| rs6743068 | 2 | 202153920 | G | A | 0.730 | 0.031 | 0.002 | 1.69e-49 |
| rs715 | 2 | 211543055 | C | T | 0.311 | -0.015 | 0.002 | 5.63e-13 |
| rs11897252 | 2 | 216250263 | T | C | 0.234 | -0.014 | 0.002 | 4.69e-10 |
| rs78058190 | 2 | 219699999 | A | G | 0.052 | 0.047 | 0.005 | 1.36e-22 |
| rs978917 | 2 | 223974443 | A | G | 0.615 | 0.018 | 0.002 | 7.38e-20 |
| rs11676298 | 2 | 227291731 | G | C | 0.191 | 0.027 | 0.002 | 2.86e-29 |
| rs34261309 | 2 | 231224638 | C | T | 0.107 | 0.022 | 0.003 | 8.5e-13 |
| rs58106596 | 2 | 232579379 | A | G | 0.226 | -0.024 | 0.002 | 3.67e-25 |
| rs13016839 | 2 | 234357513 | G | A | 0.107 | -0.026 | 0.003 | 1.03e-16 |
| rs55982282 | 2 | 24188128 | A | G | 0.135 | 0.062 | 0.003 | 8.7e-110 |
| rs1260326 | 2 | 27730940 | C | T | 0.602 | -0.024 | 0.002 | 1.44e-35 |
| rs11688289 | 2 | 40698811 | C | T | 0.189 | -0.015 | 0.002 | 6.98e-10 |
| rs149290349 | 2 | 43451957 | A | G | 0.075 | -0.068 | 0.004 | 3.29e-77 |
| rs2542573 | 2 | 54046646 | C | T | 0.331 | -0.015 | 0.002 | 1.52e-13 |
| rs4671238 | 2 | 55324905 | T | C | 0.560 | -0.014 | 0.002 | 1.05e-12 |
| rs12991188 | 2 | 62554797 | G | A | 0.598 | 0.013 | 0.002 | 3.77e-11 |
| rs6546881 | 2 | 74187420 | T | C | 0.657 | 0.013 | 0.002 | 2.17e-10 |
| rs6721663 | 2 | 7615061 | A | G | 0.159 | -0.026 | 0.003 | 9.83e-24 |
| rs7572278 | 2 | 8563029 | A | T | 0.207 | 0.020 | 0.002 | 1.04e-17 |
| rs2100139 | 3 | 108504265 | C | T | 0.658 | 0.015 | 0.002 | 2.32e-14 |
| rs1822534 | 3 | 12266804 | G | A | 0.394 | -0.034 | 0.002 | 7.22e-71 |
| rs62262391 | 3 | 123105119 | T | C | 0.225 | -0.026 | 0.002 | 3.52e-30 |
| rs332507 | 3 | 124435110 | C | T | 0.222 | -0.015 | 0.002 | 3.12e-11 |
| rs11712335 | 3 | 128210550 | C | T | 0.269 | 0.015 | 0.002 | 1.26e-12 |
| rs9835571 | 3 | 136523475 | G | T | 0.690 | 0.022 | 0.002 | 3.49e-27 |
| rs12636926 | 3 | 141660742 | C | T | 0.730 | -0.014 | 0.002 | 1.92e-10 |
| rs721377 | 3 | 14435063 | T | C | 0.102 | 0.020 | 0.003 | 5.23e-10 |
| rs9864216 | 3 | 151007262 | A | G | 0.776 | 0.024 | 0.002 | 1.57e-25 |
| rs76830965 | 3 | 159637678 | A | C | 0.117 | -0.021 | 0.003 | 2.13e-12 |
| rs2291900 | 3 | 170825920 | A | G | 0.291 | 0.017 | 0.002 | 1.49e-15 |
| rs7627511 | 3 | 171178037 | C | G | 0.219 | -0.017 | 0.002 | 2.11e-12 |
| rs2270968 | 3 | 182755209 | G | T | 0.738 | -0.013 | 0.002 | 2.46e-09 |
| rs11926572 | 3 | 183742328 | G | A | 0.375 | 0.013 | 0.002 | 7.34e-11 |
| rs77265382 | 3 | 185546150 | T | C | 0.045 | -0.033 | 0.005 | 1.17e-12 |
| rs11505019 | 3 | 188094327 | G | T | 0.411 | -0.013 | 0.002 | 3.18e-12 |
| rs6808949 | 3 | 18883999 | C | T | 0.111 | 0.023 | 0.003 | 9.16e-14 |
| rs56850780 | 3 | 195926731 | T | G | 0.078 | -0.021 | 0.004 | 3.55e-09 |
| rs2089979 | 3 | 196501413 | G | A | 0.414 | -0.035 | 0.002 | 4.1e-72 |
| rs2887944 | 3 | 27758275 | T | G | 0.436 | 0.019 | 0.002 | 1.13e-28 |
| rs74280151 | 3 | 28287735 | T | C | 0.175 | 0.029 | 0.002 | 2.22e-38 |
| rs11706384 | 3 | 39296881 | T | G | 0.236 | 0.027 | 0.002 | 7.44e-34 |
| rs73081851 | 3 | 56623617 | T | A | 0.530 | 0.016 | 0.002 | 2.48e-17 |
| rs12485738 | 3 | 56865776 | G | A | 0.628 | 0.018 | 0.002 | 5.99e-20 |
| rs35592432 | 3 | 71355240 | C | G | 0.027 | 0.109 | 0.006 | 1.68e-68 |
| rs6809697 | 3 | 71414864 | A | G | 0.136 | -0.018 | 0.003 | 7.75e-11 |
| rs5026470 | 4 | 103554722 | G | A | 0.517 | 0.037 | 0.002 | 1.45e-84 |
| rs2237040 | 4 | 110838862 | T | C | 0.406 | -0.020 | 0.002 | 3.53e-24 |
| rs7672879 | 4 | 122811172 | A | T | 0.353 | 0.013 | 0.002 | 1.08e-11 |
| rs309375 | 4 | 123681156 | A | C | 0.575 | 0.019 | 0.002 | 7.84e-23 |
| rs7696969 | 4 | 143326714 | G | T | 0.620 | -0.012 | 0.002 | 7.36e-10 |
| rs4696314 | 4 | 153005716 | C | T | 0.515 | 0.014 | 0.002 | 9.33e-14 |
| rs74408817 | 4 | 157801752 | A | C | 0.081 | -0.023 | 0.003 | 2.1e-11 |
| rs62334097 | 4 | 169301245 | G | A | 0.113 | -0.019 | 0.003 | 3.38e-10 |
| rs1344603 | 4 | 38109847 | C | T | 0.676 | 0.029 | 0.002 | 1.19e-47 |
| rs6531629 | 4 | 38358752 | C | T | 0.703 | 0.064 | 0.002 | 1e-200 |
| rs17581460 | 4 | 38456404 | C | T | 0.020 | -0.040 | 0.007 | 3.76e-09 |
| rs13105987 | 4 | 38602467 | A | G | 0.274 | 0.046 | 0.002 | 3.64e-103 |
| rs73142294 | 4 | 40301624 | T | C | 0.248 | 0.015 | 0.002 | 7.51e-12 |
| rs6554158 | 4 | 55066077 | A | G | 0.231 | 0.014 | 0.002 | 2.34e-10 |
| rs58408429 | 4 | 57769824 | C | T | 0.186 | -0.024 | 0.002 | 2.42e-22 |
| rs12640189 | 4 | 6893181 | T | C | 0.176 | 0.016 | 0.003 | 3.4e-10 |
| rs2298850 | 4 | 72614267 | C | G | 0.278 | -0.014 | 0.002 | 1.67e-11 |
| rs10027415 | 4 | 80898027 | T | C | 0.529 | -0.016 | 0.001 | 6.33e-33 |
| rs17005891 | 4 | 83547862 | A | G | 0.184 | -0.024 | 0.002 | 4.43e-43 |
| rs1431545 | 4 | 90220659 | T | C | 0.653 | 0.021 | 0.002 | 5.05e-25 |
| rs2548256 | 5 | 100148462 | T | G | 0.620 | 0.014 | 0.002 | 6.49e-13 |
| rs78270096 | 5 | 110517419 | A | G | 0.070 | -0.034 | 0.004 | 7.79e-19 |
| rs3797727 | 5 | 111079024 | A | T | 0.112 | 0.020 | 0.003 | 4.94e-11 |
| rs73239280 | 5 | 118695579 | A | G | 0.203 | -0.032 | 0.002 | 4.72e-41 |
| rs58265751 | 5 | 124377841 | C | T | 0.521 | -0.012 | 0.002 | 1.61e-10 |
| rs251391 | 5 | 127538116 | T | C | 0.747 | 0.020 | 0.002 | 1.21e-19 |
| rs10076701 | 5 | 131596419 | C | T | 0.557 | -0.023 | 0.002 | 5.52e-33 |
| rs75925066 | 5 | 134722833 | A | C | 0.023 | 0.085 | 0.006 | 1.84e-40 |
| rs1990933 | 5 | 147935890 | G | A | 0.475 | -0.012 | 0.002 | 4.86e-10 |
| rs3815725 | 5 | 150518532 | G | A | 0.520 | -0.017 | 0.002 | 3.66e-18 |
| rs11134475 | 5 | 156399950 | G | A | 0.636 | -0.014 | 0.002 | 4.42e-13 |
| rs17055818 | 5 | 157969475 | C | T | 0.252 | -0.030 | 0.002 | 2.15e-42 |
| rs55977949 | 5 | 158375669 | A | G | 0.098 | -0.030 | 0.003 | 1.15e-21 |
| rs360017 | 5 | 173207353 | G | A | 0.775 | -0.021 | 0.002 | 1.69e-20 |
| rs4976642 | 5 | 176673068 | A | T | 0.831 | 0.024 | 0.003 | 3.44e-22 |
| rs11742873 | 5 | 179244302 | T | C | 0.837 | -0.017 | 0.003 | 1.75e-10 |
| rs464609 | 5 | 34654477 | A | G | 0.544 | 0.012 | 0.002 | 5.3e-10 |
| rs11567701 | 5 | 35859863 | T | G | 0.262 | 0.054 | 0.002 | 2.4e-141 |
| rs2548496 | 5 | 52073844 | G | A | 0.208 | 0.019 | 0.002 | 3.34e-16 |
| rs2910580 | 5 | 57527886 | A | T | 0.659 | -0.014 | 0.002 | 3.57e-12 |
| rs251398 | 5 | 67509801 | C | T | 0.447 | -0.017 | 0.002 | 2.39e-19 |
| rs678393 | 5 | 72418309 | G | T | 0.262 | -0.015 | 0.002 | 2.32e-12 |
| rs160044 | 5 | 88114476 | C | T | 0.308 | 0.012 | 0.002 | 4.72e-09 |
| rs504117 | 6 | 10518350 | A | G | 0.522 | -0.013 | 0.002 | 1.34e-11 |
| rs12526696 | 6 | 108053364 | A | G | 0.161 | 0.037 | 0.003 | 6.06e-44 |
| rs6926219 | 6 | 122720806 | A | G | 0.547 | 0.027 | 0.002 | 2.23e-45 |
| rs1415701 | 6 | 130345835 | A | G | 0.267 | 0.014 | 0.002 | 9.71e-11 |
| rs72986491 | 6 | 135042596 | A | G | 0.206 | 0.017 | 0.002 | 1.28e-12 |
| rs7776054 | 6 | 135418916 | G | A | 0.261 | -0.040 | 0.002 | 9.81e-78 |
| rs17710008 | 6 | 153043035 | A | G | 0.182 | 0.021 | 0.002 | 4.45e-17 |
| rs707827 | 6 | 15327500 | A | G | 0.629 | 0.015 | 0.002 | 9.97e-15 |
| rs41267765 | 6 | 159462424 | T | C | 0.022 | -0.046 | 0.006 | 1.24e-12 |
| rs2273215 | 6 | 170586082 | A | G | 0.458 | -0.015 | 0.002 | 5.04e-14 |
| rs73735249 | 6 | 21809342 | A | G | 0.208 | 0.018 | 0.002 | 2.74e-14 |
| rs72835315 | 6 | 231878 | C | T | 0.060 | 0.035 | 0.004 | 7.09e-17 |
| rs2106072 | 6 | 30153363 | A | G | 0.127 | -0.087 | 0.003 | 1e-200 |
| rs28360986 | 6 | 30994248 | G | C | 0.095 | 0.051 | 0.003 | 3.1e-55 |
| rs9461975 | 6 | 34236459 | T | C | 0.113 | -0.031 | 0.003 | 4.24e-24 |
| rs1050979 | 6 | 410417 | G | A | 0.525 | 0.025 | 0.002 | 3.97e-38 |
| rs4711790 | 6 | 44573822 | T | C | 0.340 | -0.017 | 0.002 | 4.88e-18 |
| rs12190136 | 6 | 44767072 | A | G | 0.526 | 0.012 | 0.002 | 3.99e-10 |
| rs2294861 | 6 | 53136516 | T | C | 0.294 | 0.019 | 0.002 | 3.84e-20 |
| rs9328393 | 6 | 6902939 | A | C | 0.668 | 0.023 | 0.002 | 5.74e-31 |
| rs607465 | 6 | 7129563 | G | A | 0.066 | 0.034 | 0.004 | 4.83e-19 |
| rs1334577 | 6 | 7211751 | A | G | 0.235 | -0.017 | 0.002 | 5.29e-14 |
| rs72928038 | 6 | 90976768 | A | G | 0.177 | -0.033 | 0.002 | 4.24e-41 |
| rs78976959 | 7 | 100311666 | A | G | 0.041 | 0.033 | 0.005 | 8.41e-12 |
| rs4730737 | 7 | 116076443 | A | C | 0.920 | 0.023 | 0.004 | 9.41e-11 |
| rs9920 | 7 | 116200092 | C | T | 0.105 | 0.029 | 0.003 | 2.15e-21 |
| rs62621812 | 7 | 127015083 | A | G | 0.021 | 0.056 | 0.007 | 2.73e-16 |
| rs2727487 | 7 | 129513820 | C | A | 0.629 | -0.013 | 0.002 | 1.08e-10 |
| rs4336553 | 7 | 130743720 | G | A | 0.793 | -0.022 | 0.002 | 1.61e-21 |
| rs10269987 | 7 | 138741642 | A | G | 0.575 | -0.017 | 0.002 | 4.83e-19 |
| rs35114152 | 7 | 150294476 | G | A | 0.226 | 0.025 | 0.002 | 1.73e-27 |
| rs73165514 | 7 | 150527651 | A | G | 0.086 | 0.021 | 0.003 | 5.47e-10 |
| rs35345753 | 7 | 22740513 | G | C | 0.207 | -0.027 | 0.002 | 6.95e-30 |
| rs35121828 | 7 | 23634985 | A | G | 0.259 | 0.013 | 0.002 | 1.22e-09 |
| rs10807805 | 7 | 2753850 | G | A | 0.584 | -0.012 | 0.002 | 4.01e-10 |
| rs9648346 | 7 | 28160113 | G | C | 0.203 | -0.024 | 0.002 | 2.63e-25 |
| rs6955702 | 7 | 3157702 | G | A | 0.522 | -0.022 | 0.002 | 3.36e-31 |
| rs2710804 | 7 | 36084529 | C | T | 0.376 | 0.014 | 0.002 | 1.63e-12 |
| rs6966893 | 7 | 37385357 | A | G | 0.179 | 0.019 | 0.002 | 3.91e-14 |
| rs3735485 | 7 | 45009341 | G | A | 0.845 | 0.044 | 0.003 | 3.56e-63 |
| rs10245472 | 7 | 55147478 | A | G | 0.177 | -0.016 | 0.002 | 1.65e-10 |
| rs2908425 | 7 | 5569327 | A | G | 0.332 | 0.014 | 0.002 | 3.78e-12 |
| rs10237524 | 7 | 56036024 | A | G | 0.330 | -0.019 | 0.002 | 1.35e-21 |
| rs2347784 | 7 | 6524843 | G | C | 0.268 | 0.028 | 0.002 | 6.95e-39 |
| rs4718976 | 7 | 70113409 | C | T | 0.695 | -0.018 | 0.002 | 1.1e-17 |
| rs799166 | 7 | 73051932 | G | C | 0.133 | -0.017 | 0.003 | 7.57e-10 |
| rs10230506 | 7 | 92471518 | A | T | 0.161 | 0.021 | 0.003 | 2.86e-16 |
| rs11984234 | 7 | 949067 | A | T | 0.214 | 0.017 | 0.002 | 2.12e-12 |
| rs57098456 | 7 | 98038351 | G | A | 0.322 | -0.013 | 0.002 | 9.15e-10 |
| rs881492 | 8 | 101484638 | G | C | 0.467 | -0.019 | 0.002 | 6.56e-23 |
| rs1264593 | 8 | 102471269 | T | G | 0.495 | 0.015 | 0.002 | 2.15e-14 |
| rs28588745 | 8 | 10647044 | T | A | 0.206 | 0.022 | 0.002 | 1.25e-21 |
| rs13276352 | 8 | 11657838 | C | T | 0.209 | 0.014 | 0.002 | 2.15e-09 |
| rs2055101 | 8 | 119886923 | T | C | 0.527 | -0.013 | 0.002 | 1.84e-12 |
| rs2980888 | 8 | 126507308 | C | T | 0.697 | -0.015 | 0.002 | 1.21e-13 |
| rs10956401 | 8 | 129002419 | A | G | 0.345 | -0.025 | 0.002 | 5.05e-37 |
| rs10107362 | 8 | 130986984 | G | C | 0.361 | 0.013 | 0.002 | 8.36e-11 |
| rs7007986 | 8 | 142217115 | A | G | 0.601 | -0.022 | 0.002 | 8.5e-28 |
| rs7830045 | 8 | 18700282 | C | A | 0.436 | -0.013 | 0.002 | 1.69e-11 |
| rs56094005 | 8 | 21769432 | G | A | 0.043 | -0.040 | 0.005 | 1.61e-17 |
| rs1533065 | 8 | 22785768 | T | C | 0.345 | 0.017 | 0.002 | 4.95e-17 |
| rs113142693 | 8 | 30242076 | C | T | 0.247 | -0.017 | 0.002 | 1.46e-14 |
| rs2980821 | 8 | 40028306 | A | C | 0.425 | -0.013 | 0.002 | 4.97e-12 |
| rs6474359 | 8 | 41549194 | C | T | 0.038 | -0.051 | 0.005 | 1.13e-24 |
| rs4737010 | 8 | 41630447 | A | G | 0.229 | 0.027 | 0.002 | 2.47e-32 |
| rs10503147 | 8 | 421777 | T | C | 0.089 | 0.021 | 0.003 | 7.48e-10 |
| rs7839516 | 8 | 61386533 | C | T | 0.369 | 0.015 | 0.002 | 3.08e-15 |
| rs10109370 | 8 | 72909349 | A | C | 0.238 | -0.019 | 0.002 | 2.26e-17 |
| rs117934175 | 8 | 79069520 | A | G | 0.041 | -0.045 | 0.005 | 1.12e-19 |
| rs1441850 | 8 | 79657666 | C | T | 0.252 | 0.050 | 0.002 | 9.8e-117 |
| rs7855586 | 9 | 112678121 | G | A | 0.290 | 0.015 | 0.002 | 1.03e-12 |
| rs1002607 | 9 | 113858933 | C | T | 0.593 | -0.013 | 0.002 | 1.93e-11 |
| rs3761846 | 9 | 123689597 | T | C | 0.570 | 0.021 | 0.002 | 6.26e-27 |
| rs10985915 | 9 | 126018715 | G | A | 0.111 | -0.023 | 0.003 | 6.9e-14 |
| rs4880192 | 9 | 139927062 | G | A | 0.721 | 0.029 | 0.002 | 1.16e-40 |
| rs12336678 | 9 | 21785894 | T | C | 0.099 | -0.022 | 0.003 | 6.82e-12 |
| rs3731211 | 9 | 21986847 | A | T | 0.720 | 0.044 | 0.002 | 5.14e-95 |
| rs2065500 | 9 | 22145694 | G | A | 0.160 | -0.048 | 0.003 | 3.29e-77 |
| rs1971429 | 9 | 33776182 | A | C | 0.729 | 0.013 | 0.002 | 2.01e-09 |
| rs10814138 | 9 | 34723035 | A | C | 0.314 | -0.014 | 0.002 | 2.55e-11 |
| rs11541908 | 9 | 35705759 | A | G | 0.277 | 0.020 | 0.002 | 4.93e-22 |
| rs10973700 | 9 | 38196117 | C | G | 0.487 | -0.019 | 0.002 | 1.11e-23 |
| rs501461 | 9 | 4039727 | T | G | 0.600 | -0.018 | 0.002 | 6.76e-20 |
| rs2807303 | 9 | 82187095 | A | G | 0.346 | 0.022 | 0.002 | 2.79e-27 |
| rs62565782 | 9 | 86600887 | A | G | 0.131 | -0.022 | 0.003 | 3.75e-14 |
| rs61750929 | 9 | 91495135 | T | C | 0.056 | -0.070 | 0.004 | 1.36e-65 |
| rs3739756 | 9 | 98775235 | C | T | 0.036 | -0.032 | 0.005 | 2.88e-10 |
| rs74911261 | 11 | 108357137 | G | A | 0.974 | -0.045 | 0.006 | 8.68e-14 |
| rs4407213 | 2 | 25541636 | G | C | 0.968 | -0.042 | 0.006 | 1.49e-14 |
| rs140763487 | 3 | 112686528 | C | T | 0.986 | 0.049 | 0.008 | 9.26e-10 |
| rs146946728 | 6 | 32490363 | A | C | 0.935 | -0.039 | 0.006 | 5.08e-10 |
| rs149984502 | 7 | 50539036 | C | T | 0.981 | -0.048 | 0.007 | 1.18e-11 |
| rs75653581 | 8 | 78363128 | C | T | 0.987 | -0.061 | 0.009 | 5.45e-12 |
| rs3824667 | 10 | 8100125 | G | A | 0.827 | 0.027 | 0.003 | 1.56e-24 |
| rs28416859 | 11 | 196606 | C | A | 0.376 | -0.019 | 0.002 | 3.44e-15 |
| rs7933244 | 11 | 60267695 | C | T | 0.525 | 0.017 | 0.002 | 7.19e-19 |
| rs7113428 | 11 | 65770230 | A | G | 0.340 | 0.013 | 0.002 | 5.89e-11 |
| rs373582220 | 12 | 112847812 | A | G | 0.372 | -0.035 | 0.003 | 4.06e-40 |
| rs9513573 | 13 | 99815898 | A | G | 0.279 | -0.015 | 0.002 | 2.73e-12 |
| rs11863592 | 16 | 31301011 | G | A | 0.269 | -0.016 | 0.002 | 2.21e-13 |
| rs2667640 | 16 | 78575335 | T | C | 0.674 | 0.014 | 0.002 | 7.39e-12 |
| rs77017480 | 17 | 2064712 | A | T | 0.100 | -0.039 | 0.003 | 1.88e-32 |
| rs578102530 | 17 | 44364335 | A | G | 0.187 | -0.016 | 0.003 | 5.33e-10 |
| rs28841391 | 19 | 1193965 | G | T | 0.758 | 0.029 | 0.003 | 3.03e-31 |
| rs8101897 | 19 | 16410422 | G | A | 0.419 | -0.062 | 0.002 | 1e-200 |
| rs77232119 | 19 | 300952 | G | T | 0.273 | 0.016 | 0.002 | 1.22e-11 |
| rs12741113 | 1 | 101147149 | C | T | 0.449 | -0.018 | 0.002 | 2.26e-19 |
| rs377345022 | 1 | 101653213 | T | A | 0.265 | -0.018 | 0.002 | 2.97e-15 |
| rs327200 | 1 | 120550701 | G | C | 0.106 | 0.021 | 0.003 | 3.83e-11 |
| rs60918921 | 1 | 23850590 | A | C | 0.619 | -0.015 | 0.002 | 1.39e-12 |
| rs4068540 | 1 | 40398941 | G | A | 0.266 | 0.017 | 0.002 | 1.52e-14 |
| rs1883932 | 20 | 8609588 | T | A | 0.509 | -0.027 | 0.002 | 1.91e-44 |
| rs5995837 | 22 | 40603363 | C | G | 0.270 | 0.015 | 0.002 | 5.15e-12 |
| rs199898421 | 2 | 112193161 | G | A | 0.060 | 0.038 | 0.006 | 1.59e-10 |
| rs4673265 | 2 | 204631738 | T | C | 0.731 | -0.016 | 0.002 | 4.15e-13 |
| rs7625643 | 3 | 141150026 | G | A | 0.444 | 0.025 | 0.002 | 2.36e-36 |
| rs13063578 | 3 | 47087837 | A | T | 0.401 | 0.053 | 0.002 | 1.24e-142 |
| rs62332762 | 4 | 106143492 | T | C | 0.396 | -0.026 | 0.002 | 1.17e-39 |
| rs62638606 | 5 | 124234974 | A | T | 0.504 | -0.012 | 0.002 | 3.74e-09 |
| rs373718455 | 5 | 16634559 | A | G | 0.249 | -0.017 | 0.002 | 1.44e-13 |
| rs2408014 | 5 | 54864017 | T | A | 0.681 | -0.015 | 0.002 | 3.58e-13 |
| rs61076041 | 6 | 109741530 | G | C | 0.298 | -0.015 | 0.002 | 1.52e-11 |
| rs7763994 | 6 | 151912354 | T | A | 0.291 | 0.014 | 0.002 | 6.72e-11 |
| rs565245327 | 6 | 31614543 | T | G | 0.471 | -0.050 | 0.003 | 5.89e-84 |
| rs111959275 | 6 | 32618796 | T | G | 0.142 | 0.085 | 0.003 | 2.82e-168 |
| rs55633929 | 7 | 156595998 | A | G | 0.425 | -0.014 | 0.002 | 8.67e-12 |
| rs192364644 | 7 | 77163413 | C | G | 0.612 | -0.013 | 0.002 | 3.49e-11 |
| rs200984644 | 7 | 98931658 | G | A | 0.064 | 0.040 | 0.004 | 3.73e-23 |
| rs1976451 | 8 | 49456791 | G | T | 0.135 | -0.017 | 0.003 | 4.35e-09 |
| rs13277751 | 8 | 59415337 | G | T | 0.642 | -0.012 | 0.002 | 1.14e-09 |
| rs2791733 | 9 | 133623966 | A | C | 0.400 | -0.015 | 0.002 | 4.38e-14 |
| rs77866737 | 10 | 6075285 | D | I | 0.200 | -0.029 | 0.003 | 3.65e-29 |
| rs538811162 | 2 | 24613039 | I | D | 0.580 | -0.017 | 0.002 | 9.31e-15 |
| rs9494100 | 6 | 135195857 | I | D | 0.253 | -0.018 | 0.002 | 8.97e-14 |
| rs74835819 | 6 | 32455787 | A | G | 0.071 | 0.046 | 0.005 | 2.74e-20 |

**Table S13. 464 SNPs significantly associated with myeloid white cell count used IVs in forward MR analyses derived from Chen MH et al.** Chr: Chromosome; EA: Effect allele; NEA: Non-effect allele; EAF: Effect allele frequency.

| SNP | Chr | Pos | EA | NEA | EAF | Beta | SE | P-value |
| --- | --- | --- | --- | --- | --- | --- | --- | --- |
| rs7917772 | 10 | 104487443 | A | G | 0.631 | 0.017 | 0.002 | 3.6e-18 |
| rs72836628 | 10 | 113902454 | T | C | 0.249 | -0.014 | 0.002 | 3.02e-11 |
| rs180941 | 10 | 115720674 | A | G | 0.625 | -0.017 | 0.002 | 1.63e-18 |
| rs12784071 | 10 | 115854445 | T | C | 0.222 | 0.018 | 0.002 | 4.79e-16 |
| rs7920891 | 10 | 120895131 | A | C | 0.569 | -0.012 | 0.002 | 1.35e-10 |
| rs3781454 | 10 | 126348565 | A | G | 0.677 | 0.025 | 0.002 | 3.68e-36 |
| rs10906393 | 10 | 13536512 | T | A | 0.576 | 0.015 | 0.002 | 2.44e-16 |
| rs692594 | 10 | 18265893 | C | G | 0.489 | 0.013 | 0.002 | 1.7e-12 |
| rs10828725 | 10 | 25218243 | T | G | 0.365 | -0.050 | 0.002 | 1.1e-148 |
| rs703005 | 10 | 26758576 | T | C | 0.598 | -0.015 | 0.002 | 4.55e-16 |
| rs2993986 | 10 | 28800023 | T | C | 0.774 | 0.030 | 0.002 | 4.75e-43 |
| rs2091084 | 10 | 30492267 | C | T | 0.653 | 0.017 | 0.002 | 6.25e-19 |
| rs11252331 | 10 | 4129296 | G | T | 0.837 | 0.017 | 0.002 | 1.8e-11 |
| rs72790862 | 10 | 44880260 | C | T | 0.307 | -0.026 | 0.002 | 5.05e-38 |
| rs17011726 | 10 | 50264204 | G | C | 0.233 | -0.024 | 0.002 | 3.58e-27 |
| rs41295055 | 10 | 6111622 | T | C | 0.187 | -0.014 | 0.002 | 4.95e-09 |
| rs7082470 | 10 | 65277026 | A | G | 0.474 | -0.029 | 0.002 | 5.75e-56 |
| rs1885474 | 10 | 69566751 | G | T | 0.103 | -0.022 | 0.003 | 1.11e-12 |
| rs3747869 | 10 | 73520632 | C | A | 0.900 | 0.039 | 0.003 | 1.79e-33 |
| rs11002309 | 10 | 79594931 | T | C | 0.340 | 0.013 | 0.002 | 8.28e-11 |
| rs1880390 | 10 | 88334654 | A | C | 0.292 | -0.012 | 0.002 | 1.18e-09 |
| rs59085061 | 10 | 89681458 | G | A | 0.040 | -0.050 | 0.005 | 9e-27 |
| rs9633675 | 10 | 96210400 | C | G | 0.479 | -0.019 | 0.002 | 9.6e-25 |
| rs61863767 | 10 | 99084426 | T | C | 0.596 | 0.043 | 0.002 | 4.56e-117 |
| rs12363256 | 11 | 100520680 | A | T | 0.325 | 0.016 | 0.002 | 4.54e-16 |
| rs11225017 | 11 | 101687730 | T | C | 0.084 | 0.020 | 0.003 | 1.72e-09 |
| rs4909932 | 11 | 10475967 | G | A | 0.598 | 0.014 | 0.002 | 2.01e-13 |
| rs7934719 | 11 | 108341864 | T | C | 0.414 | 0.018 | 0.002 | 1.55e-21 |
| rs73000965 | 11 | 113982321 | A | T | 0.315 | 0.029 | 0.002 | 1.86e-47 |
| rs672058 | 11 | 116764021 | T | C | 0.880 | 0.018 | 0.003 | 1.81e-10 |
| rs148713124 | 11 | 118225200 | T | G | 0.009 | 0.096 | 0.010 | 1.35e-21 |
| rs11022177 | 11 | 12133478 | G | C | 0.311 | -0.018 | 0.002 | 3.43e-19 |
| rs8705 | 11 | 128328913 | A | G | 0.317 | -0.026 | 0.002 | 1.35e-39 |
| rs10831912 | 11 | 12856414 | C | T | 0.604 | -0.011 | 0.002 | 1.69e-09 |
| rs12792460 | 11 | 18076439 | G | C | 0.271 | 0.016 | 0.002 | 4.13e-14 |
| rs1468102 | 11 | 3004526 | G | C | 0.306 | 0.015 | 0.002 | 9.95e-14 |
| rs1232050 | 11 | 30798288 | C | G | 0.430 | -0.012 | 0.002 | 2.48e-09 |
| rs934177 | 11 | 44612714 | G | C | 0.436 | -0.011 | 0.002 | 1.48e-09 |
| rs1228024 | 11 | 47951353 | A | C | 0.661 | 0.025 | 0.002 | 2.32e-38 |
| rs579721 | 11 | 59834318 | A | G | 0.284 | -0.012 | 0.002 | 1.55e-09 |
| rs174548 | 11 | 61571348 | G | C | 0.314 | -0.025 | 0.002 | 6.38e-36 |
| rs617791 | 11 | 65702523 | C | G | 0.483 | 0.019 | 0.002 | 1.59e-24 |
| rs7177 | 11 | 69466115 | A | C | 0.529 | -0.013 | 0.002 | 1.43e-12 |
| rs17041439 | 12 | 101873240 | C | A | 0.057 | 0.038 | 0.004 | 2.96e-21 |
| rs73190675 | 12 | 109788891 | A | G | 0.050 | 0.029 | 0.004 | 1.15e-11 |
| rs10774624 | 12 | 111833788 | A | G | 0.514 | -0.065 | 0.002 | 1e-200 |
| rs11064881 | 12 | 120146925 | A | G | 0.074 | -0.032 | 0.004 | 9.92e-20 |
| rs7308123 | 12 | 122225225 | C | T | 0.181 | 0.019 | 0.002 | 2.29e-14 |
| rs9697459 | 12 | 123988435 | T | C | 0.225 | 0.014 | 0.002 | 4e-10 |
| rs9863 | 12 | 124421453 | C | T | 0.329 | -0.019 | 0.002 | 1.53e-21 |
| rs2024385 | 12 | 12888438 | A | T | 0.427 | 0.017 | 0.002 | 1.2e-19 |
| rs11048425 | 12 | 26339986 | G | C | 0.491 | 0.013 | 0.002 | 1.03e-12 |
| rs1007938 | 12 | 26802549 | G | A | 0.405 | -0.011 | 0.002 | 3.26e-09 |
| rs12580347 | 12 | 3388932 | C | T | 0.583 | -0.012 | 0.002 | 3.28e-10 |
| rs7955734 | 12 | 4333159 | G | C | 0.210 | -0.029 | 0.002 | 2.1e-37 |
| rs73108566 | 12 | 49080698 | A | T | 0.331 | -0.013 | 0.002 | 1.32e-10 |
| rs2302900 | 12 | 50599709 | C | T | 0.341 | 0.019 | 0.002 | 3.69e-22 |
| rs1700159 | 12 | 52305786 | T | C | 0.774 | 0.024 | 0.002 | 6.3e-28 |
| rs1245035 | 12 | 64976049 | A | C | 0.627 | 0.014 | 0.002 | 4.79e-14 |
| rs2286599 | 12 | 6499533 | A | G | 0.141 | 0.039 | 0.003 | 1.43e-43 |
| rs7968902 | 12 | 66363070 | G | T | 0.571 | 0.011 | 0.002 | 1.65e-09 |
| rs759488 | 12 | 68587155 | C | T | 0.705 | 0.014 | 0.002 | 2.58e-11 |
| rs4761234 | 12 | 69732105 | C | T | 0.482 | 0.015 | 0.002 | 2.03e-16 |
| rs11114149 | 12 | 79908743 | T | A | 0.683 | -0.014 | 0.002 | 7.3e-12 |
| rs11104881 | 12 | 88843474 | C | T | 0.702 | -0.025 | 0.002 | 1.59e-34 |
| rs2265146 | 13 | 114178222 | A | T | 0.281 | 0.022 | 0.002 | 4.66e-28 |
| rs3118899 | 13 | 114919294 | A | G | 0.357 | 0.014 | 0.002 | 2.6e-13 |
| rs76428106 | 13 | 28604007 | C | T | 0.013 | 0.158 | 0.009 | 3.45e-77 |
| rs138028125 | 13 | 28712689 | G | C | 0.034 | 0.054 | 0.006 | 2.54e-22 |
| rs9508005 | 13 | 28789794 | G | T | 0.094 | -0.023 | 0.003 | 4.92e-11 |
| rs7326825 | 13 | 50113450 | A | G | 0.704 | 0.021 | 0.002 | 1.73e-24 |
| rs806316 | 13 | 50838230 | G | C | 0.532 | 0.016 | 0.002 | 4.61e-18 |
| rs9526795 | 13 | 52341181 | C | T | 0.259 | -0.014 | 0.002 | 2.68e-11 |
| rs3812849 | 13 | 74701736 | C | A | 0.267 | 0.020 | 0.002 | 5.85e-21 |
| rs2439963 | 13 | 78638416 | G | A | 0.737 | -0.012 | 0.002 | 3.29e-09 |
| rs12429714 | 13 | 99649831 | T | C | 0.424 | -0.011 | 0.002 | 2.45e-09 |
| rs10146962 | 14 | 101170540 | C | T | 0.338 | -0.015 | 0.002 | 6.45e-15 |
| rs12898000 | 14 | 103835128 | C | G | 0.701 | 0.019 | 0.002 | 7.85e-20 |
| rs2494748 | 14 | 105258892 | T | C | 0.616 | 0.013 | 0.002 | 6.09e-11 |
| rs7159281 | 14 | 23756604 | G | A | 0.672 | -0.013 | 0.002 | 1.35e-10 |
| rs2038700 | 14 | 25461989 | C | T | 0.395 | 0.034 | 0.002 | 4.61e-74 |
| rs72664840 | 14 | 35596323 | T | C | 0.179 | 0.021 | 0.002 | 8.44e-18 |
| rs696 | 14 | 35871093 | T | C | 0.366 | 0.015 | 0.002 | 4.53e-15 |
| rs9323285 | 14 | 55863829 | A | C | 0.338 | 0.012 | 0.002 | 8.19e-10 |
| rs10138752 | 14 | 69179971 | T | C | 0.077 | -0.040 | 0.003 | 1.85e-31 |
| rs4903580 | 14 | 77850978 | T | C | 0.456 | 0.016 | 0.002 | 2.93e-18 |
| rs2241621 | 14 | 81737076 | C | A | 0.588 | -0.015 | 0.002 | 7.34e-16 |
| rs10498635 | 14 | 93103309 | T | C | 0.183 | -0.031 | 0.002 | 6.58e-39 |
| rs2180369 | 14 | 93516465 | C | T | 0.111 | 0.023 | 0.003 | 7.65e-14 |
| rs28576226 | 15 | 101713668 | A | G | 0.122 | 0.031 | 0.003 | 6.14e-28 |
| rs4924450 | 15 | 40597229 | A | G | 0.706 | 0.014 | 0.002 | 1.14e-10 |
| rs72726027 | 15 | 42248826 | C | T | 0.112 | -0.043 | 0.003 | 4.17e-48 |
| rs60695341 | 15 | 51010271 | T | C | 0.197 | -0.030 | 0.002 | 3.2e-38 |
| rs780142 | 15 | 62797964 | G | T | 0.274 | 0.015 | 0.002 | 2.02e-12 |
| rs2062250 | 15 | 64672002 | A | G | 0.938 | 0.051 | 0.004 | 2.12e-38 |
| rs2469147 | 15 | 66910224 | G | A | 0.158 | -0.015 | 0.003 | 1.67e-09 |
| rs62007171 | 15 | 77306285 | C | T | 0.291 | -0.015 | 0.002 | 6.22e-14 |
| rs11854390 | 15 | 80224817 | T | C | 0.562 | 0.014 | 0.002 | 2.13e-13 |
| rs7183988 | 15 | 91428589 | G | T | 0.527 | -0.016 | 0.002 | 8.74e-17 |
| rs4984768 | 16 | 1253907 | T | G | 0.363 | 0.012 | 0.002 | 3.34e-09 |
| rs9926183 | 16 | 1363878 | T | C | 0.263 | 0.019 | 0.002 | 2.86e-17 |
| rs12929950 | 16 | 2144894 | A | G | 0.077 | -0.030 | 0.004 | 7.83e-17 |
| rs61739285 | 16 | 27480797 | T | C | 0.033 | -0.034 | 0.005 | 9.98e-11 |
| rs11574938 | 16 | 30485393 | C | G | 0.521 | 0.038 | 0.002 | 8.13e-91 |
| rs28853644 | 16 | 30801027 | T | C | 0.272 | -0.018 | 0.002 | 1.64e-17 |
| rs9925985 | 16 | 31305593 | C | A | 0.269 | -0.014 | 0.002 | 1.09e-11 |
| rs6500550 | 16 | 3746241 | T | C | 0.304 | -0.021 | 0.002 | 2.65e-24 |
| rs75859969 | 16 | 50160737 | C | A | 0.156 | -0.017 | 0.003 | 5.94e-11 |
| rs11644125 | 16 | 57058974 | T | C | 0.597 | -0.018 | 0.002 | 6.36e-21 |
| rs12918121 | 16 | 67187795 | T | C | 0.018 | -0.051 | 0.007 | 1.08e-12 |
| rs117556162 | 16 | 67680806 | A | G | 0.057 | 0.027 | 0.004 | 1.99e-11 |
| rs7198940 | 16 | 81605559 | T | C | 0.522 | 0.016 | 0.002 | 3.33e-17 |
| rs247826 | 16 | 84582965 | T | C | 0.221 | 0.034 | 0.002 | 9.59e-52 |
| rs9933582 | 16 | 86016026 | G | T | 0.230 | 0.021 | 0.002 | 9.18e-22 |
| rs17232826 | 16 | 89842784 | T | C | 0.102 | -0.022 | 0.003 | 1.6e-12 |
| rs6502721 | 17 | 1362503 | T | C | 0.678 | 0.013 | 0.002 | 1.28e-10 |
| rs12936529 | 17 | 16168784 | T | C | 0.479 | -0.025 | 0.002 | 2.74e-42 |
| rs146207734 | 17 | 16472850 | T | A | 0.017 | -0.054 | 0.008 | 4.58e-11 |
| rs4925108 | 17 | 17649423 | T | C | 0.615 | -0.017 | 0.002 | 2.67e-18 |
| rs7225843 | 17 | 2001825 | C | T | 0.203 | -0.032 | 0.002 | 8.58e-43 |
| rs756818 | 17 | 2711248 | A | G | 0.172 | 0.017 | 0.002 | 2.01e-12 |
| rs16964963 | 17 | 27603316 | G | A | 0.173 | 0.022 | 0.002 | 2.54e-19 |
| rs9900730 | 17 | 28179066 | G | A | 0.504 | 0.023 | 0.002 | 1.57e-35 |
| rs1034686 | 17 | 35899141 | A | T | 0.584 | -0.013 | 0.002 | 4.87e-13 |
| rs12946510 | 17 | 37912377 | T | C | 0.472 | -0.059 | 0.002 | 1e-200 |
| rs74725931 | 17 | 38196327 | C | T | 0.037 | 0.061 | 0.005 | 3.65e-33 |
| rs111975659 | 17 | 38570168 | T | C | 0.027 | -0.036 | 0.006 | 1.34e-09 |
| rs34855406 | 17 | 40731411 | C | G | 0.275 | 0.016 | 0.002 | 5.42e-15 |
| rs35592645 | 17 | 4616387 | A | C | 0.249 | 0.017 | 0.002 | 2.38e-16 |
| rs4790752 | 17 | 5115877 | T | C | 0.451 | -0.018 | 0.002 | 2.11e-21 |
| rs56378716 | 17 | 56356502 | G | A | 0.013 | 0.108 | 0.008 | 1.7e-38 |
| rs2665405 | 17 | 57875292 | A | G | 0.548 | 0.035 | 0.002 | 5.63e-82 |
| rs2084312 | 17 | 72695211 | T | C | 0.800 | 0.029 | 0.002 | 7.67e-36 |
| rs9900613 | 17 | 74674857 | T | C | 0.431 | -0.011 | 0.002 | 1.3e-09 |
| rs8079218 | 17 | 76247744 | G | A | 0.497 | -0.015 | 0.002 | 1.11e-14 |
| rs9747839 | 17 | 81084522 | G | C | 0.476 | 0.019 | 0.002 | 4.46e-18 |
| rs11874453 | 18 | 20750400 | A | G | 0.533 | -0.012 | 0.002 | 1.36e-09 |
| rs303753 | 18 | 21074922 | A | G | 0.345 | -0.020 | 0.002 | 1.78e-23 |
| rs78285907 | 18 | 21622003 | T | A | 0.089 | 0.020 | 0.003 | 3.1e-09 |
| rs75354229 | 18 | 42078782 | T | C | 0.129 | 0.024 | 0.003 | 6.22e-19 |
| rs8084255 | 18 | 48144754 | T | C | 0.376 | 0.020 | 0.002 | 1.26e-25 |
| rs527544 | 18 | 51762866 | T | G | 0.427 | 0.014 | 0.002 | 8.21e-13 |
| rs17758695 | 18 | 60920854 | T | C | 0.029 | -0.056 | 0.006 | 1.67e-22 |
| rs12956324 | 18 | 67537270 | A | C | 0.403 | 0.014 | 0.002 | 2.1e-13 |
| rs72973711 | 18 | 74072245 | T | A | 0.067 | -0.032 | 0.004 | 1.28e-17 |
| rs7246841 | 19 | 1030431 | C | T | 0.731 | 0.022 | 0.002 | 3.78e-24 |
| rs2358581 | 19 | 10391611 | G | T | 0.733 | -0.028 | 0.002 | 9.36e-38 |
| rs56121005 | 19 | 11414706 | T | C | 0.023 | 0.040 | 0.007 | 3.03e-09 |
| rs2290669 | 19 | 16495774 | C | A | 0.786 | -0.048 | 0.002 | 1.52e-94 |
| rs4808683 | 19 | 17862925 | G | C | 0.458 | -0.013 | 0.002 | 8.23e-12 |
| rs4805881 | 19 | 33896432 | C | A | 0.665 | -0.016 | 0.002 | 1.32e-16 |
| rs2194067 | 19 | 35464727 | T | C | 0.416 | 0.013 | 0.002 | 2.31e-11 |
| rs4760 | 19 | 44153100 | G | A | 0.153 | -0.070 | 0.003 | 2.21e-161 |
| rs73036517 | 19 | 45744842 | G | A | 0.259 | -0.053 | 0.002 | 1.08e-137 |
| rs309190 | 19 | 47629264 | C | T | 0.894 | -0.030 | 0.003 | 7.05e-22 |
| rs2617802 | 19 | 49114479 | G | T | 0.207 | -0.015 | 0.002 | 1.59e-10 |
| rs35112940 | 19 | 51738917 | A | G | 0.216 | -0.026 | 0.002 | 5.21e-31 |
| rs571497 | 19 | 7827830 | A | G | 0.155 | -0.036 | 0.003 | 6.32e-45 |
| rs34202160 | 1 | 101102673 | G | T | 0.358 | -0.015 | 0.002 | 8.34e-15 |
| rs6696259 | 1 | 101221482 | C | G | 0.409 | 0.018 | 0.002 | 3.74e-22 |
| rs284317 | 1 | 10731625 | G | A | 0.498 | 0.017 | 0.002 | 3.52e-19 |
| rs9429767 | 1 | 110496087 | A | G | 0.197 | 0.017 | 0.002 | 1.03e-13 |
| rs630505 | 1 | 111737916 | C | T | 0.265 | 0.012 | 0.002 | 3.13e-09 |
| rs2476601 | 1 | 114377568 | G | A | 0.900 | 0.053 | 0.003 | 3.35e-67 |
| rs4970996 | 1 | 150506589 | C | G | 0.751 | 0.015 | 0.002 | 8.97e-13 |
| rs568035 | 1 | 156110167 | T | C | 0.067 | -0.025 | 0.004 | 9.21e-12 |
| rs34599082 | 1 | 159175494 | T | C | 0.013 | -0.151 | 0.008 | 1.93e-77 |
| rs11587213 | 1 | 161184875 | G | A | 0.182 | -0.020 | 0.002 | 7.7e-17 |
| rs61804160 | 1 | 161620089 | A | T | 0.140 | -0.030 | 0.003 | 8.08e-29 |
| rs12737843 | 1 | 167010375 | C | T | 0.202 | 0.014 | 0.002 | 5.1e-10 |
| rs3795503 | 1 | 180905694 | T | C | 0.315 | -0.015 | 0.002 | 1.83e-14 |
| rs1779809 | 1 | 182145545 | T | C | 0.334 | 0.014 | 0.002 | 1.36e-13 |
| rs41272536 | 1 | 183440531 | G | A | 0.046 | -0.034 | 0.005 | 3.65e-13 |
| rs10494783 | 1 | 198663661 | A | G | 0.052 | -0.044 | 0.004 | 4.43e-26 |
| rs4844565 | 1 | 207227138 | T | A | 0.410 | -0.011 | 0.002 | 1.01e-09 |
| rs4844390 | 1 | 207934849 | G | A | 0.220 | -0.027 | 0.002 | 4.97e-33 |
| rs35571080 | 1 | 224637790 | C | T | 0.213 | 0.025 | 0.002 | 1.32e-28 |
| rs2615061 | 1 | 225895806 | A | G | 0.117 | -0.028 | 0.003 | 3.82e-22 |
| rs6426584 | 1 | 227374949 | T | A | 0.335 | -0.012 | 0.002 | 3.66e-09 |
| rs533483 | 1 | 234765256 | A | G | 0.243 | -0.017 | 0.002 | 1.91e-15 |
| rs1886654 | 1 | 236105910 | C | T | 0.892 | -0.057 | 0.003 | 5.08e-84 |
| rs6684709 | 1 | 23850023 | C | G | 0.620 | -0.021 | 0.002 | 1.84e-27 |
| rs34298354 | 1 | 247588053 | T | C | 0.124 | -0.024 | 0.003 | 5.9e-18 |
| rs56188865 | 1 | 247606276 | C | T | 0.374 | -0.027 | 0.002 | 7e-46 |
| rs4925756 | 1 | 248048016 | T | C | 0.692 | -0.015 | 0.002 | 3.83e-13 |
| rs35755865 | 1 | 27140060 | T | C | 0.034 | -0.041 | 0.005 | 3.18e-16 |
| rs7549164 | 1 | 31224193 | T | C | 0.189 | 0.025 | 0.002 | 1.33e-24 |
| rs3917932 | 1 | 36943916 | G | C | 0.577 | -0.045 | 0.002 | 1.22e-127 |
| rs6667709 | 1 | 42201943 | T | C | 0.370 | -0.014 | 0.002 | 6.3e-13 |
| rs3754224 | 1 | 43423622 | C | T | 0.271 | -0.015 | 0.002 | 4.3e-12 |
| rs74076327 | 1 | 56979431 | C | T | 0.045 | -0.029 | 0.004 | 1.55e-10 |
| rs1933295 | 1 | 62107021 | G | A | 0.777 | -0.016 | 0.002 | 1.81e-11 |
| rs10889574 | 1 | 66149341 | A | G | 0.357 | -0.044 | 0.002 | 6.64e-106 |
| rs61780038 | 1 | 67487376 | A | C | 0.565 | -0.017 | 0.002 | 2.99e-16 |
| rs41313381 | 1 | 79411968 | A | C | 0.030 | 0.048 | 0.006 | 1.47e-18 |
| rs301819 | 1 | 8501786 | G | A | 0.583 | -0.016 | 0.002 | 3.12e-17 |
| rs10864368 | 1 | 8918313 | C | T | 0.503 | 0.022 | 0.002 | 1.15e-31 |
| rs12138789 | 1 | 9192307 | C | T | 0.238 | -0.015 | 0.002 | 2.08e-11 |
| rs150649461 | 1 | 92925654 | C | G | 0.015 | 0.068 | 0.008 | 1.33e-17 |
| rs9430574 | 1 | 9709072 | A | G | 0.330 | -0.014 | 0.002 | 1.55e-12 |
| rs34952318 | 20 | 11177055 | A | G | 0.050 | -0.033 | 0.004 | 4.24e-14 |
| rs6045615 | 20 | 1931582 | C | A | 0.330 | -0.019 | 0.002 | 1.99e-22 |
| rs4815552 | 20 | 2833578 | G | A | 0.351 | 0.012 | 0.002 | 7.61e-10 |
| rs6059938 | 20 | 33187130 | A | G | 0.486 | 0.011 | 0.002 | 4.74e-09 |
| rs67139609 | 20 | 36888321 | G | C | 0.145 | 0.017 | 0.003 | 2.47e-10 |
| rs6029234 | 20 | 39259278 | C | G | 0.624 | 0.024 | 0.002 | 7.57e-37 |
| rs763227 | 20 | 42205061 | A | G | 0.786 | 0.014 | 0.002 | 1.93e-09 |
| rs2904270 | 20 | 49185272 | A | G | 0.517 | -0.015 | 0.002 | 1.03e-16 |
| rs2327028 | 20 | 8140925 | T | C | 0.476 | -0.013 | 0.002 | 3.36e-13 |
| rs4432538 | 20 | 8607393 | A | G | 0.509 | -0.022 | 0.002 | 2.22e-33 |
| rs28574812 | 21 | 16384940 | G | A | 0.158 | -0.016 | 0.003 | 2.66e-10 |
| rs9977672 | 21 | 40463283 | A | G | 0.259 | -0.022 | 0.002 | 7.08e-25 |
| rs35990176 | 21 | 44472118 | C | A | 0.451 | 0.012 | 0.002 | 6.46e-10 |
| rs2839166 | 21 | 47656712 | C | T | 0.398 | -0.016 | 0.002 | 2.97e-17 |
| rs5746451 | 22 | 18126020 | C | T | 0.504 | 0.020 | 0.002 | 4.61e-26 |
| rs448184 | 22 | 18293967 | T | A | 0.368 | 0.015 | 0.002 | 1.61e-13 |
| rs34505104 | 22 | 24624609 | G | A | 0.305 | -0.020 | 0.002 | 3.36e-23 |
| rs9625746 | 22 | 29637658 | C | G | 0.412 | -0.018 | 0.002 | 8.77e-22 |
| rs2857633 | 22 | 30114333 | C | T | 0.727 | -0.013 | 0.002 | 2.52e-10 |
| rs2180142 | 22 | 31855248 | A | T | 0.051 | -0.025 | 0.004 | 3.17e-09 |
| rs713909 | 22 | 39532420 | C | G | 0.432 | -0.020 | 0.002 | 2.75e-26 |
| rs1033415 | 22 | 39888774 | G | A | 0.650 | 0.013 | 0.002 | 6.01e-10 |
| rs47341 | 22 | 43560763 | T | C | 0.396 | 0.016 | 0.002 | 1.01e-15 |
| rs2076211 | 22 | 44329078 | T | C | 0.158 | -0.018 | 0.003 | 1.53e-12 |
| rs796056 | 2 | 101735276 | C | A | 0.623 | 0.021 | 0.002 | 2.31e-27 |
| rs6543094 | 2 | 102372131 | C | T | 0.772 | 0.019 | 0.002 | 1.86e-17 |
| rs75726191 | 2 | 112914684 | T | C | 0.136 | 0.024 | 0.003 | 1.22e-16 |
| rs6734238 | 2 | 113841030 | G | A | 0.402 | 0.036 | 0.002 | 2.59e-77 |
| rs11688303 | 2 | 128410244 | T | C | 0.120 | 0.021 | 0.003 | 3.19e-13 |
| rs1863176 | 2 | 12931092 | C | G | 0.497 | 0.019 | 0.002 | 6.27e-24 |
| rs79716587 | 2 | 143886819 | A | G | 0.124 | -0.036 | 0.003 | 9.22e-37 |
| rs1371045 | 2 | 145787050 | C | T | 0.744 | -0.014 | 0.002 | 1.26e-11 |
| rs114427331 | 2 | 153381036 | G | C | 0.038 | -0.036 | 0.005 | 6.36e-14 |
| rs2729707 | 2 | 160687231 | G | A | 0.830 | -0.031 | 0.002 | 1.28e-36 |
| rs6746368 | 2 | 161222998 | T | A | 0.334 | 0.011 | 0.002 | 4.36e-09 |
| rs2068330 | 2 | 163237390 | G | C | 0.361 | 0.019 | 0.002 | 2.87e-22 |
| rs4632345 | 2 | 16702654 | A | G | 0.678 | -0.014 | 0.002 | 3.75e-12 |
| rs527393 | 2 | 169709757 | A | G | 0.468 | -0.022 | 0.002 | 4.34e-33 |
| rs17270882 | 2 | 182218046 | C | G | 0.239 | 0.031 | 0.002 | 4.07e-46 |
| rs7573465 | 2 | 182315885 | T | G | 0.554 | 0.036 | 0.002 | 4.66e-83 |
| rs8176528 | 2 | 188345017 | A | G | 0.297 | -0.017 | 0.002 | 6.31e-17 |
| rs12997100 | 2 | 192522423 | C | T | 0.071 | 0.027 | 0.004 | 9.38e-14 |
| rs10931934 | 2 | 202119789 | C | T | 0.600 | 0.015 | 0.002 | 1.38e-15 |
| rs1047891 | 2 | 211540507 | A | C | 0.315 | -0.021 | 0.002 | 1.6e-26 |
| rs10203838 | 2 | 213935900 | C | T | 0.418 | 0.012 | 0.002 | 4.66e-11 |
| rs1250215 | 2 | 216253039 | C | G | 0.641 | 0.012 | 0.002 | 1.08e-09 |
| rs114050631 | 2 | 219020958 | T | C | 0.011 | -0.138 | 0.010 | 1.38e-45 |
| rs58106596 | 2 | 232579379 | A | G | 0.227 | -0.024 | 0.002 | 1.85e-25 |
| rs10164769 | 2 | 237779229 | T | C | 0.743 | 0.027 | 0.002 | 7.73e-38 |
| rs1260326 | 2 | 27730940 | C | T | 0.601 | -0.033 | 0.002 | 9.83e-69 |
| rs4666068 | 2 | 28623982 | G | A | 0.509 | -0.012 | 0.002 | 1.15e-10 |
| rs633323 | 2 | 31463044 | T | A | 0.698 | 0.015 | 0.002 | 2.87e-13 |
| rs935655 | 2 | 46067445 | G | T | 0.762 | 0.015 | 0.002 | 8.11e-12 |
| rs75475627 | 2 | 54787592 | G | C | 0.077 | 0.033 | 0.004 | 1.92e-20 |
| rs10208769 | 2 | 61605614 | A | T | 0.516 | 0.022 | 0.002 | 6.48e-33 |
| rs6731993 | 2 | 65642097 | T | A | 0.408 | 0.023 | 0.002 | 3.7e-33 |
| rs60190483 | 2 | 70385245 | C | T | 0.057 | -0.031 | 0.004 | 2.43e-15 |
| rs7639292 | 3 | 107295665 | T | C | 0.168 | -0.024 | 0.002 | 2.48e-22 |
| rs13321783 | 3 | 119615375 | C | T | 0.394 | -0.012 | 0.002 | 1.58e-09 |
| rs1822534 | 3 | 12266804 | G | A | 0.394 | -0.026 | 0.002 | 8.29e-43 |
| rs3773312 | 3 | 12946142 | A | G | 0.134 | 0.018 | 0.003 | 3.67e-10 |
| rs78208917 | 3 | 132229708 | G | A | 0.090 | 0.026 | 0.003 | 7.98e-16 |
| rs2012610 | 3 | 140933565 | T | G | 0.127 | 0.028 | 0.003 | 2.99e-23 |
| rs9819371 | 3 | 141206800 | T | C | 0.065 | -0.044 | 0.004 | 5.94e-32 |
| rs73028871 | 3 | 14277058 | T | C | 0.134 | -0.017 | 0.003 | 9.3e-10 |
| rs6440732 | 3 | 150990510 | A | C | 0.828 | 0.023 | 0.002 | 1.35e-21 |
| rs56082403 | 3 | 156797225 | C | T | 0.405 | 0.015 | 0.002 | 5.5e-15 |
| rs10936588 | 3 | 169319801 | A | G | 0.636 | -0.014 | 0.002 | 6.95e-14 |
| rs4074672 | 3 | 183730295 | T | C | 0.370 | 0.018 | 0.002 | 3.87e-21 |
| rs9867398 | 3 | 185912816 | T | C | 0.094 | 0.022 | 0.003 | 2.8e-12 |
| rs2268829 | 3 | 185989567 | A | G | 0.785 | -0.014 | 0.002 | 2.66e-10 |
| rs9829114 | 3 | 196518623 | A | G | 0.414 | -0.034 | 0.002 | 7.45e-72 |
| rs11927257 | 3 | 27334294 | T | C | 0.282 | -0.013 | 0.002 | 2.97e-10 |
| rs2371108 | 3 | 27757018 | T | G | 0.389 | 0.019 | 0.002 | 3.11e-24 |
| rs11706384 | 3 | 39296881 | T | G | 0.236 | 0.016 | 0.002 | 1.72e-13 |
| rs1366045 | 3 | 42909050 | C | T | 0.384 | -0.025 | 0.002 | 5.44e-39 |
| rs116781404 | 3 | 48730203 | T | C | 0.043 | 0.028 | 0.005 | 1.46e-09 |
| rs6779340 | 3 | 58033701 | G | C | 0.337 | -0.018 | 0.002 | 4.51e-19 |
| rs832190 | 3 | 63842629 | T | C | 0.633 | -0.013 | 0.002 | 3.88e-12 |
| rs35592432 | 3 | 71355240 | C | G | 0.027 | 0.040 | 0.006 | 5.47e-11 |
| rs830623 | 3 | 71677746 | C | A | 0.170 | 0.014 | 0.002 | 3.88e-09 |
| rs4504118 | 3 | 98422588 | C | G | 0.559 | 0.013 | 0.002 | 6.34e-12 |
| rs113473633 | 4 | 103449131 | G | A | 0.026 | -0.051 | 0.006 | 3.24e-16 |
| rs144317085 | 4 | 105806108 | T | A | 0.034 | 0.050 | 0.005 | 3e-22 |
| rs10006495 | 4 | 120263315 | G | T | 0.354 | -0.016 | 0.002 | 1.35e-15 |
| rs11735662 | 4 | 145026126 | T | C | 0.033 | 0.052 | 0.005 | 4.74e-24 |
| rs2305980 | 4 | 151171269 | A | T | 0.255 | 0.014 | 0.002 | 6.67e-11 |
| rs6854855 | 4 | 152247161 | G | A | 0.487 | 0.016 | 0.002 | 9.69e-19 |
| rs140311179 | 4 | 26295540 | G | C | 0.051 | -0.026 | 0.004 | 1.08e-09 |
| rs73191188 | 4 | 3105200 | A | G | 0.353 | -0.016 | 0.002 | 1.23e-17 |
| rs28530750 | 4 | 36312542 | A | G | 0.043 | 0.056 | 0.005 | 1.83e-33 |
| rs987121 | 4 | 38329783 | T | A | 0.606 | 0.015 | 0.002 | 4.76e-16 |
| rs13132853 | 4 | 38680015 | G | A | 0.355 | 0.020 | 0.002 | 1.88e-25 |
| rs218264 | 4 | 55408875 | T | A | 0.251 | 0.026 | 0.002 | 2.52e-33 |
| rs6554195 | 4 | 55500714 | T | G | 0.484 | -0.019 | 0.002 | 3.9e-26 |
| rs28695311 | 4 | 55992105 | C | G | 0.506 | -0.011 | 0.002 | 6.36e-10 |
| rs2412771 | 4 | 57761417 | C | T | 0.417 | -0.018 | 0.002 | 4.42e-22 |
| rs6831368 | 4 | 6969919 | G | A | 0.362 | -0.020 | 0.002 | 2.02e-25 |
| rs35734242 | 4 | 706700 | C | T | 0.428 | 0.022 | 0.002 | 6.3e-29 |
| rs16846876 | 4 | 72592491 | T | A | 0.329 | -0.021 | 0.002 | 7.39e-27 |
| rs16850073 | 4 | 74703999 | T | C | 0.374 | 0.043 | 0.002 | 8.76e-109 |
| rs16850408 | 4 | 74932807 | A | C | 0.369 | 0.059 | 0.002 | 1e-200 |
| rs4640621 | 4 | 80946541 | C | A | 0.632 | -0.013 | 0.002 | 2.01e-11 |
| rs116746683 | 4 | 83471084 | T | C | 0.063 | -0.030 | 0.004 | 7.86e-15 |
| rs10028284 | 4 | 89752913 | T | A | 0.177 | -0.015 | 0.002 | 3.25e-10 |
| rs28378092 | 4 | 90228807 | T | C | 0.458 | 0.012 | 0.002 | 6.64e-10 |
| rs62387565 | 5 | 100024839 | T | C | 0.484 | 0.014 | 0.002 | 1.1e-14 |
| rs4535497 | 5 | 1107428 | A | C | 0.570 | -0.013 | 0.002 | 2.34e-11 |
| rs79692389 | 5 | 111033606 | C | G | 0.061 | 0.026 | 0.004 | 9.39e-12 |
| rs257063 | 5 | 114806819 | T | C | 0.741 | -0.014 | 0.002 | 4.91e-11 |
| rs79272926 | 5 | 118726637 | A | C | 0.222 | -0.022 | 0.002 | 1.04e-23 |
| rs6878780 | 5 | 122093740 | C | T | 0.427 | 0.013 | 0.002 | 1.02e-12 |
| rs7705526 | 5 | 1285974 | A | C | 0.327 | 0.029 | 0.002 | 8.3e-43 |
| rs11242109 | 5 | 131677047 | T | G | 0.477 | -0.033 | 0.002 | 5.08e-72 |
| rs79237520 | 5 | 134712566 | T | C | 0.023 | 0.040 | 0.006 | 2.13e-10 |
| rs4391200 | 5 | 141509537 | G | A | 0.619 | 0.023 | 0.002 | 7.86e-34 |
| rs72660299 | 5 | 150564634 | T | C | 0.063 | -0.024 | 0.004 | 7.4e-11 |
| rs4704826 | 5 | 156392082 | A | C | 0.634 | -0.016 | 0.002 | 9.71e-17 |
| rs9313822 | 5 | 159207694 | A | G | 0.052 | 0.025 | 0.004 | 9.35e-10 |
| rs10054235 | 5 | 173087427 | C | A | 0.349 | 0.013 | 0.002 | 4.9e-11 |
| rs2561758 | 5 | 173205282 | G | A | 0.723 | -0.036 | 0.002 | 4.2e-68 |
| rs13180726 | 5 | 179126457 | A | G | 0.826 | -0.021 | 0.002 | 2.71e-18 |
| rs464609 | 5 | 34654477 | A | G | 0.544 | 0.013 | 0.002 | 3e-12 |
| rs987107 | 5 | 35875227 | A | G | 0.261 | 0.020 | 0.002 | 2.52e-22 |
| rs11744663 | 5 | 57315635 | A | G | 0.190 | -0.019 | 0.002 | 1.72e-15 |
| rs2910580 | 5 | 57527886 | A | T | 0.659 | -0.015 | 0.002 | 2.83e-15 |
| rs4703881 | 5 | 71741851 | A | G | 0.879 | -0.036 | 0.003 | 7.28e-38 |
| rs2432142 | 5 | 96275201 | A | G | 0.434 | 0.013 | 0.002 | 1.09e-11 |
| rs1087924 | 6 | 10523826 | C | G | 0.531 | -0.014 | 0.002 | 4.66e-14 |
| rs9487043 | 6 | 109610474 | T | C | 0.487 | 0.023 | 0.002 | 4.35e-36 |
| rs10872223 | 6 | 121814703 | T | C | 0.183 | 0.014 | 0.002 | 3.57e-09 |
| rs549302 | 6 | 122734972 | A | G | 0.545 | 0.020 | 0.002 | 8.48e-28 |
| rs9375447 | 6 | 126874091 | G | A | 0.495 | -0.014 | 0.002 | 6.34e-14 |
| rs7776054 | 6 | 135418916 | G | A | 0.261 | -0.047 | 0.002 | 2.48e-110 |
| rs12214269 | 6 | 135846518 | A | G | 0.546 | 0.018 | 0.002 | 5.06e-23 |
| rs6924387 | 6 | 137082948 | G | A | 0.412 | 0.017 | 0.002 | 1.48e-18 |
| rs72978754 | 6 | 138053203 | C | T | 0.066 | -0.028 | 0.004 | 2.45e-14 |
| rs9390461 | 6 | 147701217 | G | A | 0.538 | 0.015 | 0.002 | 8.51e-17 |
| rs7751717 | 6 | 153005229 | A | G | 0.200 | -0.014 | 0.002 | 2.23e-09 |
| rs1738074 | 6 | 159465977 | C | T | 0.568 | -0.020 | 0.002 | 3.74e-28 |
| rs1322599 | 6 | 16758425 | T | C | 0.174 | -0.031 | 0.002 | 9.97e-36 |
| rs73735248 | 6 | 21809195 | T | C | 0.159 | 0.015 | 0.003 | 9.08e-10 |
| rs75628831 | 6 | 22316185 | A | G | 0.086 | 0.031 | 0.003 | 4.98e-20 |
| rs707910 | 6 | 29910663 | A | G | 0.187 | -0.073 | 0.002 | 1e-200 |
| rs114892859 | 6 | 31369278 | T | G | 0.019 | -0.085 | 0.007 | 1.92e-35 |
| rs9394299 | 6 | 35449326 | T | C | 0.104 | -0.028 | 0.003 | 1.21e-19 |
| rs12203592 | 6 | 396321 | T | C | 0.210 | 0.022 | 0.002 | 1.61e-21 |
| rs10948036 | 6 | 42510305 | A | C | 0.210 | 0.023 | 0.002 | 1.97e-24 |
| rs17713864 | 6 | 43695465 | G | A | 0.029 | 0.035 | 0.005 | 1.54e-10 |
| rs998584 | 6 | 43757896 | A | C | 0.484 | 0.016 | 0.002 | 9.24e-17 |
| rs729761 | 6 | 43804571 | G | T | 0.713 | 0.015 | 0.002 | 3.38e-13 |
| rs639728 | 6 | 44239922 | T | C | 0.441 | -0.012 | 0.002 | 5.99e-10 |
| rs1285886 | 6 | 7140831 | A | G | 0.197 | 0.026 | 0.002 | 5.1e-30 |
| rs4707609 | 6 | 90946479 | C | T | 0.360 | -0.021 | 0.002 | 3.21e-28 |
| rs10808139 | 7 | 104709946 | A | G | 0.119 | 0.018 | 0.003 | 6.06e-10 |
| rs342294 | 7 | 106372622 | C | T | 0.456 | -0.017 | 0.002 | 4.9e-21 |
| rs38859 | 7 | 116379094 | T | C | 0.449 | 0.011 | 0.002 | 9.71e-10 |
| rs9656395 | 7 | 130575884 | G | A | 0.095 | -0.028 | 0.003 | 2.97e-18 |
| rs7803075 | 7 | 130742066 | G | A | 0.734 | -0.024 | 0.002 | 5.47e-31 |
| rs13231262 | 7 | 148886058 | T | C | 0.656 | -0.016 | 0.002 | 3.99e-16 |
| rs4721659 | 7 | 17869147 | G | T | 0.123 | -0.018 | 0.003 | 5.3e-11 |
| rs7776857 | 7 | 22754768 | T | G | 0.658 | -0.019 | 0.002 | 1.89e-21 |
| rs7792934 | 7 | 2393016 | G | A | 0.789 | -0.015 | 0.002 | 4.69e-11 |
| rs798555 | 7 | 2759473 | C | T | 0.295 | 0.019 | 0.002 | 4.81e-20 |
| rs2158799 | 7 | 28277107 | G | C | 0.612 | 0.043 | 0.002 | 7.86e-114 |
| rs56388170 | 7 | 28724374 | T | G | 0.295 | 0.059 | 0.002 | 2.73e-185 |
| rs2710804 | 7 | 36084529 | C | T | 0.376 | 0.021 | 0.002 | 1.92e-27 |
| rs10260281 | 7 | 38270550 | C | G | 0.237 | 0.013 | 0.002 | 5.33e-10 |
| rs3735485 | 7 | 45009341 | G | A | 0.845 | 0.041 | 0.003 | 2.34e-57 |
| rs2692540 | 7 | 47465325 | G | C | 0.428 | -0.011 | 0.002 | 1.19e-09 |
| rs6583435 | 7 | 50293405 | A | C | 0.680 | 0.017 | 0.002 | 8.7e-18 |
| rs4948097 | 7 | 56056571 | G | A | 0.758 | -0.018 | 0.002 | 3.03e-16 |
| rs12540307 | 7 | 65662767 | T | C | 0.049 | -0.039 | 0.004 | 4.76e-18 |
| rs1474419 | 7 | 6692605 | C | T | 0.580 | 0.018 | 0.002 | 3.11e-22 |
| rs62466318 | 7 | 73042085 | T | C | 0.202 | -0.021 | 0.002 | 9.8e-19 |
| rs7781268 | 7 | 75748183 | A | G | 0.246 | -0.013 | 0.002 | 3.66e-09 |
| rs60466842 | 7 | 8024840 | A | C | 0.088 | -0.030 | 0.003 | 1.04e-19 |
| rs4272 | 7 | 92236829 | G | A | 0.213 | 0.035 | 0.002 | 2.41e-54 |
| rs445 | 7 | 92408370 | T | C | 0.099 | -0.095 | 0.003 | 1e-200 |
| rs12538820 | 7 | 99340049 | A | G | 0.113 | -0.019 | 0.003 | 1.62e-10 |
| rs77912140 | 8 | 100824826 | G | A | 0.044 | 0.027 | 0.004 | 1.66e-09 |
| rs3847147 | 8 | 105784723 | A | G | 0.307 | -0.012 | 0.002 | 2.99e-09 |
| rs17209630 | 8 | 106296090 | A | G | 0.281 | 0.014 | 0.002 | 3.25e-12 |
| rs11250076 | 8 | 10647823 | G | A | 0.575 | 0.021 | 0.002 | 2.44e-30 |
| rs4734879 | 8 | 106583124 | G | A | 0.276 | -0.022 | 0.002 | 1.34e-26 |
| rs71510325 | 8 | 126298640 | T | C | 0.597 | 0.014 | 0.002 | 3.34e-13 |
| rs10087240 | 8 | 129012574 | T | C | 0.458 | 0.020 | 0.002 | 6.13e-27 |
| rs145209947 | 8 | 130694185 | A | C | 0.014 | -0.075 | 0.009 | 3.19e-18 |
| rs7018391 | 8 | 131243506 | C | G | 0.140 | 0.016 | 0.003 | 3.26e-09 |
| rs7005996 | 8 | 142241681 | T | C | 0.906 | 0.024 | 0.003 | 4.06e-13 |
| rs6985508 | 8 | 142337734 | A | G | 0.358 | -0.027 | 0.002 | 1.57e-39 |
| rs12550612 | 8 | 22966769 | A | G | 0.821 | -0.033 | 0.002 | 1.09e-41 |
| rs755951 | 8 | 27226790 | C | A | 0.403 | -0.013 | 0.002 | 1.02e-12 |
| rs13248936 | 8 | 30316162 | G | A | 0.498 | 0.013 | 0.002 | 1.17e-12 |
| rs6474514 | 8 | 38835269 | C | T | 0.412 | 0.011 | 0.002 | 3.59e-09 |
| rs28571765 | 8 | 55449281 | C | T | 0.207 | -0.022 | 0.002 | 1.32e-20 |
| rs1511724 | 8 | 56796697 | G | C | 0.422 | -0.017 | 0.002 | 1.15e-18 |
| rs2875974 | 8 | 61419728 | A | G | 0.373 | 0.021 | 0.002 | 3.54e-29 |
| rs7846314 | 8 | 61650831 | T | A | 0.187 | 0.047 | 0.002 | 5.71e-88 |
| rs4276676 | 8 | 68795618 | C | T | 0.469 | -0.019 | 0.002 | 2.22e-25 |
| rs12716647 | 8 | 6901304 | C | G | 0.638 | 0.012 | 0.002 | 1.12e-10 |
| rs1947897 | 8 | 78946874 | C | G | 0.336 | -0.014 | 0.002 | 1.71e-13 |
| rs62510269 | 8 | 79012343 | G | A | 0.146 | -0.036 | 0.003 | 1.77e-43 |
| rs10102877 | 8 | 79598362 | A | G | 0.685 | -0.013 | 0.002 | 3.91e-11 |
| rs7864482 | 9 | 102268861 | G | A | 0.377 | 0.013 | 0.002 | 2e-11 |
| rs12683699 | 9 | 112738298 | G | A | 0.160 | 0.022 | 0.003 | 5.16e-18 |
| rs10980797 | 9 | 113912553 | G | A | 0.488 | -0.027 | 0.002 | 1.19e-48 |
| rs7855091 | 9 | 114655098 | A | G | 0.307 | 0.015 | 0.002 | 1.11e-13 |
| rs12343532 | 9 | 115978022 | A | T | 0.272 | -0.015 | 0.002 | 2.49e-13 |
| rs10986338 | 9 | 127191232 | A | G | 0.650 | -0.017 | 0.002 | 1.93e-17 |
| rs2519093 | 9 | 136141870 | T | C | 0.186 | -0.041 | 0.002 | 8.55e-68 |
| rs2506699 | 9 | 136946799 | G | A | 0.484 | 0.014 | 0.002 | 4.87e-14 |
| rs4413892 | 9 | 139330158 | A | G | 0.276 | 0.031 | 0.002 | 2.87e-46 |
| rs41317014 | 9 | 139929080 | T | C | 0.248 | -0.020 | 0.002 | 3.25e-19 |
| rs7036656 | 9 | 21990457 | T | C | 0.722 | 0.028 | 0.002 | 1.25e-42 |
| rs12376511 | 9 | 22142756 | C | T | 0.164 | -0.034 | 0.002 | 6.16e-43 |
| rs34881325 | 9 | 2622134 | T | C | 0.379 | 0.017 | 0.002 | 1.16e-18 |
| rs13291664 | 9 | 282738 | G | A | 0.187 | 0.026 | 0.002 | 1.57e-28 |
| rs3793537 | 9 | 35687556 | C | G | 0.290 | 0.020 | 0.002 | 1.53e-22 |
| rs10973700 | 9 | 38196117 | C | G | 0.486 | -0.013 | 0.002 | 1.36e-12 |
| rs447124 | 9 | 4746539 | T | C | 0.491 | 0.017 | 0.002 | 4.86e-20 |
| rs1887428 | 9 | 4984530 | C | G | 0.625 | -0.015 | 0.002 | 9.58e-15 |
| rs626416 | 9 | 79326680 | C | G | 0.763 | -0.017 | 0.002 | 2.81e-14 |
| rs796003 | 9 | 86595801 | T | G | 0.258 | -0.025 | 0.002 | 8.84e-33 |
| rs4877024 | 9 | 91520860 | G | A | 0.381 | -0.012 | 0.002 | 5.91e-10 |
| rs10992394 | 9 | 95433830 | A | G | 0.230 | 0.015 | 0.002 | 2.14e-12 |
| rs28821528 | 6 | 32528913 | A | G | 0.412 | -0.036 | 0.003 | 3.23e-45 |
| rs4746522 | 10 | 64437684 | A | G | 0.278 | 0.013 | 0.002 | 7.17e-10 |
| rs71468750 | 11 | 120205560 | A | G | 0.413 | 0.014 | 0.002 | 1.39e-11 |
| rs14408 | 11 | 308314 | C | T | 0.368 | 0.044 | 0.002 | 2.97e-93 |
| rs374039502 | 13 | 108960385 | A | T | 0.022 | -0.065 | 0.007 | 2.13e-20 |
| rs576566496 | 17 | 38140269 | A | G | 0.026 | 0.087 | 0.006 | 3.22e-42 |
| rs79135050 | 17 | 38215203 | C | A | 0.030 | 0.068 | 0.006 | 1.44e-28 |
| rs76306941 | 17 | 61944620 | G | T | 0.581 | -0.012 | 0.002 | 1.04e-09 |
| rs55874505 | 18 | 43816804 | G | T | 0.584 | -0.019 | 0.002 | 5.5e-20 |
| rs74176367 | 19 | 1180039 | C | T | 0.355 | 0.014 | 0.002 | 2.58e-10 |
| rs143998157 | 1 | 161561236 | T | C | 0.392 | -0.014 | 0.002 | 2.54e-11 |
| rs6676472 | 1 | 200294113 | T | C | 0.712 | 0.014 | 0.002 | 1.96e-11 |
| rs7554039 | 1 | 22303188 | A | G | 0.243 | 0.016 | 0.002 | 1.15e-12 |
| rs140958972 | 1 | 46428213 | G | A | 0.435 | -0.020 | 0.002 | 6.18e-25 |
| rs12121236 | 1 | 56617911 | A | C | 0.352 | 0.015 | 0.002 | 4.41e-14 |
| rs3016112 | 22 | 17517388 | T | C | 0.750 | -0.018 | 0.002 | 9.57e-16 |
| rs58270357 | 2 | 112236636 | T | C | 0.081 | -0.026 | 0.004 | 4.03e-10 |
| rs377110161 | 2 | 23963462 | G | A | 0.128 | 0.037 | 0.003 | 1.56e-36 |
| rs4599108 | 2 | 85543222 | T | C | 0.488 | -0.016 | 0.002 | 1.31e-16 |
| rs150918492 | 3 | 128389659 | A | G | 0.265 | -0.027 | 0.002 | 9.95e-36 |
| rs13063578 | 3 | 47087837 | A | T | 0.401 | 0.029 | 0.002 | 1.25e-44 |
| rs11130982 | 3 | 64728312 | G | T | 0.700 | 0.012 | 0.002 | 3.53e-09 |
| rs11960649 | 5 | 148203144 | C | A | 0.553 | -0.020 | 0.002 | 3.58e-24 |
| rs13165502 | 5 | 54972181 | A | T | 0.652 | -0.014 | 0.002 | 9.3e-12 |
| rs66514959 | 5 | 68611583 | A | G | 0.444 | -0.022 | 0.002 | 5.66e-30 |
| rs71536939 | 6 | 170486950 | G | A | 0.783 | 0.018 | 0.003 | 2.38e-11 |
| rs76819034 | 6 | 32457814 | C | T | 0.437 | 0.070 | 0.002 | 1e-200 |
| rs573635286 | 6 | 32489556 | T | G | 0.238 | -0.074 | 0.003 | 1.61e-144 |
| rs1761678 | 6 | 7090640 | T | G | 0.871 | -0.020 | 0.003 | 2.25e-11 |
| rs201150852 | 6 | 87837608 | C | T | 0.458 | -0.017 | 0.002 | 3.78e-18 |
| rs28555390 | 7 | 137607217 | T | C | 0.620 | 0.012 | 0.002 | 2.31e-09 |
| rs10104995 | 8 | 103901234 | T | C | 0.231 | -0.022 | 0.002 | 4.74e-21 |
| rs59697075 | 8 | 130618150 | T | C | 0.587 | -0.039 | 0.002 | 1.82e-85 |
| rs10808538 | 8 | 87169063 | A | G | 0.296 | 0.013 | 0.002 | 3.6e-10 |
| rs10815052 | 9 | 4648694 | T | C | 0.453 | -0.012 | 0.002 | 1.28e-09 |
| rs1288649 | 9 | 91426717 | G | T | 0.612 | -0.021 | 0.002 | 7.74e-25 |
| rs6502497 | 17 | 16102530 | D | I | 0.464 | 0.021 | 0.002 | 5.67e-24 |
| rs2409752 | 8 | 11092685 | D | I | 0.535 | 0.017 | 0.002 | 4.18e-16 |

**Table S14. 347 SNPs significantly associated with eosinophil percentage of white cells used IVs in forward MR analyses derived from Vuckovic D et al.** Chr: Chromosome; EA: Effect allele; NEA: Non-effect allele; EAF: Effect allele frequency.

| SNP | Chr | Pos | EA | NEA | EAF | Beta | SE | P-value |
| --- | --- | --- | --- | --- | --- | --- | --- | --- |
| rs159963 | 1 | 8504421 | A | C | 0.583 | 0.025 | 0.002 | 1.4e-30 |
| rs4908832 | 1 | 9338959 | T | C | 0.387 | 0.016 | 0.002 | 1.2e-13 |
| rs57959755 | 1 | 9358890 | A | G | 0.162 | 0.018 | 0.003 | 8.3e-10 |
| rs35249183 | 1 | 12099345 | G | A | 0.100 | 0.038 | 0.004 | 1.8e-25 |
| rs10799794 | 1 | 23701118 | A | C | 0.736 | -0.014 | 0.002 | 4.2e-09 |
| rs2229586 | 1 | 24200903 | C | G | 0.570 | 0.022 | 0.002 | 2.5e-24 |
| rs7416513 | 1 | 26647949 | C | G | 0.762 | -0.017 | 0.002 | 1.5e-11 |
| rs76022305 | 1 | 28631300 | T | C | 0.069 | -0.032 | 0.004 | 2.8e-14 |
| rs1004870 | 1 | 42370787 | T | C | 0.589 | 0.024 | 0.002 | 3.3e-29 |
| rs567910619 | 1 | 65210790 | T | C | 0.011 | -0.080 | 0.011 | 1.5e-13 |
| rs12408934 | 1 | 65423447 | A | G | 0.103 | -0.047 | 0.004 | 9.3e-41 |
| rs2802469 | 1 | 66017386 | T | C | 0.644 | -0.014 | 0.002 | 1.4e-10 |
| rs6672038 | 1 | 87751670 | T | C | 0.119 | 0.039 | 0.003 | 1e-32 |
| rs1088318 | 1 | 93733057 | T | C | 0.379 | -0.021 | 0.002 | 2.1e-22 |
| rs76336747 | 1 | 101648318 | A | G | 0.114 | -0.043 | 0.003 | 2.8e-38 |
| rs2793925 | 1 | 120652158 | A | T | 0.684 | 0.017 | 0.002 | 7.1e-14 |
| rs2596251 | 1 | 145103928 | A | G | 0.388 | 0.020 | 0.003 | 4.6e-09 |
| rs11204682 | 1 | 150595537 | T | G | 0.223 | -0.035 | 0.003 | 2.8e-41 |
| rs111548294 | 1 | 150911337 | T | C | 0.013 | 0.078 | 0.010 | 2.6e-16 |
| rs2242193 | 1 | 154995503 | A | G | 0.420 | 0.015 | 0.002 | 2.1e-11 |
| rs6672031 | 1 | 172796505 | G | C | 0.487 | 0.020 | 0.002 | 4.8e-21 |
| rs1099448 | 1 | 173231898 | T | C | 0.467 | 0.019 | 0.002 | 1.1e-19 |
| rs6658698 | 1 | 181060791 | A | G | 0.618 | -0.015 | 0.002 | 5.5e-12 |
| rs17668272 | 1 | 198623842 | T | G | 0.118 | -0.043 | 0.003 | 2.8e-39 |
| rs1036332 | 1 | 199012478 | C | A | 0.738 | -0.033 | 0.002 | 1.5e-42 |
| rs10900595 | 1 | 204511602 | A | C | 0.713 | 0.016 | 0.002 | 3.9e-12 |
| rs3849269 | 1 | 206676143 | T | C | 0.530 | -0.013 | 0.002 | 2.1e-09 |
| rs59150958 | 1 | 221106304 | T | C | 0.152 | -0.026 | 0.003 | 1.8e-18 |
| rs12142635 | 1 | 226976332 | A | C | 0.057 | -0.030 | 0.005 | 1.3e-10 |
| rs4362021 | 1 | 234696177 | A | T | 0.534 | 0.013 | 0.002 | 4.4e-09 |
| rs4660129 | 1 | 236094087 | G | A | 0.892 | 0.025 | 0.003 | 1.9e-13 |
| rs4149909 | 1 | 242023898 | G | A | 0.033 | -0.040 | 0.006 | 2.9e-11 |
| rs346835 | 2 | 8438693 | T | C | 0.329 | -0.027 | 0.002 | 3.7e-32 |
| rs62105489 | 2 | 8757326 | T | C | 0.053 | -0.030 | 0.005 | 5.6e-10 |
| rs780092 | 2 | 27743154 | G | A | 0.161 | 0.021 | 0.003 | 2e-13 |
| rs11127154 | 2 | 28685967 | T | A | 0.492 | -0.023 | 0.002 | 1.9e-25 |
| rs336039 | 2 | 38258827 | T | C | 0.599 | -0.013 | 0.002 | 2e-09 |
| rs77059113 | 2 | 43672508 | G | T | 0.073 | -0.032 | 0.004 | 1.7e-15 |
| rs11126037 | 2 | 65663158 | A | C | 0.327 | -0.014 | 0.002 | 2.7e-10 |
| rs6546661 | 2 | 71277382 | C | T | 0.304 | -0.017 | 0.002 | 1.7e-13 |
| rs11126424 | 2 | 74262353 | C | T | 0.306 | -0.016 | 0.002 | 1.2e-12 |
| rs2366640 | 2 | 85810734 | C | A | 0.319 | -0.014 | 0.002 | 2.8e-09 |
| rs2579500 | 2 | 97201682 | A | G | 0.390 | 0.050 | 0.002 | 9.7e-116 |
| rs6543250 | 2 | 98640981 | G | A | 0.604 | -0.015 | 0.002 | 2.1e-11 |
| rs55948744 | 2 | 102805911 | G | T | 0.052 | 0.030 | 0.005 | 9.7e-10 |
| rs12470864 | 2 | 102926362 | A | G | 0.388 | 0.069 | 0.002 | 1e-200 |
| rs66494224 | 2 | 112002314 | C | T | 0.226 | 0.031 | 0.003 | 1e-34 |
| rs4848100 | 2 | 112388532 | T | C | 0.768 | 0.028 | 0.003 | 3.1e-29 |
| rs1257192 | 2 | 134991024 | G | A | 0.812 | 0.017 | 0.003 | 2.3e-10 |
| rs4954388 | 2 | 136808142 | T | C | 0.772 | -0.016 | 0.003 | 3.2e-10 |
| rs12472034 | 2 | 136828273 | G | C | 0.324 | -0.014 | 0.002 | 5.3e-10 |
| rs1427508 | 2 | 145367024 | A | T | 0.826 | 0.021 | 0.003 | 4.1e-14 |
| rs10195713 | 2 | 158520905 | T | C | 0.864 | 0.031 | 0.003 | 1.4e-22 |
| rs6734311 | 2 | 182139508 | A | T | 0.630 | 0.013 | 0.002 | 2.5e-09 |
| rs539299744 | 2 | 197006644 | T | A | 0.135 | -0.021 | 0.003 | 1.5e-11 |
| rs1263595 | 2 | 207947633 | C | T | 0.287 | -0.014 | 0.002 | 2.9e-09 |
| rs715 | 2 | 211543055 | C | T | 0.311 | 0.025 | 0.002 | 4e-28 |
| rs12619285 | 2 | 213824045 | G | A | 0.265 | -0.063 | 0.002 | 8.1e-151 |
| rs35409523 | 2 | 213908457 | A | G | 0.076 | 0.054 | 0.004 | 1.1e-40 |
| rs34855578 | 2 | 231114990 | G | C | 0.186 | 0.026 | 0.003 | 1.7e-21 |
| rs1057258 | 2 | 234115629 | T | C | 0.178 | -0.037 | 0.003 | 1.3e-41 |
| rs34290285 | 2 | 242698640 | A | G | 0.256 | -0.046 | 0.002 | 4.4e-80 |
| rs76474320 | 2 | 242806022 | C | T | 0.078 | 0.033 | 0.004 | 1.1e-16 |
| rs13084872 | 3 | 3068148 | T | C | 0.359 | 0.014 | 0.002 | 3.5e-10 |
| rs6787336 | 3 | 3153194 | A | G | 0.287 | 0.037 | 0.002 | 5.6e-55 |
| rs73019235 | 3 | 3164321 | A | T | 0.123 | 0.020 | 0.003 | 1.1e-09 |
| rs155266 | 3 | 10230728 | A | T | 0.571 | 0.018 | 0.002 | 1e-16 |
| rs2960422 | 3 | 12334991 | A | G | 0.602 | -0.029 | 0.002 | 7.3e-41 |
| rs9872485 | 3 | 16918405 | G | T | 0.164 | -0.017 | 0.003 | 2.4e-09 |
| rs13073683 | 3 | 25393837 | C | T | 0.399 | 0.015 | 0.002 | 3e-12 |
| rs6795177 | 3 | 27800734 | G | A | 0.479 | 0.015 | 0.002 | 1.2e-12 |
| rs7646283 | 3 | 33046480 | T | C | 0.367 | 0.027 | 0.002 | 6e-34 |
| rs2228467 | 3 | 42906116 | C | T | 0.061 | 0.050 | 0.004 | 2.2e-29 |
| rs4072859 | 3 | 49032205 | C | G | 0.659 | 0.027 | 0.002 | 9.7e-31 |
| rs9815974 | 3 | 71439472 | A | G | 0.665 | -0.016 | 0.002 | 3.4e-12 |
| rs4677079 | 3 | 72125863 | T | G | 0.498 | 0.015 | 0.002 | 6.4e-13 |
| rs7649812 | 3 | 98449191 | G | C | 0.242 | 0.018 | 0.002 | 5.8e-13 |
| rs4618204 | 3 | 101281534 | C | T | 0.444 | 0.019 | 0.002 | 5.4e-18 |
| rs9868582 | 3 | 112052927 | T | G | 0.452 | -0.026 | 0.002 | 2.8e-33 |
| rs2399441 | 3 | 112587012 | C | T | 0.350 | -0.020 | 0.002 | 2.2e-19 |
| rs2715275 | 3 | 121781689 | G | A | 0.202 | 0.021 | 0.003 | 5.6e-16 |
| rs9877627 | 3 | 128249249 | G | A | 0.174 | -0.085 | 0.003 | 1e-200 |
| rs2712399 | 3 | 128432953 | G | C | 0.409 | -0.030 | 0.002 | 2.8e-42 |
| rs115702896 | 3 | 129066698 | C | T | 0.019 | -0.052 | 0.008 | 3.4e-11 |
| rs6441306 | 3 | 159952278 | G | A | 0.433 | -0.013 | 0.002 | 1.4e-09 |
| rs191674933 | 3 | 169488691 | T | A | 0.240 | 0.025 | 0.003 | 3.6e-23 |
| rs73046033 | 3 | 171527955 | G | T | 0.116 | -0.022 | 0.003 | 7.3e-11 |
| rs9815874 | 3 | 188441161 | T | C | 0.299 | 0.027 | 0.002 | 2.1e-32 |
| rs9818987 | 3 | 194402430 | T | C | 0.306 | 0.015 | 0.002 | 1.2e-10 |
| rs12152276 | 3 | 196368501 | G | A | 0.081 | -0.035 | 0.004 | 2.9e-19 |
| rs1828803 | 4 | 2689449 | A | C | 0.393 | 0.015 | 0.002 | 2.8e-11 |
| rs13139941 | 4 | 7071741 | G | A | 0.803 | -0.017 | 0.003 | 2.7e-10 |
| rs76981581 | 4 | 38416046 | G | C | 0.048 | -0.032 | 0.005 | 2.6e-10 |
| rs13112758 | 4 | 38555527 | T | C | 0.283 | 0.014 | 0.002 | 1.9e-09 |
| rs12331049 | 4 | 38655772 | T | C | 0.212 | 0.074 | 0.003 | 2.6e-175 |
| rs2254075 | 4 | 39030183 | G | A | 0.666 | -0.015 | 0.002 | 1.4e-10 |
| rs4694639 | 4 | 74699054 | T | C | 0.568 | 0.017 | 0.002 | 2.9e-14 |
| rs13138355 | 4 | 83545976 | T | C | 0.185 | -0.040 | 0.003 | 1.3e-47 |
| rs13107325 | 4 | 103188709 | T | C | 0.075 | -0.030 | 0.004 | 2.5e-13 |
| rs6827756 | 4 | 123184411 | C | T | 0.637 | -0.023 | 0.002 | 8.9e-26 |
| rs2390353 | 4 | 123602097 | C | T | 0.232 | 0.023 | 0.003 | 3.6e-19 |
| rs13120371 | 4 | 139092719 | G | A | 0.327 | 0.015 | 0.002 | 1.4e-11 |
| rs57199846 | 4 | 153353956 | T | A | 0.297 | 0.019 | 0.002 | 2.8e-16 |
| rs2853677 | 5 | 1287194 | A | G | 0.577 | 0.024 | 0.002 | 2.8e-28 |
| rs11737939 | 5 | 10622792 | T | C | 0.165 | 0.025 | 0.003 | 4.3e-18 |
| rs16903574 | 5 | 14610309 | G | C | 0.076 | 0.036 | 0.004 | 2.4e-18 |
| rs4594881 | 5 | 35846815 | T | G | 0.341 | -0.034 | 0.002 | 3.4e-51 |
| rs4703730 | 5 | 76549688 | T | C | 0.517 | -0.016 | 0.002 | 2.1e-13 |
| rs34495 | 5 | 98265807 | T | G | 0.304 | -0.021 | 0.002 | 2.8e-19 |
| rs6884604 | 5 | 110084154 | C | T | 0.089 | 0.026 | 0.004 | 6.3e-12 |
| rs141405779 | 5 | 110424932 | A | G | 0.138 | -0.056 | 0.003 | 1.5e-74 |
| rs35771899 | 5 | 130865179 | A | T | 0.685 | -0.023 | 0.002 | 2e-21 |
| rs2188962 | 5 | 131770805 | T | C | 0.428 | -0.065 | 0.002 | 1e-200 |
| rs244689 | 5 | 133422816 | G | A | 0.877 | -0.022 | 0.003 | 6.4e-12 |
| rs6580223 | 5 | 141489027 | T | G | 0.617 | 0.033 | 0.002 | 3.8e-50 |
| rs56330463 | 5 | 148200011 | C | T | 0.552 | 0.049 | 0.002 | 5e-114 |
| rs6869605 | 5 | 150452866 | C | A | 0.123 | -0.022 | 0.003 | 5.5e-12 |
| rs11748149 | 5 | 156977393 | A | G | 0.106 | 0.021 | 0.003 | 5.8e-10 |
| rs114925538 | 5 | 169696756 | T | C | 0.051 | -0.037 | 0.005 | 4.8e-14 |
| rs35716097 | 5 | 176806636 | T | C | 0.298 | 0.015 | 0.002 | 1.5e-10 |
| rs2666969 | 6 | 336410 | G | A | 0.254 | -0.037 | 0.003 | 1.3e-30 |
| rs9504361 | 6 | 577820 | G | A | 0.446 | -0.029 | 0.002 | 5e-39 |
| rs3132682 | 6 | 30044388 | C | G | 0.588 | -0.047 | 0.002 | 2.7e-104 |
| rs9266321 | 6 | 31330360 | C | G | 0.184 | 0.045 | 0.003 | 1.7e-58 |
| rs192955957 | 6 | 32486965 | T | C | 0.104 | 0.040 | 0.005 | 2.3e-18 |
| rs28383314 | 6 | 32587213 | C | T | 0.625 | 0.040 | 0.002 | 6.3e-74 |
| rs12206050 | 6 | 33564296 | T | A | 0.185 | 0.033 | 0.003 | 3.2e-33 |
| rs9462094 | 6 | 35513603 | T | C | 0.431 | -0.024 | 0.002 | 5.1e-21 |
| rs11751949 | 6 | 42219296 | A | G | 0.071 | -0.034 | 0.004 | 5.8e-16 |
| rs76236447 | 6 | 42225993 | C | G | 0.072 | 0.039 | 0.004 | 2e-21 |
| rs62414592 | 6 | 42516867 | A | G | 0.192 | -0.019 | 0.003 | 1.4e-12 |
| rs9369573 | 6 | 45689356 | T | C | 0.185 | 0.017 | 0.003 | 7e-10 |
| rs2025489 | 6 | 87814959 | G | A | 0.527 | 0.021 | 0.002 | 1.5e-22 |
| rs905670 | 6 | 90958502 | A | G | 0.351 | -0.035 | 0.002 | 1.2e-55 |
| rs12208103 | 6 | 107442431 | T | C | 0.376 | -0.033 | 0.002 | 1.2e-50 |
| rs783646 | 6 | 117286220 | C | G | 0.817 | -0.022 | 0.003 | 4.5e-15 |
| rs9402682 | 6 | 135406183 | T | G | 0.240 | 0.036 | 0.003 | 2.9e-45 |
| rs4548024 | 6 | 138165744 | C | T | 0.237 | -0.017 | 0.003 | 2.2e-11 |
| rs391875 | 6 | 149795136 | A | G | 0.405 | 0.016 | 0.002 | 5.5e-13 |
| rs3093023 | 6 | 167534290 | A | G | 0.433 | 0.013 | 0.002 | 1.3e-09 |
| rs6955702 | 7 | 3157702 | G | A | 0.524 | 0.025 | 0.002 | 2.3e-30 |
| rs10950642 | 7 | 17016646 | A | G | 0.363 | -0.021 | 0.002 | 1.4e-20 |
| rs57834782 | 7 | 20502828 | A | T | 0.245 | -0.056 | 0.002 | 2e-114 |
| rs12537614 | 7 | 22789551 | C | G | 0.592 | -0.023 | 0.002 | 3.3e-26 |
| rs2158799 | 7 | 28277107 | G | C | 0.610 | -0.021 | 0.002 | 8.1e-21 |
| rs60600003 | 7 | 37382465 | G | T | 0.101 | 0.038 | 0.004 | 1.2e-26 |
| rs12536178 | 7 | 45178748 | T | C | 0.223 | 0.015 | 0.003 | 4.6e-09 |
| rs61198215 | 7 | 50329697 | A | G | 0.376 | 0.024 | 0.002 | 8.5e-27 |
| rs12718731 | 7 | 50436948 | G | A | 0.399 | -0.013 | 0.002 | 8e-10 |
| rs80271829 | 7 | 50443141 | C | T | 0.086 | -0.030 | 0.004 | 2e-15 |
| rs62466318 | 7 | 73042085 | T | C | 0.203 | 0.019 | 0.003 | 2e-12 |
| rs13226583 | 7 | 75454152 | T | A | 0.116 | -0.062 | 0.003 | 1.3e-76 |
| rs55879743 | 7 | 75470364 | T | C | 0.067 | 0.077 | 0.004 | 7.6e-70 |
| rs4279 | 7 | 91764128 | A | C | 0.395 | 0.018 | 0.002 | 1e-16 |
| rs73187852 | 7 | 101878336 | T | C | 0.275 | 0.016 | 0.002 | 9.8e-12 |
| rs12705849 | 7 | 112782556 | A | G | 0.407 | -0.021 | 0.002 | 1.5e-22 |
| rs3925331 | 7 | 124446137 | G | C | 0.287 | 0.014 | 0.002 | 4.4e-09 |
| rs6959832 | 7 | 135289854 | A | G | 0.507 | -0.015 | 0.002 | 1e-11 |
| rs6975036 | 7 | 138780350 | A | G | 0.218 | 0.023 | 0.003 | 4.8e-19 |
| rs12530946 | 7 | 148887942 | G | A | 0.614 | 0.048 | 0.002 | 1.8e-106 |
| rs2979256 | 8 | 8871710 | T | C | 0.450 | 0.018 | 0.002 | 4.3e-17 |
| rs2119690 | 8 | 19859539 | A | G | 0.288 | 0.014 | 0.002 | 2.1e-09 |
| rs4871849 | 8 | 22964719 | A | G | 0.708 | -0.027 | 0.002 | 2.7e-31 |
| rs73681119 | 8 | 27272123 | G | C | 0.324 | -0.015 | 0.002 | 1.9e-10 |
| rs45577137 | 8 | 48651633 | G | A | 0.045 | -0.066 | 0.005 | 2.7e-33 |
| rs74915527 | 8 | 48806383 | T | C | 0.097 | 0.026 | 0.004 | 3.5e-13 |
| rs7846314 | 8 | 61650831 | T | A | 0.187 | -0.051 | 0.003 | 3e-76 |
| rs6989099 | 8 | 66898262 | C | T | 0.317 | -0.017 | 0.002 | 3.9e-14 |
| rs10957978 | 8 | 81285139 | T | G | 0.647 | -0.016 | 0.002 | 1.6e-12 |
| rs13255935 | 8 | 95965685 | A | C | 0.513 | 0.013 | 0.002 | 1.7e-09 |
| rs1493935 | 8 | 119848956 | A | G | 0.669 | 0.020 | 0.002 | 7e-18 |
| rs35779726 | 8 | 126154899 | C | G | 0.131 | 0.020 | 0.003 | 1.7e-10 |
| rs11786536 | 8 | 129000416 | A | G | 0.164 | -0.025 | 0.003 | 2.6e-17 |
| rs77959102 | 8 | 130602928 | C | T | 0.143 | -0.060 | 0.003 | 1.5e-83 |
| rs4236746 | 8 | 130699861 | G | A | 0.975 | 0.062 | 0.007 | 4.6e-20 |
| rs2294023 | 8 | 134126039 | G | A | 0.219 | 0.015 | 0.003 | 4.7e-09 |
| rs10087637 | 8 | 142239318 | A | C | 0.637 | -0.015 | 0.002 | 3.2e-11 |
| rs34173062 | 8 | 145158607 | A | G | 0.072 | 0.058 | 0.004 | 4e-40 |
| rs460631 | 9 | 4851440 | G | A | 0.885 | 0.023 | 0.003 | 8.8e-12 |
| rs10974980 | 9 | 5140671 | G | C | 0.270 | 0.029 | 0.002 | 1.6e-33 |
| rs4142528 | 9 | 6172296 | A | T | 0.672 | -0.077 | 0.002 | 1e-200 |
| rs1547258 | 9 | 6523056 | C | T | 0.708 | -0.021 | 0.002 | 3e-19 |
| rs10810657 | 9 | 16884586 | A | T | 0.625 | -0.015 | 0.002 | 5.6e-11 |
| rs11789226 | 9 | 34975508 | C | T | 0.501 | -0.014 | 0.002 | 4.1e-11 |
| rs7869321 | 9 | 86520782 | T | C | 0.255 | 0.031 | 0.002 | 4.9e-36 |
| rs76179738 | 9 | 94127317 | T | C | 0.185 | -0.027 | 0.003 | 1.6e-23 |
| rs4742774 | 9 | 102562927 | C | T | 0.378 | 0.013 | 0.002 | 2.6e-09 |
| rs496475 | 9 | 113638236 | G | T | 0.387 | 0.033 | 0.002 | 8.9e-52 |
| rs1322054 | 9 | 117669299 | G | A | 0.435 | -0.026 | 0.002 | 1e-33 |
| rs7040707 | 9 | 117680732 | G | A | 0.048 | -0.034 | 0.005 | 3.5e-10 |
| rs605642 | 9 | 135880469 | A | C | 0.582 | 0.027 | 0.002 | 2.5e-34 |
| rs671050 | 9 | 136001966 | C | T | 0.674 | -0.014 | 0.002 | 1.4e-09 |
| rs2810489 | 9 | 136922700 | A | G | 0.258 | -0.028 | 0.002 | 4.1e-31 |
| rs11253517 | 10 | 970524 | A | G | 0.238 | 0.034 | 0.003 | 1.3e-42 |
| rs11255507 | 10 | 8109615 | G | T | 0.178 | 0.021 | 0.003 | 6e-14 |
| rs2646438 | 10 | 8565675 | A | G | 0.566 | -0.020 | 0.002 | 3.3e-20 |
| rs1775554 | 10 | 9054340 | C | A | 0.423 | -0.040 | 0.002 | 3.1e-76 |
| rs2992335 | 10 | 26727934 | G | C | 0.598 | -0.030 | 0.002 | 5.8e-44 |
| rs2807740 | 10 | 28784483 | T | C | 0.769 | 0.027 | 0.003 | 6e-27 |
| rs17482472 | 10 | 44859618 | A | G | 0.099 | -0.024 | 0.004 | 4e-11 |
| rs10995251 | 10 | 64398466 | T | C | 0.369 | -0.040 | 0.002 | 1.7e-74 |
| rs72834751 | 10 | 64559465 | T | C | 0.013 | -0.077 | 0.010 | 3.1e-14 |
| rs61855876 | 10 | 65357541 | T | C | 0.163 | -0.017 | 0.003 | 2.1e-09 |
| rs10998535 | 10 | 70805373 | T | C | 0.215 | 0.018 | 0.003 | 2.2e-12 |
| rs564443 | 10 | 89807917 | A | G | 0.163 | 0.022 | 0.003 | 1.5e-14 |
| rs7897422 | 10 | 90849704 | C | T | 0.191 | 0.022 | 0.003 | 1.2e-16 |
| rs7918084 | 10 | 94429467 | T | C | 0.550 | 0.029 | 0.002 | 6.4e-42 |
| rs3862006 | 10 | 111750764 | A | G | 0.148 | -0.018 | 0.003 | 4.4e-09 |
| rs7080536 | 10 | 115348046 | A | G | 0.043 | -0.050 | 0.005 | 2.6e-21 |
| rs3781455 | 10 | 126347688 | C | T | 0.305 | 0.018 | 0.002 | 1.6e-15 |
| rs28475163 | 11 | 327143 | A | G | 0.234 | -0.034 | 0.003 | 1.7e-42 |
| rs61871200 | 11 | 2959313 | T | A | 0.769 | -0.021 | 0.003 | 7.3e-17 |
| rs360124 | 11 | 9802228 | C | G | 0.904 | -0.023 | 0.004 | 1.1e-10 |
| rs34439695 | 11 | 33901483 | T | C | 0.035 | -0.039 | 0.006 | 4.2e-11 |
| rs12577643 | 11 | 47467172 | T | A | 0.326 | 0.016 | 0.002 | 7.2e-13 |
| rs34108746 | 11 | 57146225 | G | A | 0.077 | -0.039 | 0.004 | 1.9e-22 |
| rs663743 | 11 | 64107735 | A | G | 0.341 | 0.015 | 0.002 | 5.7e-11 |
| rs669371 | 11 | 65674153 | T | G | 0.542 | 0.028 | 0.002 | 6.3e-38 |
| rs7936323 | 11 | 76293758 | A | G | 0.478 | 0.042 | 0.002 | 2.2e-85 |
| rs11236813 | 11 | 76343427 | C | G | 0.101 | -0.028 | 0.004 | 3e-15 |
| rs4409785 | 11 | 95311422 | C | T | 0.172 | 0.018 | 0.003 | 4.8e-10 |
| rs637064 | 11 | 108140909 | T | C | 0.555 | 0.021 | 0.002 | 4.4e-23 |
| rs10466533 | 11 | 116612128 | G | A | 0.074 | -0.030 | 0.004 | 1.1e-13 |
| rs11216183 | 11 | 116781545 | A | C | 0.092 | 0.022 | 0.004 | 1.9e-09 |
| rs73005426 | 11 | 118681083 | G | A | 0.211 | 0.022 | 0.003 | 1.4e-17 |
| rs171270 | 11 | 126207745 | G | C | 0.285 | 0.019 | 0.002 | 1.4e-15 |
| rs10893844 | 11 | 128185850 | C | G | 0.502 | 0.026 | 0.002 | 1.2e-33 |
| rs3016776 | 11 | 128606508 | T | C | 0.472 | -0.017 | 0.002 | 6.2e-15 |
| rs12820863 | 12 | 4318723 | T | C | 0.351 | 0.020 | 0.002 | 2.9e-18 |
| rs10849448 | 12 | 6493351 | G | A | 0.753 | -0.022 | 0.003 | 2.6e-18 |
| rs4931002 | 12 | 32143169 | A | C | 0.779 | -0.024 | 0.003 | 8.7e-21 |
| rs11559982 | 12 | 54711574 | G | A | 0.556 | 0.014 | 0.002 | 3.9e-11 |
| rs1689510 | 12 | 56396768 | C | G | 0.338 | 0.029 | 0.002 | 8.9e-39 |
| rs3024971 | 12 | 57493727 | G | T | 0.107 | -0.038 | 0.003 | 2.9e-28 |
| rs11105213 | 12 | 89624206 | G | A | 0.139 | -0.020 | 0.003 | 2.6e-10 |
| rs4760384 | 12 | 92579100 | C | T | 0.171 | -0.031 | 0.003 | 6.9e-28 |
| rs3858607 | 12 | 94536803 | C | G | 0.378 | -0.014 | 0.002 | 1.4e-10 |
| rs140526537 | 12 | 110553366 | T | C | 0.020 | 0.057 | 0.008 | 1.4e-12 |
| rs10774624 | 12 | 111833788 | A | G | 0.515 | -0.080 | 0.002 | 1e-200 |
| rs535850137 | 12 | 112018317 | A | G | 0.407 | 0.028 | 0.003 | 6.2e-25 |
| rs78691875 | 12 | 113070264 | A | C | 0.021 | -0.044 | 0.007 | 2.5e-09 |
| rs12811109 | 12 | 123471094 | A | G | 0.199 | -0.019 | 0.003 | 2.4e-12 |
| rs12861824 | 13 | 31079184 | C | T | 0.273 | -0.019 | 0.003 | 2.8e-13 |
| rs12019493 | 13 | 40373190 | T | C | 0.600 | 0.014 | 0.002 | 4.1e-11 |
| rs2065633 | 13 | 41252265 | C | T | 0.290 | 0.038 | 0.002 | 9.1e-59 |
| rs12876545 | 13 | 43057478 | G | A | 0.300 | 0.025 | 0.002 | 4.6e-27 |
| rs60970458 | 13 | 50424159 | T | G | 0.217 | 0.018 | 0.003 | 2e-12 |
| rs1326125 | 13 | 50888893 | C | A | 0.361 | 0.021 | 0.002 | 2.6e-21 |
| rs9600233 | 13 | 74693401 | A | G | 0.159 | -0.027 | 0.003 | 5.7e-20 |
| rs9557201 | 13 | 99989538 | G | A | 0.272 | 0.018 | 0.002 | 3.6e-14 |
| rs912131 | 13 | 100032346 | G | A | 0.702 | 0.028 | 0.002 | 6.2e-33 |
| rs9525192 | 13 | 114961339 | G | A | 0.531 | -0.017 | 0.002 | 8.8e-16 |
| rs1951121 | 14 | 23429729 | G | T | 0.398 | -0.015 | 0.002 | 3.1e-11 |
| rs2239633 | 14 | 23589057 | A | G | 0.484 | 0.035 | 0.002 | 4.8e-62 |
| rs8020739 | 14 | 35882492 | T | G | 0.643 | 0.023 | 0.002 | 3.7e-24 |
| rs61988304 | 14 | 37699544 | G | A | 0.287 | 0.016 | 0.002 | 3.2e-11 |
| rs6573020 | 14 | 55871452 | T | C | 0.434 | 0.018 | 0.002 | 3.6e-17 |
| rs762810 | 14 | 65544367 | A | C | 0.361 | 0.016 | 0.002 | 2.3e-12 |
| rs11624310 | 14 | 69240079 | G | A | 0.564 | 0.014 | 0.002 | 2.9e-10 |
| rs12882409 | 14 | 73425462 | A | G | 0.176 | -0.018 | 0.003 | 2.7e-10 |
| rs175705 | 14 | 75975648 | G | C | 0.719 | 0.037 | 0.002 | 6.4e-54 |
| rs3742704 | 14 | 88477882 | C | A | 0.092 | 0.023 | 0.004 | 4.1e-10 |
| rs67856193 | 14 | 93024616 | G | C | 0.308 | 0.020 | 0.002 | 3.6e-18 |
| rs11555542 | 14 | 94417531 | C | T | 0.063 | 0.064 | 0.004 | 1.5e-48 |
| rs8012557 | 14 | 103326936 | C | G | 0.563 | -0.014 | 0.002 | 1.9e-11 |
| rs62006172 | 15 | 38904527 | A | G | 0.042 | -0.038 | 0.005 | 4.1e-13 |
| rs1655558 | 15 | 41787585 | G | T | 0.548 | 0.019 | 0.002 | 3.5e-19 |
| rs35311230 | 15 | 43633497 | C | G | 0.200 | 0.022 | 0.003 | 3.2e-16 |
| rs11071528 | 15 | 60701797 | C | G | 0.807 | 0.019 | 0.003 | 1.9e-12 |
| rs34986765 | 15 | 61069201 | C | T | 0.131 | -0.023 | 0.003 | 4.4e-13 |
| rs11638671 | 15 | 63795628 | C | T | 0.343 | -0.015 | 0.002 | 7.9e-12 |
| rs17293632 | 15 | 67442596 | T | C | 0.236 | 0.030 | 0.003 | 2.8e-33 |
| rs28362902 | 15 | 74712178 | A | G | 0.133 | 0.019 | 0.003 | 4.3e-09 |
| rs2115535 | 15 | 80195199 | A | G | 0.568 | 0.030 | 0.002 | 6.6e-44 |
| rs1138358 | 15 | 80263345 | C | A | 0.264 | -0.026 | 0.002 | 1.8e-27 |
| rs13511 | 15 | 86284817 | C | T | 0.577 | -0.019 | 0.002 | 5.1e-19 |
| rs5011651 | 15 | 91166613 | A | G | 0.759 | 0.019 | 0.002 | 2.2e-14 |
| rs4785903 | 16 | 2838274 | C | T | 0.755 | -0.034 | 0.002 | 1.7e-43 |
| rs9934736 | 16 | 3519252 | G | A | 0.047 | 0.033 | 0.005 | 1.3e-10 |
| rs9927527 | 16 | 11212681 | G | A | 0.350 | -0.045 | 0.002 | 1.7e-88 |
| rs2074572 | 16 | 27356359 | T | C | 0.359 | 0.028 | 0.002 | 5.1e-36 |
| rs8060511 | 16 | 30101596 | A | C | 0.480 | -0.027 | 0.002 | 1.7e-35 |
| rs8044535 | 16 | 30464034 | A | T | 0.568 | -0.013 | 0.002 | 5.6e-10 |
| rs72800841 | 16 | 31014247 | A | G | 0.250 | -0.015 | 0.002 | 1.9e-09 |
| rs9924483 | 16 | 48646258 | T | C | 0.310 | -0.026 | 0.002 | 4.9e-30 |
| rs9925905 | 16 | 57080866 | A | T | 0.473 | -0.015 | 0.002 | 2.5e-12 |
| rs73595580 | 16 | 67636767 | A | G | 0.083 | 0.027 | 0.004 | 2.4e-12 |
| rs1170439 | 16 | 68608511 | C | T | 0.779 | 0.024 | 0.003 | 3.9e-20 |
| rs73575076 | 16 | 79744148 | G | A | 0.314 | -0.015 | 0.002 | 1.2e-10 |
| rs301161 | 16 | 85810349 | A | G | 0.850 | 0.025 | 0.003 | 1.6e-17 |
| rs12924774 | 16 | 86003804 | C | G | 0.352 | 0.018 | 0.002 | 2.5e-16 |
| rs17175830 | 16 | 88558164 | A | G | 0.237 | 0.030 | 0.003 | 2.8e-33 |
| rs112036266 | 17 | 2834143 | T | C | 0.176 | 0.017 | 0.003 | 5.7e-10 |
| rs34210653 | 17 | 4535314 | A | G | 0.021 | -0.172 | 0.007 | 6.5e-118 |
| rs12601936 | 17 | 7172609 | G | A | 0.605 | 0.013 | 0.002 | 8.3e-10 |
| rs74480102 | 17 | 7742601 | A | G | 0.043 | -0.068 | 0.005 | 3.8e-38 |
| rs1838149 | 17 | 33819302 | A | G | 0.423 | -0.020 | 0.002 | 4e-19 |
| rs35130019 | 17 | 37983142 | G | A | 0.034 | -0.085 | 0.006 | 2e-48 |
| rs2109221 | 17 | 38830056 | A | G | 0.578 | 0.015 | 0.002 | 1.6e-11 |
| rs9906974 | 17 | 43822772 | C | T | 0.219 | -0.038 | 0.003 | 8.4e-49 |
| rs12952581 | 17 | 47448346 | A | G | 0.362 | 0.029 | 0.002 | 3.5e-39 |
| rs11079348 | 17 | 56438624 | T | G | 0.171 | 0.019 | 0.003 | 4.6e-11 |
| rs62086903 | 17 | 66016006 | C | T | 0.231 | 0.029 | 0.003 | 2.6e-30 |
| rs180506 | 17 | 68274205 | A | G | 0.776 | -0.024 | 0.003 | 2.2e-21 |
| rs11077961 | 17 | 81012749 | G | A | 0.361 | 0.013 | 0.002 | 1.7e-09 |
| rs2847286 | 18 | 12817815 | G | A | 0.352 | -0.015 | 0.002 | 1.2e-11 |
| rs4578769 | 18 | 20395104 | T | G | 0.749 | 0.023 | 0.002 | 1.1e-20 |
| rs57633475 | 18 | 46469650 | G | A | 0.122 | -0.025 | 0.003 | 3.4e-14 |
| rs73963711 | 18 | 60874413 | T | C | 0.210 | 0.023 | 0.003 | 1.2e-17 |
| rs954954 | 18 | 60902328 | C | A | 0.105 | -0.034 | 0.003 | 6.1e-22 |
| rs17758695 | 18 | 60920854 | T | C | 0.029 | -0.104 | 0.006 | 1.8e-61 |
| rs1395269 | 18 | 61377644 | G | T | 0.301 | -0.023 | 0.002 | 3.8e-23 |
| rs3848640 | 19 | 1037986 | G | A | 0.329 | -0.021 | 0.002 | 6.4e-20 |
| rs36084354 | 19 | 1079959 | A | G | 0.092 | -0.034 | 0.004 | 1.3e-20 |
| rs757230 | 19 | 1102131 | G | A | 0.469 | -0.013 | 0.002 | 1.9e-09 |
| rs61731111 | 19 | 3179517 | T | C | 0.011 | -0.132 | 0.010 | 5.1e-38 |
| rs448998 | 19 | 16411418 | G | A | 0.732 | 0.023 | 0.002 | 4e-22 |
| rs410867 | 19 | 16427111 | G | A | 0.216 | -0.065 | 0.003 | 3.3e-138 |
| rs11669369 | 19 | 16654394 | C | G | 0.107 | 0.029 | 0.003 | 5.3e-17 |
| rs117710327 | 19 | 33726578 | A | C | 0.067 | -0.037 | 0.004 | 8.8e-18 |
| rs4802399 | 19 | 38899999 | A | G | 0.043 | 0.045 | 0.005 | 2.4e-17 |
| rs412884 | 19 | 40219449 | C | T | 0.672 | 0.056 | 0.002 | 2.4e-135 |
| rs531660643 | 19 | 45251806 | T | G | 0.023 | -0.069 | 0.007 | 6.9e-21 |
| rs7252363 | 19 | 46354793 | A | G | 0.353 | 0.026 | 0.002 | 1.2e-30 |
| rs11084096 | 19 | 52128795 | A | G | 0.296 | -0.017 | 0.002 | 1.4e-12 |
| rs11667325 | 19 | 52301063 | A | G | 0.663 | -0.019 | 0.002 | 8.7e-18 |
| rs6080761 | 20 | 17629162 | A | G | 0.419 | 0.015 | 0.002 | 6.2e-12 |
| rs80054178 | 20 | 30294682 | C | T | 0.023 | -0.080 | 0.007 | 7.3e-29 |
| rs6141755 | 20 | 31163565 | T | G | 0.236 | -0.016 | 0.003 | 9.7e-11 |
| rs6031302 | 20 | 42656310 | G | C | 0.728 | -0.020 | 0.002 | 2.5e-16 |
| rs6126042 | 20 | 49162186 | T | C | 0.085 | -0.037 | 0.004 | 2.3e-21 |
| rs17728960 | 20 | 50129716 | C | T | 0.068 | -0.029 | 0.004 | 1.3e-11 |
| rs2223043 | 21 | 16438793 | G | A | 0.305 | 0.021 | 0.002 | 4.2e-18 |
| rs2823327 | 21 | 16898807 | T | C | 0.187 | 0.017 | 0.003 | 4.3e-10 |
| rs1000005 | 21 | 34433051 | C | G | 0.591 | -0.021 | 0.002 | 3.6e-21 |
| rs34872427 | 21 | 36294044 | C | T | 0.415 | -0.018 | 0.002 | 2.8e-17 |
| rs2242886 | 21 | 36387806 | T | C | 0.071 | -0.099 | 0.004 | 1.4e-125 |
| rs9979383 | 21 | 36715761 | T | C | 0.630 | 0.031 | 0.002 | 3.9e-44 |
| rs150229150 | 21 | 45641853 | T | C | 0.125 | -0.020 | 0.003 | 6.7e-10 |
| rs1811069 | 22 | 21982054 | G | T | 0.191 | 0.018 | 0.003 | 2.7e-11 |
| rs7288670 | 22 | 24621826 | G | A | 0.306 | -0.019 | 0.002 | 2.8e-16 |
| rs140074 | 22 | 31725634 | G | C | 0.486 | 0.028 | 0.002 | 1.5e-40 |
| rs41280005 | 22 | 37316133 | A | G | 0.167 | -0.029 | 0.003 | 5e-24 |
| rs228951 | 22 | 37529798 | G | A | 0.363 | -0.022 | 0.002 | 1.6e-22 |
| rs2413631 | 22 | 41359786 | A | G | 0.250 | 0.015 | 0.002 | 3.2e-09 |
| rs743002 | 22 | 41404939 | C | T | 0.066 | -0.063 | 0.004 | 1.6e-48 |
| rs78023664 | 22 | 46647844 | A | G | 0.092 | -0.029 | 0.004 | 6.6e-15 |

**Table S15. 115 SNPs significantly associated with basophil percentage of white cells used IVs in forward MR analyses derived from Vuckovic D et al.** Chr: Chromosome; EA: Effect allele; NEA: Non-effect allele; EAF: Effect allele frequency.

| SNP | Chr | Pos | EA | NEA | EAF | Beta | SE | P-value |
| --- | --- | --- | --- | --- | --- | --- | --- | --- |
| rs35076930 | 1 | 17677636 | T | C | 0.293 | 0.021 | 0.003 | 1.6e-13 |
| rs657624 | 1 | 20888348 | A | G | 0.332 | -0.014 | 0.002 | 4e-09 |
| rs12097415 | 1 | 21506276 | A | T | 0.547 | -0.020 | 0.002 | 1.4e-19 |
| rs10788980 | 1 | 54892472 | G | A | 0.447 | 0.014 | 0.002 | 1.4e-10 |
| rs6679677 | 1 | 114303808 | A | C | 0.101 | 0.030 | 0.004 | 4e-16 |
| rs11587409 | 1 | 118143444 | C | G | 0.163 | -0.020 | 0.003 | 1.3e-11 |
| rs12075 | 1 | 159175354 | A | G | 0.579 | 0.027 | 0.002 | 2.9e-33 |
| rs6671847 | 1 | 161478810 | A | G | 0.510 | 0.018 | 0.002 | 3.9e-16 |
| rs145510048 | 1 | 161605454 | C | T | 0.332 | -0.020 | 0.002 | 4.6e-17 |
| rs4987358 | 1 | 169665551 | T | G | 0.270 | 0.015 | 0.002 | 1.7e-09 |
| rs12123922 | 1 | 205140436 | A | G | 0.441 | -0.032 | 0.002 | 5e-48 |
| rs12562618 | 1 | 212450399 | G | A | 0.186 | -0.043 | 0.003 | 1.6e-50 |
| rs483954 | 1 | 212620214 | G | A | 0.395 | 0.018 | 0.002 | 2.1e-15 |
| rs28434172 | 1 | 224614625 | T | C | 0.208 | -0.017 | 0.003 | 5.4e-10 |
| rs2208568 | 1 | 236090155 | C | T | 0.893 | 0.112 | 0.004 | 1e-200 |
| rs62105478 | 2 | 8735869 | A | G | 0.052 | -0.029 | 0.005 | 4.3e-09 |
| rs2384575 | 2 | 27387429 | C | T | 0.291 | 0.015 | 0.002 | 2.6e-09 |
| rs75475627 | 2 | 54787592 | G | C | 0.076 | -0.027 | 0.004 | 1.8e-10 |
| rs12470883 | 2 | 65651851 | A | G | 0.409 | 0.014 | 0.002 | 8.2e-10 |
| rs3755266 | 2 | 103042712 | A | G | 0.482 | 0.015 | 0.002 | 1.4e-11 |
| rs75490288 | 2 | 145377211 | A | G | 0.175 | -0.021 | 0.003 | 8.5e-13 |
| rs78862952 | 2 | 146506667 | C | T | 0.092 | 0.027 | 0.004 | 3.2e-12 |
| rs57338137 | 2 | 213857250 | C | T | 0.077 | 0.029 | 0.004 | 1.3e-12 |
| rs4324460 | 3 | 3120542 | G | T | 0.219 | 0.018 | 0.003 | 2.4e-11 |
| rs1669340 | 3 | 3198380 | T | G | 0.839 | -0.025 | 0.003 | 2.6e-17 |
| rs6766037 | 3 | 16908186 | G | A | 0.218 | -0.040 | 0.003 | 1.5e-50 |
| rs3181077 | 3 | 46250652 | T | C | 0.718 | -0.044 | 0.002 | 7.8e-70 |
| rs6782812 | 3 | 128317997 | A | G | 0.893 | 0.086 | 0.004 | 2.9e-127 |
| rs3774298 | 3 | 187460099 | A | G | 0.688 | -0.020 | 0.002 | 2.6e-16 |
| rs28530750 | 4 | 36312542 | A | G | 0.043 | -0.033 | 0.006 | 2.4e-09 |
| rs16850073 | 4 | 74703999 | T | C | 0.375 | -0.014 | 0.002 | 1.6e-09 |
| rs9131 | 4 | 74963049 | T | C | 0.601 | 0.020 | 0.002 | 2.9e-19 |
| rs72667739 | 4 | 87877499 | C | A | 0.168 | -0.034 | 0.003 | 1.1e-29 |
| rs10013613 | 4 | 103423493 | T | G | 0.402 | -0.017 | 0.002 | 7e-14 |
| rs12513896 | 5 | 10688755 | C | G | 0.252 | 0.016 | 0.003 | 1.2e-10 |
| rs13188960 | 5 | 35853319 | T | G | 0.280 | -0.016 | 0.002 | 7.8e-11 |
| rs34500 | 5 | 98294885 | A | G | 0.084 | -0.035 | 0.004 | 4e-18 |
| rs2271352 | 5 | 126091428 | C | G | 0.226 | 0.048 | 0.003 | 1.5e-71 |
| rs35074529 | 5 | 150838293 | C | T | 0.571 | -0.017 | 0.002 | 1e-14 |
| rs724745 | 6 | 22340396 | G | T | 0.504 | 0.021 | 0.002 | 6.9e-21 |
| rs915125 | 6 | 82463376 | T | C | 0.279 | -0.033 | 0.002 | 2.7e-41 |
| rs72895231 | 6 | 82679396 | A | C | 0.596 | -0.015 | 0.002 | 1.3e-10 |
| rs9376098 | 6 | 135499460 | A | T | 0.349 | 0.031 | 0.002 | 5.6e-41 |
| rs9389286 | 6 | 135640965 | C | G | 0.527 | -0.013 | 0.002 | 1.7e-09 |
| rs852444 | 7 | 5556062 | G | A | 0.399 | -0.015 | 0.002 | 3.3e-11 |
| rs73049252 | 7 | 8016602 | A | G | 0.059 | -0.035 | 0.005 | 1.1e-13 |
| rs62447197 | 7 | 50417632 | G | A | 0.107 | 0.033 | 0.004 | 2.6e-20 |
| rs1186222 | 7 | 75247329 | T | C | 0.462 | -0.028 | 0.002 | 4.2e-36 |
| rs8179 | 7 | 92236164 | C | T | 0.792 | -0.022 | 0.003 | 9.3e-16 |
| rs445 | 7 | 92408370 | T | C | 0.096 | -0.055 | 0.004 | 4.6e-48 |
| rs11772895 | 7 | 143081942 | C | G | 0.277 | 0.039 | 0.002 | 1.5e-55 |
| rs2977799 | 8 | 6696927 | G | A | 0.511 | 0.031 | 0.002 | 8.3e-44 |
| rs2741684 | 8 | 6788418 | G | C | 0.693 | 0.019 | 0.002 | 1.4e-15 |
| rs73523802 | 8 | 6878935 | G | A | 0.271 | 0.045 | 0.003 | 8.3e-60 |
| rs6983894 | 8 | 6938131 | A | C | 0.046 | -0.043 | 0.006 | 1.4e-14 |
| rs45577137 | 8 | 48651633 | G | A | 0.045 | -0.057 | 0.006 | 1.4e-23 |
| rs6995515 | 8 | 61511366 | C | A | 0.371 | -0.014 | 0.002 | 3.4e-09 |
| rs7846314 | 8 | 61650831 | T | A | 0.187 | -0.031 | 0.003 | 2.5e-28 |
| rs7832357 | 8 | 126516197 | G | A | 0.342 | -0.015 | 0.002 | 4e-11 |
| rs4836579 | 9 | 130528548 | T | C | 0.573 | -0.014 | 0.002 | 3.5e-10 |
| rs11253511 | 10 | 964832 | T | C | 0.238 | 0.050 | 0.003 | 6.6e-82 |
| rs11595895 | 10 | 26734518 | A | C | 0.403 | 0.019 | 0.002 | 8.9e-17 |
| rs7082470 | 10 | 65277026 | A | G | 0.471 | 0.019 | 0.002 | 1.9e-17 |
| rs55922628 | 10 | 77191441 | A | C | 0.611 | -0.018 | 0.002 | 2e-14 |
| rs56014906 | 10 | 102103508 | G | A | 0.368 | -0.019 | 0.002 | 5.1e-16 |
| rs2256368 | 10 | 114186624 | A | G | 0.904 | -0.026 | 0.004 | 1.1e-11 |
| rs3781452 | 10 | 126355129 | T | C | 0.633 | 0.015 | 0.002 | 1.4e-10 |
| rs7116520 | 11 | 4053377 | A | G | 0.410 | 0.016 | 0.002 | 6.4e-12 |
| rs74472890 | 11 | 72946279 | C | T | 0.049 | 0.121 | 0.005 | 1.7e-124 |
| rs149709671 | 11 | 74382388 | A | G | 0.034 | 0.036 | 0.006 | 4.5e-09 |
| rs72964941 | 11 | 88025009 | T | C | 0.093 | 0.024 | 0.004 | 7.5e-10 |
| rs4753251 | 11 | 89654536 | A | T | 0.730 | -0.016 | 0.003 | 5.3e-10 |
| rs10893844 | 11 | 128185850 | C | G | 0.502 | -0.015 | 0.002 | 4.6e-11 |
| rs146970669 | 12 | 27103449 | A | G | 0.093 | 0.030 | 0.004 | 7.5e-15 |
| rs117053853 | 12 | 51720047 | A | G | 0.011 | 0.113 | 0.011 | 4.3e-26 |
| rs2118138 | 12 | 66698144 | G | A | 0.528 | 0.015 | 0.002 | 3.1e-12 |
| rs75084335 | 12 | 76981753 | G | C | 0.069 | 0.033 | 0.004 | 6.4e-14 |
| rs2952110 | 12 | 106661316 | G | A | 0.169 | 0.020 | 0.003 | 1.8e-11 |
| rs4475963 | 12 | 129302255 | G | T | 0.370 | -0.019 | 0.002 | 8.6e-17 |
| rs149391212 | 14 | 23544999 | T | C | 0.016 | 0.068 | 0.009 | 3.7e-14 |
| rs2239630 | 14 | 23589349 | G | A | 0.558 | 0.029 | 0.002 | 4e-38 |
| rs11158159 | 14 | 57857162 | C | G | 0.773 | 0.029 | 0.003 | 3.4e-26 |
| rs12878807 | 14 | 67964688 | A | T | 0.478 | -0.013 | 0.002 | 3.5e-09 |
| rs72721631 | 14 | 75807762 | A | C | 0.238 | -0.021 | 0.003 | 9e-16 |
| rs2289511 | 14 | 88454910 | A | G | 0.582 | -0.019 | 0.002 | 1.3e-16 |
| rs12438453 | 15 | 42678609 | G | A | 0.122 | 0.025 | 0.003 | 2.3e-13 |
| rs2009833 | 15 | 50193553 | A | G | 0.360 | 0.015 | 0.002 | 3.4e-10 |
| rs66871053 | 15 | 50558793 | C | T | 0.179 | 0.018 | 0.003 | 2.5e-09 |
| rs7496362 | 15 | 65758874 | G | C | 0.366 | -0.017 | 0.002 | 8e-14 |
| rs2415042 | 15 | 69649385 | G | C | 0.627 | 0.017 | 0.002 | 1.5e-13 |
| rs7166645 | 15 | 83740277 | A | G | 0.387 | 0.014 | 0.002 | 1.4e-09 |
| rs2074585 | 15 | 91009484 | A | G | 0.514 | -0.040 | 0.002 | 2.2e-74 |
| rs8026614 | 15 | 100051136 | T | C | 0.472 | -0.016 | 0.002 | 1.7e-12 |
| rs875740 | 16 | 16123048 | A | C | 0.666 | -0.018 | 0.002 | 8.7e-14 |
| rs879627 | 16 | 88522541 | G | C | 0.437 | 0.016 | 0.002 | 1.9e-12 |
| rs12941811 | 17 | 38159335 | C | T | 0.578 | -0.013 | 0.002 | 2.1e-09 |
| rs28730837 | 17 | 56355397 | A | G | 0.016 | 0.092 | 0.009 | 5.2e-26 |
| rs56378716 | 17 | 56356502 | G | A | 0.013 | 0.060 | 0.010 | 1.8e-09 |
| rs17758695 | 18 | 60920854 | T | C | 0.029 | -0.086 | 0.007 | 2.4e-39 |
| rs62132278 | 19 | 836654 | T | C | 0.256 | -0.034 | 0.003 | 4.5e-37 |
| rs2336068 | 19 | 5813519 | C | A | 0.276 | 0.016 | 0.003 | 6.1e-10 |
| rs1609712 | 19 | 8566634 | A | G | 0.167 | -0.033 | 0.003 | 1.2e-28 |
| rs11554159 | 19 | 18285944 | A | G | 0.270 | -0.017 | 0.002 | 2.8e-12 |
| rs11879838 | 19 | 19742751 | C | G | 0.747 | -0.022 | 0.003 | 3.3e-18 |
| rs78744187 | 19 | 33754548 | T | C | 0.081 | -0.087 | 0.004 | 5e-102 |
| rs34158728 | 19 | 38903032 | A | G | 0.029 | 0.102 | 0.007 | 2.3e-53 |
| rs453827 | 19 | 40224901 | A | G | 0.670 | -0.020 | 0.002 | 2.8e-17 |
| rs56827178 | 20 | 31178577 | A | G | 0.230 | -0.017 | 0.003 | 1.2e-10 |
| rs6141779 | 20 | 31259366 | T | C | 0.145 | 0.025 | 0.003 | 2.9e-15 |
| rs4931 | 20 | 43530234 | C | A | 0.271 | -0.024 | 0.002 | 1.3e-21 |
| rs150498232 | 21 | 36792624 | G | A | 0.027 | 0.043 | 0.007 | 2.5e-09 |
| rs11702209 | 21 | 38808428 | G | C | 0.118 | -0.024 | 0.003 | 4.9e-12 |
| rs2836154 | 21 | 39485842 | T | C | 0.336 | 0.021 | 0.002 | 3.8e-19 |
| rs761702 | 22 | 41404970 | A | G | 0.065 | 0.037 | 0.005 | 2.6e-16 |
| rs5766582 | 22 | 45632528 | T | C | 0.475 | -0.021 | 0.002 | 6e-21 |

**Table S16. 295 SNPs significantly associated with neutrophil percentage of white cells used IVs in forward MR analyses derived from Vuckovic D et al.** Chr: Chromosome; EA: Effect allele; NEA: Non-effect allele; EAF: Effect allele frequency.

| SNP | Chr | Pos | EA | NEA | EAF | Beta | SE | P-value |
| --- | --- | --- | --- | --- | --- | --- | --- | --- |
| rs11121242 | 1 | 8906301 | G | A | 0.513 | 0.016 | 0.002 | 3.1e-14 |
| rs66731853 | 1 | 20916238 | A | G | 0.325 | -0.015 | 0.002 | 1.6e-10 |
| rs4543768 | 1 | 22329645 | C | T | 0.321 | 0.016 | 0.002 | 5.5e-13 |
| rs79567479 | 1 | 27011533 | C | T | 0.080 | -0.037 | 0.004 | 1.9e-20 |
| rs3917932 | 1 | 36943916 | G | C | 0.577 | -0.038 | 0.002 | 5.2e-67 |
| rs12718444 | 1 | 43409179 | T | G | 0.173 | -0.019 | 0.003 | 1.6e-11 |
| rs7537229 | 1 | 56906274 | A | G | 0.896 | 0.045 | 0.004 | 2.1e-37 |
| rs2767481 | 1 | 66015394 | C | T | 0.705 | 0.027 | 0.002 | 5.1e-29 |
| rs2755253 | 1 | 67470843 | T | C | 0.707 | 0.018 | 0.002 | 1.1e-13 |
| rs41313381 | 1 | 79411968 | A | C | 0.031 | 0.052 | 0.006 | 1.5e-17 |
| rs115340020 | 1 | 92766438 | A | G | 0.028 | 0.055 | 0.006 | 2.1e-17 |
| rs11166538 | 1 | 101513708 | G | A | 0.220 | -0.021 | 0.003 | 4.6e-16 |
| rs33967958 | 1 | 101701481 | C | T | 0.318 | 0.015 | 0.002 | 3.6e-10 |
| rs2494015 | 1 | 111734122 | T | C | 0.684 | 0.021 | 0.002 | 5e-20 |
| rs7512697 | 1 | 150218242 | T | C | 0.284 | -0.015 | 0.002 | 4e-10 |
| rs6694531 | 1 | 150683512 | A | C | 0.514 | 0.033 | 0.002 | 2.1e-52 |
| rs2479868 | 1 | 158580069 | T | C | 0.266 | -0.021 | 0.002 | 3.1e-18 |
| rs3026940 | 1 | 159130696 | T | A | 0.008 | -0.120 | 0.014 | 8.1e-19 |
| rs12131588 | 1 | 197666111 | A | G | 0.217 | 0.017 | 0.003 | 3.5e-11 |
| rs7530375 | 1 | 212371305 | G | T | 0.452 | -0.015 | 0.002 | 1e-11 |
| rs6540754 | 1 | 212878652 | T | A | 0.664 | -0.015 | 0.002 | 4.4e-11 |
| rs5011752 | 1 | 221044762 | A | G | 0.675 | -0.019 | 0.002 | 1e-16 |
| rs34791963 | 1 | 224662703 | C | T | 0.252 | 0.015 | 0.003 | 2.4e-09 |
| rs7552783 | 1 | 227174145 | C | T | 0.512 | -0.025 | 0.002 | 3.6e-26 |
| rs822829 | 1 | 234962529 | C | A | 0.879 | 0.024 | 0.003 | 1.6e-13 |
| rs9970896 | 1 | 236104981 | T | A | 0.888 | -0.027 | 0.003 | 3.1e-15 |
| rs61838753 | 1 | 247569300 | C | A | 0.672 | -0.014 | 0.002 | 4.5e-10 |
| rs56188865 | 1 | 247606276 | C | T | 0.372 | -0.016 | 0.002 | 1.9e-13 |
| rs12105729 | 2 | 7614263 | G | A | 0.159 | 0.021 | 0.003 | 1.7e-12 |
| rs10779936 | 2 | 23950200 | A | G | 0.715 | -0.017 | 0.002 | 2.6e-12 |
| rs7578575 | 2 | 25488819 | A | T | 0.293 | -0.027 | 0.002 | 8.7e-29 |
| rs12988231 | 2 | 25593537 | C | T | 0.315 | -0.014 | 0.002 | 2.4e-09 |
| rs6721395 | 2 | 27280127 | A | G | 0.700 | -0.019 | 0.002 | 9.8e-16 |
| rs62149448 | 2 | 40697617 | C | T | 0.189 | 0.019 | 0.003 | 1.2e-11 |
| rs6705577 | 2 | 43359275 | C | G | 0.271 | -0.021 | 0.002 | 1.4e-18 |
| rs113542380 | 2 | 43464818 | A | G | 0.075 | 0.086 | 0.004 | 4.3e-96 |
| rs62137602 | 2 | 53963242 | C | A | 0.184 | 0.020 | 0.003 | 2e-13 |
| rs74625883 | 2 | 65663381 | A | G | 0.176 | -0.025 | 0.003 | 3.2e-18 |
| rs4599108 | 2 | 85543222 | T | C | 0.487 | -0.015 | 0.002 | 1.2e-11 |
| rs1921622 | 2 | 102966067 | A | G | 0.542 | -0.019 | 0.002 | 3.1e-18 |
| rs1969232 | 2 | 111597636 | G | A | 0.683 | -0.014 | 0.002 | 6.4e-10 |
| rs3789087 | 2 | 111791653 | T | C | 0.163 | 0.028 | 0.003 | 1.1e-21 |
| rs55706446 | 2 | 112239082 | T | C | 0.075 | 0.041 | 0.004 | 3.5e-21 |
| rs13409371 | 2 | 113838145 | A | G | 0.396 | 0.028 | 0.002 | 1.9e-36 |
| rs10188023 | 2 | 136981135 | A | G | 0.907 | 0.023 | 0.004 | 6.2e-10 |
| rs354719 | 2 | 143892139 | T | A | 0.825 | 0.026 | 0.003 | 2.2e-20 |
| rs12611922 | 2 | 160462749 | G | T | 0.374 | 0.023 | 0.002 | 1.2e-24 |
| rs6718526 | 2 | 161214175 | C | T | 0.798 | -0.019 | 0.003 | 1.9e-12 |
| rs55954384 | 2 | 182017630 | G | A | 0.315 | 0.024 | 0.002 | 2.9e-25 |
| rs17365418 | 2 | 182173530 | G | A | 0.442 | -0.018 | 0.002 | 4.4e-17 |
| rs3185378 | 2 | 202151439 | C | G | 0.641 | -0.019 | 0.002 | 4.6e-17 |
| rs114050631 | 2 | 219020958 | T | C | 0.011 | -0.121 | 0.011 | 3.4e-28 |
| rs113414093 | 2 | 219859171 | A | G | 0.047 | -0.043 | 0.006 | 6.1e-14 |
| rs11676298 | 2 | 227291731 | G | C | 0.192 | -0.025 | 0.003 | 1.5e-20 |
| rs6741554 | 2 | 232272500 | G | A | 0.248 | 0.016 | 0.003 | 4.2e-10 |
| rs2568569 | 2 | 237796392 | G | A | 0.739 | 0.015 | 0.002 | 1.8e-09 |
| rs13066322 | 3 | 12269266 | T | C | 0.417 | 0.021 | 0.002 | 3.1e-21 |
| rs1563656 | 3 | 28520417 | A | T | 0.414 | -0.017 | 0.002 | 1.4e-15 |
| rs11706384 | 3 | 39296881 | T | G | 0.236 | -0.022 | 0.003 | 2.5e-18 |
| rs13063578 | 3 | 47087837 | A | T | 0.401 | -0.030 | 0.002 | 2.6e-39 |
| rs71298382 | 3 | 71382067 | G | A | 0.027 | -0.076 | 0.007 | 5.6e-28 |
| rs1456117 | 3 | 123078908 | T | G | 0.217 | 0.023 | 0.003 | 5.8e-18 |
| rs115697464 | 3 | 128175328 | C | T | 0.015 | 0.054 | 0.009 | 1.9e-09 |
| rs76937118 | 3 | 128316589 | T | C | 0.050 | -0.039 | 0.005 | 2.4e-14 |
| rs6782812 | 3 | 128317997 | A | G | 0.893 | -0.079 | 0.003 | 8.8e-114 |
| rs143699489 | 3 | 140927121 | G | A | 0.127 | 0.022 | 0.003 | 2.7e-11 |
| rs2871960 | 3 | 141121814 | C | A | 0.445 | -0.017 | 0.002 | 2e-15 |
| rs28465436 | 3 | 169489082 | C | A | 0.248 | -0.016 | 0.003 | 4.4e-10 |
| rs1523475 | 3 | 187444210 | C | T | 0.806 | 0.017 | 0.003 | 6.6e-10 |
| rs789858 | 3 | 194405966 | T | C | 0.403 | 0.014 | 0.002 | 5.8e-10 |
| rs73221129 | 4 | 729603 | G | A | 0.359 | 0.019 | 0.002 | 1.1e-17 |
| rs6831368 | 4 | 6969919 | G | A | 0.362 | -0.015 | 0.002 | 1.8e-11 |
| rs28530750 | 4 | 36312542 | A | G | 0.043 | 0.038 | 0.005 | 1.7e-12 |
| rs6531601 | 4 | 38015513 | A | T | 0.422 | -0.013 | 0.002 | 2e-09 |
| rs6841652 | 4 | 38363262 | C | T | 0.680 | -0.044 | 0.002 | 3.2e-81 |
| rs6827279 | 4 | 38566132 | T | C | 0.539 | 0.024 | 0.002 | 6.7e-26 |
| rs218265 | 4 | 55408999 | C | T | 0.155 | 0.032 | 0.003 | 5.6e-27 |
| rs13121174 | 4 | 55497446 | A | T | 0.381 | -0.016 | 0.002 | 1e-12 |
| rs16850073 | 4 | 74703999 | T | C | 0.375 | 0.045 | 0.002 | 7.4e-90 |
| rs146149115 | 4 | 74768492 | G | A | 0.010 | -0.085 | 0.011 | 5.5e-14 |
| rs546829 | 4 | 74956372 | T | A | 0.626 | -0.056 | 0.002 | 1.1e-140 |
| rs36114828 | 4 | 80923829 | A | C | 0.739 | 0.020 | 0.002 | 1.4e-15 |
| rs11726195 | 4 | 103538911 | G | C | 0.521 | -0.027 | 0.002 | 4.5e-35 |
| rs2189234 | 4 | 106075498 | G | T | 0.618 | -0.034 | 0.002 | 4.7e-53 |
| rs309375 | 4 | 123681156 | A | C | 0.572 | -0.019 | 0.002 | 2e-17 |
| rs4696314 | 4 | 153005716 | C | T | 0.512 | -0.017 | 0.002 | 2.6e-15 |
| rs2853677 | 5 | 1287194 | A | G | 0.577 | -0.025 | 0.002 | 2.1e-31 |
| rs11744391 | 5 | 33241655 | A | C | 0.392 | -0.013 | 0.002 | 1.5e-09 |
| rs11567701 | 5 | 35859863 | T | G | 0.260 | -0.035 | 0.002 | 8.8e-47 |
| rs4865744 | 5 | 52082774 | G | A | 0.921 | 0.026 | 0.004 | 9.5e-11 |
| rs11741775 | 5 | 68590395 | T | C | 0.443 | -0.020 | 0.002 | 2.7e-20 |
| rs10942300 | 5 | 71745412 | T | C | 0.880 | -0.033 | 0.003 | 6.6e-24 |
| rs27643 | 5 | 87994702 | G | T | 0.721 | 0.020 | 0.002 | 4.7e-17 |
| rs2927608 | 5 | 96252432 | A | G | 0.446 | 0.013 | 0.002 | 4.4e-09 |
| rs62385260 | 5 | 131967540 | A | G | 0.188 | -0.027 | 0.003 | 1.3e-22 |
| rs75925066 | 5 | 134722833 | A | C | 0.023 | -0.049 | 0.007 | 9.4e-12 |
| rs6580229 | 5 | 141510754 | G | A | 0.617 | 0.015 | 0.002 | 3e-11 |
| rs56330463 | 5 | 148200011 | C | T | 0.552 | -0.035 | 0.002 | 5.4e-57 |
| rs7720470 | 5 | 149398480 | G | A | 0.625 | -0.018 | 0.002 | 1e-15 |
| rs6878978 | 5 | 157950125 | T | G | 0.278 | 0.018 | 0.002 | 1.2e-13 |
| rs12659737 | 5 | 176742506 | C | T | 0.840 | -0.028 | 0.003 | 9.3e-22 |
| rs61408162 | 5 | 179042400 | T | C | 0.357 | 0.017 | 0.002 | 9.4e-15 |
| rs27474 | 5 | 179342289 | C | A | 0.395 | 0.015 | 0.002 | 3.4e-11 |
| rs3777755 | 6 | 12159699 | T | C | 0.310 | 0.019 | 0.002 | 1.5e-15 |
| rs2560775 | 6 | 13525365 | A | G | 0.692 | -0.017 | 0.002 | 6.7e-14 |
| rs1267500 | 6 | 14715825 | T | C | 0.810 | 0.021 | 0.003 | 9e-14 |
| rs6911302 | 6 | 24820052 | C | A | 0.551 | 0.025 | 0.002 | 2.7e-31 |
| rs3128900 | 6 | 29833057 | T | G | 0.845 | 0.046 | 0.003 | 2.8e-53 |
| rs2071596 | 6 | 31506691 | A | G | 0.163 | 0.027 | 0.003 | 4e-20 |
| rs116787876 | 6 | 32491139 | G | A | 0.422 | -0.028 | 0.003 | 1.7e-26 |
| rs2185798 | 6 | 41773576 | C | G | 0.479 | -0.016 | 0.002 | 3e-14 |
| rs11755487 | 6 | 42506099 | A | G | 0.210 | 0.028 | 0.003 | 1.3e-25 |
| rs1533584 | 6 | 53221183 | C | T | 0.301 | -0.014 | 0.002 | 2.2e-09 |
| rs915125 | 6 | 82463376 | T | C | 0.279 | -0.027 | 0.002 | 2.4e-29 |
| rs9362417 | 6 | 87983690 | G | A | 0.490 | -0.029 | 0.002 | 3e-40 |
| rs12526696 | 6 | 108053364 | A | G | 0.161 | -0.024 | 0.003 | 8e-16 |
| rs12664455 | 6 | 109590739 | T | C | 0.293 | 0.016 | 0.002 | 2e-11 |
| rs13203252 | 6 | 109672736 | G | A | 0.300 | 0.025 | 0.002 | 4.8e-26 |
| rs9398642 | 6 | 121883200 | T | C | 0.144 | 0.018 | 0.003 | 1.7e-09 |
| rs72959041 | 6 | 127454893 | A | G | 0.049 | -0.033 | 0.005 | 5.3e-11 |
| rs928722 | 6 | 137973832 | T | C | 0.221 | 0.017 | 0.003 | 4.3e-11 |
| rs149110519 | 6 | 144385777 | T | C | 0.036 | -0.072 | 0.006 | 2.4e-34 |
| rs212409 | 6 | 159470058 | A | G | 0.553 | -0.014 | 0.002 | 1.7e-10 |
| rs1012657 | 6 | 167525142 | C | A | 0.473 | 0.013 | 0.002 | 1.4e-09 |
| rs6796 | 7 | 6502367 | C | T | 0.278 | -0.040 | 0.002 | 5.4e-61 |
| rs73049270 | 7 | 8021646 | T | C | 0.058 | -0.034 | 0.005 | 2.1e-13 |
| rs212837 | 7 | 26695215 | C | T | 0.621 | -0.013 | 0.002 | 2.2e-09 |
| rs2158799 | 7 | 28277107 | G | C | 0.610 | 0.043 | 0.002 | 7.3e-83 |
| rs56388170 | 7 | 28724374 | T | G | 0.292 | 0.055 | 0.002 | 3.7e-117 |
| rs60600003 | 7 | 37382465 | G | T | 0.101 | -0.025 | 0.004 | 7.6e-12 |
| rs10259698 | 7 | 40069862 | G | A | 0.421 | 0.013 | 0.002 | 3.5e-09 |
| rs876036 | 7 | 50307710 | C | T | 0.320 | 0.027 | 0.002 | 5.3e-32 |
| rs149007767 | 7 | 50370254 | T | C | 0.162 | 0.044 | 0.003 | 3e-48 |
| rs4416724 | 7 | 64982809 | C | A | 0.101 | 0.029 | 0.004 | 2.6e-15 |
| rs42033 | 7 | 92237533 | T | A | 0.208 | 0.028 | 0.003 | 1.4e-25 |
| rs445 | 7 | 92408370 | T | C | 0.096 | -0.045 | 0.004 | 2.3e-34 |
| rs6969234 | 7 | 97987986 | C | T | 0.493 | -0.018 | 0.002 | 4.3e-17 |
| rs6956344 | 7 | 99359151 | T | C | 0.086 | -0.026 | 0.004 | 6.3e-12 |
| rs62621812 | 7 | 127015083 | A | G | 0.020 | -0.062 | 0.008 | 3.1e-15 |
| rs12531590 | 7 | 148888461 | C | A | 0.624 | -0.017 | 0.002 | 1e-14 |
| rs3735080 | 7 | 150217309 | T | C | 0.230 | -0.026 | 0.003 | 1.7e-23 |
| rs4840639 | 8 | 6698413 | A | T | 0.510 | 0.013 | 0.002 | 6.9e-10 |
| rs4841133 | 8 | 9183664 | G | A | 0.909 | 0.039 | 0.004 | 9.6e-25 |
| rs1864585 | 8 | 10732905 | G | A | 0.742 | -0.022 | 0.002 | 7.8e-19 |
| rs34215892 | 8 | 21767240 | A | G | 0.028 | 0.044 | 0.007 | 3.4e-11 |
| rs13272050 | 8 | 23406241 | T | C | 0.171 | 0.017 | 0.003 | 4.9e-09 |
| rs6468341 | 8 | 30279355 | C | T | 0.742 | 0.016 | 0.002 | 1.1e-10 |
| rs4737009 | 8 | 41630405 | A | G | 0.236 | -0.025 | 0.003 | 1.6e-23 |
| rs45577137 | 8 | 48651633 | G | A | 0.045 | 0.066 | 0.006 | 3.6e-32 |
| rs7846314 | 8 | 61650831 | T | A | 0.187 | 0.067 | 0.003 | 4.8e-128 |
| rs2953475 | 8 | 79638138 | A | C | 0.250 | -0.031 | 0.002 | 2.1e-35 |
| rs667927 | 8 | 103912414 | T | A | 0.366 | -0.015 | 0.002 | 8.6e-12 |
| rs2954027 | 8 | 126485294 | A | T | 0.468 | 0.027 | 0.002 | 1e-34 |
| rs4871622 | 8 | 126618736 | T | C | 0.666 | 0.014 | 0.002 | 1.1e-09 |
| rs16904123 | 8 | 130607775 | T | G | 0.187 | 0.034 | 0.003 | 5.2e-34 |
| rs13276309 | 8 | 142338552 | G | A | 0.359 | -0.014 | 0.002 | 3.9e-10 |
| rs385893 | 9 | 4763176 | C | T | 0.524 | 0.026 | 0.002 | 1.4e-33 |
| rs2066361 | 9 | 6218960 | A | T | 0.258 | -0.017 | 0.002 | 2.7e-11 |
| rs3731211 | 9 | 21986847 | A | T | 0.722 | -0.027 | 0.002 | 4.7e-30 |
| rs2065500 | 9 | 22145694 | G | A | 0.158 | 0.032 | 0.003 | 4.4e-27 |
| rs10971930 | 9 | 34116083 | C | T | 0.127 | -0.027 | 0.003 | 3.3e-17 |
| rs2585668 | 9 | 38200670 | C | T | 0.505 | 0.013 | 0.002 | 1.2e-09 |
| rs10867416 | 9 | 82324152 | C | T | 0.656 | 0.013 | 0.002 | 3.9e-09 |
| rs61750929 | 9 | 91495135 | T | C | 0.056 | 0.060 | 0.005 | 8.4e-37 |
| rs10980797 | 9 | 113912553 | G | A | 0.489 | 0.023 | 0.002 | 1.8e-26 |
| rs10817483 | 9 | 116097941 | A | C | 0.357 | -0.014 | 0.002 | 8.4e-10 |
| rs72759286 | 9 | 126985858 | C | T | 0.199 | -0.022 | 0.003 | 2.5e-16 |
| rs455381 | 9 | 136905577 | A | C | 0.266 | 0.020 | 0.002 | 3.4e-16 |
| rs3812565 | 9 | 139272502 | C | T | 0.397 | 0.020 | 0.002 | 8.6e-20 |
| rs61839660 | 10 | 6094697 | T | C | 0.098 | 0.032 | 0.004 | 8.4e-19 |
| rs7916672 | 10 | 13534234 | T | C | 0.600 | 0.013 | 0.002 | 1.1e-09 |
| rs2807742 | 10 | 28781367 | A | G | 0.770 | 0.018 | 0.003 | 1.4e-12 |
| rs2288619 | 10 | 45939818 | T | C | 0.070 | 0.027 | 0.004 | 3.1e-10 |
| rs7084861 | 10 | 50303536 | T | C | 0.236 | 0.017 | 0.003 | 4.5e-11 |
| rs10740054 | 10 | 63702586 | A | T | 0.680 | 0.015 | 0.002 | 6.7e-11 |
| rs4469770 | 10 | 65013912 | G | A | 0.784 | -0.021 | 0.003 | 4.9e-16 |
| rs182049218 | 10 | 72332726 | T | C | 0.035 | -0.037 | 0.006 | 4.2e-10 |
| rs2802372 | 10 | 81047575 | C | A | 0.436 | -0.016 | 0.002 | 8.1e-13 |
| rs1539330 | 10 | 94502738 | T | C | 0.134 | -0.022 | 0.003 | 3.3e-11 |
| rs10882881 | 10 | 99010067 | A | T | 0.600 | 0.020 | 0.002 | 1.8e-19 |
| rs7912643 | 10 | 112027936 | G | A | 0.358 | -0.013 | 0.002 | 3e-09 |
| rs10885079 | 10 | 112797803 | A | G | 0.704 | 0.015 | 0.002 | 3.4e-10 |
| rs11198861 | 10 | 121049819 | A | G | 0.512 | -0.015 | 0.002 | 2e-11 |
| rs11245331 | 10 | 126382353 | A | G | 0.110 | 0.032 | 0.003 | 1.2e-20 |
| rs11603142 | 11 | 300228 | G | C | 0.342 | 0.014 | 0.002 | 2.4e-09 |
| rs14408 | 11 | 308314 | C | T | 0.368 | 0.037 | 0.002 | 6.2e-59 |
| rs11599958 | 11 | 10358095 | T | C | 0.289 | -0.014 | 0.002 | 3.8e-09 |
| rs11033388 | 11 | 36074218 | T | A | 0.418 | -0.014 | 0.002 | 5.4e-11 |
| rs11039216 | 11 | 47406592 | T | C | 0.533 | -0.013 | 0.002 | 3.3e-09 |
| rs7396753 | 11 | 60021508 | C | T | 0.592 | 0.016 | 0.002 | 3.2e-13 |
| rs61897795 | 11 | 61618169 | G | A | 0.164 | -0.022 | 0.003 | 1.7e-14 |
| rs7927894 | 11 | 76301316 | T | C | 0.392 | -0.014 | 0.002 | 9.3e-11 |
| rs72996108 | 11 | 100452869 | A | C | 0.097 | 0.023 | 0.004 | 3.6e-10 |
| rs2606724 | 11 | 113957880 | A | G | 0.455 | 0.019 | 0.002 | 2.4e-17 |
| rs11243 | 11 | 119178640 | G | A | 0.045 | 0.033 | 0.005 | 5.1e-10 |
| rs58432776 | 11 | 122514281 | A | C | 0.380 | -0.032 | 0.002 | 4.4e-46 |
| rs10790962 | 11 | 128385169 | A | G | 0.469 | -0.019 | 0.002 | 2.6e-17 |
| rs34038797 | 12 | 740009 | G | C | 0.479 | 0.015 | 0.002 | 3e-12 |
| rs2364482 | 12 | 6502131 | G | T | 0.197 | -0.035 | 0.003 | 1.4e-37 |
| rs117266223 | 12 | 10543436 | T | A | 0.075 | -0.035 | 0.004 | 7.4e-17 |
| rs11045886 | 12 | 21386493 | C | A | 0.166 | 0.020 | 0.003 | 1.4e-11 |
| rs113736796 | 12 | 48213720 | G | C | 0.039 | 0.047 | 0.006 | 2.9e-16 |
| rs1793937 | 12 | 48375475 | G | C | 0.401 | -0.015 | 0.002 | 3.4e-12 |
| rs190801344 | 12 | 53735778 | G | C | 0.019 | -0.051 | 0.008 | 3.2e-10 |
| rs4761234 | 12 | 69732105 | C | T | 0.485 | 0.024 | 0.002 | 2.1e-29 |
| rs4842610 | 12 | 88811689 | T | C | 0.705 | -0.014 | 0.002 | 1.2e-09 |
| rs35700167 | 12 | 108165223 | T | G | 0.347 | -0.014 | 0.002 | 3.8e-10 |
| rs653178 | 12 | 112007756 | T | C | 0.518 | 0.050 | 0.002 | 1.2e-120 |
| rs28413626 | 12 | 123861452 | A | G | 0.214 | -0.026 | 0.003 | 1.8e-22 |
| rs12428172 | 13 | 28645757 | G | A | 0.544 | 0.013 | 0.002 | 2.4e-09 |
| rs9549201 | 13 | 40977348 | C | T | 0.634 | 0.015 | 0.002 | 1.2e-11 |
| rs9532679 | 13 | 41522338 | C | A | 0.150 | 0.032 | 0.003 | 4.9e-25 |
| rs2044732 | 13 | 42833400 | G | A | 0.214 | 0.022 | 0.003 | 1.8e-16 |
| rs1570621 | 13 | 47170118 | C | T | 0.848 | 0.019 | 0.003 | 9.6e-10 |
| rs9530281 | 13 | 74679161 | A | G | 0.716 | 0.022 | 0.002 | 1.5e-19 |
| rs2997995 | 13 | 114160506 | T | C | 0.295 | 0.018 | 0.002 | 1.1e-13 |
| rs7981577 | 13 | 114835802 | C | T | 0.437 | 0.014 | 0.002 | 2.7e-11 |
| rs2239630 | 14 | 23589349 | G | A | 0.558 | -0.021 | 0.002 | 1.4e-21 |
| rs2038700 | 14 | 25461989 | C | T | 0.394 | 0.017 | 0.002 | 4.5e-15 |
| rs17091786 | 14 | 35565856 | T | A | 0.200 | -0.021 | 0.003 | 1.9e-14 |
| rs73267743 | 14 | 64978142 | T | G | 0.230 | -0.021 | 0.003 | 1.1e-15 |
| rs34765661 | 14 | 69852940 | C | T | 0.065 | 0.040 | 0.004 | 7.5e-20 |
| rs175702 | 14 | 75971133 | C | A | 0.532 | -0.018 | 0.002 | 8.6e-17 |
| rs10146346 | 14 | 93083572 | A | G | 0.408 | -0.013 | 0.002 | 1.2e-09 |
| rs8008502 | 14 | 103359492 | A | G | 0.202 | -0.024 | 0.003 | 1.6e-18 |
| rs4990175 | 14 | 103838509 | A | G | 0.534 | -0.014 | 0.002 | 3.6e-10 |
| rs10851372 | 15 | 39246754 | G | A | 0.409 | -0.013 | 0.002 | 1.2e-09 |
| rs12440045 | 15 | 41782684 | C | A | 0.529 | -0.018 | 0.002 | 4.7e-17 |
| rs4924590 | 15 | 42233673 | A | T | 0.244 | 0.020 | 0.003 | 2.3e-15 |
| rs28885693 | 15 | 51278413 | A | G | 0.459 | -0.013 | 0.002 | 1.3e-09 |
| rs28727558 | 15 | 66016711 | A | T | 0.170 | 0.024 | 0.003 | 1.5e-16 |
| rs4480762 | 15 | 75139426 | G | A | 0.581 | 0.020 | 0.002 | 1.7e-19 |
| rs168108 | 15 | 86183873 | G | C | 0.651 | 0.016 | 0.002 | 6.2e-13 |
| rs28539372 | 15 | 91021412 | A | T | 0.319 | -0.025 | 0.002 | 3.2e-27 |
| rs67175901 | 15 | 101748227 | T | C | 0.108 | 0.039 | 0.003 | 6.1e-29 |
| rs12935657 | 16 | 11219041 | A | G | 0.246 | 0.017 | 0.003 | 3.1e-11 |
| rs61739285 | 16 | 27480797 | T | C | 0.034 | -0.035 | 0.006 | 2.9e-09 |
| rs11150589 | 16 | 30482494 | C | T | 0.517 | -0.023 | 0.002 | 2.3e-25 |
| rs74250734 | 16 | 50212589 | G | T | 0.095 | -0.023 | 0.004 | 3.2e-10 |
| rs36026517 | 16 | 74597758 | C | G | 0.198 | 0.022 | 0.003 | 1e-16 |
| rs391855 | 16 | 85928621 | T | A | 0.423 | 0.043 | 0.002 | 6.1e-86 |
| rs305064 | 16 | 85974142 | C | T | 0.648 | -0.022 | 0.002 | 2.4e-22 |
| rs7202624 | 16 | 86016175 | C | G | 0.811 | -0.023 | 0.003 | 8.2e-17 |
| rs9905106 | 17 | 1373518 | C | T | 0.735 | 0.020 | 0.002 | 8e-17 |
| rs9674744 | 17 | 2729980 | T | C | 0.277 | -0.016 | 0.002 | 1.7e-10 |
| rs75921867 | 17 | 7241277 | T | C | 0.163 | 0.019 | 0.003 | 8.8e-10 |
| rs9910148 | 17 | 16169009 | A | G | 0.476 | -0.038 | 0.002 | 5.7e-70 |
| rs8070995 | 17 | 17167492 | T | C | 0.137 | -0.020 | 0.003 | 3.2e-10 |
| rs113865680 | 17 | 29278821 | C | T | 0.204 | -0.020 | 0.003 | 1.1e-12 |
| rs4580194 | 17 | 38126477 | G | A | 0.435 | 0.071 | 0.002 | 1e-200 |
| rs11654706 | 17 | 38147293 | G | A | 0.168 | -0.021 | 0.003 | 4.2e-13 |
| rs8067341 | 17 | 41079374 | C | T | 0.476 | -0.017 | 0.002 | 2.8e-14 |
| rs71375338 | 17 | 44332793 | A | G | 0.269 | 0.028 | 0.002 | 1.3e-30 |
| rs8178414 | 17 | 56345363 | T | C | 0.013 | 0.100 | 0.010 | 2e-25 |
| rs34097845 | 17 | 56358429 | T | C | 0.057 | 0.054 | 0.005 | 1.9e-29 |
| rs8079549 | 17 | 73066982 | T | G | 0.458 | 0.015 | 0.002 | 1.1e-11 |
| rs4789294 | 17 | 74419177 | G | A | 0.217 | -0.020 | 0.003 | 5.7e-15 |
| rs4969405 | 17 | 79122996 | T | G | 0.420 | 0.014 | 0.002 | 7.2e-11 |
| rs7231555 | 18 | 19699122 | G | A | 0.088 | 0.026 | 0.004 | 2.8e-11 |
| rs75763843 | 18 | 42062380 | C | A | 0.129 | 0.031 | 0.003 | 1.2e-21 |
| rs66836460 | 18 | 60837508 | A | C | 0.136 | -0.023 | 0.003 | 8e-13 |
| rs3177609 | 18 | 74071078 | C | T | 0.077 | -0.028 | 0.004 | 2.6e-12 |
| rs454854 | 18 | 77253866 | C | G | 0.679 | 0.015 | 0.002 | 4.5e-10 |
| rs35140707 | 19 | 1078297 | T | C | 0.100 | 0.038 | 0.004 | 2.5e-25 |
| rs6510834 | 19 | 5008673 | C | T | 0.657 | 0.025 | 0.002 | 1.2e-27 |
| rs35251378 | 19 | 10459969 | A | G | 0.293 | -0.028 | 0.002 | 1.6e-32 |
| rs345627 | 19 | 13917264 | T | C | 0.349 | -0.021 | 0.002 | 7.5e-21 |
| rs10423726 | 19 | 16444697 | A | C | 0.580 | 0.033 | 0.002 | 3.9e-48 |
| rs7251806 | 19 | 16499569 | T | C | 0.841 | 0.065 | 0.003 | 1.1e-108 |
| rs79355165 | 19 | 18522031 | G | A | 0.120 | 0.027 | 0.003 | 2.2e-16 |
| rs867616 | 19 | 19757682 | T | C | 0.177 | -0.018 | 0.003 | 3.7e-10 |
| rs12151289 | 19 | 33751852 | C | G | 0.030 | 0.059 | 0.006 | 8e-20 |
| rs56898944 | 19 | 36662925 | A | T | 0.114 | 0.022 | 0.004 | 2.8e-09 |
| rs4760 | 19 | 44153100 | G | A | 0.156 | -0.050 | 0.003 | 7.5e-63 |
| rs349048 | 19 | 44300937 | T | C | 0.476 | 0.013 | 0.002 | 8.6e-10 |
| rs7255933 | 19 | 45766729 | A | G | 0.258 | -0.015 | 0.002 | 1.7e-09 |
| rs466477 | 19 | 47679798 | C | T | 0.743 | 0.018 | 0.002 | 4e-13 |
| rs8102492 | 19 | 49140809 | C | A | 0.796 | 0.034 | 0.003 | 2.4e-36 |
| rs4630833 | 20 | 1930798 | C | T | 0.263 | 0.032 | 0.002 | 5.3e-38 |
| rs8114815 | 20 | 3757520 | C | T | 0.267 | -0.018 | 0.002 | 2.5e-13 |
| rs7261425 | 20 | 20068635 | G | C | 0.279 | -0.018 | 0.002 | 2.8e-13 |
| rs8114375 | 20 | 31196471 | G | A | 0.394 | -0.015 | 0.002 | 1e-10 |
| rs6071837 | 20 | 38496424 | G | A | 0.247 | -0.015 | 0.003 | 1.7e-09 |
| rs112111275 | 20 | 39198317 | A | G | 0.064 | -0.029 | 0.004 | 1e-10 |
| rs2143606 | 20 | 42838550 | G | A | 0.455 | -0.022 | 0.002 | 9.9e-24 |
| rs259967 | 20 | 57816060 | C | T | 0.528 | 0.025 | 0.002 | 1.5e-29 |
| rs6011066 | 20 | 62364180 | A | G | 0.674 | 0.018 | 0.002 | 1.4e-14 |
| rs1736148 | 21 | 16813212 | C | T | 0.430 | -0.013 | 0.002 | 1e-09 |
| rs2836884 | 21 | 40467643 | C | T | 0.276 | -0.017 | 0.002 | 9.4e-12 |
| rs5747308 | 22 | 18133500 | C | A | 0.505 | 0.016 | 0.002 | 3.6e-13 |
| rs5754104 | 22 | 21916361 | A | G | 0.187 | 0.026 | 0.003 | 4.3e-21 |
| rs118839 | 22 | 28380770 | C | T | 0.482 | 0.014 | 0.002 | 1.4e-10 |
| rs4823076 | 22 | 30519414 | A | G | 0.606 | -0.022 | 0.002 | 1.6e-23 |
| rs5757886 | 22 | 40636076 | C | T | 0.748 | 0.018 | 0.002 | 1.2e-13 |
| rs5758307 | 22 | 41750219 | G | A | 0.229 | -0.026 | 0.003 | 3.4e-23 |

**Table S17. 371 SNPs significantly associated with monocyte percentage of white cells used IVs in forward MR analyses derived from Vuckovic D et al.** Chr: Chromosome; EA: Effect allele; NEA: Non-effect allele; EAF: Effect allele frequency.

| SNP | Chr | Pos | EA | NEA | EAF | Beta | SE | P-value |
| --- | --- | --- | --- | --- | --- | --- | --- | --- |
| rs67289919 | 1 | 2954208 | A | G | 0.183 | 0.017 | 0.003 | 9.3e-10 |
| rs12124851 | 1 | 8903931 | G | A | 0.316 | 0.016 | 0.002 | 1.2e-11 |
| rs79412885 | 1 | 9241839 | A | G | 0.073 | 0.046 | 0.004 | 6.7e-29 |
| rs6703602 | 1 | 10378451 | G | C | 0.127 | 0.025 | 0.003 | 9.2e-15 |
| rs2501299 | 1 | 22345647 | T | C | 0.661 | 0.019 | 0.002 | 5.8e-17 |
| rs12140788 | 1 | 23659214 | A | C | 0.182 | -0.022 | 0.003 | 7.5e-16 |
| rs7416513 | 1 | 26647949 | C | G | 0.762 | 0.034 | 0.002 | 7.5e-44 |
| rs78169804 | 1 | 27155747 | T | G | 0.081 | 0.038 | 0.004 | 1.4e-22 |
| rs3917932 | 1 | 36943916 | G | C | 0.577 | 0.025 | 0.002 | 6e-31 |
| rs11800162 | 1 | 36970512 | T | C | 0.020 | -0.052 | 0.008 | 1.6e-11 |
| rs11209092 | 1 | 41346344 | C | T | 0.599 | -0.015 | 0.002 | 3.8e-12 |
| rs11206004 | 1 | 53210958 | A | G | 0.470 | -0.013 | 0.002 | 1.2e-09 |
| rs7516441 | 1 | 60140247 | A | G | 0.124 | 0.027 | 0.003 | 3.8e-16 |
| rs11579758 | 1 | 65445542 | A | G | 0.757 | 0.017 | 0.002 | 3.7e-12 |
| rs1892534 | 1 | 66105944 | T | C | 0.375 | 0.021 | 0.002 | 3.8e-22 |
| rs17391694 | 1 | 78623626 | T | C | 0.138 | -0.018 | 0.003 | 4.6e-09 |
| rs12118443 | 1 | 90038588 | A | G | 0.489 | -0.015 | 0.002 | 3.6e-12 |
| rs17575497 | 1 | 92315896 | T | C | 0.013 | 0.365 | 0.010 | 1e-200 |
| rs143705965 | 1 | 93055538 | T | C | 0.029 | -0.125 | 0.006 | 1.5e-84 |
| rs4847398 | 1 | 93578402 | T | C | 0.620 | 0.018 | 0.002 | 3.3e-17 |
| rs11166469 | 1 | 100991634 | C | G | 0.756 | 0.017 | 0.002 | 5.2e-12 |
| rs663045 | 1 | 108743059 | C | G | 0.534 | -0.018 | 0.002 | 1.1e-16 |
| rs11102144 | 1 | 111195493 | G | A | 0.169 | -0.023 | 0.003 | 5e-16 |
| rs11464707 | 1 | 118161020 | G | T | 0.164 | 0.027 | 0.003 | 3.6e-21 |
| rs11204682 | 1 | 150595537 | T | G | 0.223 | -0.064 | 0.003 | 1e-140 |
| rs3014880 | 1 | 153344374 | G | C | 0.116 | 0.038 | 0.003 | 7.3e-31 |
| rs12741203 | 1 | 161139951 | T | C | 0.300 | 0.024 | 0.002 | 8.7e-25 |
| rs5030738 | 1 | 161599654 | T | G | 0.018 | -0.064 | 0.008 | 1.3e-15 |
| rs12725126 | 1 | 179427632 | A | G | 0.188 | -0.016 | 0.003 | 4.1e-09 |
| rs2281415 | 1 | 185392845 | T | C | 0.321 | -0.023 | 0.002 | 3.4e-23 |
| rs79273924 | 1 | 198639132 | T | C | 0.044 | -0.033 | 0.005 | 5.8e-10 |
| rs11582805 | 1 | 205653056 | T | C | 0.460 | 0.019 | 0.002 | 3.1e-19 |
| rs1663626 | 1 | 212422567 | A | C | 0.399 | 0.026 | 0.002 | 8e-34 |
| rs2738752 | 1 | 221052765 | G | A | 0.331 | -0.027 | 0.002 | 4.6e-33 |
| rs113292043 | 1 | 223231657 | T | C | 0.067 | -0.028 | 0.004 | 3e-11 |
| rs34791963 | 1 | 224662703 | C | T | 0.252 | -0.016 | 0.002 | 1e-10 |
| rs12593 | 1 | 227172290 | T | C | 0.465 | 0.017 | 0.002 | 3.2e-13 |
| rs72749964 | 1 | 234740975 | A | G | 0.189 | 0.023 | 0.003 | 6.5e-17 |
| rs4626924 | 1 | 234909298 | T | C | 0.551 | -0.036 | 0.002 | 1.7e-63 |
| rs6429432 | 1 | 236107241 | C | A | 0.892 | -0.053 | 0.003 | 5.8e-56 |
| rs113566032 | 1 | 247495385 | C | T | 0.376 | 0.021 | 0.002 | 2e-21 |
| rs3811444 | 1 | 248039451 | T | C | 0.334 | -0.015 | 0.002 | 5.6e-12 |
| rs186975 | 2 | 8442736 | C | G | 0.394 | 0.017 | 0.002 | 1.7e-15 |
| rs138730630 | 2 | 24250088 | A | G | 0.133 | -0.038 | 0.003 | 4.6e-34 |
| rs56180058 | 2 | 25337155 | T | C | 0.160 | 0.018 | 0.003 | 1.4e-09 |
| rs1260326 | 2 | 27730940 | C | T | 0.606 | 0.040 | 0.002 | 5e-78 |
| rs6715862 | 2 | 42328575 | G | A | 0.505 | 0.013 | 0.002 | 1.9e-10 |
| rs28498283 | 2 | 43360065 | T | A | 0.256 | 0.029 | 0.002 | 1.5e-31 |
| rs6750285 | 2 | 64920032 | A | G | 0.244 | 0.024 | 0.002 | 3.7e-22 |
| rs79709502 | 2 | 65087232 | G | C | 0.222 | 0.015 | 0.003 | 3e-09 |
| rs11694714 | 2 | 65663936 | T | C | 0.328 | -0.019 | 0.002 | 1.2e-17 |
| rs72844410 | 2 | 85723006 | C | T | 0.204 | 0.018 | 0.003 | 2.3e-11 |
| rs7593080 | 2 | 86145787 | T | C | 0.572 | 0.016 | 0.002 | 6.5e-14 |
| rs62153700 | 2 | 96581338 | T | C | 0.305 | 0.018 | 0.002 | 8.3e-15 |
| rs3789098 | 2 | 111738371 | A | G | 0.159 | -0.048 | 0.003 | 1.2e-60 |
| rs150449635 | 2 | 111752151 | C | T | 0.022 | 0.148 | 0.008 | 1.6e-86 |
| rs140796214 | 2 | 111861072 | T | A | 0.017 | 0.071 | 0.008 | 6.4e-17 |
| rs72836346 | 2 | 111876613 | C | G | 0.078 | 0.051 | 0.004 | 4.5e-37 |
| rs7604081 | 2 | 127866535 | C | T | 0.056 | -0.036 | 0.005 | 4.8e-15 |
| rs7604460 | 2 | 136773638 | C | T | 0.115 | -0.023 | 0.003 | 5.9e-12 |
| rs2056295 | 2 | 136886897 | G | A | 0.737 | -0.044 | 0.002 | 1.2e-74 |
| rs146048066 | 2 | 145277924 | G | A | 0.036 | 0.040 | 0.006 | 2.4e-10 |
| rs13000983 | 2 | 148561383 | C | T | 0.295 | 0.017 | 0.002 | 6.3e-13 |
| rs10185155 | 2 | 160553228 | A | G | 0.374 | -0.032 | 0.002 | 4.6e-50 |
| rs4664330 | 2 | 161333128 | C | G | 0.316 | -0.020 | 0.002 | 1e-18 |
| rs13023380 | 2 | 163154363 | A | G | 0.518 | 0.014 | 0.002 | 2.8e-11 |
| rs11680095 | 2 | 181825956 | T | C | 0.593 | 0.026 | 0.002 | 1.9e-32 |
| rs6740847 | 2 | 182308352 | G | A | 0.564 | 0.114 | 0.002 | 1e-200 |
| rs12693280 | 2 | 182424718 | G | A | 0.689 | -0.022 | 0.002 | 3.3e-22 |
| rs190303734 | 2 | 203491226 | T | A | 0.295 | 0.019 | 0.002 | 1.3e-16 |
| rs114050631 | 2 | 219020958 | T | C | 0.011 | 0.117 | 0.011 | 1e-27 |
| rs10206184 | 2 | 225748532 | A | T | 0.157 | -0.035 | 0.003 | 1.4e-32 |
| rs6742228 | 2 | 232267116 | G | A | 0.327 | -0.013 | 0.002 | 3e-09 |
| rs11693819 | 2 | 232566206 | A | G | 0.186 | 0.018 | 0.003 | 2.6e-11 |
| rs3749171 | 2 | 241569692 | T | C | 0.182 | -0.032 | 0.003 | 1.5e-31 |
| rs1584063 | 3 | 12266855 | A | G | 0.456 | -0.014 | 0.002 | 3.2e-11 |
| rs11721007 | 3 | 14260417 | A | G | 0.335 | 0.020 | 0.002 | 4.5e-18 |
| rs12485694 | 3 | 24346109 | G | A | 0.583 | -0.017 | 0.002 | 7.6e-15 |
| rs10510607 | 3 | 28286261 | T | C | 0.174 | -0.017 | 0.003 | 4.1e-10 |
| rs73058593 | 3 | 39281374 | C | G | 0.217 | 0.033 | 0.003 | 4.1e-37 |
| rs2228467 | 3 | 42906116 | C | T | 0.061 | 0.149 | 0.004 | 1e-200 |
| rs34847985 | 3 | 45949753 | G | A | 0.260 | -0.020 | 0.002 | 1.2e-15 |
| rs2213290 | 3 | 46406367 | T | C | 0.409 | 0.046 | 0.002 | 1.4e-96 |
| rs9853113 | 3 | 57397738 | A | T | 0.462 | -0.015 | 0.002 | 2.4e-12 |
| rs13079034 | 3 | 71813219 | T | C | 0.112 | -0.027 | 0.003 | 1.7e-15 |
| rs55890339 | 3 | 72386266 | T | C | 0.222 | 0.024 | 0.003 | 4e-20 |
| rs73133996 | 3 | 98429219 | C | G | 0.440 | 0.015 | 0.002 | 7.9e-13 |
| rs1456117 | 3 | 123078908 | T | G | 0.217 | -0.017 | 0.003 | 3.4e-11 |
| rs2939820 | 3 | 128127643 | A | G | 0.294 | -0.016 | 0.002 | 8e-12 |
| rs72983394 | 3 | 128226451 | C | T | 0.023 | -0.068 | 0.007 | 1.1e-21 |
| rs2712421 | 3 | 128287743 | T | G | 0.654 | 0.069 | 0.002 | 1e-200 |
| rs116026278 | 3 | 128323904 | T | C | 0.014 | 0.064 | 0.009 | 4.1e-13 |
| rs9828891 | 3 | 140967436 | T | C | 0.568 | 0.013 | 0.002 | 6.5e-10 |
| rs11916906 | 3 | 141314133 | T | A | 0.401 | -0.029 | 0.002 | 1.1e-39 |
| rs115379740 | 3 | 152031848 | C | T | 0.021 | 0.044 | 0.007 | 4e-09 |
| rs9290357 | 3 | 168819643 | G | T | 0.889 | 0.021 | 0.003 | 7.5e-10 |
| rs12490581 | 3 | 169565030 | A | G | 0.318 | 0.014 | 0.002 | 8.9e-10 |
| rs79287178 | 3 | 172294500 | A | G | 0.031 | -0.057 | 0.006 | 4.7e-19 |
| rs939334 | 3 | 183749731 | C | T | 0.328 | -0.019 | 0.002 | 1.4e-16 |
| rs12485444 | 3 | 188135783 | A | T | 0.545 | -0.029 | 0.002 | 1.1e-40 |
| rs114503621 | 4 | 730272 | C | T | 0.314 | -0.028 | 0.002 | 1e-33 |
| rs871134 | 4 | 7044380 | T | C | 0.574 | 0.029 | 0.002 | 2.9e-41 |
| rs79804860 | 4 | 26306658 | T | C | 0.049 | 0.030 | 0.005 | 1.4e-09 |
| rs28478131 | 4 | 38039008 | C | A | 0.314 | -0.013 | 0.002 | 3.4e-09 |
| rs10213458 | 4 | 38366390 | G | A | 0.682 | -0.018 | 0.002 | 4.7e-15 |
| rs7686818 | 4 | 38682612 | G | A | 0.809 | -0.019 | 0.003 | 4.1e-12 |
| rs2711981 | 4 | 39039258 | T | C | 0.663 | -0.018 | 0.002 | 5.7e-15 |
| rs16850073 | 4 | 74703999 | T | C | 0.375 | -0.027 | 0.002 | 6.6e-34 |
| rs518867 | 4 | 74955050 | C | T | 0.623 | 0.036 | 0.002 | 1.1e-59 |
| rs4693593 | 4 | 84143528 | C | T | 0.354 | -0.028 | 0.002 | 2.6e-35 |
| rs34333163 | 4 | 103283117 | G | A | 0.071 | -0.027 | 0.004 | 4.5e-11 |
| rs11723120 | 4 | 103502551 | A | G | 0.375 | -0.025 | 0.002 | 3.7e-29 |
| rs62343137 | 4 | 145885961 | C | G | 0.371 | -0.016 | 0.002 | 4.4e-13 |
| rs10069690 | 5 | 1279790 | T | C | 0.258 | -0.016 | 0.002 | 6.9e-11 |
| rs11960511 | 5 | 35883241 | G | A | 0.259 | -0.016 | 0.002 | 1.3e-10 |
| rs28722705 | 5 | 55453942 | T | A | 0.148 | 0.031 | 0.003 | 9.4e-26 |
| rs11741775 | 5 | 68590395 | T | C | 0.443 | 0.013 | 0.002 | 9.5e-10 |
| rs6865582 | 5 | 78201833 | G | C | 0.462 | -0.014 | 0.002 | 6.8e-11 |
| rs73187140 | 5 | 90320859 | C | T | 0.036 | 0.035 | 0.006 | 1.6e-09 |
| rs327806 | 5 | 98280018 | T | G | 0.083 | -0.026 | 0.004 | 3.2e-11 |
| rs12521723 | 5 | 132273808 | T | A | 0.163 | -0.020 | 0.003 | 9.6e-12 |
| rs620498 | 5 | 133869270 | G | A | 0.160 | -0.024 | 0.003 | 7.3e-17 |
| rs2082382 | 5 | 148200553 | A | G | 0.548 | 0.026 | 0.002 | 3.6e-33 |
| rs6579771 | 5 | 149482262 | T | C | 0.267 | 0.035 | 0.002 | 4.8e-49 |
| rs193549 | 5 | 149500236 | T | C | 0.323 | -0.054 | 0.002 | 4.6e-123 |
| rs1081071 | 5 | 158381663 | C | T | 0.390 | -0.015 | 0.002 | 8.1e-12 |
| rs56998314 | 5 | 179242418 | G | A | 0.438 | 0.026 | 0.002 | 3.5e-34 |
| rs12203592 | 6 | 396321 | T | C | 0.220 | -0.017 | 0.003 | 1.3e-10 |
| rs3748059 | 6 | 6900951 | G | A | 0.799 | -0.016 | 0.003 | 1e-09 |
| rs509227 | 6 | 10535591 | C | A | 0.522 | -0.022 | 0.002 | 3.2e-25 |
| rs6906830 | 6 | 16711060 | A | C | 0.380 | -0.014 | 0.002 | 1e-10 |
| rs9264400 | 6 | 31229413 | T | C | 0.808 | -0.061 | 0.003 | 6.4e-115 |
| rs11751074 | 6 | 31518157 | A | G | 0.033 | 0.053 | 0.006 | 3.1e-19 |
| rs140932965 | 6 | 31561913 | T | C | 0.011 | -0.069 | 0.011 | 1.2e-10 |
| rs190452998 | 6 | 31606528 | A | G | 0.020 | -0.113 | 0.011 | 2.8e-26 |
| rs28746843 | 6 | 32634137 | C | A | 0.240 | -0.035 | 0.003 | 1.2e-35 |
| rs45482297 | 6 | 35265535 | A | G | 0.030 | 0.063 | 0.006 | 8.8e-24 |
| rs72664259 | 6 | 41986269 | G | A | 0.244 | 0.041 | 0.002 | 1.9e-61 |
| rs10948036 | 6 | 42510305 | A | C | 0.210 | -0.028 | 0.003 | 9.8e-27 |
| rs79894332 | 6 | 44591721 | A | G | 0.101 | -0.045 | 0.004 | 1.8e-37 |
| rs809942 | 6 | 53443269 | A | G | 0.198 | -0.021 | 0.003 | 1.9e-15 |
| rs915125 | 6 | 82463376 | T | C | 0.279 | 0.051 | 0.002 | 2.4e-102 |
| rs4706287 | 6 | 87806496 | G | A | 0.571 | 0.021 | 0.002 | 5.4e-22 |
| rs62414606 | 6 | 89836176 | A | G | 0.304 | -0.015 | 0.002 | 5.2e-11 |
| rs60066732 | 6 | 90935383 | G | A | 0.362 | 0.027 | 0.002 | 2e-34 |
| rs2783019 | 6 | 107422239 | C | G | 0.336 | -0.020 | 0.002 | 5.5e-19 |
| rs77791617 | 6 | 114083374 | T | C | 0.060 | 0.039 | 0.004 | 3.6e-18 |
| rs12190916 | 6 | 131045693 | A | C | 0.153 | -0.023 | 0.003 | 7.6e-15 |
| rs12525521 | 6 | 144371265 | A | G | 0.095 | -0.027 | 0.004 | 7.9e-14 |
| rs149110519 | 6 | 144385777 | T | C | 0.036 | 0.107 | 0.006 | 1.6e-76 |
| rs2292334 | 6 | 160858188 | A | G | 0.390 | -0.015 | 0.002 | 1e-11 |
| rs2273215 | 6 | 170586082 | A | G | 0.459 | -0.013 | 0.002 | 3.5e-09 |
| rs7786990 | 7 | 5499748 | G | T | 0.207 | 0.016 | 0.003 | 7.5e-10 |
| rs6796 | 7 | 6502367 | C | T | 0.278 | 0.050 | 0.002 | 4e-100 |
| rs56195338 | 7 | 8107922 | A | G | 0.058 | 0.042 | 0.005 | 4.5e-20 |
| rs10259620 | 7 | 27202289 | G | A | 0.787 | 0.017 | 0.003 | 7e-11 |
| rs498475 | 7 | 28256240 | A | G | 0.632 | -0.018 | 0.002 | 1.2e-15 |
| rs2158799 | 7 | 28277107 | G | C | 0.610 | -0.019 | 0.002 | 7.8e-19 |
| rs56388170 | 7 | 28724374 | T | G | 0.292 | -0.039 | 0.002 | 1.2e-62 |
| rs73109480 | 7 | 44811221 | T | C | 0.071 | 0.029 | 0.004 | 4.9e-12 |
| rs1870027 | 7 | 50258234 | A | G | 0.312 | -0.079 | 0.002 | 1e-200 |
| rs149007767 | 7 | 50370254 | T | C | 0.162 | -0.092 | 0.003 | 1e-200 |
| rs11766800 | 7 | 50435617 | T | A | 0.322 | 0.026 | 0.002 | 4.1e-31 |
| rs41430449 | 7 | 50798525 | G | C | 0.067 | -0.034 | 0.004 | 1.7e-15 |
| rs2690418 | 7 | 65181112 | C | G | 0.544 | -0.014 | 0.002 | 2.4e-11 |
| rs33951980 | 7 | 73029437 | T | C | 0.130 | 0.038 | 0.003 | 3.3e-33 |
| rs2282979 | 7 | 92264993 | C | T | 0.269 | -0.041 | 0.002 | 2.8e-66 |
| rs12532293 | 7 | 106886032 | C | T | 0.518 | 0.015 | 0.002 | 4.4e-12 |
| rs3778754 | 7 | 128575552 | G | C | 0.467 | 0.013 | 0.002 | 1.7e-09 |
| rs8180748 | 7 | 134740199 | A | C | 0.485 | -0.014 | 0.002 | 1.2e-10 |
| rs7785014 | 7 | 137881690 | C | T | 0.672 | -0.026 | 0.002 | 9.5e-31 |
| rs9640381 | 7 | 143082027 | C | T | 0.488 | 0.014 | 0.002 | 1.2e-11 |
| rs4840638 | 8 | 6698248 | G | T | 0.536 | -0.033 | 0.002 | 6.8e-53 |
| rs17078436 | 8 | 6827165 | C | T | 0.016 | 0.100 | 0.009 | 2e-31 |
| rs7826487 | 8 | 6880925 | G | A | 0.116 | -0.061 | 0.003 | 8.3e-76 |
| rs6988319 | 8 | 6915673 | G | C | 0.608 | 0.021 | 0.002 | 1.7e-21 |
| rs11774798 | 8 | 21758823 | G | C | 0.222 | 0.020 | 0.003 | 7.6e-15 |
| rs72607398 | 8 | 22941873 | G | C | 0.200 | 0.023 | 0.003 | 3.1e-18 |
| rs55650056 | 8 | 23080829 | G | C | 0.185 | 0.017 | 0.003 | 5.9e-10 |
| rs7007639 | 8 | 38820363 | T | C | 0.447 | -0.013 | 0.002 | 4.5e-09 |
| rs145718079 | 8 | 48641192 | A | G | 0.010 | -0.097 | 0.011 | 8.1e-18 |
| rs45577137 | 8 | 48651633 | G | A | 0.045 | -0.082 | 0.005 | 1.8e-50 |
| rs2719233 | 8 | 56886156 | G | A | 0.193 | 0.020 | 0.003 | 9.1e-14 |
| rs10100873 | 8 | 59464803 | G | T | 0.305 | 0.014 | 0.002 | 6.7e-10 |
| rs7846314 | 8 | 61650831 | T | A | 0.187 | -0.049 | 0.003 | 1.4e-73 |
| rs6990105 | 8 | 79601887 | C | T | 0.814 | 0.020 | 0.003 | 2.4e-13 |
| rs10103921 | 8 | 82041714 | G | A | 0.419 | -0.017 | 0.002 | 3.1e-15 |
| rs6469603 | 8 | 116596188 | T | C | 0.563 | -0.033 | 0.002 | 7.5e-54 |
| rs28393360 | 8 | 126619477 | A | T | 0.477 | -0.019 | 0.002 | 4.7e-19 |
| rs10094748 | 8 | 130454047 | C | G | 0.084 | -0.026 | 0.004 | 1.4e-11 |
| rs7843207 | 8 | 130570063 | A | C | 0.506 | -0.065 | 0.002 | 1e-200 |
| rs837224 | 8 | 130971985 | A | G | 0.770 | -0.016 | 0.003 | 1.2e-10 |
| rs11136301 | 8 | 144650510 | A | G | 0.346 | -0.013 | 0.002 | 2.5e-09 |
| rs4741775 | 9 | 307266 | G | A | 0.702 | 0.021 | 0.002 | 4.2e-20 |
| rs12005199 | 9 | 4763491 | A | G | 0.311 | -0.017 | 0.002 | 2.4e-12 |
| rs1887428 | 9 | 4984530 | C | G | 0.625 | 0.015 | 0.002 | 4.1e-11 |
| rs3731246 | 9 | 21971989 | G | C | 0.105 | -0.024 | 0.003 | 1.9e-12 |
| rs10965258 | 9 | 22153663 | G | A | 0.073 | 0.043 | 0.004 | 1.2e-25 |
| rs11557154 | 9 | 34107505 | T | C | 0.127 | 0.032 | 0.003 | 2.8e-23 |
| rs10780209 | 9 | 91472127 | A | G | 0.477 | 0.040 | 0.002 | 2.3e-81 |
| rs7029413 | 9 | 91558264 | A | G | 0.322 | -0.019 | 0.002 | 3.6e-17 |
| rs34374605 | 9 | 113338631 | A | G | 0.050 | -0.033 | 0.005 | 2.7e-11 |
| rs118012336 | 9 | 113731641 | C | T | 0.013 | 0.064 | 0.010 | 2.5e-11 |
| rs10126013 | 9 | 113838318 | A | G | 0.596 | 0.068 | 0.002 | 1e-200 |
| rs1930303 | 9 | 114071879 | A | G | 0.663 | -0.061 | 0.002 | 2.7e-160 |
| rs75943501 | 9 | 114162372 | G | A | 0.052 | 0.056 | 0.005 | 1.4e-31 |
| rs4979025 | 9 | 114421035 | G | T | 0.559 | 0.022 | 0.002 | 1.1e-24 |
| rs72759286 | 9 | 126985858 | C | T | 0.199 | 0.044 | 0.003 | 5.8e-62 |
| rs10739694 | 9 | 130526605 | G | A | 0.588 | -0.017 | 0.002 | 5.2e-15 |
| rs12216891 | 9 | 136127366 | T | C | 0.059 | -0.037 | 0.005 | 3.4e-16 |
| rs1038193 | 9 | 137855206 | G | C | 0.264 | -0.016 | 0.002 | 2.9e-11 |
| rs3812591 | 9 | 139341612 | C | T | 0.275 | -0.026 | 0.002 | 6.8e-29 |
| rs71477838 | 10 | 3827920 | T | C | 0.148 | -0.019 | 0.003 | 1.7e-10 |
| rs11598318 | 10 | 7544309 | A | C | 0.051 | -0.030 | 0.005 | 1.4e-09 |
| rs2646424 | 10 | 8463115 | C | G | 0.326 | -0.026 | 0.002 | 1.3e-29 |
| rs2094285 | 10 | 22418702 | A | G | 0.225 | 0.023 | 0.003 | 4.5e-20 |
| rs2807740 | 10 | 28784483 | T | C | 0.769 | -0.019 | 0.003 | 6.9e-14 |
| rs1184089 | 10 | 61645590 | G | A | 0.673 | -0.017 | 0.002 | 2e-14 |
| rs4595427 | 10 | 65012944 | C | A | 0.472 | 0.020 | 0.002 | 2.7e-21 |
| rs3747866 | 10 | 73521578 | A | G | 0.061 | 0.028 | 0.004 | 4.5e-10 |
| rs10824722 | 10 | 80905542 | T | C | 0.246 | 0.021 | 0.003 | 8.7e-17 |
| rs113452214 | 10 | 89786760 | T | C | 0.026 | 0.040 | 0.007 | 1.7e-09 |
| rs370527799 | 10 | 94036630 | A | T | 0.304 | 0.014 | 0.002 | 3.4e-09 |
| rs11189155 | 10 | 99108947 | A | G | 0.257 | 0.031 | 0.002 | 1.6e-37 |
| rs11817777 | 10 | 101264592 | T | C | 0.107 | 0.029 | 0.003 | 1.6e-17 |
| rs11190141 | 10 | 101292390 | T | C | 0.372 | -0.038 | 0.002 | 1.3e-68 |
| rs10885396 | 10 | 114711755 | T | C | 0.541 | -0.019 | 0.002 | 1.8e-19 |
| rs41287142 | 10 | 121353054 | T | C | 0.023 | -0.044 | 0.007 | 5.7e-10 |
| rs3781454 | 10 | 126348565 | A | G | 0.681 | -0.019 | 0.002 | 4.1e-17 |
| rs7115703 | 11 | 306920 | A | T | 0.513 | -0.053 | 0.002 | 6.8e-139 |
| rs907612 | 11 | 1874221 | T | C | 0.380 | -0.029 | 0.002 | 5.2e-39 |
| rs2633082 | 11 | 47248050 | G | C | 0.808 | -0.027 | 0.003 | 2.7e-24 |
| rs11246680 | 11 | 51584437 | A | G | 0.947 | 0.032 | 0.005 | 3.1e-10 |
| rs1373064 | 11 | 60168200 | T | C | 0.637 | 0.016 | 0.002 | 1.7e-12 |
| rs28456 | 11 | 61589481 | G | A | 0.314 | 0.027 | 0.002 | 8.7e-32 |
| rs74472890 | 11 | 72946279 | C | T | 0.049 | -0.040 | 0.005 | 2.5e-16 |
| rs2606724 | 11 | 113957880 | A | G | 0.455 | -0.033 | 0.002 | 4.9e-51 |
| rs77669868 | 11 | 114040394 | G | A | 0.170 | -0.026 | 0.003 | 1.1e-19 |
| rs6589695 | 11 | 118644323 | G | C | 0.957 | 0.036 | 0.005 | 4.8e-12 |
| rs6589940 | 11 | 122518609 | G | A | 0.387 | 0.024 | 0.002 | 7.1e-29 |
| rs548116 | 11 | 128538015 | C | T | 0.190 | 0.022 | 0.003 | 3.2e-16 |
| rs2159599 | 12 | 710441 | G | A | 0.258 | 0.016 | 0.003 | 1.6e-10 |
| rs10849448 | 12 | 6493351 | G | A | 0.753 | -0.063 | 0.003 | 5.8e-138 |
| rs10772280 | 12 | 10581040 | A | C | 0.875 | -0.046 | 0.003 | 1.2e-45 |
| rs4149083 | 12 | 21380630 | T | A | 0.161 | -0.019 | 0.003 | 1.5e-11 |
| rs4082413 | 12 | 30783184 | G | C | 0.511 | 0.018 | 0.002 | 5.8e-17 |
| rs190801344 | 12 | 53735778 | G | C | 0.019 | 0.080 | 0.008 | 5.4e-24 |
| rs11170652 | 12 | 54099076 | A | G | 0.194 | -0.022 | 0.003 | 3.4e-16 |
| rs35979828 | 12 | 54685880 | T | C | 0.070 | -0.048 | 0.004 | 1.6e-30 |
| rs1800973 | 12 | 69744014 | A | C | 0.061 | 0.115 | 0.004 | 2.1e-149 |
| rs8181612 | 12 | 89904698 | A | T | 0.707 | 0.014 | 0.002 | 4.2e-09 |
| rs4964677 | 12 | 108720446 | C | T | 0.389 | 0.032 | 0.002 | 6e-48 |
| rs7314538 | 12 | 110050110 | G | A | 0.441 | 0.013 | 0.002 | 2.7e-09 |
| rs10849930 | 12 | 111739236 | C | T | 0.161 | 0.018 | 0.003 | 2.8e-10 |
| rs11065286 | 12 | 121271734 | C | T | 0.555 | 0.015 | 0.002 | 2.9e-12 |
| rs7485656 | 12 | 125315647 | G | A | 0.141 | 0.032 | 0.003 | 6.7e-26 |
| rs7326855 | 13 | 28555260 | A | G | 0.180 | -0.020 | 0.003 | 9.4e-13 |
| rs188175496 | 13 | 28595940 | T | C | 0.024 | -0.075 | 0.008 | 1.9e-23 |
| rs76428106 | 13 | 28604007 | C | T | 0.013 | 0.483 | 0.010 | 1e-200 |
| rs17086242 | 13 | 28611221 | C | T | 0.041 | -0.057 | 0.005 | 1.9e-26 |
| rs138028125 | 13 | 28712689 | G | C | 0.035 | 0.109 | 0.006 | 4.5e-74 |
| rs1892548 | 13 | 41002641 | C | T | 0.657 | -0.064 | 0.002 | 2.2e-175 |
| rs9532580 | 13 | 41244260 | C | T | 0.264 | -0.036 | 0.002 | 6.9e-51 |
| rs809845 | 13 | 50724055 | T | C | 0.443 | 0.015 | 0.002 | 5e-13 |
| rs73217470 | 13 | 72513420 | G | A | 0.047 | 0.070 | 0.005 | 2.1e-43 |
| rs111371822 | 13 | 92005134 | G | A | 0.117 | 0.030 | 0.003 | 5.2e-19 |
| rs9521345 | 13 | 110039905 | A | G | 0.354 | -0.026 | 0.002 | 6.7e-32 |
| rs76760541 | 13 | 114191214 | G | A | 0.169 | -0.020 | 0.003 | 4.8e-12 |
| rs6560956 | 13 | 114816003 | A | G | 0.414 | 0.014 | 0.002 | 2.1e-11 |
| rs2239630 | 14 | 23589349 | G | A | 0.558 | 0.051 | 0.002 | 5.1e-126 |
| rs2038700 | 14 | 25461989 | C | T | 0.394 | 0.017 | 0.002 | 4.7e-15 |
| rs8015119 | 14 | 35568788 | T | C | 0.109 | -0.032 | 0.003 | 1.9e-21 |
| rs7146939 | 14 | 65583581 | C | T | 0.581 | 0.013 | 0.002 | 9.7e-10 |
| rs150362244 | 14 | 69884941 | G | A | 0.065 | -0.035 | 0.004 | 3.1e-16 |
| rs4903311 | 14 | 75827841 | T | C | 0.621 | -0.015 | 0.002 | 2.8e-12 |
| rs8016326 | 14 | 103846716 | A | G | 0.737 | 0.055 | 0.002 | 3.2e-113 |
| rs55633823 | 14 | 105954705 | T | C | 0.251 | 0.016 | 0.002 | 2.3e-10 |
| rs484943 | 15 | 40398754 | C | T | 0.662 | 0.022 | 0.002 | 3.9e-21 |
| rs149698681 | 15 | 42651900 | C | G | 0.021 | 0.103 | 0.008 | 8.6e-42 |
| rs150844304 | 15 | 43726625 | C | A | 0.025 | -0.059 | 0.007 | 4.9e-18 |
| rs3743044 | 15 | 50773787 | G | A | 0.028 | 0.046 | 0.006 | 1.7e-12 |
| rs1533378 | 15 | 64646777 | T | C | 0.879 | 0.039 | 0.003 | 2.2e-32 |
| rs12594547 | 15 | 65784751 | A | G | 0.185 | -0.035 | 0.003 | 1.6e-37 |
| rs3765066 | 15 | 75140854 | A | G | 0.672 | -0.022 | 0.002 | 2.5e-22 |
| rs11854390 | 15 | 80224817 | T | C | 0.564 | 0.038 | 0.002 | 1.6e-69 |
| rs7180804 | 15 | 80259074 | A | G | 0.258 | -0.078 | 0.002 | 1e-200 |
| rs338555 | 15 | 86236101 | A | G | 0.636 | -0.020 | 0.002 | 2.4e-20 |
| rs11856737 | 15 | 99094203 | G | C | 0.183 | 0.023 | 0.003 | 2.6e-17 |
| rs3784526 | 15 | 101716797 | A | G | 0.107 | -0.044 | 0.003 | 6.8e-37 |
| rs8047143 | 16 | 1349659 | G | T | 0.277 | -0.015 | 0.002 | 3.1e-10 |
| rs12599911 | 16 | 4062436 | T | G | 0.567 | -0.019 | 0.002 | 2.8e-19 |
| rs8049116 | 16 | 4150530 | T | C | 0.117 | 0.028 | 0.003 | 5.1e-17 |
| rs12325238 | 16 | 10981518 | A | C | 0.265 | 0.019 | 0.002 | 2e-15 |
| rs28449958 | 16 | 28528781 | A | G | 0.343 | -0.013 | 0.002 | 4.7e-09 |
| rs9783783 | 16 | 30078492 | C | G | 0.502 | -0.013 | 0.002 | 1.7e-09 |
| rs12598049 | 16 | 53316854 | G | A | 0.299 | -0.020 | 0.002 | 1.4e-17 |
| rs12924604 | 16 | 74601486 | A | G | 0.198 | -0.032 | 0.003 | 3.4e-34 |
| rs374029 | 16 | 85725953 | A | G | 0.297 | 0.018 | 0.002 | 1.2e-14 |
| rs428578 | 16 | 85917944 | G | A | 0.348 | -0.077 | 0.002 | 1e-200 |
| rs305071 | 16 | 85949271 | A | G | 0.120 | -0.108 | 0.003 | 1e-200 |
| rs2176777 | 16 | 86012521 | C | G | 0.813 | 0.056 | 0.003 | 3.2e-95 |
| rs16940328 | 16 | 86076108 | A | G | 0.113 | 0.035 | 0.003 | 6.6e-26 |
| rs3743860 | 16 | 89818491 | C | T | 0.416 | 0.013 | 0.002 | 4.3e-09 |
| rs9900036 | 17 | 4739161 | G | T | 0.344 | -0.014 | 0.002 | 1.2e-09 |
| rs61759532 | 17 | 7240391 | T | C | 0.247 | 0.022 | 0.003 | 5.5e-18 |
| rs74480102 | 17 | 7742601 | A | G | 0.043 | 0.068 | 0.005 | 4.4e-39 |
| rs541313781 | 17 | 16749390 | G | A | 0.028 | -0.048 | 0.008 | 2e-10 |
| rs2948542 | 17 | 25856486 | G | A | 0.587 | -0.018 | 0.002 | 2.6e-17 |
| rs894606 | 17 | 27909352 | T | C | 0.420 | 0.015 | 0.002 | 9.3e-13 |
| rs7218453 | 17 | 32535460 | T | A | 0.221 | -0.028 | 0.003 | 1.3e-27 |
| rs57968500 | 17 | 38145828 | G | A | 0.388 | -0.066 | 0.002 | 1e-200 |
| rs79364306 | 17 | 41627966 | T | C | 0.060 | -0.027 | 0.004 | 2.2e-09 |
| rs4997213 | 17 | 44186267 | A | G | 0.180 | -0.020 | 0.003 | 1.2e-13 |
| rs11656379 | 17 | 47273515 | G | A | 0.162 | -0.019 | 0.003 | 4.3e-11 |
| rs8178414 | 17 | 56345363 | T | C | 0.013 | -0.070 | 0.009 | 1.1e-13 |
| rs34097845 | 17 | 56358429 | T | C | 0.057 | -0.137 | 0.005 | 4.3e-184 |
| rs7216675 | 17 | 57882674 | C | T | 0.079 | -0.058 | 0.004 | 6.3e-49 |
| rs734232 | 17 | 72765731 | A | G | 0.463 | -0.020 | 0.002 | 1.7e-21 |
| rs79647649 | 17 | 80295875 | C | T | 0.042 | -0.047 | 0.005 | 3.1e-19 |
| rs12946146 | 17 | 81079703 | T | C | 0.486 | 0.055 | 0.002 | 1.2e-125 |
| rs149519071 | 17 | 81152877 | C | T | 0.331 | 0.020 | 0.003 | 3.9e-10 |
| rs238135 | 18 | 3448979 | C | T | 0.290 | -0.020 | 0.002 | 1.5e-16 |
| rs303752 | 18 | 21074255 | A | G | 0.408 | 0.016 | 0.002 | 3.5e-13 |
| rs4800173 | 18 | 21619931 | G | A | 0.804 | 0.021 | 0.003 | 2.7e-14 |
| rs8098694 | 18 | 60780199 | A | G | 0.441 | -0.016 | 0.002 | 2e-13 |
| rs954954 | 18 | 60902328 | C | A | 0.105 | -0.028 | 0.003 | 2.5e-16 |
| rs17758695 | 18 | 60920854 | T | C | 0.029 | -0.073 | 0.006 | 1.6e-31 |
| rs3177609 | 18 | 74071078 | C | T | 0.077 | 0.058 | 0.004 | 9.9e-49 |
| rs3786228 | 18 | 77464170 | C | T | 0.383 | -0.022 | 0.002 | 3.1e-23 |
| rs138303849 | 19 | 837174 | C | G | 0.254 | 0.028 | 0.003 | 2.7e-28 |
| rs376882860 | 19 | 3174514 | T | C | 0.080 | -0.033 | 0.004 | 1.7e-13 |
| rs57900491 | 19 | 4995295 | T | A | 0.341 | -0.018 | 0.002 | 5.4e-15 |
| rs413141 | 19 | 6675989 | G | A | 0.863 | 0.033 | 0.003 | 1.5e-26 |
| rs3786602 | 19 | 16730196 | C | T | 0.346 | -0.015 | 0.002 | 4e-12 |
| rs11666267 | 19 | 17660300 | A | G | 0.566 | -0.014 | 0.002 | 1.9e-10 |
| rs11086102 | 19 | 18398628 | C | G | 0.633 | 0.035 | 0.002 | 3.2e-57 |
| rs3859570 | 19 | 18510925 | C | T | 0.425 | -0.045 | 0.002 | 4.1e-96 |
| rs7256148 | 19 | 30345526 | G | A | 0.498 | -0.014 | 0.002 | 7.5e-11 |
| rs62126615 | 19 | 33749710 | T | C | 0.167 | 0.028 | 0.003 | 3.4e-22 |
| rs12151289 | 19 | 33751852 | C | G | 0.030 | -0.094 | 0.006 | 7.9e-50 |
| rs8103728 | 19 | 33900257 | G | C | 0.670 | 0.015 | 0.002 | 9e-12 |
| rs28528757 | 19 | 36732215 | A | G | 0.226 | -0.016 | 0.003 | 8.9e-11 |
| rs4760 | 19 | 44153100 | G | A | 0.156 | 0.045 | 0.003 | 8.8e-54 |
| rs62116961 | 19 | 44283843 | C | G | 0.376 | -0.043 | 0.002 | 1.1e-83 |
| rs12982781 | 19 | 45786555 | C | T | 0.339 | -0.024 | 0.002 | 1.1e-26 |
| rs73052804 | 19 | 52155401 | T | C | 0.072 | -0.025 | 0.004 | 1.9e-09 |
| rs62143206 | 19 | 54326212 | T | G | 0.211 | -0.078 | 0.003 | 1.6e-196 |
| rs602444 | 19 | 55246731 | A | C | 0.614 | -0.017 | 0.002 | 3.7e-12 |
| rs1883932 | 20 | 8609588 | T | A | 0.508 | -0.015 | 0.002 | 6e-12 |
| rs6121172 | 20 | 30296095 | C | A | 0.288 | -0.020 | 0.002 | 2.9e-17 |
| rs4911236 | 20 | 31082056 | A | G | 0.308 | -0.034 | 0.002 | 2e-46 |
| rs61114499 | 20 | 31192357 | A | G | 0.259 | 0.060 | 0.002 | 1.4e-132 |
| rs4142441 | 20 | 42839620 | G | A | 0.151 | 0.044 | 0.003 | 1.6e-47 |
| rs932905 | 20 | 48796131 | A | G | 0.281 | 0.022 | 0.002 | 2.3e-20 |
| rs17196752 | 20 | 48887268 | T | C | 0.189 | -0.079 | 0.003 | 1.9e-182 |
| rs6020468 | 20 | 48967103 | A | G | 0.195 | -0.051 | 0.003 | 7.9e-80 |
| rs2254953 | 20 | 61574660 | G | A | 0.722 | 0.014 | 0.002 | 3.1e-09 |
| rs2823002 | 21 | 16373836 | T | A | 0.158 | -0.022 | 0.003 | 1.8e-14 |
| rs11088296 | 21 | 36237546 | T | C | 0.262 | 0.014 | 0.002 | 2.2e-09 |
| rs2834754 | 21 | 36463774 | G | C | 0.706 | 0.015 | 0.002 | 2.4e-10 |
| rs80109907 | 21 | 39850955 | A | C | 0.009 | -0.107 | 0.011 | 1.1e-21 |
| rs4817987 | 21 | 40465901 | T | C | 0.266 | 0.019 | 0.002 | 6.2e-15 |
| rs17176611 | 21 | 47676948 | C | A | 0.172 | 0.019 | 0.003 | 5.6e-12 |
| rs41409548 | 22 | 17579495 | A | G | 0.033 | -0.096 | 0.006 | 1.9e-50 |
| rs5748937 | 22 | 17675324 | T | C | 0.054 | -0.060 | 0.005 | 1.3e-32 |
| rs9605235 | 22 | 17702355 | A | G | 0.094 | -0.023 | 0.004 | 2.6e-10 |
| rs143283354 | 22 | 24646573 | T | C | 0.275 | -0.016 | 0.002 | 4.6e-11 |
| rs2015580 | 22 | 32326021 | T | C | 0.312 | -0.017 | 0.002 | 5.8e-13 |
| rs62234274 | 22 | 43549646 | T | C | 0.573 | -0.023 | 0.002 | 4.5e-26 |
| rs738408 | 22 | 44324730 | T | C | 0.216 | 0.023 | 0.003 | 1.2e-19 |
| rs8138399 | 22 | 50044824 | T | C | 0.669 | 0.014 | 0.002 | 1.4e-10 |

**Table S18. 307 SNPs significantly associated with lymphocyte percentage of white cells used IVs in forward MR analyses derived from Vuckovic D et al.** Chr: Chromosome; EA: Effect allele; NEA: Non-effect allele; EAF: Effect allele frequency.

| SNP | Chr | Pos | EA | NEA | EAF | Beta | SE | P-value |
| --- | --- | --- | --- | --- | --- | --- | --- | --- |
| rs1569419 | 1 | 2996602 | C | T | 0.767 | -0.016 | 0.003 | 2.2e-10 |
| rs11121242 | 1 | 8906301 | G | A | 0.513 | -0.015 | 0.002 | 2.7e-12 |
| rs589942 | 1 | 20916080 | G | C | 0.338 | 0.017 | 0.002 | 3e-14 |
| rs79567479 | 1 | 27011533 | C | T | 0.080 | 0.029 | 0.004 | 1.9e-13 |
| rs3917932 | 1 | 36943916 | G | C | 0.577 | 0.032 | 0.002 | 9.7e-48 |
| rs80200046 | 1 | 43423932 | A | G | 0.172 | 0.024 | 0.003 | 3.3e-17 |
| rs6703960 | 1 | 46232723 | T | C | 0.457 | 0.013 | 0.002 | 7.1e-10 |
| rs10789009 | 1 | 56331635 | G | A | 0.562 | 0.013 | 0.002 | 1.1e-09 |
| rs7537229 | 1 | 56906274 | A | G | 0.896 | -0.045 | 0.004 | 1e-37 |
| rs2767481 | 1 | 66015394 | C | T | 0.705 | -0.022 | 0.002 | 7.1e-20 |
| rs1925408 | 1 | 67365034 | T | C | 0.735 | -0.022 | 0.002 | 6.8e-19 |
| rs41313381 | 1 | 79411968 | A | C | 0.031 | -0.069 | 0.006 | 4.1e-30 |
| rs150649461 | 1 | 92925654 | C | G | 0.015 | -0.114 | 0.009 | 1.5e-35 |
| rs2148404 | 1 | 101225444 | A | G | 0.590 | -0.022 | 0.002 | 2e-23 |
| rs9433774 | 1 | 101755299 | T | C | 0.617 | -0.016 | 0.002 | 7.3e-13 |
| rs325927 | 1 | 111698348 | C | T | 0.654 | -0.019 | 0.002 | 1.7e-17 |
| rs7555995 | 1 | 150212707 | G | C | 0.286 | 0.017 | 0.002 | 3.7e-12 |
| rs3894771 | 1 | 150789961 | T | A | 0.448 | 0.022 | 0.002 | 2.4e-23 |
| rs34599082 | 1 | 159175494 | T | C | 0.014 | 0.121 | 0.009 | 5.2e-39 |
| rs571841 | 1 | 160791411 | T | C | 0.426 | 0.021 | 0.002 | 1.4e-21 |
| rs10489481 | 1 | 185391167 | A | G | 0.321 | 0.016 | 0.002 | 1.6e-12 |
| rs16841904 | 1 | 197701992 | T | C | 0.213 | -0.022 | 0.003 | 5.6e-17 |
| rs7555082 | 1 | 198598663 | A | G | 0.118 | 0.027 | 0.003 | 1.5e-15 |
| rs1434282 | 1 | 199010721 | T | C | 0.725 | 0.015 | 0.002 | 7e-10 |
| rs55734382 | 1 | 201019059 | T | C | 0.333 | 0.016 | 0.002 | 9.3e-12 |
| rs4311947 | 1 | 212880752 | T | G | 0.711 | 0.016 | 0.002 | 6.5e-12 |
| rs7552783 | 1 | 227174145 | C | T | 0.512 | 0.021 | 0.002 | 7e-19 |
| rs6697464 | 1 | 235007116 | T | A | 0.291 | -0.014 | 0.002 | 4.5e-09 |
| rs9970896 | 1 | 236104981 | T | A | 0.888 | 0.030 | 0.003 | 2e-18 |
| rs61838753 | 1 | 247569300 | C | A | 0.672 | 0.014 | 0.002 | 9.3e-10 |
| rs12105729 | 2 | 7614263 | G | A | 0.159 | -0.023 | 0.003 | 2.3e-14 |
| rs10779936 | 2 | 23950200 | A | G | 0.715 | 0.022 | 0.002 | 1.4e-20 |
| rs7578575 | 2 | 25488819 | A | T | 0.293 | 0.036 | 0.002 | 7.3e-51 |
| rs1124649 | 2 | 27260469 | A | G | 0.299 | -0.019 | 0.002 | 4e-16 |
| rs4952073 | 2 | 31457981 | C | T | 0.703 | -0.017 | 0.002 | 3.2e-12 |
| rs11688289 | 2 | 40698811 | C | T | 0.189 | -0.019 | 0.003 | 7.1e-12 |
| rs6705577 | 2 | 43359275 | C | G | 0.271 | 0.018 | 0.002 | 1.9e-13 |
| rs113542380 | 2 | 43464818 | A | G | 0.075 | -0.087 | 0.004 | 5e-99 |
| rs698853 | 2 | 54053872 | G | A | 0.276 | -0.017 | 0.002 | 4e-12 |
| rs2860773 | 2 | 65665641 | T | G | 0.408 | -0.020 | 0.002 | 1.3e-19 |
| rs10048745 | 2 | 68962137 | A | G | 0.256 | -0.021 | 0.002 | 8.2e-18 |
| rs4599108 | 2 | 85543222 | T | C | 0.486 | 0.014 | 0.002 | 2.5e-10 |
| rs6419573 | 2 | 103027103 | C | T | 0.780 | 0.026 | 0.003 | 2.2e-23 |
| rs10210955 | 2 | 111599282 | C | G | 0.490 | 0.025 | 0.002 | 1.6e-31 |
| rs55706446 | 2 | 112239082 | T | C | 0.075 | -0.061 | 0.004 | 5e-45 |
| rs13409371 | 2 | 113838145 | A | G | 0.396 | -0.025 | 0.002 | 1.1e-29 |
| rs80018307 | 2 | 129030338 | C | T | 0.050 | 0.033 | 0.005 | 5.1e-11 |
| rs6750955 | 2 | 136982502 | A | C | 0.907 | -0.029 | 0.004 | 3.6e-15 |
| rs354719 | 2 | 143892139 | T | A | 0.825 | -0.025 | 0.003 | 6.3e-18 |
| rs34030812 | 2 | 144248905 | C | T | 0.367 | -0.014 | 0.002 | 3.8e-10 |
| rs12052715 | 2 | 160677375 | G | C | 0.726 | 0.022 | 0.002 | 2e-19 |
| rs6746841 | 2 | 161280888 | A | G | 0.747 | 0.027 | 0.003 | 1.4e-27 |
| rs199514773 | 2 | 181925838 | A | T | 0.641 | 0.024 | 0.002 | 3.3e-24 |
| rs6760805 | 2 | 182157839 | G | A | 0.388 | 0.028 | 0.002 | 2.6e-37 |
| rs13029501 | 2 | 182334753 | T | G | 0.563 | -0.019 | 0.002 | 9.7e-19 |
| rs75507031 | 2 | 202160762 | T | C | 0.732 | 0.022 | 0.002 | 2.3e-20 |
| rs114050631 | 2 | 219020958 | T | C | 0.011 | 0.098 | 0.011 | 3.4e-19 |
| rs78058190 | 2 | 219699999 | A | G | 0.050 | 0.046 | 0.006 | 1.5e-16 |
| rs6713887 | 2 | 220026369 | T | G | 0.638 | -0.015 | 0.002 | 1.9e-11 |
| rs11676298 | 2 | 227291731 | G | C | 0.192 | 0.020 | 0.003 | 5.5e-14 |
| rs57003673 | 2 | 231123460 | C | A | 0.108 | 0.023 | 0.003 | 2.8e-11 |
| rs6741554 | 2 | 232272500 | G | A | 0.248 | -0.015 | 0.002 | 1e-09 |
| rs2568569 | 2 | 237796392 | G | A | 0.739 | -0.015 | 0.002 | 2.7e-09 |
| rs9826367 | 3 | 12294202 | G | A | 0.449 | -0.016 | 0.002 | 3.7e-14 |
| rs75831154 | 3 | 28284622 | G | C | 0.175 | 0.032 | 0.003 | 2.2e-29 |
| rs11706384 | 3 | 39296881 | T | G | 0.236 | 0.016 | 0.003 | 7.2e-10 |
| rs7433284 | 3 | 42872590 | A | G | 0.383 | 0.021 | 0.002 | 5.1e-22 |
| rs13063578 | 3 | 47087837 | A | T | 0.401 | 0.036 | 0.002 | 2.4e-55 |
| rs71298382 | 3 | 71382067 | G | A | 0.027 | 0.091 | 0.007 | 6.6e-39 |
| rs13314303 | 3 | 71421314 | T | C | 0.138 | -0.019 | 0.003 | 2.5e-09 |
| rs1584335 | 3 | 98394871 | C | T | 0.606 | -0.015 | 0.002 | 2.7e-11 |
| rs6781942 | 3 | 119758049 | G | A | 0.372 | 0.015 | 0.002 | 3.2e-12 |
| rs4474982 | 3 | 123068820 | A | G | 0.177 | -0.019 | 0.003 | 8.5e-12 |
| rs6782228 | 3 | 128323424 | C | G | 0.268 | 0.038 | 0.002 | 1.5e-53 |
| rs7625643 | 3 | 141150026 | G | A | 0.446 | 0.021 | 0.002 | 1.6e-21 |
| rs6440859 | 3 | 152977859 | A | T | 0.574 | 0.013 | 0.002 | 1.1e-09 |
| rs1565568 | 3 | 171176050 | A | T | 0.196 | -0.018 | 0.003 | 1.1e-10 |
| rs68062772 | 3 | 195171795 | G | A | 0.183 | 0.019 | 0.003 | 1.2e-11 |
| rs6831368 | 4 | 6969919 | G | A | 0.362 | 0.021 | 0.002 | 1.6e-20 |
| rs6531603 | 4 | 38031267 | C | T | 0.668 | 0.019 | 0.002 | 1.9e-16 |
| rs12649942 | 4 | 38261175 | C | T | 0.352 | 0.014 | 0.002 | 1.6e-09 |
| rs6841652 | 4 | 38363262 | C | T | 0.680 | 0.060 | 0.002 | 5.5e-148 |
| rs6827279 | 4 | 38566132 | T | C | 0.539 | -0.036 | 0.002 | 8.7e-59 |
| rs218265 | 4 | 55408999 | C | T | 0.155 | -0.032 | 0.003 | 1.5e-26 |
| rs6554196 | 4 | 55501109 | G | C | 0.382 | 0.020 | 0.002 | 2.3e-19 |
| rs16850073 | 4 | 74703999 | T | C | 0.375 | -0.039 | 0.002 | 1.7e-68 |
| rs146149115 | 4 | 74768492 | G | A | 0.010 | 0.079 | 0.011 | 2.2e-12 |
| rs546829 | 4 | 74956372 | T | A | 0.626 | 0.047 | 0.002 | 7.1e-103 |
| rs28830602 | 4 | 80908833 | G | A | 0.629 | -0.020 | 0.002 | 4e-20 |
| rs11726195 | 4 | 103538911 | G | C | 0.522 | 0.033 | 0.002 | 1e-53 |
| rs6533181 | 4 | 106074826 | G | T | 0.632 | 0.038 | 0.002 | 8.1e-64 |
| rs309375 | 4 | 123681156 | A | C | 0.572 | 0.020 | 0.002 | 1.2e-19 |
| rs60428870 | 4 | 144896336 | A | G | 0.616 | -0.015 | 0.002 | 1.3e-10 |
| rs4696314 | 4 | 153005716 | C | T | 0.512 | 0.022 | 0.002 | 3.9e-24 |
| rs2853677 | 5 | 1287194 | A | G | 0.577 | 0.020 | 0.002 | 2.7e-20 |
| rs3822412 | 5 | 10689768 | G | A | 0.369 | -0.015 | 0.002 | 4e-11 |
| rs11567701 | 5 | 35859863 | T | G | 0.260 | 0.042 | 0.002 | 6.1e-67 |
| rs4865744 | 5 | 52082774 | G | A | 0.921 | -0.028 | 0.004 | 1.5e-12 |
| rs13154469 | 5 | 68607707 | C | T | 0.441 | 0.016 | 0.002 | 5.6e-14 |
| rs10942300 | 5 | 71745412 | T | C | 0.880 | 0.038 | 0.003 | 9.4e-30 |
| rs304151 | 5 | 88125853 | G | T | 0.268 | 0.020 | 0.002 | 3e-16 |
| rs2910687 | 5 | 96276877 | T | C | 0.435 | -0.014 | 0.002 | 1.4e-10 |
| rs114378220 | 5 | 110566360 | T | C | 0.074 | -0.033 | 0.004 | 2.4e-14 |
| rs2250127 | 5 | 127388844 | A | G | 0.247 | -0.018 | 0.002 | 9.9e-13 |
| rs3846730 | 5 | 131806780 | T | C | 0.237 | -0.022 | 0.003 | 2.4e-18 |
| rs7700687 | 5 | 141491985 | T | C | 0.617 | -0.020 | 0.002 | 3e-20 |
| rs56330463 | 5 | 148200011 | C | T | 0.552 | 0.022 | 0.002 | 6.7e-23 |
| rs9313777 | 5 | 157945599 | G | T | 0.281 | -0.023 | 0.002 | 1.6e-22 |
| rs535833686 | 5 | 158347179 | G | A | 0.024 | -0.047 | 0.008 | 8.8e-10 |
| rs55775466 | 5 | 172006296 | G | A | 0.120 | -0.020 | 0.003 | 8.4e-10 |
| rs12659737 | 5 | 176742506 | C | T | 0.840 | 0.032 | 0.003 | 4e-27 |
| rs55785724 | 5 | 176817583 | T | A | 0.312 | -0.015 | 0.002 | 8.7e-11 |
| rs72835315 | 6 | 231878 | C | T | 0.058 | 0.031 | 0.005 | 3.2e-11 |
| rs1050979 | 6 | 410417 | G | A | 0.528 | 0.014 | 0.002 | 1.7e-10 |
| rs3777755 | 6 | 12159699 | T | C | 0.310 | -0.019 | 0.002 | 7.8e-17 |
| rs2560775 | 6 | 13525365 | A | G | 0.692 | 0.020 | 0.002 | 4.6e-18 |
| rs1267498 | 6 | 14715910 | C | T | 0.807 | -0.024 | 0.003 | 5.9e-19 |
| rs4713208 | 6 | 29283579 | C | T | 0.616 | 0.013 | 0.002 | 2.9e-09 |
| rs28883698 | 6 | 29846390 | A | G | 0.200 | -0.043 | 0.003 | 3.7e-34 |
| rs73390981 | 6 | 31243578 | A | G | 0.078 | 0.065 | 0.004 | 3.4e-57 |
| rs7758790 | 6 | 31552850 | C | T | 0.226 | 0.023 | 0.003 | 1.2e-18 |
| rs113571892 | 6 | 32489037 | C | T | 0.251 | -0.029 | 0.003 | 2.2e-24 |
| rs17880292 | 6 | 32552000 | T | C | 0.043 | -0.055 | 0.007 | 4.7e-16 |
| rs11758943 | 6 | 42501162 | C | G | 0.208 | -0.018 | 0.003 | 5.5e-12 |
| rs1533584 | 6 | 53221183 | C | T | 0.301 | 0.017 | 0.002 | 8.2e-13 |
| rs915125 | 6 | 82463376 | T | C | 0.279 | 0.018 | 0.002 | 4.9e-14 |
| rs3734187 | 6 | 87969737 | T | C | 0.490 | 0.022 | 0.002 | 2.3e-25 |
| rs12526696 | 6 | 108053364 | A | G | 0.161 | 0.033 | 0.003 | 3.9e-29 |
| rs9487023 | 6 | 109590004 | G | A | 0.448 | -0.024 | 0.002 | 4.2e-28 |
| rs577721086 | 6 | 127440047 | C | T | 0.050 | 0.030 | 0.005 | 1.2e-09 |
| rs12203816 | 6 | 135529561 | T | C | 0.401 | 0.017 | 0.002 | 1.1e-14 |
| rs1012515 | 6 | 137216847 | T | A | 0.494 | 0.015 | 0.002 | 5.3e-12 |
| rs142761146 | 6 | 138004508 | A | T | 0.181 | 0.018 | 0.003 | 2.1e-10 |
| rs566028 | 6 | 147520965 | C | T | 0.452 | 0.016 | 0.002 | 4e-13 |
| rs6956301 | 7 | 971124 | A | G | 0.667 | -0.016 | 0.002 | 2.9e-12 |
| rs6955702 | 7 | 3157702 | G | A | 0.524 | -0.021 | 0.002 | 7.3e-23 |
| rs2347784 | 7 | 6524843 | G | C | 0.271 | 0.031 | 0.002 | 2.5e-36 |
| rs11763992 | 7 | 22725522 | T | A | 0.305 | -0.014 | 0.002 | 1.7e-09 |
| rs6978504 | 7 | 26695880 | G | C | 0.344 | -0.016 | 0.002 | 4.4e-13 |
| rs498475 | 7 | 28256240 | A | G | 0.633 | 0.016 | 0.002 | 1.5e-12 |
| rs2158799 | 7 | 28277107 | G | C | 0.610 | -0.038 | 0.002 | 7.1e-67 |
| rs56388170 | 7 | 28724374 | T | G | 0.292 | -0.049 | 0.002 | 1.6e-95 |
| rs532636967 | 7 | 40060707 | C | T | 0.461 | -0.015 | 0.002 | 3.6e-11 |
| rs149007767 | 7 | 50370254 | T | C | 0.162 | -0.023 | 0.003 | 2.8e-14 |
| rs2943379 | 7 | 65174354 | T | C | 0.098 | -0.021 | 0.004 | 3.7e-09 |
| rs2108321 | 7 | 74297551 | A | T | 0.285 | 0.019 | 0.002 | 1e-14 |
| rs42035 | 7 | 92239531 | G | A | 0.245 | -0.023 | 0.003 | 1.5e-19 |
| rs445 | 7 | 92408370 | T | C | 0.096 | 0.068 | 0.004 | 7.2e-77 |
| rs56229643 | 7 | 97865936 | C | T | 0.491 | 0.022 | 0.002 | 1.4e-23 |
| rs776739 | 7 | 99283696 | C | T | 0.082 | 0.031 | 0.004 | 1.6e-15 |
| rs9920 | 7 | 116200092 | C | T | 0.105 | 0.026 | 0.004 | 1.1e-13 |
| rs62621812 | 7 | 127015083 | A | G | 0.020 | 0.071 | 0.008 | 1e-19 |
| rs62485905 | 7 | 138741586 | T | C | 0.551 | -0.013 | 0.002 | 2.2e-09 |
| rs3735080 | 7 | 150217309 | T | C | 0.230 | 0.024 | 0.003 | 1.1e-21 |
| rs7457817 | 7 | 156596400 | C | T | 0.418 | -0.013 | 0.002 | 9.5e-10 |
| rs17453815 | 8 | 5549619 | G | A | 0.231 | -0.020 | 0.003 | 1.8e-14 |
| rs28494016 | 8 | 9187961 | C | A | 0.792 | -0.018 | 0.003 | 1.2e-11 |
| rs4840514 | 8 | 10612925 | A | G | 0.312 | -0.022 | 0.002 | 2.2e-21 |
| rs12550367 | 8 | 22793451 | C | T | 0.347 | 0.015 | 0.002 | 1.3e-11 |
| rs2979489 | 8 | 30280833 | A | G | 0.742 | -0.021 | 0.002 | 2.3e-18 |
| rs66593272 | 8 | 41548911 | T | A | 0.037 | -0.033 | 0.006 | 4.1e-09 |
| rs4737009 | 8 | 41630405 | A | G | 0.236 | 0.025 | 0.003 | 1.7e-23 |
| rs45577137 | 8 | 48651633 | G | A | 0.045 | -0.033 | 0.006 | 1.9e-09 |
| rs7846314 | 8 | 61650831 | T | A | 0.187 | -0.047 | 0.003 | 3.8e-66 |
| rs10111413 | 8 | 72891748 | A | G | 0.231 | -0.016 | 0.003 | 1.2e-10 |
| rs75653581 | 8 | 78363128 | T | C | 0.013 | 0.068 | 0.010 | 5.5e-12 |
| rs2919917 | 8 | 79657263 | C | T | 0.250 | 0.041 | 0.002 | 8.5e-62 |
| rs4521730 | 8 | 101180650 | A | G | 0.185 | 0.019 | 0.003 | 1.8e-11 |
| rs6988147 | 8 | 101459320 | T | C | 0.863 | -0.020 | 0.003 | 1.1e-10 |
| rs6988140 | 8 | 126480543 | T | C | 0.543 | -0.024 | 0.002 | 1e-24 |
| rs1865223 | 8 | 130606476 | T | G | 0.568 | 0.040 | 0.002 | 5e-74 |
| rs16904181 | 8 | 130976074 | A | G | 0.301 | 0.016 | 0.002 | 3.3e-11 |
| rs2219143 | 9 | 2622278 | A | G | 0.393 | -0.016 | 0.002 | 3e-12 |
| rs385893 | 9 | 4763176 | C | T | 0.524 | -0.026 | 0.002 | 8.9e-34 |
| rs3731211 | 9 | 21986847 | A | T | 0.722 | 0.026 | 0.002 | 4.7e-27 |
| rs2065500 | 9 | 22145694 | G | A | 0.158 | -0.027 | 0.003 | 1.8e-19 |
| rs10971930 | 9 | 34116083 | C | T | 0.127 | 0.020 | 0.003 | 8.2e-10 |
| rs10867416 | 9 | 82324152 | C | T | 0.656 | -0.016 | 0.002 | 1.4e-12 |
| rs61750929 | 9 | 91495135 | T | C | 0.056 | -0.055 | 0.005 | 2.5e-31 |
| rs2120229 | 9 | 123440875 | G | T | 0.491 | 0.014 | 0.002 | 2.5e-10 |
| rs2519093 | 9 | 136141870 | T | C | 0.184 | 0.027 | 0.003 | 1.1e-22 |
| rs28545822 | 9 | 139217805 | C | T | 0.467 | 0.014 | 0.002 | 2.1e-10 |
| rs11145986 | 9 | 139319847 | G | A | 0.290 | -0.019 | 0.002 | 9.3e-16 |
| rs61839660 | 10 | 6094697 | T | C | 0.098 | -0.046 | 0.004 | 1.1e-36 |
| rs2388486 | 10 | 8099021 | C | T | 0.830 | 0.024 | 0.003 | 1.1e-16 |
| rs2992327 | 10 | 26711899 | T | A | 0.273 | -0.016 | 0.002 | 1.1e-10 |
| rs2807742 | 10 | 28781367 | A | G | 0.770 | -0.022 | 0.003 | 2.9e-17 |
| rs17156360 | 10 | 44922040 | T | C | 0.132 | 0.020 | 0.003 | 3.6e-10 |
| rs56197478 | 10 | 50301892 | G | A | 0.221 | -0.017 | 0.003 | 8.5e-11 |
| rs7090445 | 10 | 63721176 | T | C | 0.660 | -0.022 | 0.002 | 5.1e-23 |
| rs142239370 | 10 | 72382869 | C | A | 0.040 | 0.053 | 0.005 | 3.8e-22 |
| rs1250546 | 10 | 81032532 | G | A | 0.437 | 0.019 | 0.002 | 7.2e-19 |
| rs1539330 | 10 | 94502738 | T | C | 0.134 | 0.025 | 0.003 | 6.2e-15 |
| rs10882827 | 10 | 98371790 | T | C | 0.179 | -0.018 | 0.003 | 1.4e-10 |
| rs11189132 | 10 | 99086195 | C | G | 0.396 | 0.025 | 0.002 | 5.9e-29 |
| rs11198861 | 10 | 121049819 | A | G | 0.512 | 0.016 | 0.002 | 2.7e-13 |
| rs2885521 | 10 | 126373313 | T | C | 0.110 | -0.029 | 0.003 | 7.3e-17 |
| rs14408 | 11 | 308314 | C | T | 0.368 | -0.031 | 0.002 | 3.8e-44 |
| rs12289940 | 11 | 4224589 | A | T | 0.378 | -0.013 | 0.002 | 1.4e-09 |
| rs4756265 | 11 | 36078171 | G | A | 0.418 | 0.016 | 0.002 | 1.8e-13 |
| rs4647725 | 11 | 47245389 | C | T | 0.157 | -0.023 | 0.003 | 1.9e-14 |
| rs7232 | 11 | 59940599 | A | T | 0.378 | 0.019 | 0.002 | 1.3e-17 |
| rs61897795 | 11 | 61618169 | G | A | 0.164 | 0.021 | 0.003 | 8.4e-13 |
| rs11231693 | 11 | 63862612 | A | G | 0.055 | -0.028 | 0.005 | 3.2e-09 |
| rs554169857 | 11 | 65686473 | A | C | 0.020 | -0.048 | 0.008 | 1.8e-09 |
| rs7930161 | 11 | 78268878 | A | C | 0.826 | 0.020 | 0.003 | 2.9e-12 |
| rs72992361 | 11 | 100427512 | A | C | 0.082 | -0.025 | 0.004 | 9.8e-11 |
| rs680113 | 11 | 108165406 | A | G | 0.555 | -0.016 | 0.002 | 3.7e-14 |
| rs73000965 | 11 | 113982321 | A | T | 0.317 | -0.014 | 0.002 | 4e-09 |
| rs73564493 | 11 | 119056273 | A | G | 0.073 | -0.026 | 0.004 | 1.5e-10 |
| rs11218725 | 11 | 122509237 | G | A | 0.377 | 0.029 | 0.002 | 9e-40 |
| rs4520607 | 11 | 128406775 | C | T | 0.525 | 0.021 | 0.002 | 1.6e-21 |
| rs2364482 | 12 | 6502131 | G | T | 0.197 | 0.041 | 0.003 | 4.1e-51 |
| rs7960048 | 12 | 12554861 | G | T | 0.466 | 0.013 | 0.002 | 1.3e-09 |
| rs35071163 | 12 | 12883507 | C | G | 0.479 | 0.017 | 0.002 | 2e-15 |
| rs113736796 | 12 | 48213720 | G | C | 0.039 | -0.047 | 0.006 | 1.7e-16 |
| rs1793937 | 12 | 48375475 | G | C | 0.401 | 0.015 | 0.002 | 2.3e-11 |
| rs75639074 | 12 | 56367901 | G | C | 0.063 | 0.026 | 0.004 | 4.8e-09 |
| rs4761234 | 12 | 69732105 | C | T | 0.485 | -0.020 | 0.002 | 7.4e-21 |
| rs4842610 | 12 | 88811689 | T | C | 0.705 | 0.015 | 0.002 | 2.8e-10 |
| rs653178 | 12 | 112007756 | T | C | 0.518 | -0.041 | 0.002 | 2e-81 |
| rs11064881 | 12 | 120146925 | A | G | 0.073 | 0.026 | 0.004 | 4.5e-10 |
| rs28413626 | 12 | 123861452 | A | G | 0.214 | 0.032 | 0.003 | 2.1e-34 |
| rs117808697 | 13 | 28513538 | T | A | 0.073 | -0.027 | 0.005 | 4.4e-09 |
| rs2504235 | 13 | 28612886 | G | A | 0.638 | -0.024 | 0.002 | 2.8e-27 |
| rs9532679 | 13 | 41522338 | C | A | 0.150 | -0.034 | 0.003 | 2.2e-29 |
| rs9533100 | 13 | 42996548 | T | G | 0.536 | -0.021 | 0.002 | 3.8e-23 |
| rs9316484 | 13 | 50645164 | C | T | 0.220 | -0.018 | 0.003 | 2.9e-12 |
| rs67483792 | 13 | 72503638 | T | C | 0.043 | -0.040 | 0.005 | 2.1e-13 |
| rs9530281 | 13 | 74679161 | A | G | 0.716 | -0.031 | 0.002 | 2.7e-39 |
| rs7981577 | 13 | 114835802 | C | T | 0.437 | -0.020 | 0.002 | 5e-20 |
| rs2004925 | 14 | 25445362 | A | G | 0.393 | -0.026 | 0.002 | 6.7e-31 |
| rs10131567 | 14 | 35864878 | T | G | 0.285 | -0.020 | 0.002 | 4.6e-17 |
| rs11848179 | 14 | 64977053 | G | A | 0.318 | 0.025 | 0.002 | 9.9e-27 |
| rs112506826 | 14 | 69975724 | T | G | 0.030 | -0.042 | 0.006 | 5.7e-11 |
| rs9323612 | 14 | 75968608 | G | A | 0.331 | -0.021 | 0.002 | 8.5e-20 |
| rs61975764 | 14 | 93014929 | A | G | 0.465 | -0.019 | 0.002 | 2.9e-18 |
| rs72699870 | 14 | 93116351 | C | T | 0.195 | 0.022 | 0.003 | 2.6e-16 |
| rs11625841 | 14 | 99787017 | C | T | 0.619 | -0.014 | 0.002 | 7.1e-10 |
| rs8008502 | 14 | 103359492 | A | G | 0.202 | 0.031 | 0.003 | 5.8e-30 |
| rs1132639 | 15 | 41597437 | A | T | 0.295 | 0.018 | 0.002 | 1.4e-14 |
| rs4924590 | 15 | 42233673 | A | T | 0.244 | -0.027 | 0.003 | 9.1e-28 |
| rs60820695 | 15 | 48584883 | T | C | 0.134 | -0.022 | 0.003 | 8.6e-12 |
| rs6494476 | 15 | 64657496 | T | C | 0.879 | -0.022 | 0.003 | 2.5e-11 |
| rs77365378 | 15 | 69972289 | T | C | 0.046 | 0.032 | 0.005 | 8.5e-10 |
| rs12908814 | 15 | 75132490 | G | C | 0.629 | -0.017 | 0.002 | 4.9e-15 |
| rs17832339 | 15 | 90557494 | T | G | 0.039 | 0.037 | 0.006 | 2.1e-11 |
| rs28539372 | 15 | 91021412 | A | T | 0.319 | 0.035 | 0.002 | 1.3e-50 |
| rs67175901 | 15 | 101748227 | T | C | 0.108 | -0.031 | 0.003 | 3.2e-19 |
| rs447361 | 16 | 4148240 | G | T | 0.669 | 0.015 | 0.002 | 3.2e-11 |
| rs61739285 | 16 | 27480797 | T | C | 0.034 | 0.044 | 0.006 | 7.8e-14 |
| rs11150589 | 16 | 30482494 | C | T | 0.517 | 0.027 | 0.002 | 1.5e-36 |
| rs7192652 | 16 | 57075180 | G | A | 0.479 | 0.023 | 0.002 | 2.3e-25 |
| rs11640346 | 16 | 81603848 | T | G | 0.537 | -0.014 | 0.002 | 2.7e-10 |
| rs1519978 | 16 | 85935211 | T | A | 0.168 | -0.022 | 0.003 | 3.2e-14 |
| rs72803323 | 16 | 89373324 | T | C | 0.040 | -0.038 | 0.005 | 6.8e-12 |
| rs9905106 | 17 | 1373518 | C | T | 0.735 | -0.024 | 0.002 | 3.4e-23 |
| rs9674692 | 17 | 2730030 | T | G | 0.277 | 0.018 | 0.002 | 6.3e-14 |
| rs28662192 | 17 | 4604497 | C | T | 0.294 | 0.019 | 0.002 | 1.8e-15 |
| rs9895848 | 17 | 4738958 | T | C | 0.345 | -0.019 | 0.002 | 1.3e-16 |
| rs9910148 | 17 | 16169009 | A | G | 0.476 | 0.038 | 0.002 | 5.9e-69 |
| rs4924997 | 17 | 19422725 | T | A | 0.460 | -0.015 | 0.002 | 4.9e-12 |
| rs4239225 | 17 | 38127112 | A | G | 0.472 | -0.069 | 0.002 | 1e-200 |
| rs11654706 | 17 | 38147293 | G | A | 0.168 | 0.021 | 0.003 | 7.9e-13 |
| rs146346285 | 17 | 38897220 | T | C | 0.014 | -0.074 | 0.010 | 1.8e-14 |
| rs8081535 | 17 | 41080195 | A | G | 0.436 | 0.018 | 0.002 | 1.2e-14 |
| rs1706725 | 17 | 44324185 | G | A | 0.236 | -0.028 | 0.003 | 1.1e-27 |
| rs28730837 | 17 | 56355397 | A | G | 0.016 | -0.060 | 0.008 | 1.9e-12 |
| rs138284624 | 17 | 56385573 | T | C | 0.013 | -0.103 | 0.010 | 6.8e-27 |
| rs13342015 | 17 | 73067482 | A | G | 0.304 | -0.015 | 0.002 | 4.8e-11 |
| rs4789294 | 17 | 74419177 | G | A | 0.217 | 0.026 | 0.003 | 6.4e-23 |
| rs11659109 | 17 | 75375859 | T | C | 0.483 | -0.016 | 0.002 | 3.5e-14 |
| rs7231555 | 18 | 19699122 | G | A | 0.088 | -0.034 | 0.004 | 1e-18 |
| rs75763843 | 18 | 42062380 | C | A | 0.129 | -0.032 | 0.003 | 6.2e-23 |
| rs9947316 | 18 | 43777831 | G | T | 0.559 | 0.013 | 0.002 | 5.8e-10 |
| rs66836460 | 18 | 60837508 | A | C | 0.136 | 0.021 | 0.003 | 9.9e-12 |
| rs56249713 | 18 | 67533332 | C | T | 0.420 | 0.016 | 0.002 | 4.1e-13 |
| rs454854 | 18 | 77253866 | C | G | 0.679 | -0.015 | 0.002 | 6.1e-11 |
| rs36084354 | 19 | 1079959 | A | G | 0.092 | -0.038 | 0.004 | 2.4e-24 |
| rs28841391 | 19 | 1193965 | G | T | 0.759 | 0.024 | 0.003 | 9.1e-19 |
| rs2569703 | 19 | 10404227 | G | C | 0.544 | -0.032 | 0.002 | 4.2e-49 |
| rs537835324 | 19 | 12994335 | T | C | 0.191 | -0.019 | 0.003 | 6.4e-11 |
| rs345627 | 19 | 13917264 | T | C | 0.349 | 0.023 | 0.002 | 7.6e-25 |
| rs10423726 | 19 | 16444697 | A | C | 0.580 | -0.036 | 0.002 | 3.1e-54 |
| rs7251806 | 19 | 16499569 | T | C | 0.841 | -0.088 | 0.003 | 1e-194 |
| rs4530278 | 19 | 33752994 | T | G | 0.601 | 0.018 | 0.002 | 6.9e-16 |
| rs59058760 | 19 | 40811757 | C | T | 0.068 | 0.026 | 0.004 | 7.9e-10 |
| rs4760 | 19 | 44153100 | G | A | 0.156 | 0.039 | 0.003 | 3e-39 |
| rs11673093 | 19 | 45742094 | A | G | 0.260 | 0.027 | 0.002 | 1.5e-28 |
| rs8102492 | 19 | 49140809 | C | A | 0.796 | -0.036 | 0.003 | 3e-40 |
| rs1028397 | 20 | 1929662 | A | G | 0.331 | -0.033 | 0.002 | 1.3e-47 |
| rs1010608 | 20 | 3784593 | A | G | 0.236 | 0.018 | 0.003 | 4.3e-12 |
| rs73125628 | 20 | 20066701 | T | C | 0.278 | 0.015 | 0.002 | 1.3e-10 |
| rs910083 | 20 | 31378690 | C | A | 0.447 | 0.016 | 0.002 | 5.6e-14 |
| rs6073369 | 20 | 42832789 | T | C | 0.314 | 0.019 | 0.002 | 2.4e-16 |
| rs2230604 | 20 | 49196284 | T | C | 0.084 | 0.029 | 0.004 | 7.3e-14 |
| rs259981 | 20 | 57748369 | A | T | 0.359 | -0.026 | 0.002 | 3.7e-31 |
| rs4809330 | 20 | 62349586 | G | A | 0.668 | -0.020 | 0.002 | 2.3e-18 |
| rs1736023 | 21 | 16812882 | T | C | 0.430 | 0.014 | 0.002 | 1.9e-10 |
| rs2212931 | 21 | 39859190 | G | A | 0.417 | -0.014 | 0.002 | 1.8e-10 |
| rs220262 | 21 | 43482046 | A | T | 0.482 | -0.016 | 0.002 | 2.2e-13 |
| rs5747308 | 22 | 18133500 | C | A | 0.505 | -0.019 | 0.002 | 7.9e-18 |
| rs5998509 | 22 | 21917479 | T | C | 0.188 | -0.033 | 0.003 | 2.5e-33 |
| rs2144922 | 22 | 28923317 | A | G | 0.878 | 0.027 | 0.003 | 2.7e-16 |
| rs2412971 | 22 | 30494371 | A | G | 0.446 | 0.028 | 0.002 | 3e-37 |
| rs5757886 | 22 | 40636076 | C | T | 0.747 | -0.017 | 0.002 | 2e-12 |
| rs5758307 | 22 | 41750219 | G | A | 0.229 | 0.028 | 0.003 | 1.6e-27 |

**Table S19. 9 SNPs significantly associated with COVID-19 severity (very severe respiratory confirmed covid vs. population) used IVs in reverse MR analyses derived from the COVID-19 Host Genetics Initiative.** Chr: Chromosome; EA: Effect allele; NEA: Non-effect allele; EAF: Effect allele frequency.

| SNP | Chr | Pos | EA | NEA | EAF | Beta | SE | P-value |
| --- | --- | --- | --- | --- | --- | --- | --- | --- |
| rs35081325 | 3 | 45889921 | T | A | 0.075 | 0.626 | 0.045 | 5.754e-45 |
| rs111837807 | 6 | 31121232 | C | T | 0.100 | 0.295 | 0.043 | 5.656e-12 |
| rs622568 | 7 | 54647894 | C | A | 0.146 | 0.226 | 0.037 | 1.044e-09 |
| rs2237698 | 7 | 107607902 | T | C | 0.090 | 0.237 | 0.040 | 2.414e-09 |
| rs10860891 | 12 | 103014757 | A | C | 0.885 | -0.239 | 0.040 | 1.636e-09 |
| rs2384074 | 12 | 113382977 | T | C | 0.676 | 0.198 | 0.028 | 2.104e-12 |
| rs77534576 | 17 | 47940666 | T | C | 0.035 | 0.460 | 0.075 | 8.525e-10 |
| rs2109069 | 19 | 4719443 | A | G | 0.329 | 0.257 | 0.028 | 6.115e-20 |
| rs13050728 | 21 | 34615210 | C | T | 0.663 | -0.200 | 0.029 | 2.437e-12 |

**Table S20. 5 SNPs significantly associated with COVID-19 severity (hospitalized covid vs. population) used IVs in reverse MR analyses derived from the COVID-19 Host Genetics Initiative.** Chr: Chromosome; EA: Effect allele; NEA: Non-effect allele; EAF: Effect allele frequency.

| SNP | Chr | Pos | EA | NEA | EAF | Beta | SE | P-value |
| --- | --- | --- | --- | --- | --- | --- | --- | --- |
| rs35081325 | 3 | 45889921 | T | A | 0.081 | 0.488 | 0.032 | 3.684e-54 |
| rs622568 | 7 | 54647894 | C | A | 0.151 | 0.154 | 0.026 | 3.644e-09 |
| rs2660 | 12 | 113357442 | A | G | 0.690 | 0.116 | 0.019 | 2.005e-09 |
| rs2109069 | 19 | 4719443 | A | G | 0.323 | 0.151 | 0.020 | 2.937e-14 |
| rs13050728 | 21 | 34615210 | C | T | 0.653 | -0.168 | 0.020 | 7.437e-17 |

**Table S21. 7 SNPs significantly associated with COVID-19 susceptibility (covid vs. population) used IVs in reverse MR analyses derived from the COVID-19 Host Genetics Initiative.** Chr: Chromosome; EA: Effect allele; NEA: Non-effect allele; EAF: Effect allele frequency.

| SNP | Chr | Pos | EA | NEA | EAF | Beta | SE | P-value |
| --- | --- | --- | --- | --- | --- | --- | --- | --- |
| rs4971066 | 1 | 155105882 | G | T | 0.178 | -0.077 | 0.013 | 1.015e-08 |
| rs2271616 | 3 | 45838013 | T | G | 0.118 | 0.156 | 0.015 | 3.608e-25 |
| rs17078348 | 3 | 45847241 | G | A | 0.100 | 0.092 | 0.016 | 1.196e-08 |
| rs10936744 | 3 | 101433440 | T | C | 0.359 | -0.063 | 0.010 | 3.511e-10 |
| rs579459 | 9 | 136154168 | T | C | 0.790 | -0.094 | 0.011 | 2.183e-16 |
| rs757405 | 12 | 113406945 | A | T | 0.709 | 0.069 | 0.011 | 1.639e-10 |
| rs12482060 | 21 | 34611571 | G | C | 0.338 | 0.062 | 0.011 | 3.958e-09 |
